## Supplementary Data 7 for "Effective high-throughput RT-qPCR screening for SARS-CoV-2 infections in children": analysis_ensemble1.py

```
#---------Import---------
import sys
import time
from tqdm import tqdm
import numpy as np
import matplotlib.pyplot as plt
import matplotlib as mpl
import pandas as pd
sys.path.append('../lib/')
from models import *
from networks import *
from sim_loops import *
from utilities import *
from functions import *
from matplotlib.ticker import FormatStrFormatter
from matplotlib.ticker import PercentFormatter
import matplotlib.ticker as mtick

Text_files_path = '../../Text_files/SEIR_ensemble/'

print('Hola mundo \n')

N=20
N_groups = 12
NN = N*N_groups

p_exts = [1e-4, 1e-3]
R0s = np.flip(np.array([2.5, 4.5, 7.5]))
variants  = np.flip(['WT', r'$\alpha$', r'$\delta$'])
markers_R0 = np.flip(['^', 's', 'o'])
#R0s = np.array([0.5])

Q_days = np.array([0, 14])
#colors_p_ext = plt.cm.Blues(np.linspace(0,1,len(p_exts)+2))
colors_R0 = np.flip(np.array([['tab:blue']*3, ['tab:green']*3, ['tab:red']*3], dtype = object))

#testing_frequencies = ['weekly', 'weekly2', 'semiweekly', 'semiweekly2', 'triweekly', 'never']
testing_frequencies = ['never', 'weekly2', 'semiweekly2', 'triweekly']
testing_frequencies_names = ['No test', 'weekly', 'semiweekly', 'triweekly']

data_dict = {}

for i, d in enumerate(Q_days):
	fig3, ax3 = plt.subplots(figsize = (10,8), gridspec_kw={'bottom': 0.2, 'left': 0.16, 'top':.95, 'right':.95})
	for r, R0 in enumerate(R0s):
		colors_p_ext = colors_R0[r]

		for l, p_ext in enumerate(p_exts):
			positions = np.array([])
			values = np.array([])
			errors = np.array([])
			ax3.text(.18+(1/len(p_exts))*(l), .95, r'$P_{\mathrm{ext}} =$%.e'%(p_ext), fontsize = 16, transform=ax3.transAxes)
			
			for t, testing_frequency in enumerate(testing_frequencies):
				#fig2, ax2 = plt.subplots(figsize = (10,8), gridspec_kw={'bottom': 0.13, 'left': 0.14})
				#fig3, ax3 = plt.subplots(figsize = (10,8), gridspec_kw={'bottom': 0.13, 'left': 0.14})
				infile = open(Text_files_path+'I_cummulative_pop_R0-%.1f_p_ext-%.2e_d-%.d_'%(R0, p_ext, d)+'testing_frequency-'+testing_frequency+'_interactions-1_0.pkl','rb')
				I_cum = (pickle.load(infile))
				infile.close()
				avg = np.mean(I_cum)
				var = np.var(I_cum)

				infile_no_interactions = open(Text_files_path+'I_cummulative_pop_R0-%.1f_p_ext-%.2e_d-%.d_'%(R0, p_ext, d)+'testing_frequency-'+testing_frequency+'_interactions-0_0.pkl','rb')				
				I_cum_no_interactions = (pickle.load(infile_no_interactions))
				infile_no_interactions.close()
				avg_no_interactions = np.mean(I_cum_no_interactions)
				var_no_interactions = np.var(I_cum_no_interactions)

				if(testing_frequency=='never'):
					avg_no_test = avg
					avg_no_interactions_no_test = avg_no_interactions
					var_no_test = var
					var_no_interactions_no_test = var_no_interactions

				pos_violin = 4*l + 1*t

				T = avg-avg_no_interactions
				T_NT = avg_no_test-avg_no_interactions_no_test

				D_T = np.sqrt(var + var_no_interactions)
				D_T_NT = np.sqrt(var_no_test + var_no_interactions_no_test)

				positions = np.append(positions, pos_violin)
				values = np.append(values, 1-(T)/(T_NT))
				#print(d, R0, p_ext, testing_frequency, 1-(T)/(T_NT))

				data_dict[(d, p_ext, R0, testing_frequency)] = {'Prevented transmission': 1-(T)/(T_NT)}


			# --- Plot 3 ---
			#ax3.scatter(pos_violin, 1-(I_cum_avg-I_cum_avg_no_interactions)/I_cum_avg_no_test, color = colors_p_ext[l], marker = markers_R0[r], s = 50)
			ax3.plot(positions, values, color = colors_p_ext[l], marker = markers_R0[r], ms = 12, linestyle = '--')


		ax3.vlines([k*4 - 0.5 for k in np.arange(4)], 0, 1, color='black', linestyle = 'dotted')
		my_plot_layout(ax=ax3, xlabel = 'Testing frequency', ylabel = 'Prevented Transmissions', yscale='linear')
		ax3.set_xticks(np.arange(0, (len(testing_frequencies))*len(p_exts) , 1))
		ax3.set_xticklabels(testing_frequencies_names*len(p_exts))
		ax3.tick_params('x', labelsize = 16,  rotation=45)
		ax3.yaxis.set_major_formatter(PercentFormatter(1))
		ax3.set_ylim(0,1.1)
		ax3.set_xlim(-1,len(p_exts)*4)
		ax3.set_title( '%d'%(d) + ' Isolations days',fontsize = 22)
		#ax3.legend(fontsize = 22, loc = 1)
		fig3.savefig('../../Figures/1_Extended_Model/ensemble/1/PrevTrans_d-%d'%(d)+'.pdf')
		plt.close(fig3)

data_frame = pd.DataFrame.from_dict(data_dict)
data_frame.to_csv(Text_files_path+'../data/data_simulations_Nov.csv')
data_frame.to_excel(Text_files_path+'../data/data_simulations_Nov.xlsx')
```
