## Supplementary Data 7 for "Effective high-throughput RT-qPCR screening for SARS-CoV-2 infections in children": extra_figures.py

```
#---------Import---------
import sys
import time
from tqdm import tqdm
import numpy as np
import matplotlib.pyplot as plt
import matplotlib as mpl
import pandas as pd
sys.path.append('../lib/')
from models import *
from networks import *
from sim_loops import *
from utilities import *
from functions import *
from matplotlib.ticker import FormatStrFormatter
import matplotlib.ticker as mtick

Text_files_path = '../Text_files/SEIR_ensemble/'


colors = ['#C5835D', '#E7BF63']
lamdas = [1/3, 1/6]
labels = [r'$\tau_{p}$', r'$\tau_{i}$']
xlabels = ['Pre-infectious period', 'Infectious period']
t = np.arange(1, 25)
fig, ax = plt.subplots(1, 2, figsize = (12, 6), gridspec_kw={'wspace':.32, 'bottom':.15})
for l, lamda in enumerate(lamdas):
	p_ext = lamda*np.exp(-lamda*t)
	ax[l].bar(t, p_ext, color = colors[l])
	ax[l].vlines(1/lamda, 0, 0.24, color = colors[l], label = labels[l], linestyle = 'dotted', lw = 3)
	ax[l].set_ylim(0, 0.25)
	my_plot_layout(ax=ax[l], xlabel = xlabels[l], ylabel = 'Probability')
	ax[l].legend(fontsize= 24)

fig.savefig('../../Figures/tau_distributions.pdf')
```
