## Supplementary Data 7 for "Effective high-throughput RT-qPCR screening for SARS-CoV-2 infections in children": FARZ.py

```
#################################################
# Adapted from https://github.com/rabbanyk/FARZ
#################################################

import random
import bisect
import math
import os 

def random_choice(values, weights=None , size = 1, replace = True):
    if weights is None:
        i = int(random.random() * len(values))
    else :
        total = 0
        cum_weights = []
        for w in weights:
            total += w
            cum_weights.append(total)
        x = random.random() * total
        i = bisect.bisect(cum_weights, x)
    if size <=1: 
        if len(values)>i: return values[i] 
        else: return None
    else: 
        cval = [values[j] for j in range(len(values)) if replace or i!=j]
        if weights is None: cwei=None 
        else: cwei = [weights[j] for j in range(len(weights)) if replace or i!=j]
        tmp= random_choice(cval, cwei, size-1, replace)
        if not isinstance(tmp,list): tmp = [tmp]
        tmp.append(values[i])
        return tmp 

class Comms:
     def __init__(self, k):
         self.k = k
         self.groups = [[] for i in range(k)]
         self.memberships = {}
         
     def add(self, cluster_id, i, s = 1):
         if i not in  [m[0] for m in self.groups[cluster_id]]:
            self.groups[cluster_id].append((i,s)) 
            if i in self.memberships:
                self.memberships[i].append((cluster_id,s))
            else:
                self.memberships[i] =[(cluster_id,s)] 
     def write_groups(self, path):
         with open(path, 'w') as f:
             for g in self.groups:
                 for i,s in g:
                    f.write(str(i) + ' ')
                 f.write('\n')

             
            
class Graph:
    def __init__(self,directed=False, weighted=False):
        self.n = 0
        self.counter = 0
        self.max_degree = 0
        self.directed = directed
        self.weighted = weighted
        self.edge_list = [] 
        self.edge_time = []
        self.deg = []
        self.neigh =  [[]]
        return 

    def add_node(self):
        self.deg.append(0)
        self.neigh.append([])
        self.n+=1
    
    def weight(self, u, v):
        for i,w in self.neigh[u]:
            if i == v: return w
        return 0
    
    def is_neigh(self, u, v):
        for i,_ in self.neigh[u]:
            if i == v: return True
        return False
    
    def add_edge(self, u, v, w=1):
        if u==v: return
        if not self.weighted : w =1
        self.edge_list.append((u,v,w) if u<v or self.directed else  (v,u,w))
        self.edge_time.append(self.counter)
        self.counter +=1
        self.neigh[u].append((v,w))
        self.deg[v]+=w
        if  self.deg[v]>self.max_degree: self.max_degree = self.deg[v]
        
        if not self.directed: #if directed deg is indegree, outdegree = len(negh)
            self.neigh[v].append((u,w))
            self.deg[u]+=w
            if  self.deg[u]>self.max_degree: self.max_degree = self.deg[u]

        return 
    
     
    def to_nx(self):
        import networkx as nx
        G=nx.Graph()
        for u,v, w in self.edge_list:
            G.add_edge(u, v)
            # G.add_edges_from(self.edge_list)
        return G
    
    def to_nx(self, C):
        import networkx as nx
        G=nx.Graph()
        for i in range(self.n):
            # This line works for networkx 1.10:
            # G.add_node(i, {'c':str(sorted(C.memberships[i]))})
            # This line works for networkx 2.2:
            G.add_node(i, c=str(sorted(C.memberships[i])))
            # G.add_node(i, {'c':int(C.memberships[i][0][0])})
        for i in range(len(self.edge_list)):
        # for u,v, w in self.edge_list:
            u,v, w = self.edge_list[i]
            G.add_edge(u, v, weight=w, capacity=self.edge_time[i])
            # G.add_edges_from(self.edge_list)
        return G
    
    def to_ig(self):
        G=ig.Graph()
        G.add_edges(self.edge_list)
        return G 
 
 
    def write_edgelist(self, path):
         with open(path, 'w') as f:
             for i,j,w in self.edge_list:
                 f.write(str(i) + '\t'+str(j) + '\n')

 
def Q(G, C):
    q = 0.0
    m = 2 * len(G.edge_list)
    for c in C.groups:
        for i,_ in c:
            for j,_ in c:
                q+= G.weight(i,j) - (G.deg[i]*G.deg[j]/(2*m))
    q /= 2*m
    return q

def common_neighbour(i, G, normalize=True):
    p = {}
    for k,wik in G.neigh[i]:
        for j,wjk in G.neigh[k]:
            if j in p: p[j]+=(wik * wjk) 
            else: p[j]= (wik * wjk)
    if len(p)<=0 or not normalize: return p
    maxp = p[max(p, key = lambda i: p[i])]
    for j in p:  p[j] = p[j]*1.0 / maxp
    return p

def choose_community(i, G, C, alpha, beta, gamma, epsilon):
    mids =[k for  k,uik in C.memberships[i]]
    if random.random()< beta: #inside
        cids = mids
    else:     
        cids = [j for j in range(len(C.groups)) if j not in mids] #:  cids.append(j)

    return cids[ int(random.random()*len(cids))] if len(cids)>0 else None

def degree_similarity(i, ids, G, gamma, normalize = True):
    p = [0]*len(ids)
    for ij,j in enumerate(ids):
        p[ij] =  (G.deg[j] -G.deg[i])**2
    if len(p)<=0 or not normalize: return p
    maxp = max(p)
    if maxp==0: return p
    p = [pi*1.0/maxp if gamma<0 else 1-pi*1.0/maxp for pi in p]
    return p

def combine (a,b,alpha,gamma):
    return (a**alpha) / ((b+1)**gamma)

def choose_node(i,c, G, C, alpha, beta, gamma, epsilon):
    ids = [j for j,_ in C.groups[c] if j !=i ]
    #   also remove nodes that are already connected from the candidate list
    for k,_ in G.neigh[i]: 
        if k in ids: ids.remove(k) 

    norma = False
    cn = common_neighbour(i, G, normalize=norma)
    trim_ids = [id for id in ids if id in cn]
    dd = degree_similarity(i, trim_ids, G, gamma, normalize=norma)
    
    if random.random()<epsilon or len(trim_ids)<=0:
        tmp = int(random.random() * len(ids))
        if tmp==0: return  None
        return ids[tmp], epsilon
    else:
        p = [0 for j in range(len(trim_ids))]
        for ind in range(len(trim_ids)):
            j = trim_ids[ind]
            p[ind] = (cn[j]**alpha )/ ((dd[ind]+1)** gamma) 
            
        if(sum(p)==0): return  None
        tmp = random_choice(range(len(p)), p ) #, size=1, replace = False)
        # TODO add weights /direction/attributes
        if tmp is None: return  None
        return trim_ids[tmp], p[tmp]

 
def connect_neighbor(i, j, pj, c, b,  G, C, beta):
    if b<=0: return 
    ids = [id for id,_ in C.groups[c]]
    for k,wjk in G.neigh[j]:
        if (random.random() <b and k!=i and (k in ids or random.random()>beta)):
            G.add_edge(i,k,wjk*pj)
                    
def connect(i, b,  G, C, alpha, beta, gamma, epsilon):
    #Choose community
    c = choose_community(i, G, C, alpha, beta, gamma, epsilon)
    if c is None: return
    #Choose node within community
    tmp = choose_node(i, c, G, C, alpha, beta, gamma, epsilon)
    if tmp is None: return
    j, pj = tmp 
    G.add_edge(i,j,pj)
    connect_neighbor(i, j, pj , c, b,  G, C, beta)
            
def select_node(G, method = 'uniform'):
    if method=='uniform':   
        return int(random.random() * G.n) # uniform
    else:
        if method == 'older_less_active': p = [(i+1) for i in range(G.n)] # older less active
        elif method == 'younger_less_active' :  p = [G.n-i for i in range(G.n)] # younger less active
        else:  p = [1 for i in range(G.n)] # uniform
        return  random_choice(range(len(p)), p ) #, size=1, replace = False)[0]

def assign(i, C, e=1, r=1, q = 0.5):
    p = [e +len(c) for c in C.groups]
    id = random_choice(range(C.k),p )
    C.add(id, i)
    for j in range(1,r): #todo add strength for fuzzy
        if (random.random()<q): 
              id = random_choice(range(C.k),p )
              C.add(id, i)
    return
 
def print_setting(n,m,k,alpha,beta,gamma, phi,r,q,epsilon,weighted,directed):
    print('n:',n,'m:', m ,'k:', k,'alpha:', alpha,'beta:', beta,'gamma:', gamma,)
    if phi!=default_FARZ_setting['phi']: print('phi:', phi, )
    if r!=default_FARZ_setting['r']: print('r:', r,)
    if q!=default_FARZ_setting['q']: print('pr:', q, )
    if epsilon!=default_FARZ_setting['epsilon']:'epsilon:', epsilon, 
    print('weighted' if weighted else '', 'directed' if directed else '')
    
def realize(n, m,  k, b=0.0,  alpha=0.4, beta=0.5, gamma=0.1, phi=1, r=1, q = 0.5, epsilon = 0.0000001, weighted =False, directed=False):
    # print_setting(n,m,k,alpha,beta,gamma, phi,r,q,epsilon,weighted,directed)
    G =  Graph()
    C = Comms(k)
    for i in range(n):
    # if i%10==0: print('-- ',G.n, len(G.edge_list))
        G.add_node()
        assign(i, C, phi, r, q)
        connect(i,b, G, C, alpha, beta, gamma, epsilon)
        for e in range(1,m):
            j = select_node(G) 
            connect(j, b, G, C, alpha, beta, gamma, epsilon)        
    return G,C


def props():
    import plotNets as pltn
    import matplotlib as mpl
    mpl.rcParams['axes.unicode_minus']=False
    graphs = []
    names = []
    params = default_FARZ_setting.copy()
    for alp, gam in [(0.5,0.5), (0.8,0.2), (.5,-0.5), (0.2,-0.8)]:
        params[ "alpha"]=alp
        params[ "gamma"]=gam
        print(str(params))
        G, C =realize(**params)
        print('n=',G.n,' e=', len(G.edge_list))
        print('Q=',Q(G,C))
        G = G.to_nx(C)
        pltn.printGraphStats(G)
        graphs.append(G.to_undirected())
        # name = 'F'+str(params)
        name = '$\\alpha ='+str(params[ "alpha"]) +',\; \\gamma='+str(params[ "gamma"])+"$"
        names.append(name)
        nx.write_gml(graphs[-1], "farz-"+str(params[ "alpha"])+str(params[ "gamma"])+'.gml')
    
    pltn.plot_dists(graphs,names)

def write_to_file(G,C,path, name,format,params):
    if not os.path.exists(path+'/'): os.makedirs(path+'/')
    print('n=',G.n,' e=', len(G.edge_list), 'generated, writing to ', path+'/'+name, ' in', format)
    if format == 'gml':
        import networkx as nx
        G = G.to_nx(C)
        if not params['directed']: G = G.to_undirected()
        nx.write_gml(G, path+'/'+name+'.gml')
        return G
    if format == 'list': 
        G.write_edgelist(path+'/'+name+'.dat')
        C.write_groups( path+'/'+name+'.lgt')
        return G
 

default_ranges = {'beta':(0.5,1,0.05), 'k':(2,50,5), 'm':(2,11,1) , 'phi':(1,100,10), 'r':(1,10,1), 'q':(0.0,1,0.1)}
default_FARZ_setting = {"n":1000, "k":4, "m":5, "alpha":0.5,"gamma":0.5, "beta":.8, "phi":1, "o":1, 'q':0.5,  "b":0.0, "epsilon":0.0000001, 'directed':False, 'weighted':False}
default_batch_setting= {'vari':None, 'arange':None, 'repeat':1, 'path':'.', 'net_name':'network', 'format':'gml', 'farz_params':None}
supported_formats = ['gml','list']
def generate( vari =None, arange =None, repeat = 1, path ='.', net_name = 'network',format ='gml', farz_params= default_FARZ_setting.copy()):
    def get_range(s,e,i):
        res =[]
        while s<=e +1e-6:
            res.append(s)
            s +=i
        return res
    
    if vari is None:
        for r in range(repeat):
            G, C =realize(**farz_params)

            import networkx as nx
            G = G.to_nx(C)
            if not farz_params['directed']: G = G.to_undirected()


            name = net_name+( str(r+1) if repeat>1 else '') 
            # G = write_to_file(G,C,path,name,format,farz_params)
           
            # print(len([memtup[0][0] for memtup in C.memberships.values()]))
            # import numpy
            # (unique, counts) = numpy.unique( [memtup[0][0] for memtup in C.memberships.values()], return_counts=True )
            # print(numpy.asarray((unique, counts)).T)
            # exit()

            node_communities = {node: [c[0] for c in comm_tup] for node, comm_tup in C.memberships.items()}
            
        return G, node_communities

    if arange ==None:
        arange = default_ranges[vari]
    for i,var in enumerate(get_range(arange[0],arange[1],arange[2])): 
        for r in range(repeat):
            farz_params[vari] = var
            print('s',i+1, r+1, str(farz_params))
            G, C =realize(**farz_params)
            name = 'S'+str(i+1)+'-'+net_name+ (str(r+1) if repeat>1 else '') 
            write_to_file(G,C,path,name,format,farz_params)

    


import sys
def main(argv):
    import getopt
    FARZsetting = default_FARZ_setting.copy()
    batch_setting= default_batch_setting.copy()
    try:    
        opts, args = getopt.getopt(argv,"ho:s:v:c:f:n:k:m:a:b:g:p:r:q:t:e:dw",\
                                   ["output=","path=","repeat=","vary=",'range=','format=',"alpha=","beta=","gamma=",'phi=','overlap=','oProb=','epsilon=','cneigh=','directed','weighted'])
    except getopt.GetoptError:
        print('invalid command, try -h to see usage and options')
        sys.exit(2)
    for opt, arg in opts:
        if opt == '-h':
            print('*** examples:')
            print('+ example 1: generate a network with 1000 nodes and about 5x1000 edges (m=5), with 4 communities, where 90% of edges fall within communities (beta=0.9)')
            print('> python FARZ.py -n 1000 -m 5 -k 4 --beta 0.9\n')
            print('+ example 2: generate a network with properties of example 1, where alpha = 0.2 and gamma = -0.8')
            print('> python FARZ.py -n 1000 -m 5 -k 4 --beta 0.9 --alpha 0.2 --gamma -0.8 \n')
            print('+ example 3: generate 10 sample networks with properties of example 1 and save them into ./data')
            print('> python FARZ.py --path ./data -s 10 -n 1000 -m 5 -k 4 --beta 0.9\n')
            print('+ example 4: repeat example 2, for beta that varies from 0.5 to 1 with 0.05 increments')
            print('> python FARZ.py --path ./data -s 10 -v beta -c [0.5,1,0.05] -n 1000 -m 5 -k 4 \n')
            print('+ example 5: generate overlapping communities, where each node belongs to at most 3 communities and the portion of overlapping nodes varies')
            print('python FARZ.py -r 3 -v q --path ./datavrq -s 5 --format list\n')
            
            print('*** parameters:')
            print('-n: number of nodes, default (1000)')
            print('-m: half the average degree of nodes, default (5)')
            print('-k: number of communities, default (4)')
            print('-b [or --beta]: the strength of community structure, i.e. the probability of edges to be formed within communities, default (0.8)')
            print('-a [or --alpha]: the strength of common neighbor\'s effect on edge formation edges, default (0.5)')
            print('-g [or --gamma]: the strength of degree similarity effect on edge formation, default (0.5), can be negative for networks with negative degree correlation')
            print('-p [or --phi]: the constant added to all community sizes, higher number makes the communities more balanced in size, default (1), which results in power law distribution for community sizes')
            print('-r: the number of communities each node can belong to, default (1)' )
            print('-q: the probability of a node belonging to the multiple communities, default (0.5)' )
            print('-e [or --epsilon]: the probability of noisy/random edges, default (0.0000001)')
            print('-t: the probability of also connecting to the neighbors of a node each nodes connects to. The default value is (0), but could be increased to a small number to achieve higher clustering coefficient. \n')

            print('*** batch parameters:')
            print('-s: the number of networks to be sampled with the given properties, default (1)' )
            print('-o: the name of the output network, default (network)')
            print('--path : the path to write the network(s) to, default (.)')
            print('-f [or --format]: the format of output, list or gml, default (gml)')
            print('-v: the parameter to vary and sample networks for, default (None)')
            print('-c: the range to change the given parameter, should be in format of [s,e,inc]')
            #print('default FARZ parameters are :\n', default_FARZ_setting)
            #print('default batch generator parameters are :\n', default_batch_setting)
            
            sys.exit()
            
        elif opt in ("-o", "--output"):
            batch_setting['net_name'] = arg
        elif opt in ("--path"):
            batch_setting['path'] = arg
        elif opt in ("-f", "--format"):
            if arg in supported_formats:
                batch_setting['format'] = arg
            else:
                print('Format not supported , choose from ',supported_formats,' or try -h to see the usage and options')
                sys.exit(2)                
        elif opt in ("-s","--repeat"):
            try: batch_setting['repeat'] = int(arg) 
            except ValueError:
                print('Invalid Number , try -h to see the usage and options')
                sys.exit(2)            
        elif opt in ("-v", "--vary"):
            if (arg in list(default_ranges.keys())):
                batch_setting['vari'] = arg
            else:
                print('Invalid variable, choose form :', list(default_ranges.keys()), ', try -h to see the usage and options')
                sys.exit(2)
        elif opt in ("-c", "--range"):
            try:
                arange = [float(s) for s in arg[1:-1].split(',')]
                batch_setting['arange'] = arange
            except Error:
                print('Invalid range, should have the following form : [start,end,incrementBy], try -h to see the usage and options ')
                sys.exit(2)               
        elif opt in ("-n"):
            try: FARZsetting['n'] = int(arg) 
            except ValueError:
                print('Invalid Number , try -h to see the usage and options')
                sys.exit(2)
        elif opt in ("-k"):
            try: FARZsetting['k'] = int(arg)
            except ValueError:
                print('Invalid Number , try -h to see the usage and options')
                sys.exit(2)
        elif opt in ("-m"):
            try: FARZsetting['m'] = int(arg)
            except ValueError:
                print('Invalid Number , try -h to see the usage and options')
                sys.exit(2)
        elif opt in ("-a","--alpha"):
            try: FARZsetting['alpha'] = float(arg)  
            except ValueError:
                print('Invalid Number , try -h to see the usage and options')
                sys.exit(2)         
        elif opt in ("-b","--beta"):
            try: FARZsetting['beta'] = float(arg)
            except ValueError:
                print('Invalid Number , try -h to see the usage and options')
                sys.exit(2)                        
        elif opt in ("-g","--gamma"):
            try: FARZsetting['gamma'] = float(arg)
            except ValueError:
                print('Invalid Number , try -h to see the usage and options')
                sys.exit(2)  
        elif opt in ("-p","--phi"):
            try: FARZsetting['phi'] = int(arg)
            except ValueError:
                print('Invalid Number , try -h to see the usage and options')
                sys.exit(2)         
        elif opt in ("-r","--overlap"):
            try: FARZsetting['r'] = int(arg)
            except ValueError:
                print('Invalid Number , try -h to see the usage and options')
                sys.exit(2)      
        elif opt in ("-q","--oProb"):
            try: FARZsetting['q'] = float(arg)
            except ValueError:
                print('Invalid Number , try -h to see the usage and options')
                sys.exit(2)
        elif opt in ("-d","--directed"):
            FARZsetting['directed'] = True
        elif opt in ("-w","--wighted"):
            FARZsetting['weighted'] = True                           
        elif opt in ("-t","--cneigh"):
            try: FARZsetting['b'] = float(arg)
            except ValueError:
                print('Invalid Number , try -h to see the usage and options')
                sys.exit(2)         
        elif opt in ("-e","--epsilon"):
            try: FARZsetting['epsilon'] = float(arg)         
            except ValueError:
                print('Invalid Number , try -h to see the usage and options')
                sys.exit(2)
                
    batch_setting['farz_params'] = FARZsetting
    print('generating FARZ benchmark(s) ... ')
    generate( **batch_setting)
          

if __name__ == "__main__":
   main(sys.argv[1:])


# generate(farz_params={"n":25000, 
#                         "k":4, 
#                         "m":5, 
#                         "alpha":0.5,
#                         "gamma":0.5, 
#                         "beta":.8, 
#                         "phi":1, 
#                         "o":1, 
#                         'q':0.5,  
#                         "b":0.0, 
#                         "epsilon":0.0000001, 
#                         'directed':False, 
#                         'weighted':False})
   
   
# python FARZ.py --path ./dataVb55 -s 10 -v beta
# python FARZ.py --path ./dataVb82 -s 10 -v beta --alpha 0.8 --gamma 0.2
# python FARZ.py --path ./dataVb5-5 -s 10 -v beta --alpha 0.5 --gamma -0.5
# python FARZ.py --path ./dataVb2-8 -s 10 -v beta --alpha 0.2 --gamma -0.8
```
