## Supplementary Data 7 for "Effective high-throughput RT-qPCR screening for SARS-CoV-2 infections in children": functions.py

```
import numpy as np
import matplotlib.pyplot as plt
#from Bio import Phylo
from io import StringIO
from matplotlib.lines import Line2D
from datetime import datetime, timedelta
import scipy.special as sc
import seaborn as sns
import pickle
import json
from scipy.optimize import curve_fit

	

#----------------- Models -----------------


#----------------- Functions -----------------

def my_linear_func(x, a, b):

    return a + b*x

def my_quadratic_func(x, a, b, c):

    return np.log(a)+np.log(np.sqrt(-b)) + b*(x-c)**2

def my_plot_layout(ax, yscale = 'linear', xscale = 'linear', ticks_labelsize = 24, xlabel = '', ylabel = '', title = '', x_fontsize=24, y_fontsize = 24, t_fontsize = 24):
    ax.tick_params(labelsize = ticks_labelsize)
    ax.set_yscale(yscale)
    ax.set_xscale(xscale)
    ax.set_xlabel(xlabel, fontsize = x_fontsize)
    ax.set_ylabel(ylabel, fontsize = y_fontsize)
    ax.set_title(title, fontsize = t_fontsize)


#----------------- Plots -----------------


#----------------- Generate files -----------------


#----------------- Plots for ensemble averages -----------------


#----------------------------------------------------------------
```
