## Supplementary Data 7 for "Effective high-throughput RT-qPCR screening for SARS-CoV-2 infections in children": models.py

```
from __future__ import absolute_import
from __future__ import division
from __future__ import print_function

import networkx as networkx
import numpy as numpy
import scipy as scipy
import scipy.integrate


########################################################
#@@@@@@@@@@@@@@@@@@@@@@@@@@@@@@@@@@@@@@@@@@@@@@@@@@@@@@#
#@                                                    @#
#@  BASIC SEIRS MODELS                                @#
#@                                                    @#
#@@@@@@@@@@@@@@@@@@@@@@@@@@@@@@@@@@@@@@@@@@@@@@@@@@@@@@#
########################################################

class SEIRSModel():
    """
    A class to simulate the Deterministic SEIRS Model
    ===================================================
    Params: beta    Rate of transmission (exposure) 
            sigma   Rate of infection (upon exposure) 
            gamma   Rate of recovery (upon infection) 
            xi      Rate of re-susceptibility (upon recovery)  
            mu_I    Rate of infection-related death  
            mu_0    Rate of baseline death   
            nu      Rate of baseline birth
            
            beta_Q  Rate of transmission (exposure) for individuals with detected infections
            sigma_Q Rate of infection (upon exposure) for individuals with detected infections
            gamma_Q Rate of recovery (upon infection) for individuals with detected infections
            mu_Q    Rate of infection-related death for individuals with detected infections
            theta_E Rate of baseline testing for exposed individuals
            theta_I Rate of baseline testing for infectious individuals
            psi_E   Probability of positive test results for exposed individuals
            psi_I   Probability of positive test results for exposed individuals
            q       Probability of quarantined individuals interacting with others
            
            initE   Init number of exposed individuals       
            initI   Init number of infectious individuals      
            initQ_E Init number of detected infectious individuals
            initQ_I Init number of detected infectious individuals   
            initR   Init number of recovered individuals     
            initF   Init number of infection-related fatalities
                    (all remaining nodes initialized susceptible)   
    """

    def __init__(self, initN, beta, sigma, gamma, xi=0, mu_I=0, mu_0=0, nu=0, p=0,
                        beta_Q=None, sigma_Q=None, gamma_Q=None, mu_Q=None, 
                        theta_E=0, theta_I=0, psi_E=0, psi_I=0, q=0,
                        initE=0, initI=10, initQ_E=0, initQ_I=0, initR=0, initF=0):

        #~~~~~~~~~~~~~~~~~~~~~~~~~~~~~~~~~~~~~~~~
        # Model Parameters:
        #~~~~~~~~~~~~~~~~~~~~~~~~~~~~~~~~~~~~~~~~
        self.beta   = beta  
        self.sigma  = sigma 
        self.gamma  = gamma 
        self.xi     = xi    
        self.mu_I   = mu_I  
        self.mu_0   = mu_0  
        self.nu     = nu    
        self.p      = p     

        # Testing-related parameters:
        self.beta_Q   = beta_Q  if beta_Q is not None else self.beta
        self.sigma_Q  = sigma_Q if sigma_Q is not None else self.sigma
        self.gamma_Q  = gamma_Q if gamma_Q is not None else self.gamma
        self.mu_Q     = mu_Q    if mu_Q is not None else self.mu_I
        self.theta_E  = theta_E if theta_E is not None else self.theta_E
        self.theta_I  = theta_I if theta_I is not None else self.theta_I
        self.psi_E    = psi_E   if psi_E is not None else self.psi_E
        self.psi_I    = psi_I   if psi_I is not None else self.psi_I
        self.q        = q       if q is not None else self.q

        #~~~~~~~~~~~~~~~~~~~~~~~~~~~~~~~~~~~~~~~~
        # Initialize Timekeeping:
        #~~~~~~~~~~~~~~~~~~~~~~~~~~~~~~~~~~~~~~~~
        self.t       = 0
        self.tmax    = 0 # will be set when run() is called
        self.tseries = numpy.array([0])
        
        #~~~~~~~~~~~~~~~~~~~~~~~~~~~~~~~~~~~~~~~~
        # Initialize Counts of inidividuals with each state:
        #~~~~~~~~~~~~~~~~~~~~~~~~~~~~~~~~~~~~~~~~
        self.N          = numpy.array([int(initN)])
        self.numE       = numpy.array([int(initE)])
        self.numI       = numpy.array([int(initI)])
        self.numQ_E     = numpy.array([int(initQ_E)])
        self.numQ_I     = numpy.array([int(initQ_I)])
        self.numR       = numpy.array([int(initR)])
        self.numF       = numpy.array([int(initF)])
        self.numS       = numpy.array([self.N[-1] - self.numE[-1] - self.numI[-1] - self.numQ_E[-1] - self.numQ_I[-1] - self.numR[-1] - self.numF[-1]])
        assert(self.numS[0] >= 0), "The specified initial population size N must be greater than or equal to the initial compartment counts."


#^^^^^^^^^^^^^^^^^^^^^^^^^^^^^^^^^^^^^^^^
#^^^^^^^^^^^^^^^^^^^^^^^^^^^^^^^^^^^^^^^^

    @staticmethod
    def system_dfes(t, variables, beta, sigma, gamma, xi, mu_I, mu_0, nu,
                                  beta_Q, sigma_Q, gamma_Q, mu_Q, theta_E, theta_I, psi_E, psi_I, q):

        S, E, I, Q_E, Q_I, R, F = variables    # varibles is a list with compartment counts as elements 

        N   = S + E + I + Q_E + Q_I + R

        dS  = - (beta*S*I)/N - q*(beta_Q*S*Q_I)/N + xi*R + nu*N - mu_0*S

        dE  = (beta*S*I)/N + q*(beta_Q*S*Q_I)/N - sigma*E - theta_E*psi_E*E - mu_0*E

        dI  = sigma*E - gamma*I - mu_I*I - theta_I*psi_I*I - mu_0*I

        dDE = theta_E*psi_E*E - sigma_Q*Q_E - mu_0*Q_E

        dDI = theta_I*psi_I*I + sigma_Q*Q_E - gamma_Q*Q_I - mu_Q*Q_I - mu_0*Q_I

        dR  = gamma*I + gamma_Q*Q_I - xi*R - mu_0*R

        dF  = mu_I*I + mu_Q*Q_I

        return [dS, dE, dI, dDE, dDI, dR, dF]


#^^^^^^^^^^^^^^^^^^^^^^^^^^^^^^^^^^^^^^^^
#^^^^^^^^^^^^^^^^^^^^^^^^^^^^^^^^^^^^^^^^

    def run_epoch(self, runtime, dt=0.1):

        #~~~~~~~~~~~~~~~~~~~~~~~~~~~~~~~~~~~~~~~~
        # Create a list of times at which the ODE solver should output system values.
        # Append this list of times as the model's timeseries
        t_eval    = numpy.arange(start=self.t, stop=self.t+runtime, step=dt)

        # Define the range of time values for the integration:
        t_span          = [self.t, self.t+runtime]

        # Define the initial conditions as the system's current state:
        # (which will be the t=0 condition if this is the first run of this model, 
        # else where the last sim left off)

        init_cond       = [self.numS[-1], self.numE[-1], self.numI[-1], self.numQ_E[-1], self.numQ_I[-1], self.numR[-1], self.numF[-1]]

        #~~~~~~~~~~~~~~~~~~~~~~~~~~~~~~~~~~~~~~~~
        # Solve the system of differential eqns:
        #~~~~~~~~~~~~~~~~~~~~~~~~~~~~~~~~~~~~~~~~
        solution        = scipy.integrate.solve_ivp(lambda t, X: SEIRSModel.system_dfes(t, X, self.beta, self.sigma, self.gamma, self.xi, self.mu_I, self.mu_0, self.nu,
                                                                                            self.beta_Q, self.sigma_Q, self.gamma_Q, self.mu_Q, self.theta_E, self.theta_I, self.psi_E, self.psi_I, self.q
                                                                                        ), 
                                                        t_span=t_span, y0=init_cond, t_eval=t_eval
                                                   )

        #~~~~~~~~~~~~~~~~~~~~~~~~~~~~~~~~~~~~~~~~
        # Store the solution output as the model's time series and data series:
        #~~~~~~~~~~~~~~~~~~~~~~~~~~~~~~~~~~~~~~~~
        self.tseries    = numpy.append(self.tseries, solution['t'])
        self.numS       = numpy.append(self.numS, solution['y'][0])
        self.numE       = numpy.append(self.numE, solution['y'][1])
        self.numI       = numpy.append(self.numI, solution['y'][2])
        self.numQ_E       = numpy.append(self.numQ_E, solution['y'][3])
        self.numQ_I       = numpy.append(self.numQ_I, solution['y'][4])
        self.numR       = numpy.append(self.numR, solution['y'][5])
        self.numF       = numpy.append(self.numF, solution['y'][6])

        self.t = self.tseries[-1]


#^^^^^^^^^^^^^^^^^^^^^^^^^^^^^^^^^^^^^^^^
#^^^^^^^^^^^^^^^^^^^^^^^^^^^^^^^^^^^^^^^^

    def run(self, T, dt=0.1, checkpoints=None, verbose=False):

        if(T>0):
            self.tmax += T
        else:
            return False
        
        #~~~~~~~~~~~~~~~~~~~~~~~~~~~~~~~~~~~~~~~~
        # Pre-process checkpoint values:
        #~~~~~~~~~~~~~~~~~~~~~~~~~~~~~~~~~~~~~~~~
        if(checkpoints):
            numCheckpoints = len(checkpoints['t'])
            paramNames = ['beta', 'sigma', 'gamma', 'xi', 'mu_I', 'mu_0', 'nu',
                          'beta_Q', 'sigma_Q', 'gamma_Q', 'mu_Q',
                          'theta_E', 'theta_I', 'psi_E', 'psi_I', 'q']
            for param in paramNames:
                # For params that don't have given checkpoint values (or bad value given), 
                # set their checkpoint values to the value they have now for all checkpoints.
                if(param not in list(checkpoints.keys())
                    or not isinstance(checkpoints[param], (list, numpy.ndarray)) 
                    or len(checkpoints[param])!=numCheckpoints):
                    checkpoints[param] = [getattr(self, param)]*numCheckpoints

        #%%%%%%%%%%%%%%%%%%%%%%%%%%%%%%%%%%%%%%%%
        # Run the simulation loop:
        #%%%%%%%%%%%%%%%%%%%%%%%%%%%%%%%%%%%%%%%%
        if(not checkpoints):
            self.run_epoch(runtime=self.tmax, dt=dt)

            #~~~~~~~~~~~~~~~~~~~~~~~~~~~~~~~~~~~~~~~~

            print("t = %.2f" % self.t)
            if(verbose):
                print("\t S   = " + str(self.numS[-1]))
                print("\t E   = " + str(self.numE[-1]))
                print("\t I   = " + str(self.numI[-1]))
                print("\t Q_E = " + str(self.numQ_E[-1]))
                print("\t Q_I = " + str(self.numQ_I[-1]))
                print("\t R   = " + str(self.numR[-1]))
                print("\t F   = " + str(self.numF[-1]))
                    

        else: # checkpoints provided
            for checkpointIdx, checkpointTime in enumerate(checkpoints['t']):
                # Run the sim until the next checkpoint time:
                self.run_epoch(runtime=checkpointTime-self.t, dt=dt)
                # Having reached the checkpoint, update applicable parameters:
                print("[Checkpoint: Updating parameters]")
                for param in paramNames:
                    setattr(self, param, checkpoints[param][checkpointIdx])

                #~~~~~~~~~~~~~~~~~~~~~~~~~~~~~~~~~~~~~~~~

                print("t = %.2f" % self.t)
                if(verbose):
                    print("\t S   = " + str(self.numS[-1]))
                    print("\t E   = " + str(self.numE[-1]))
                    print("\t I   = " + str(self.numI[-1]))
                    print("\t Q_E = " + str(self.numQ_E[-1]))
                    print("\t Q_I = " + str(self.numQ_I[-1]))
                    print("\t R   = " + str(self.numR[-1]))
                    print("\t F   = " + str(self.numF[-1]))

            if(self.t < self.tmax):
                self.run_epoch(runtime=self.tmax-self.t, dt=dt)

        return True

#^^^^^^^^^^^^^^^^^^^^^^^^^^^^^^^^^^^^^^^^
#^^^^^^^^^^^^^^^^^^^^^^^^^^^^^^^^^^^^^^^^

    def total_num_susceptible(self, t_idx=None):
        if(t_idx is None):
            return (self.numS[:])            
        else:
            return (self.numS[t_idx])

#^^^^^^^^^^^^^^^^^^^^^^^^^^^^^^^^^^^^^^^^

    def total_num_infected(self, t_idx=None):
        if(t_idx is None):
            return (self.numE[:] + self.numI[:] + self.numQ_E[:] + self.numQ_I[:])  
        else:
            return (self.numE[t_idx] + self.numI[t_idx] + self.numQ_E[t_idx] + self.numQ_I[t_idx])  

#^^^^^^^^^^^^^^^^^^^^^^^^^^^^^^^^^^^^^^^^

    def total_num_isolated(self, t_idx=None):
        if(t_idx is None):
            return (self.numQ_E[:] + self.numQ_I[:])
        else:
            return (self.numQ_E[t_idx] + self.numQ_I[t_idx])

#^^^^^^^^^^^^^^^^^^^^^^^^^^^^^^^^^^^^^^^^

    def total_num_recovered(self, t_idx=None):
        if(t_idx is None):
            return (self.numR[:])            
        else:
            return (self.numR[t_idx]) 


#^^^^^^^^^^^^^^^^^^^^^^^^^^^^^^^^^^^^^^^^
#^^^^^^^^^^^^^^^^^^^^^^^^^^^^^^^^^^^^^^^^

    def plot(self, ax=None,  plot_S='line', plot_E='line', plot_I='line',plot_R='line', plot_F='line',
                            plot_Q_E='line', plot_Q_I='line', combine_Q=True,
                            color_S='tab:green', color_E='orange', color_I='crimson', color_R='tab:blue', color_F='black',
                            color_Q_E='mediumorchid', color_Q_I='mediumorchid', color_reference='#E0E0E0',
                            dashed_reference_results=None, dashed_reference_label='reference', 
                            shaded_reference_results=None, shaded_reference_label='reference', 
                            vlines=[], vline_colors=[], vline_styles=[], vline_labels=[],
                            ylim=None, xlim=None, legend=True, title=None, side_title=None, plot_percentages=True):

        import matplotlib.pyplot as pyplot

        #~~~~~~~~~~~~~~~~~~~~~~~~~~~~~~~~~~~~~~~~
        # Create an Axes object if None provided:
        #~~~~~~~~~~~~~~~~~~~~~~~~~~~~~~~~~~~~~~~~
        if(not ax):
            fig, ax = pyplot.subplots()

        #~~~~~~~~~~~~~~~~~~~~~~~~~~~~~~~~~~~~~~~~
        # Prepare data series to be plotted:
        #~~~~~~~~~~~~~~~~~~~~~~~~~~~~~~~~~~~~~~~~
        Fseries     = self.numF/self.N if plot_percentages else self.numF
        Eseries     = self.numE/self.N if plot_percentages else self.numE
        Dseries     = (self.numQ_E+self.numQ_I)/self.N if plot_percentages else (self.numQ_E+self.numQ_I)
        Q_Eseries   = self.numQ_E/self.N if plot_percentages else self.numQ_E
        Q_Iseries   = self.numQ_I/self.N if plot_percentages else self.numQ_I
        Iseries     = self.numI/self.N if plot_percentages else self.numI
        Rseries     = self.numR/self.N if plot_percentages else self.numR
        Sseries     = self.numS/self.N if plot_percentages else self.numS 

        #~~~~~~~~~~~~~~~~~~~~~~~~~~~~~~~~~~~~~~~~
        # Draw the reference data:      
        #~~~~~~~~~~~~~~~~~~~~~~~~~~~~~~~~~~~~~~~~
        if(dashed_reference_results):
            dashedReference_tseries  = dashed_reference_results.tseries[::int(self.N/100)]
            dashedReference_IDEstack = (dashed_reference_results.numI + dashed_reference_results.numQ_I + dashed_reference_results.numQ_E + dashed_reference_results.numE)[::int(self.N/100)] / (self.N if plot_percentages else 1)
            ax.plot(dashedReference_tseries, dashedReference_IDEstack, color='#E0E0E0', linestyle='--', label='$I+D+E$ ('+dashed_reference_label+')', zorder=0)
        if(shaded_reference_results):
            shadedReference_tseries  = shaded_reference_results.tseries
            shadedReference_IDEstack = (shaded_reference_results.numI + shaded_reference_results.numQ_I + shaded_reference_results.numQ_E + shaded_reference_results.numE) / (self.N if plot_percentages else 1)
            ax.fill_between(shaded_reference_results.tseries, shadedReference_IDEstack, 0, color='#EFEFEF', label='$I+D+E$ ('+shaded_reference_label+')', zorder=0)
            ax.plot(shaded_reference_results.tseries, shadedReference_IDEstack, color='#E0E0E0', zorder=1)

        #~~~~~~~~~~~~~~~~~~~~~~~~~~~~~~~~~~~~~~~~
        # Draw the stacked variables:
        #~~~~~~~~~~~~~~~~~~~~~~~~~~~~~~~~~~~~~~~~
        topstack = numpy.zeros_like(self.tseries)
        if(any(Fseries) and plot_F=='stacked'):
            ax.fill_between(numpy.ma.masked_where(Fseries<=0, self.tseries), numpy.ma.masked_where(Fseries<=0, topstack+Fseries), topstack, color=color_F, alpha=0.5, label='$F$', zorder=2)
            ax.plot(        numpy.ma.masked_where(Fseries<=0, self.tseries), numpy.ma.masked_where(Fseries<=0, topstack+Fseries),           color=color_F, zorder=3)
            topstack = topstack+Fseries
        if(any(Eseries) and plot_E=='stacked'):
            ax.fill_between(numpy.ma.masked_where(Eseries<=0, self.tseries), numpy.ma.masked_where(Eseries<=0, topstack+Eseries), topstack, color=color_E, alpha=0.5, label='$E$', zorder=2)
            ax.plot(        numpy.ma.masked_where(Eseries<=0, self.tseries), numpy.ma.masked_where(Eseries<=0, topstack+Eseries),           color=color_E, zorder=3)
            topstack = topstack+Eseries
        if(combine_Q and plot_Q_E=='stacked' and plot_Q_I=='stacked'):
            ax.fill_between(numpy.ma.masked_where(Dseries<=0, self.tseries), numpy.ma.masked_where(Dseries<=0, topstack+Dseries), topstack, color=color_Q_E, alpha=0.5, label='$Q_{all}$', zorder=2)
            ax.plot(        numpy.ma.masked_where(Dseries<=0, self.tseries), numpy.ma.masked_where(Dseries<=0, topstack+Dseries),           color=color_Q_E, zorder=3)
            topstack = topstack+Dseries
        else:
            if(any(Q_Eseries) and plot_Q_E=='stacked'):
                ax.fill_between(numpy.ma.masked_where(Q_Eseries<=0, self.tseries), numpy.ma.masked_where(Q_Eseries<=0, topstack+Q_Eseries), topstack, color=color_Q_E, alpha=0.5, label='$Q_E$', zorder=2)
                ax.plot(        numpy.ma.masked_where(Q_Eseries<=0, self.tseries), numpy.ma.masked_where(Q_Eseries<=0, topstack+Q_Eseries),           color=color_Q_E, zorder=3)
                topstack = topstack+Q_Eseries
            if(any(Q_Iseries) and plot_Q_I=='stacked'):
                ax.fill_between(numpy.ma.masked_where(Q_Iseries<=0, self.tseries), numpy.ma.masked_where(Q_Iseries<=0, topstack+Q_Iseries), topstack, color=color_Q_I, alpha=0.5, label='$Q_I$', zorder=2)
                ax.plot(        numpy.ma.masked_where(Q_Iseries<=0, self.tseries), numpy.ma.masked_where(Q_Iseries<=0, topstack+Q_Iseries),           color=color_Q_I, zorder=3)
                topstack = topstack+Q_Iseries
        if(any(Iseries) and plot_I=='stacked'):
            ax.fill_between(numpy.ma.masked_where(Iseries<=0, self.tseries), numpy.ma.masked_where(Iseries<=0, topstack+Iseries), topstack, color=color_I, alpha=0.5, label='$I$', zorder=2)
            ax.plot(        numpy.ma.masked_where(Iseries<=0, self.tseries), numpy.ma.masked_where(Iseries<=0, topstack+Iseries),           color=color_I, zorder=3)
            topstack = topstack+Iseries
        if(any(Rseries) and plot_R=='stacked'):
            ax.fill_between(numpy.ma.masked_where(Rseries<=0, self.tseries), numpy.ma.masked_where(Rseries<=0, topstack+Rseries), topstack, color=color_R, alpha=0.5, label='$R$', zorder=2)
            ax.plot(        numpy.ma.masked_where(Rseries<=0, self.tseries), numpy.ma.masked_where(Rseries<=0, topstack+Rseries),           color=color_R, zorder=3)
            topstack = topstack+Rseries
        if(any(Sseries) and plot_S=='stacked'):
            ax.fill_between(numpy.ma.masked_where(Sseries<=0, self.tseries), numpy.ma.masked_where(Sseries<=0, topstack+Sseries), topstack, color=color_S, alpha=0.5, label='$S$', zorder=2)
            ax.plot(        numpy.ma.masked_where(Sseries<=0, self.tseries), numpy.ma.masked_where(Sseries<=0, topstack+Sseries),           color=color_S, zorder=3)
            topstack = topstack+Sseries
        

        #~~~~~~~~~~~~~~~~~~~~~~~~~~~~~~~~~~~~~~~~
        # Draw the shaded variables:
        #~~~~~~~~~~~~~~~~~~~~~~~~~~~~~~~~~~~~~~~~
        if(any(Fseries) and plot_F=='shaded'):
            ax.fill_between(numpy.ma.masked_where(Fseries<=0, self.tseries), numpy.ma.masked_where(Fseries<=0, Fseries), 0, color=color_F, alpha=0.5, label='$F$', zorder=4)
            ax.plot(        numpy.ma.masked_where(Fseries<=0, self.tseries), numpy.ma.masked_where(Fseries<=0, Fseries),    color=color_F, zorder=5)
        if(any(Eseries) and plot_E=='shaded'):
            ax.fill_between(numpy.ma.masked_where(Eseries<=0, self.tseries), numpy.ma.masked_where(Eseries<=0, Eseries), 0, color=color_E, alpha=0.5, label='$E$', zorder=4)
            ax.plot(        numpy.ma.masked_where(Eseries<=0, self.tseries), numpy.ma.masked_where(Eseries<=0, Eseries),    color=color_E, zorder=5)
        if(combine_Q and (any(Dseries) and plot_Q_E=='shaded' and plot_Q_E=='shaded')):
            ax.fill_between(numpy.ma.masked_where(Dseries<=0, self.tseries), numpy.ma.masked_where(Dseries<=0, Dseries), 0, color=color_Q_E, alpha=0.5, label='$Q_{all}$', zorder=4)
            ax.plot(        numpy.ma.masked_where(Dseries<=0, self.tseries), numpy.ma.masked_where(Dseries<=0, Dseries),    color=color_Q_E, zorder=5)
        else:
            if(any(Q_Eseries) and plot_Q_E=='shaded'):
                ax.fill_between(numpy.ma.masked_where(Q_Eseries<=0, self.tseries), numpy.ma.masked_where(Q_Eseries<=0, Q_Eseries), 0, color=color_Q_E, alpha=0.5, label='$Q_E$', zorder=4)
                ax.plot(        numpy.ma.masked_where(Q_Eseries<=0, self.tseries), numpy.ma.masked_where(Q_Eseries<=0, Q_Eseries),    color=color_Q_E, zorder=5)
            if(any(Q_Iseries) and plot_Q_I=='shaded'):
                ax.fill_between(numpy.ma.masked_where(Q_Iseries<=0, self.tseries), numpy.ma.masked_where(Q_Iseries<=0, Q_Iseries), 0, color=color_Q_I, alpha=0.5, label='$Q_I$', zorder=4)
                ax.plot(        numpy.ma.masked_where(Q_Iseries<=0, self.tseries), numpy.ma.masked_where(Q_Iseries<=0, Q_Iseries),    color=color_Q_I, zorder=5)
        if(any(Iseries) and plot_I=='shaded'):
            ax.fill_between(numpy.ma.masked_where(Iseries<=0, self.tseries), numpy.ma.masked_where(Iseries<=0, Iseries), 0, color=color_I, alpha=0.5, label='$I$', zorder=4)
            ax.plot(        numpy.ma.masked_where(Iseries<=0, self.tseries), numpy.ma.masked_where(Iseries<=0, Iseries),    color=color_I, zorder=5)
        if(any(Sseries) and plot_S=='shaded'):
            ax.fill_between(numpy.ma.masked_where(Sseries<=0, self.tseries), numpy.ma.masked_where(Sseries<=0, Sseries), 0, color=color_S, alpha=0.5, label='$S$', zorder=4)
            ax.plot(        numpy.ma.masked_where(Sseries<=0, self.tseries), numpy.ma.masked_where(Sseries<=0, Sseries),    color=color_S, zorder=5)
        if(any(Rseries) and plot_R=='shaded'):
            ax.fill_between(numpy.ma.masked_where(Rseries<=0, self.tseries), numpy.ma.masked_where(Rseries<=0, Rseries), 0, color=color_R, alpha=0.5, label='$R$', zorder=4)
            ax.plot(        numpy.ma.masked_where(Rseries<=0, self.tseries), numpy.ma.masked_where(Rseries<=0, Rseries),    color=color_R, zorder=5)

        #~~~~~~~~~~~~~~~~~~~~~~~~~~~~~~~~~~~~~~~~
        # Draw the line variables:
        #~~~~~~~~~~~~~~~~~~~~~~~~~~~~~~~~~~~~~~~~        
        if(any(Fseries) and plot_F=='line'):
            ax.plot(numpy.ma.masked_where(Fseries<=0, self.tseries), numpy.ma.masked_where(Fseries<=0, Fseries), color=color_F, label='$F$', zorder=6)
        if(any(Eseries) and plot_E=='line'):
            ax.plot(numpy.ma.masked_where(Eseries<=0, self.tseries), numpy.ma.masked_where(Eseries<=0, Eseries), color=color_E, label='$E$', zorder=6)
        if(combine_Q and (any(Dseries) and plot_Q_E=='line' and plot_Q_E=='line')):
            ax.plot(numpy.ma.masked_where(Dseries<=0, self.tseries), numpy.ma.masked_where(Dseries<=0, Dseries), color=color_Q_E, label='$Q_{all}$', zorder=6)
        else:
            if(any(Q_Eseries) and plot_Q_E=='line'):
                ax.plot(numpy.ma.masked_where(Q_Eseries<=0, self.tseries), numpy.ma.masked_where(Q_Eseries<=0, Q_Eseries), color=color_Q_E, label='$Q_E$', zorder=6)
            if(any(Q_Iseries) and plot_Q_I=='line'):
                ax.plot(numpy.ma.masked_where(Q_Iseries<=0, self.tseries), numpy.ma.masked_where(Q_Iseries<=0, Q_Iseries), color=color_Q_I, label='$Q_I$', zorder=6)
        if(any(Iseries) and plot_I=='line'):
            ax.plot(numpy.ma.masked_where(Iseries<=0, self.tseries), numpy.ma.masked_where(Iseries<=0, Iseries), color=color_I, label='$I$', zorder=6)
        if(any(Sseries) and plot_S=='line'):
            ax.plot(numpy.ma.masked_where(Sseries<=0, self.tseries), numpy.ma.masked_where(Sseries<=0, Sseries), color=color_S, label='$S$', zorder=6)
        if(any(Rseries) and plot_R=='line'):
            ax.plot(numpy.ma.masked_where(Rseries<=0, self.tseries), numpy.ma.masked_where(Rseries<=0, Rseries), color=color_R, label='$R$', zorder=6)

        #~~~~~~~~~~~~~~~~~~~~~~~~~~~~~~~~~~~~~~~~
        # Draw the vertical line annotations:
        #~~~~~~~~~~~~~~~~~~~~~~~~~~~~~~~~~~~~~~~~
        if(len(vlines)>0 and len(vline_colors)==0):
            vline_colors = ['gray']*len(vlines)
        if(len(vlines)>0 and len(vline_labels)==0):
            vline_labels = [None]*len(vlines)
        if(len(vlines)>0 and len(vline_styles)==0):
            vline_styles = [':']*len(vlines)
        for vline_x, vline_color, vline_style, vline_label in zip(vlines, vline_colors, vline_styles, vline_labels):
            if(vline_x is not None):
                ax.axvline(x=vline_x, color=vline_color, linestyle=vline_style, alpha=1, label=vline_label)

        #~~~~~~~~~~~~~~~~~~~~~~~~~~~~~~~~~~~~~~~~
        # Draw the plot labels:
        #~~~~~~~~~~~~~~~~~~~~~~~~~~~~~~~~~~~~~~~~
        ax.set_xlabel('days')
        ax.set_ylabel('percent of population' if plot_percentages else 'number of individuals')
        ax.set_xlim(0, (max(self.tseries) if not xlim else xlim))
        ax.set_ylim(0, ylim)
        if(plot_percentages):
            ax.set_yticklabels(['{:,.0%}'.format(y) for y in ax.get_yticks()])
        if(legend):
            legend_handles, legend_labels = ax.get_legend_handles_labels()
            ax.legend(legend_handles[::-1], legend_labels[::-1], loc='upper right', facecolor='white', edgecolor='none', framealpha=0.9, prop={'size': 8})
        if(title):
            ax.set_title(title, size=12)
        if(side_title):
            ax.annotate(side_title, (0, 0.5), xytext=(-45, 0), ha='right', va='center',
                size=12, rotation=90, xycoords='axes fraction', textcoords='offset points')
       
        return ax


#^^^^^^^^^^^^^^^^^^^^^^^^^^^^^^^^^^^^^^^^
#^^^^^^^^^^^^^^^^^^^^^^^^^^^^^^^^^^^^^^^^

    def figure_basic(self, plot_S='line', plot_E='line', plot_I='line',plot_R='line', plot_F='line',
                        plot_Q_E='line', plot_Q_I='line', combine_Q=True,
                        color_S='tab:green', color_E='orange', color_I='crimson', color_R='tab:blue', color_F='black',
                        color_Q_E='mediumorchid', color_Q_I='mediumorchid', color_reference='#E0E0E0',
                        dashed_reference_results=None, dashed_reference_label='reference', 
                        shaded_reference_results=None, shaded_reference_label='reference', 
                        vlines=[], vline_colors=[], vline_styles=[], vline_labels=[],
                        ylim=None, xlim=None, legend=True, title=None, side_title=None, plot_percentages=True,
                        figsize=(12,8), use_seaborn=True, show=True):

        import matplotlib.pyplot as pyplot

        fig, ax = pyplot.subplots(figsize=figsize)

        if(use_seaborn):
            import seaborn
            seaborn.set_style('ticks')
            seaborn.despine()

        self.plot(ax=ax, plot_S=plot_S, plot_E=plot_E, plot_I=plot_I,plot_R=plot_R, plot_F=plot_F,
                        plot_Q_E=plot_Q_E, plot_Q_I=plot_Q_I, combine_Q=combine_Q,
                        color_S=color_S, color_E=color_E, color_I=color_I, color_R=color_R, color_F=color_F,
                        color_Q_E=color_Q_E, color_Q_I=color_Q_I, color_reference=color_reference,
                        dashed_reference_results=dashed_reference_results, dashed_reference_label=dashed_reference_label, 
                        shaded_reference_results=shaded_reference_results, shaded_reference_label=shaded_reference_label, 
                        vlines=vlines, vline_colors=vline_colors, vline_styles=vline_styles, vline_labels=vline_labels,
                        ylim=ylim, xlim=xlim, legend=legend, title=title, side_title=side_title, plot_percentages=plot_percentages)

        if(show):
            pyplot.show()

        return fig, ax


#^^^^^^^^^^^^^^^^^^^^^^^^^^^^^^^^^^^^^^^^
#^^^^^^^^^^^^^^^^^^^^^^^^^^^^^^^^^^^^^^^^

    def figure_infections(self, plot_S=False, plot_E='stacked', plot_I='stacked',plot_R=False, plot_F=False,
                            plot_Q_E='stacked', plot_Q_I='stacked', combine_Q=True,
                            color_S='tab:green', color_E='orange', color_I='crimson', color_R='tab:blue', color_F='black',
                            color_Q_E='mediumorchid', color_Q_I='mediumorchid', color_reference='#E0E0E0',
                            dashed_reference_results=None, dashed_reference_label='reference', 
                            shaded_reference_results=None, shaded_reference_label='reference', 
                            vlines=[], vline_colors=[], vline_styles=[], vline_labels=[],
                            ylim=None, xlim=None, legend=True, title=None, side_title=None, plot_percentages=True,
                            figsize=(12,8), use_seaborn=True, show=True):

        import matplotlib.pyplot as pyplot

        fig, ax = pyplot.subplots(figsize=figsize)

        if(use_seaborn):
            import seaborn
            seaborn.set_style('ticks')
            seaborn.despine()

        self.plot(ax=ax, plot_S=plot_S, plot_E=plot_E, plot_I=plot_I,plot_R=plot_R, plot_F=plot_F,
                        plot_Q_E=plot_Q_E, plot_Q_I=plot_Q_I, combine_Q=combine_Q,
                        color_S=color_S, color_E=color_E, color_I=color_I, color_R=color_R, color_F=color_F,
                        color_Q_E=color_Q_E, color_Q_I=color_Q_I, color_reference=color_reference,
                        dashed_reference_results=dashed_reference_results, dashed_reference_label=dashed_reference_label, 
                        shaded_reference_results=shaded_reference_results, shaded_reference_label=shaded_reference_label, 
                        vlines=vlines, vline_colors=vline_colors, vline_styles=vline_styles, vline_labels=vline_labels, 
                        ylim=ylim, xlim=xlim, legend=legend, title=title, side_title=side_title, plot_percentages=plot_percentages)

        if(show):
            pyplot.show()

        return fig, ax


#%%%%%%%%%%%%%%%%%%%%%%%%%%%%%%%%%%%%%%%%%%%%%%%%%%
#%%%%%%%%%%%%%%%%%%%%%%%%%%%%%%%%%%%%%%%%%%%%%%%%%%
#%%%%%%%%%%%%%%%%%%%%%%%%%%%%%%%%%%%%%%%%%%%%%%%%%%
#%%%%%%%%%%%%%%%%%%%%%%%%%%%%%%%%%%%%%%%%%%%%%%%%%%


class SEIRSNetworkModel():
    """
    A class to simulate the SEIRS Stochastic Network Model
    ======================================================
    Params: 
            G               Network adjacency matrix (numpy array) or Networkx graph object.
            beta            Rate of transmission (global interactions)
            beta_local      Rate(s) of transmission between adjacent individuals (optional)
            sigma           Rate of progression to infectious state (inverse of latent period)             
            gamma           Rate of recovery (inverse of symptomatic infectious period)           
            mu_I            Rate of infection-related death
            xi              Rate of re-susceptibility (upon recovery)
            mu_0            Rate of baseline death
            nu              Rate of baseline birth                        
            p               Probability of individuals interacting with global population            
            
            G_Q             Quarantine adjacency matrix (numpy array) or Networkx graph object.
            beta_Q          Rate of transmission for isolated individuals (global interactions)
            beta_Q_local    Rate(s) of transmission (exposure) for adjacent isolated individuals (optional)              
            sigma_Q         Rate of progression to infectious state for isolated individuals               
            gamma_Q         Rate of recovery for isolated individuals             
            mu_Q            Rate of infection-related death for isolated individuals
            q               Probability of isolated individuals interacting with global population
            isolation_time  Time to remain in isolation upon positive test, self-isolation, etc.

            theta_E         Rate of random testing for exposed individuals             
            theta_I         Rate of random testing for infectious individuals              
            phi_E           Rate of testing when a close contact has tested positive for exposed individuals               
            phi_I           Rate of testing when a close contact has tested positive for infectious  individuals                
            psi_E           Probability of positive test for exposed individuals               
            psi_I           Probability of positive test for infectious  individuals                
            
            initE           Initial number of exposed individuals
            initI           Initial number of infectious individuals
            initR           Initial number of recovered individuals     
            initF           Initial number of infection-related fatalities
            initQ_S         Initial number of isolated susceptible individuals
            initQ_E         Initial number of isolated exposed individuals
            initQ_I         Initial number of isolated infectious individuals
            initQ_R         Initial number of isolated recovered individuals
                            (all remaining nodes initialized susceptible)   
    """
    def __init__(self, G, beta, sigma, gamma,
                    mu_I=0, alpha=1.0, xi=0, mu_0=0, nu=0, f=0, p=0,  
                    beta_local=None, beta_pairwise_mode='infected', delta=None, delta_pairwise_mode=None,
                    G_Q=None, beta_Q=None, beta_Q_local=None, sigma_Q=None, gamma_Q=None, mu_Q=None, alpha_Q=None, delta_Q=None,
                    theta_E=0, theta_I=0, phi_E=0, phi_I=0, psi_E=1, psi_I=1, q=0, isolation_time=14,
                    initE=0, initI=0, initR=0, initF=0, initQ_E=0, initQ_I=0, 
                    transition_mode='exponential_rates', node_groups=None, store_Xseries=False, seed=None,
                    o=1, prevalence_ext=0):

        if(seed is not None):
            numpy.random.seed(seed)
            self.seed = seed

        #~~~~~~~~~~~~~~~~~~~~~~~~~~~~~~~~~~~~~~~~
        # Model Parameters:
        #~~~~~~~~~~~~~~~~~~~~~~~~~~~~~~~~~~~~~~~~
        self.parameters = { 'G':G, 'G_Q':G_Q,
                            'beta':beta, 'sigma':sigma, 'gamma':gamma, 'mu_I':mu_I, 
                            'xi':xi, 'mu_0':mu_0, 'nu':nu, 'f':f, 'p':p, 
                            'beta_local':beta_local, 'beta_pairwise_mode':beta_pairwise_mode,
                            'alpha':alpha, 'delta':delta, 'delta_pairwise_mode':delta_pairwise_mode,
                            'beta_Q':beta_Q, 'beta_Q_local':beta_Q_local, 'sigma_Q':sigma_Q, 'gamma_Q':gamma_Q, 'mu_Q':mu_Q,
                            'alpha_Q':alpha_Q, 'delta_Q':delta_Q,
                            'theta_E':theta_E, 'theta_I':theta_I, 'phi_E':phi_E, 'phi_I':phi_I, 'psi_E':psi_E, 'psi_I':psi_I, 
                            'q':q, 'isolation_time':isolation_time,
                            'initE':initE, 'initI':initI, 'initR':initR, 'initF':initF, 
                            'initQ_E':initQ_E, 'initQ_I':initQ_I }
        self.update_parameters()
        self.o = o
        self.prevalence_ext = prevalence_ext

        #~~~~~~~~~~~~~~~~~~~~~~~~~~~~~~~~~~~~~~~~
        # Each node can undergo 4-6 transitions (sans vitality/re-susceptibility returns to S state),
        # so there are ~numNodes*6 events/timesteps expected; initialize numNodes*6 timestep slots to start 
        # (will be expanded during run if needed for some reason)
        self.tseries    = numpy.zeros(6*self.numNodes)
        self.numS       = numpy.zeros(6*self.numNodes)
        self.numE       = numpy.zeros(6*self.numNodes)
        self.numI       = numpy.zeros(6*self.numNodes)
        self.numR       = numpy.zeros(6*self.numNodes)
        self.numF       = numpy.zeros(6*self.numNodes)
        self.numQ_E     = numpy.zeros(6*self.numNodes)
        self.numQ_I     = numpy.zeros(6*self.numNodes)
        self.N          = numpy.zeros(6*self.numNodes)

        #~~~~~~~~~~~~~~~~~~~~~~~~~~~~~~~~~~~~~~~~
        # Initialize Timekeeping:
        #~~~~~~~~~~~~~~~~~~~~~~~~~~~~~~~~~~~~~~~~
        self.t          = 0
        self.tmax       = 0 # will be set when run() is called
        self.tidx       = 0
        self.tseries[0] = 0

        # Vectors holding the time that each node has been in a given state or in isolation:
        self.timer_state     = numpy.zeros((self.numNodes,1))
        self.timer_isolation = numpy.zeros(self.numNodes)
        self.isolationTime   = isolation_time
        
        #~~~~~~~~~~~~~~~~~~~~~~~~~~~~~~~~~~~~~~~~
        # Initialize Counts of inidividuals with each state:
        #~~~~~~~~~~~~~~~~~~~~~~~~~~~~~~~~~~~~~~~~
        self.numE[0]        = int(initE)
        self.numI[0]        = int(initI)
        self.numR[0]        = int(initR)
        self.numF[0]        = int(initF)
        self.numQ_E[0]      = int(initQ_E)
        self.numQ_I[0]      = int(initQ_I)
        self.numS[0]        = (self.numNodes - self.numE[0] - self.numI[0] - self.numR[0] - self.numQ_E[0] - self.numQ_I[0] - self.numF[0])
        self.N[0]           = self.numNodes - self.numF[0]

        #~~~~~~~~~~~~~~~~~~~~~~~~~~~~~~~~~~~~~~~~
        # Node states:
        #~~~~~~~~~~~~~~~~~~~~~~~~~~~~~~~~~~~~~~~~
        self.S          = 1
        self.E          = 2
        self.I          = 3
        self.R          = 4
        self.F          = 5
        self.Q_E        = 6
        self.Q_I        = 7
        
        self.X = numpy.array( [self.S]*int(self.numS[0]) + [self.E]*int(self.numE[0]) + [self.I]*int(self.numI[0]) 
                               + [self.R]*int(self.numR[0]) + [self.F]*int(self.numF[0])
                               + [self.Q_E]*int(self.numQ_E[0]) + [self.Q_I]*int(self.numQ_I[0])
                            ).reshape((self.numNodes,1))
        numpy.random.shuffle(self.X)

        self.store_Xseries = store_Xseries
        if(store_Xseries):
            self.Xseries        = numpy.zeros(shape=(6*self.numNodes, self.numNodes), dtype='uint8')
            self.Xseries[0,:]   = self.X.T

        self.transitions =  { 
                                'StoE': {'currentState':self.S, 'newState':self.E},
                                'EtoI': {'currentState':self.E, 'newState':self.I},
                                'ItoR': {'currentState':self.I, 'newState':self.R},
                                'ItoF': {'currentState':self.I, 'newState':self.F},
                                'RtoS': {'currentState':self.R, 'newState':self.S},
                                'EtoQE': {'currentState':self.E, 'newState':self.Q_E},
                                'ItoQI': {'currentState':self.I, 'newState':self.Q_I},
                                'QEtoQI': {'currentState':self.Q_E, 'newState':self.Q_I},
                                'QItoR': {'currentState':self.Q_I, 'newState':self.R},
                                'QItoF': {'currentState':self.Q_I, 'newState':self.F},
                                '_toS': {'currentState':True, 'newState':self.S},
                            }

        self.transition_mode = transition_mode

        #~~~~~~~~~~~~~~~~~~~~~~~~~~~~~~~~~~~~~~~~
        # Initialize other node metadata:
        #~~~~~~~~~~~~~~~~~~~~~~~~~~~~~~~~~~~~~~~~
        self.tested      = numpy.array([False]*self.numNodes).reshape((self.numNodes,1))
        self.positive    = numpy.array([False]*self.numNodes).reshape((self.numNodes,1))
        self.numTested   = numpy.zeros(6*self.numNodes) 
        self.numPositive = numpy.zeros(6*self.numNodes) 

        self.testedInCurrentState = numpy.array([False]*self.numNodes).reshape((self.numNodes,1))

        self.infectionsLog = []

        #~~~~~~~~~~~~~~~~~~~~~~~~~~~~~~~~~~~~~~~~
        # Initialize node subgroup data series:
        #~~~~~~~~~~~~~~~~~~~~~~~~~~~~~~~~~~~~~~~~
        self.nodeGroupData = None
        if(node_groups):
            self.nodeGroupData = {}
            for groupName, nodeList in node_groups.items():
                self.nodeGroupData[groupName] = {'nodes':   numpy.array(nodeList),
                                                 'mask':    numpy.isin(range(self.numNodes), nodeList).reshape((self.numNodes,1))}
                self.nodeGroupData[groupName]['numS']           = numpy.zeros(6*self.numNodes)
                self.nodeGroupData[groupName]['numE']           = numpy.zeros(6*self.numNodes)
                self.nodeGroupData[groupName]['numI']           = numpy.zeros(6*self.numNodes)
                self.nodeGroupData[groupName]['numR']           = numpy.zeros(6*self.numNodes)
                self.nodeGroupData[groupName]['numF']           = numpy.zeros(6*self.numNodes)
                self.nodeGroupData[groupName]['numQ_E']         = numpy.zeros(6*self.numNodes)
                self.nodeGroupData[groupName]['numQ_I']         = numpy.zeros(6*self.numNodes)
                self.nodeGroupData[groupName]['N']              = numpy.zeros(6*self.numNodes)
                self.nodeGroupData[groupName]['numPositive']    = numpy.zeros(6*self.numNodes)
                self.nodeGroupData[groupName]['numTested']      = numpy.zeros(6*self.numNodes)
                self.nodeGroupData[groupName]['numS'][0]        = numpy.count_nonzero(self.nodeGroupData[groupName]['mask']*self.X==self.S)
                self.nodeGroupData[groupName]['numE'][0]        = numpy.count_nonzero(self.nodeGroupData[groupName]['mask']*self.X==self.E)
                self.nodeGroupData[groupName]['numI'][0]        = numpy.count_nonzero(self.nodeGroupData[groupName]['mask']*self.X==self.I)
                self.nodeGroupData[groupName]['numR'][0]        = numpy.count_nonzero(self.nodeGroupData[groupName]['mask']*self.X==self.R)
                self.nodeGroupData[groupName]['numF'][0]        = numpy.count_nonzero(self.nodeGroupData[groupName]['mask']*self.X==self.F)
                self.nodeGroupData[groupName]['numQ_E'][0]      = numpy.count_nonzero(self.nodeGroupData[groupName]['mask']*self.X==self.Q_E)
                self.nodeGroupData[groupName]['numQ_I'][0]      = numpy.count_nonzero(self.nodeGroupData[groupName]['mask']*self.X==self.Q_I)
                self.nodeGroupData[groupName]['N'][0]           = self.numNodes - self.numF[0]

         
#^^^^^^^^^^^^^^^^^^^^^^^^^^^^^^^^^^^^^^^^
#^^^^^^^^^^^^^^^^^^^^^^^^^^^^^^^^^^^^^^^^

    def update_parameters(self):

        #~~~~~~~~~~~~~~~~~~~~~~~~~~~~~~~~~~~~~~~~
        # Model graphs:
        #~~~~~~~~~~~~~~~~~~~~~~~~~~~~~~~~~~~~~~~~
        self.G = self.parameters['G']
        # Adjacency matrix:
        if type(self.G)==numpy.ndarray:
            self.A = scipy.sparse.csr_matrix(self.G)
        elif type(self.G)==networkx.classes.graph.Graph:
            self.A = networkx.adj_matrix(self.G) # adj_matrix gives scipy.sparse csr_matrix
        else:
            raise BaseException("Input an adjacency matrix or networkx object only.")
        self.numNodes   = int(self.A.shape[1])
        self.degree     = numpy.asarray(self.node_degrees(self.A)).astype(float)
        #----------------------------------------
        if(self.parameters['G_Q'] is None):
            self.G_Q = self.G # If no Q graph is provided, use G in its place
        else:
            self.G_Q = self.parameters['G_Q']
        # Quarantine Adjacency matrix:
        if type(self.G_Q)==numpy.ndarray:
            self.A_Q = scipy.sparse.csr_matrix(self.G_Q)
        elif type(self.G_Q)==networkx.classes.graph.Graph:
            self.A_Q = networkx.adj_matrix(self.G_Q) # adj_matrix gives scipy.sparse csr_matrix
        else:
            raise BaseException("Input an adjacency matrix or networkx object only.")
        self.numNodes_Q   = int(self.A_Q.shape[1])
        self.degree_Q     = numpy.asarray(self.node_degrees(self.A_Q)).astype(float)
        #----------------------------------------
        assert(self.numNodes == self.numNodes_Q), "The normal and quarantine adjacency graphs must be of the same size."

        #~~~~~~~~~~~~~~~~~~~~~~~~~~~~~~~~~~~~~~~~
        # Model parameters:
        #~~~~~~~~~~~~~~~~~~~~~~~~~~~~~~~~~~~~~~~~
        self.beta           = numpy.array(self.parameters['beta']).reshape((self.numNodes, 1))          if isinstance(self.parameters['beta'], (list, numpy.ndarray)) else numpy.full(fill_value=self.parameters['beta'], shape=(self.numNodes,1))
        self.sigma          = numpy.array(self.parameters['sigma']).reshape((self.numNodes, 1))         if isinstance(self.parameters['sigma'], (list, numpy.ndarray)) else numpy.full(fill_value=self.parameters['sigma'], shape=(self.numNodes,1))
        self.gamma          = numpy.array(self.parameters['gamma']).reshape((self.numNodes, 1))         if isinstance(self.parameters['gamma'], (list, numpy.ndarray)) else numpy.full(fill_value=self.parameters['gamma'], shape=(self.numNodes,1))
        self.mu_I           = numpy.array(self.parameters['mu_I']).reshape((self.numNodes, 1))          if isinstance(self.parameters['mu_I'], (list, numpy.ndarray)) else numpy.full(fill_value=self.parameters['mu_I'], shape=(self.numNodes,1))
        self.alpha          = numpy.array(self.parameters['alpha']).reshape((self.numNodes, 1))         if isinstance(self.parameters['alpha'], (list, numpy.ndarray)) else numpy.full(fill_value=self.parameters['alpha'], shape=(self.numNodes,1))
        self.xi             = numpy.array(self.parameters['xi']).reshape((self.numNodes, 1))            if isinstance(self.parameters['xi'], (list, numpy.ndarray)) else numpy.full(fill_value=self.parameters['xi'], shape=(self.numNodes,1))
        self.mu_0           = numpy.array(self.parameters['mu_0']).reshape((self.numNodes, 1))          if isinstance(self.parameters['mu_0'], (list, numpy.ndarray)) else numpy.full(fill_value=self.parameters['mu_0'], shape=(self.numNodes,1))
        self.nu             = numpy.array(self.parameters['nu']).reshape((self.numNodes, 1))            if isinstance(self.parameters['nu'], (list, numpy.ndarray)) else numpy.full(fill_value=self.parameters['nu'], shape=(self.numNodes,1))
        self.f              = numpy.array(self.parameters['f']).reshape((self.numNodes, 1))             if isinstance(self.parameters['f'], (list, numpy.ndarray)) else numpy.full(fill_value=self.parameters['f'], shape=(self.numNodes,1))
        self.p              = numpy.array(self.parameters['p']).reshape((self.numNodes, 1))             if isinstance(self.parameters['p'], (list, numpy.ndarray)) else numpy.full(fill_value=self.parameters['p'], shape=(self.numNodes,1))
        
        self.rand_f = numpy.random.rand(self.f.shape[0], self.f.shape[1])
       
        #----------------------------------------
        # Testing-related parameters:
        #----------------------------------------
        self.beta_Q         = (numpy.array(self.parameters['beta_Q']).reshape((self.numNodes, 1))       if isinstance(self.parameters['beta_Q'], (list, numpy.ndarray)) else numpy.full(fill_value=self.parameters['beta_Q'], shape=(self.numNodes,1))) if self.parameters['beta_Q'] is not None else self.beta
        self.sigma_Q        = (numpy.array(self.parameters['sigma_Q']).reshape((self.numNodes, 1))      if isinstance(self.parameters['sigma_Q'], (list, numpy.ndarray)) else numpy.full(fill_value=self.parameters['sigma_Q'], shape=(self.numNodes,1))) if self.parameters['sigma_Q'] is not None else self.sigma
        self.gamma_Q        = (numpy.array(self.parameters['gamma_Q']).reshape((self.numNodes, 1))  if isinstance(self.parameters['gamma_Q'], (list, numpy.ndarray)) else numpy.full(fill_value=self.parameters['gamma_Q'], shape=(self.numNodes,1))) if self.parameters['gamma_Q'] is not None else self.gamma
        self.mu_Q           = (numpy.array(self.parameters['mu_Q']).reshape((self.numNodes, 1))  if isinstance(self.parameters['mu_Q'], (list, numpy.ndarray)) else numpy.full(fill_value=self.parameters['mu_Q'], shape=(self.numNodes,1))) if self.parameters['mu_Q'] is not None else self.mu_I
        self.alpha_Q        = (numpy.array(self.parameters['alpha_Q']).reshape((self.numNodes, 1))      if isinstance(self.parameters['alpha_Q'], (list, numpy.ndarray)) else numpy.full(fill_value=self.parameters['alpha_Q'], shape=(self.numNodes,1))) if self.parameters['alpha_Q'] is not None else self.alpha
        self.theta_E        = numpy.array(self.parameters['theta_E']).reshape((self.numNodes, 1))       if isinstance(self.parameters['theta_E'], (list, numpy.ndarray)) else numpy.full(fill_value=self.parameters['theta_E'], shape=(self.numNodes,1))
        self.theta_I        = numpy.array(self.parameters['theta_I']).reshape((self.numNodes, 1))     if isinstance(self.parameters['theta_I'], (list, numpy.ndarray)) else numpy.full(fill_value=self.parameters['theta_I'], shape=(self.numNodes,1))
        self.phi_E          = numpy.array(self.parameters['phi_E']).reshape((self.numNodes, 1))         if isinstance(self.parameters['phi_E'], (list, numpy.ndarray)) else numpy.full(fill_value=self.parameters['phi_E'], shape=(self.numNodes,1))
        self.phi_I          = numpy.array(self.parameters['phi_I']).reshape((self.numNodes, 1))       if isinstance(self.parameters['phi_I'], (list, numpy.ndarray)) else numpy.full(fill_value=self.parameters['phi_I'], shape=(self.numNodes,1))
        self.psi_E          = numpy.array(self.parameters['psi_E']).reshape((self.numNodes, 1))           if isinstance(self.parameters['psi_E'], (list, numpy.ndarray)) else numpy.full(fill_value=self.parameters['psi_E'], shape=(self.numNodes,1))
        self.psi_I          = numpy.array(self.parameters['psi_I']).reshape((self.numNodes, 1))         if isinstance(self.parameters['psi_I'], (list, numpy.ndarray)) else numpy.full(fill_value=self.parameters['psi_I'], shape=(self.numNodes,1))
        self.q              = numpy.array(self.parameters['q']).reshape((self.numNodes, 1))             if isinstance(self.parameters['q'], (list, numpy.ndarray)) else numpy.full(fill_value=self.parameters['q'], shape=(self.numNodes,1))

        #----------------------------------------

        self.beta_pairwise_mode = self.parameters['beta_pairwise_mode']

        #----------------------------------------
        # Global transmission parameters:
        #----------------------------------------
        if(self.beta_pairwise_mode == 'infected' or self.beta_pairwise_mode is None):
            self.beta_global         = numpy.full_like(self.beta, fill_value=numpy.mean(self.beta))
            self.beta_Q_global       = numpy.full_like(self.beta_Q, fill_value=numpy.mean(self.beta_Q))
        elif(self.beta_pairwise_mode == 'infectee'):
            self.beta_global         = self.beta      
            self.beta_Q_global       = self.beta_Q    
        elif(self.beta_pairwise_mode == 'min'):
            self.beta_global         = numpy.minimum(self.beta, numpy.mean(beta)) 
            self.beta_Q_global       = numpy.minimum(self.beta_Q, numpy.mean(beta_Q)) 
        elif(self.beta_pairwise_mode == 'max'):
            self.beta_global         = numpy.maximum(self.beta, numpy.mean(beta)) 
            self.beta_Q_global       = numpy.maximum(self.beta_Q, numpy.mean(beta_Q)) 
        elif(self.beta_pairwise_mode == 'mean'):
            self.beta_global         = (self.beta + numpy.full_like(self.beta, fill_value=numpy.mean(self.beta)))/2
            self.beta_Q_global       = (self.beta_Q + numpy.full_like(self.beta_Q, fill_value=numpy.mean(self.beta_Q)))/2
            
        #----------------------------------------
        # Local transmission parameters:
        #----------------------------------------
        self.beta_local         = self.beta      if self.parameters['beta_local'] is None      else numpy.array(self.parameters['beta_local'])      if isinstance(self.parameters['beta_local'], (list, numpy.ndarray))      else numpy.full(fill_value=self.parameters['beta_local'], shape=(self.numNodes,1))
        self.beta_Q_local       = self.beta_Q    if self.parameters['beta_Q_local'] is None    else numpy.array(self.parameters['beta_Q_local'])    if isinstance(self.parameters['beta_Q_local'], (list, numpy.ndarray))    else numpy.full(fill_value=self.parameters['beta_Q_local'], shape=(self.numNodes,1))

        #----------------------------------------
        if(self.beta_local.ndim == 2 and self.beta_local.shape[0] == self.numNodes and self.beta_local.shape[1] == self.numNodes):
            self.A_beta_pairwise = self.beta_local
        elif((self.beta_local.ndim == 1 and self.beta_local.shape[0] == self.numNodes) or (self.beta_local.ndim == 2 and (self.beta_local.shape[0] == self.numNodes or self.beta_local.shape[1] == self.numNodes))):
            self.beta_local = self.beta_local.reshape((self.numNodes,1))
            # Pre-multiply beta values by the adjacency matrix ("transmission weight connections")
            A_beta_pairwise_byInfected = scipy.sparse.csr_matrix.multiply(self.A, self.beta_local.T).tocsr()
            A_beta_pairwise_byInfectee = scipy.sparse.csr_matrix.multiply(self.A, self.beta_local).tocsr()    
            #------------------------------
            # Compute the effective pairwise beta values as a function of the infected/infectee pair:
            if(self.beta_pairwise_mode == 'infected'):
                self.A_beta_pairwise = A_beta_pairwise_byInfected
            elif(self.beta_pairwise_mode == 'infectee'):
                self.A_beta_pairwise = A_beta_pairwise_byInfectee
            elif(self.beta_pairwise_mode == 'min'):
                self.A_beta_pairwise = scipy.sparse.csr_matrix.minimum(A_beta_pairwise_byInfected, A_beta_pairwise_byInfectee)
            elif(self.beta_pairwise_mode == 'max'):
                self.A_beta_pairwise = scipy.sparse.csr_matrix.maximum(A_beta_pairwise_byInfected, A_beta_pairwise_byInfectee)
            elif(self.beta_pairwise_mode == 'mean' or self.beta_pairwise_mode is None):
                self.A_beta_pairwise = (A_beta_pairwise_byInfected + A_beta_pairwise_byInfectee)/2
            else:
                print("Unrecognized beta_pairwise_mode value (support for 'infected', 'infectee', 'min', 'max', and 'mean').")
        else:
            print("Invalid values given for beta_local (expected 1xN list/array or NxN 2d array)")
        #----------------------------------------
        if(self.beta_Q_local.ndim == 2 and self.beta_Q_local.shape[0] == self.numNodes and self.beta_Q_local.shape[1] == self.numNodes):
            self.A_Q_beta_Q_pairwise = self.beta_Q_local
        elif((self.beta_Q_local.ndim == 1 and self.beta_Q_local.shape[0] == self.numNodes) or (self.beta_Q_local.ndim == 2 and (self.beta_Q_local.shape[0] == self.numNodes or self.beta_Q_local.shape[1] == self.numNodes))):
            self.beta_Q_local = self.beta_Q_local.reshape((self.numNodes,1))
            # Pre-multiply beta_Q values by the isolation adjacency matrix ("transmission weight connections")
            A_Q_beta_Q_pairwise_byInfected      = scipy.sparse.csr_matrix.multiply(self.A_Q, self.beta_Q_local.T).tocsr()
            A_Q_beta_Q_pairwise_byInfectee      = scipy.sparse.csr_matrix.multiply(self.A_Q, self.beta_Q_local).tocsr()
            #------------------------------
            # Compute the effective pairwise beta values as a function of the infected/infectee pair:
            if(self.beta_pairwise_mode == 'infected'):
                self.A_Q_beta_Q_pairwise = A_Q_beta_Q_pairwise_byInfected
            elif(self.beta_pairwise_mode == 'infectee'):
                self.A_Q_beta_Q_pairwise = A_Q_beta_Q_pairwise_byInfectee
            elif(self.beta_pairwise_mode == 'min'):
                self.A_Q_beta_Q_pairwise = scipy.sparse.csr_matrix.minimum(A_Q_beta_Q_pairwise_byInfected, A_Q_beta_Q_pairwise_byInfectee)
            elif(self.beta_pairwise_mode == 'max'):
                self.A_Q_beta_Q_pairwise = scipy.sparse.csr_matrix.maximum(A_Q_beta_Q_pairwise_byInfected, A_Q_beta_Q_pairwise_byInfectee)
            elif(self.beta_pairwise_mode == 'mean' or self.beta_pairwise_mode is None):
                self.A_Q_beta_Q_pairwise = (A_Q_beta_Q_pairwise_byInfected + A_Q_beta_Q_pairwise_byInfectee)/2
            else:
                print("Unrecognized beta_pairwise_mode value (support for 'infected', 'infectee', 'min', 'max', and 'mean').")
        else:
            print("Invalid values given for beta_Q_local (expected 1xN list/array or NxN 2d array)")
        #----------------------------------------

        #----------------------------------------
        # Degree-based transmission scaling parameters:
        #----------------------------------------
        self.delta_pairwise_mode = self.parameters['delta_pairwise_mode']
        with numpy.errstate(divide='ignore'): # ignore log(0) warning, then convert log(0) = -inf -> 0.0
            self.delta               = numpy.log(self.degree)/numpy.log(numpy.mean(self.degree))     if self.parameters['delta'] is None   else numpy.array(self.parameters['delta'])   if isinstance(self.parameters['delta'], (list, numpy.ndarray))   else numpy.full(fill_value=self.parameters['delta'], shape=(self.numNodes,1))
            self.delta_Q             = numpy.log(self.degree_Q)/numpy.log(numpy.mean(self.degree_Q)) if self.parameters['delta_Q'] is None else numpy.array(self.parameters['delta_Q']) if isinstance(self.parameters['delta_Q'], (list, numpy.ndarray)) else numpy.full(fill_value=self.parameters['delta_Q'], shape=(self.numNodes,1))
        self.delta[numpy.isneginf(self.delta)] = 0.0
        self.delta_Q[numpy.isneginf(self.delta_Q)] = 0.0
        #----------------------------------------
        if(self.delta.ndim == 2 and self.delta.shape[0] == self.numNodes and self.delta.shape[1] == self.numNodes):
            self.A_delta_pairwise = self.delta
        elif((self.delta.ndim == 1 and self.delta.shape[0] == self.numNodes) or (self.delta.ndim == 2 and (self.delta.shape[0] == self.numNodes or self.delta.shape[1] == self.numNodes))):
            self.delta = self.delta.reshape((self.numNodes,1))
            # Pre-multiply delta values by the adjacency matrix ("transmission weight connections")
            A_delta_pairwise_byInfected = scipy.sparse.csr_matrix.multiply(self.A, self.delta.T).tocsr()
            A_delta_pairwise_byInfectee = scipy.sparse.csr_matrix.multiply(self.A, self.delta).tocsr()    
            #------------------------------
            # Compute the effective pairwise delta values as a function of the infected/infectee pair:
            if(self.delta_pairwise_mode == 'infected'):
                self.A_delta_pairwise = A_delta_pairwise_byInfected
            elif(self.delta_pairwise_mode == 'infectee'):
                self.A_delta_pairwise = A_delta_pairwise_byInfectee
            elif(self.delta_pairwise_mode == 'min'):
                self.A_delta_pairwise = scipy.sparse.csr_matrix.minimum(A_delta_pairwise_byInfected, A_delta_pairwise_byInfectee)
            elif(self.delta_pairwise_mode == 'max'):
                self.A_delta_pairwise = scipy.sparse.csr_matrix.maximum(A_delta_pairwise_byInfected, A_delta_pairwise_byInfectee)
            elif(self.delta_pairwise_mode == 'mean'):
                self.A_delta_pairwise = (A_delta_pairwise_byInfected + A_delta_pairwise_byInfectee)/2
            elif(self.delta_pairwise_mode is None):
                self.A_delta_pairwise = self.A
            else:
                print("Unrecognized delta_pairwise_mode value (support for 'infected', 'infectee', 'min', 'max', and 'mean').")
        else:
            print("Invalid values given for delta (expected 1xN list/array or NxN 2d array)")
        #----------------------------------------
        if(self.delta_Q.ndim == 2 and self.delta_Q.shape[0] == self.numNodes and self.delta_Q.shape[1] == self.numNodes):
            self.A_Q_delta_Q_pairwise = self.delta_Q
        elif((self.delta_Q.ndim == 1 and self.delta_Q.shape[0] == self.numNodes) or (self.delta_Q.ndim == 2 and (self.delta_Q.shape[0] == self.numNodes or self.delta_Q.shape[1] == self.numNodes))):
            self.delta_Q = self.delta_Q.reshape((self.numNodes,1))
            # Pre-multiply delta_Q values by the isolation adjacency matrix ("transmission weight connections")
            A_Q_delta_Q_pairwise_byInfected      = scipy.sparse.csr_matrix.multiply(self.A_Q, self.delta_Q).tocsr()
            A_Q_delta_Q_pairwise_byInfectee      = scipy.sparse.csr_matrix.multiply(self.A_Q, self.delta_Q.T).tocsr()
            #------------------------------
            # Compute the effective pairwise delta values as a function of the infected/infectee pair:
            if(self.delta_pairwise_mode == 'infected'):
                self.A_Q_delta_Q_pairwise = A_Q_delta_Q_pairwise_byInfected
            elif(self.delta_pairwise_mode == 'infectee'):
                self.A_Q_delta_Q_pairwise = A_Q_delta_Q_pairwise_byInfectee
            elif(self.delta_pairwise_mode == 'min'):
                self.A_Q_delta_Q_pairwise = scipy.sparse.csr_matrix.minimum(A_Q_delta_Q_pairwise_byInfected, A_Q_delta_Q_pairwise_byInfectee)
            elif(self.delta_pairwise_mode == 'max'):
                self.A_Q_delta_Q_pairwise = scipy.sparse.csr_matrix.maximum(A_Q_delta_Q_pairwise_byInfected, A_Q_delta_Q_pairwise_byInfectee)
            elif(self.delta_pairwise_mode == 'mean'):
                self.A_Q_delta_Q_pairwise = (A_Q_delta_Q_pairwise_byInfected + A_Q_delta_Q_pairwise_byInfectee)/2
            elif(self.delta_pairwise_mode is None):
                self.A_Q_delta_Q_pairwise = self.A
            else:
                print("Unrecognized delta_pairwise_mode value (support for 'infected', 'infectee', 'min', 'max', and 'mean').")
        else:
            print("Invalid values given for delta_Q (expected 1xN list/array or NxN 2d array)")

        #----------------------------------------
        # Pre-calculate the pairwise delta*beta values:
        #----------------------------------------
        self.A_deltabeta          = scipy.sparse.csr_matrix.multiply(self.A_delta_pairwise, self.A_beta_pairwise)
        self.A_Q_deltabeta_Q      = scipy.sparse.csr_matrix.multiply(self.A_Q_delta_Q_pairwise, self.A_Q_beta_Q_pairwise)
    

#^^^^^^^^^^^^^^^^^^^^^^^^^^^^^^^^^^^^^^^^
#^^^^^^^^^^^^^^^^^^^^^^^^^^^^^^^^^^^^^^^^

    def node_degrees(self, Amat):
        return Amat.sum(axis=0).reshape(self.numNodes,1)   # sums of adj matrix cols


#^^^^^^^^^^^^^^^^^^^^^^^^^^^^^^^^^^^^^^^^
#^^^^^^^^^^^^^^^^^^^^^^^^^^^^^^^^^^^^^^^^

    def total_num_susceptible(self, t_idx=None):
        if(t_idx is None):
            return (self.numS[:])            
        else:
            return (self.numS[t_idx])

#^^^^^^^^^^^^^^^^^^^^^^^^^^^^^^^^^^^^^^^^

    def total_num_infected(self, t_idx=None):
        if(t_idx is None):
            return (self.numE[:] + self.numI[:] + self.numQ_E[:] + self.numQ_I[:])            
        else:
            return (self.numE[t_idx] + self.numI[t_idx] + self.numQ_E[t_idx] + self.numQ_I[t_idx])

#^^^^^^^^^^^^^^^^^^^^^^^^^^^^^^^^^^^^^^^^

    def total_num_isolated(self, t_idx=None):
        if(t_idx is None):
            return (self.numQ_E[:] + self.numQ_I[:])            
        else:
            return (self.numQ_E[t_idx] + self.numQ_I[t_idx])            

#^^^^^^^^^^^^^^^^^^^^^^^^^^^^^^^^^^^^^^^^

    def total_num_tested(self, t_idx=None):
        if(t_idx is None):
            return (self.numTested[:])            
        else:
            return (self.numTested[t_idx])

#^^^^^^^^^^^^^^^^^^^^^^^^^^^^^^^^^^^^^^^^

    def total_num_positive(self, t_idx=None):
        if(t_idx is None):
            return (self.numPositive[:])            
        else:
            return (self.numPositive[t_idx])

#^^^^^^^^^^^^^^^^^^^^^^^^^^^^^^^^^^^^^^^^

    def total_num_recovered(self, t_idx=None):
        if(t_idx is None):
            return (self.numR[:])            
        else:
            return (self.numR[t_idx])


#^^^^^^^^^^^^^^^^^^^^^^^^^^^^^^^^^^^^^^^^
#^^^^^^^^^^^^^^^^^^^^^^^^^^^^^^^^^^^^^^^^
        
    def calc_propensities(self):

        #~~~~~~~~~~~~~~~~~~~~~~~~~~~~~~~~~~~~~~~~
        # Pre-calculate matrix multiplication terms that may be used in multiple propensity calculations,
        # and check to see if their computation is necessary before doing the multiplication
        #------------------------------------

        self.transmissionTerms_I = numpy.zeros(shape=(self.numNodes,1))      
        if(numpy.any(self.numI[self.tidx])):
            self.transmissionTerms_I = numpy.asarray(scipy.sparse.csr_matrix.dot(self.A_deltabeta, self.X==self.I))

        #------------------------------------

        self.transmissionTerms_Q = numpy.zeros(shape=(self.numNodes,1))
        if(numpy.any(self.numQ_I[self.tidx])):
            self.transmissionTerms_Q = numpy.asarray(scipy.sparse.csr_matrix.dot(self.A_Q_deltabeta_Q, self.X==self.Q_I))

        #------------------------------------

        numContacts_Q = numpy.zeros(shape=(self.numNodes,1))
        if(numpy.any(self.positive) and (numpy.any(self.phi_E) or numpy.any(self.phi_I))):
            numContacts_Q = numpy.asarray(scipy.sparse.csr_matrix.dot(self.A, ((self.positive)&(self.X!=self.R)&(self.X!=self.F))))

        #~~~~~~~~~~~~~~~~~~~~~~~~~~~~~~~~~~~~~~~~

        propensities_StoE       = (self.alpha *
                                    (self.o*(self.beta_global*self.prevalence_ext) + 
                                        (1-self.o)*(self.p*((self.beta_global*self.numI[self.tidx] + self.q*self.beta_Q_global*self.numQ_I[self.tidx])/self.N[self.tidx])
                                     + (1-self.p)*(numpy.divide(self.transmissionTerms_I, self.degree, out=numpy.zeros_like(self.degree), where=self.degree!=0)
                                                  +numpy.divide(self.transmissionTerms_Q, self.degree_Q, out=numpy.zeros_like(self.degree_Q), where=self.degree_Q!=0)))
                                  ))*(self.X==self.S)

        #~~~~~~~~~~~~~~~~~~~~~~~~~~~~~~~~~~~~~~~~

        if(self.transition_mode == 'time_in_state'):

            propensities_EtoI     = 1e5 * ((self.X==self.E) & numpy.greater(self.timer_state, 1/self.sigma))

            propensities_ItoR     = 1e5 * ((self.X==self.I) & numpy.greater(self.timer_state, 1/self.gamma) & numpy.greater_equal(self.rand_f, self.f))

            propensities_ItoF     = 1e5 * ((self.X==self.I) & numpy.greater(self.timer_state, 1/self.mu_I) & numpy.less(self.rand_f, self.f))

            propensities_EtoQE    = numpy.zeros_like(propensities_StoE)

            propensities_ItoQI    = numpy.zeros_like(propensities_StoE)

            propensities_QEtoQI   = 1e5 * ((self.X==self.Q_E) & numpy.greater(self.timer_state, 1/self.sigma_Q))

            propensities_QItoR    = 1e5 * ((self.X==self.Q_I) & numpy.greater(self.timer_state, 1/self.gamma_Q) & numpy.greater_equal(self.rand_f, self.f))

            propensities_QItoF    = 1e5 * ((self.X==self.Q_I) & numpy.greater(self.timer_state, 1/self.mu_Q) & numpy.less(self.rand_f, self.f))

            propensities_RtoS     = 1e5 * ((self.X==self.R) & numpy.greater(self.timer_state, 1/self.xi))

            propensities__toS     = 1e5 * ((self.X!=self.F) & numpy.greater(self.timer_state, 1/self.nu))            

        #~~~~~~~~~~~~~~~~~~~~~~~~~~~~~~~~~~~~~~~~

        else: # exponential_rates

            propensities_EtoI        = self.sigma * (self.X==self.E)

            propensities_ItoR        = self.gamma * ((self.X==self.I) & (numpy.greater_equal(self.rand_f, self.f)))

            propensities_ItoF        = self.mu_I * ((self.X==self.I) & (numpy.less(self.rand_f, self.f)))

            propensities_EtoQE       = (self.theta_E + self.phi_E*numContacts_Q)*self.psi_E * (self.X==self.E)

            propensities_ItoQI       = (self.theta_I + self.phi_I*numContacts_Q)*self.psi_I * (self.X==self.I)

            propensities_QEtoQI      = self.sigma_Q * (self.X==self.Q_E)

            propensities_QItoR       = self.gamma_Q * ((self.X==self.Q_I) & (numpy.greater_equal(self.rand_f, self.f)))

            propensities_QItoF       = self.mu_Q * ((self.X==self.Q_I) & (numpy.less(self.rand_f, self.f)))

            propensities_RtoS        = self.xi * (self.X==self.R)

            propensities__toS        = self.nu * (self.X!=self.F)

        


            

        #~~~~~~~~~~~~~~~~~~~~~~~~~~~~~~~~~~~~~~~~

        propensities = numpy.hstack([propensities_StoE, propensities_EtoI, 
                                     propensities_ItoR, propensities_ItoF, 
                                     propensities_EtoQE, propensities_ItoQI, propensities_QEtoQI, 
                                     propensities_QItoR, propensities_QItoF,
                                     propensities_RtoS, propensities__toS])

        columns = ['StoE', 'EtoI', 'ItoR', 'ItoF', 'EtoQE', 'ItoQI', 'QEtoQI', 'QItoR', 'QItoF', 'RtoS', '_toS']

        return propensities, columns


#^^^^^^^^^^^^^^^^^^^^^^^^^^^^^^^^^^^^^^^^
#^^^^^^^^^^^^^^^^^^^^^^^^^^^^^^^^^^^^^^^^

    def set_isolation(self, node, isolate):
        # Move this node in/out of the appropriate isolation state:
        if(isolate == True):
            if(self.X[node] == self.E):
                self.X[node] = self.Q_E
                self.timer_state[node] = 0
            elif(self.X[node] == self.I):
                self.X[node] = self.Q_I
                self.timer_state[node] = 0
        elif(isolate == False):
            if(self.X[node] == self.Q_E):
                self.X[node] = self.E
                self.timer_state[node] = 0
            elif(self.X[node] == self.Q_I):
                self.X[node] = self.I
                self.timer_state[node] = 0
        # Reset the isolation timer:
        self.timer_isolation[node] = 0

#^^^^^^^^^^^^^^^^^^^^^^^^^^^^^^^^^^^^^^^^

    def set_tested(self, node, tested):
        self.tested[node] = tested
        self.testedInCurrentState[node] = tested

#^^^^^^^^^^^^^^^^^^^^^^^^^^^^^^^^^^^^^^^^

    def set_positive(self, node, positive):
        self.positive[node] = positive

#^^^^^^^^^^^^^^^^^^^^^^^^^^^^^^^^^^^^^^^^

    def introduce_exposures(self, num_new_exposures):
        exposedNodes = numpy.random.choice(range(self.numNodes), size=num_new_exposures, replace=False)
        for exposedNode in exposedNodes:
            if(self.X[exposedNode]==self.S):
                self.X[exposedNode] = self.E


#^^^^^^^^^^^^^^^^^^^^^^^^^^^^^^^^^^^^^^^^
#^^^^^^^^^^^^^^^^^^^^^^^^^^^^^^^^^^^^^^^^   

    def increase_data_series_length(self):
        self.tseries     = numpy.pad(self.tseries, [(0, 6*self.numNodes)], mode='constant', constant_values=0)
        self.numS        = numpy.pad(self.numS, [(0, 6*self.numNodes)], mode='constant', constant_values=0)
        self.numE        = numpy.pad(self.numE, [(0, 6*self.numNodes)], mode='constant', constant_values=0)
        self.numI        = numpy.pad(self.numI, [(0, 6*self.numNodes)], mode='constant', constant_values=0)
        self.numR        = numpy.pad(self.numR, [(0, 6*self.numNodes)], mode='constant', constant_values=0)
        self.numF        = numpy.pad(self.numF, [(0, 6*self.numNodes)], mode='constant', constant_values=0)
        self.numQ_E      = numpy.pad(self.numQ_E, [(0, 6*self.numNodes)], mode='constant', constant_values=0)
        self.numQ_I      = numpy.pad(self.numQ_I, [(0, 6*self.numNodes)], mode='constant', constant_values=0)
        self.N           = numpy.pad(self.N, [(0, 6*self.numNodes)], mode='constant', constant_values=0)
        self.numTested   = numpy.pad(self.numTested, [(0, 6*self.numNodes)], mode='constant', constant_values=0)
        self.numPositive = numpy.pad(self.numPositive, [(0, 6*self.numNodes)], mode='constant', constant_values=0)

        if(self.store_Xseries):
            self.Xseries = numpy.pad(self.Xseries, [(0, 6*self.numNodes), (0,0)], mode='constant', constant_values=0)

        if(self.nodeGroupData):
            for groupName in self.nodeGroupData:
                self.nodeGroupData[groupName]['numS']        = numpy.pad(self.nodeGroupData[groupName]['numS'], [(0, 6*self.numNodes)], mode='constant', constant_values=0)
                self.nodeGroupData[groupName]['numE']        = numpy.pad(self.nodeGroupData[groupName]['numE'], [(0, 6*self.numNodes)], mode='constant', constant_values=0)
                self.nodeGroupData[groupName]['numI']        = numpy.pad(self.nodeGroupData[groupName]['numI'], [(0, 6*self.numNodes)], mode='constant', constant_values=0)
                self.nodeGroupData[groupName]['numR']        = numpy.pad(self.nodeGroupData[groupName]['numR'], [(0, 6*self.numNodes)], mode='constant', constant_values=0)
                self.nodeGroupData[groupName]['numF']        = numpy.pad(self.nodeGroupData[groupName]['numF'], [(0, 6*self.numNodes)], mode='constant', constant_values=0)
                self.nodeGroupData[groupName]['numQ_E']      = numpy.pad(self.nodeGroupData[groupName]['numQ_E'], [(0, 6*self.numNodes)], mode='constant', constant_values=0)
                self.nodeGroupData[groupName]['numQ_I']      = numpy.pad(self.nodeGroupData[groupName]['numQ_I'], [(0, 6*self.numNodes)], mode='constant', constant_values=0)
                self.nodeGroupData[groupName]['N']           = numpy.pad(self.nodeGroupData[groupName]['N'], [(0, 6*self.numNodes)], mode='constant', constant_values=0)
                self.nodeGroupData[groupName]['numTested']   = numpy.pad(self.nodeGroupData[groupName]['numTested'], [(0, 6*self.numNodes)], mode='constant', constant_values=0)
                self.nodeGroupData[groupName]['numPositive'] = numpy.pad(self.nodeGroupData[groupName]['numPositive'], [(0, 6*self.numNodes)], mode='constant', constant_values=0)

        return None

#^^^^^^^^^^^^^^^^^^^^^^^^^^^^^^^^^^^^^^^^ 

    def finalize_data_series(self):
        self.tseries     = numpy.array(self.tseries, dtype=float)[:self.tidx+1]
        self.numS        = numpy.array(self.numS, dtype=float)[:self.tidx+1]
        self.numE        = numpy.array(self.numE, dtype=float)[:self.tidx+1]
        self.numI        = numpy.array(self.numI, dtype=float)[:self.tidx+1]
        self.numR        = numpy.array(self.numR, dtype=float)[:self.tidx+1]
        self.numF        = numpy.array(self.numF, dtype=float)[:self.tidx+1]
        self.numQ_E      = numpy.array(self.numQ_E, dtype=float)[:self.tidx+1]
        self.numQ_I      = numpy.array(self.numQ_I, dtype=float)[:self.tidx+1]
        self.N           = numpy.array(self.N, dtype=float)[:self.tidx+1]
        self.numTested   = numpy.array(self.numTested, dtype=float)[:self.tidx+1]
        self.numPositive = numpy.array(self.numPositive, dtype=float)[:self.tidx+1]

        if(self.store_Xseries):
            self.Xseries = self.Xseries[:self.tidx+1, :]

        if(self.nodeGroupData):
            for groupName in self.nodeGroupData:
                self.nodeGroupData[groupName]['numS']        = numpy.array(self.nodeGroupData[groupName]['numS'], dtype=float)[:self.tidx+1]
                self.nodeGroupData[groupName]['numE']        = numpy.array(self.nodeGroupData[groupName]['numE'], dtype=float)[:self.tidx+1]
                self.nodeGroupData[groupName]['numI']        = numpy.array(self.nodeGroupData[groupName]['numI'], dtype=float)[:self.tidx+1]
                self.nodeGroupData[groupName]['numR']        = numpy.array(self.nodeGroupData[groupName]['numR'], dtype=float)[:self.tidx+1]
                self.nodeGroupData[groupName]['numF']        = numpy.array(self.nodeGroupData[groupName]['numF'], dtype=float)[:self.tidx+1]
                self.nodeGroupData[groupName]['numQ_E']      = numpy.array(self.nodeGroupData[groupName]['numQ_E'], dtype=float)[:self.tidx+1]
                self.nodeGroupData[groupName]['numQ_I']      = numpy.array(self.nodeGroupData[groupName]['numQ_I'], dtype=float)[:self.tidx+1]
                self.nodeGroupData[groupName]['N']           = numpy.array(self.nodeGroupData[groupName]['N'], dtype=float)[:self.tidx+1]
                self.nodeGroupData[groupName]['numTested']   = numpy.array(self.nodeGroupData[groupName]['numTested'], dtype=float)[:self.tidx+1]
                self.nodeGroupData[groupName]['numPositive'] = numpy.array(self.nodeGroupData[groupName]['numPositive'], dtype=float)[:self.tidx+1]

        return None

#^^^^^^^^^^^^^^^^^^^^^^^^^^^^^^^^^^^^^^^^
#^^^^^^^^^^^^^^^^^^^^^^^^^^^^^^^^^^^^^^^^     

    def run_iteration(self):

        if(self.tidx >= len(self.tseries)-1):
            # Room has run out in the timeseries storage arrays; double the size of these arrays:
            self.increase_data_series_length()

        #~~~~~~~~~~~~~~~~~~~~~~~~~~~~~~~~~~~~~~~~
        # Generate 2 random numbers uniformly distributed in (0,1)
        #~~~~~~~~~~~~~~~~~~~~~~~~~~~~~~~~~~~~~~~~
        r1 = numpy.random.rand()
        r2 = numpy.random.rand()

        #~~~~~~~~~~~~~~~~~~~~~~~~~~~~~~~~~~~~~~~~
        # Calculate propensities
        #~~~~~~~~~~~~~~~~~~~~~~~~~~~~~~~~~~~~~~~~
        propensities, transitionTypes = self.calc_propensities()

        if(propensities.sum() > 0):

            #~~~~~~~~~~~~~~~~~~~~~~~~~~~~~~~~~~~~~~~~
            # Calculate alpha
            #~~~~~~~~~~~~~~~~~~~~~~~~~~~~~~~~~~~~~~~~
            propensities_flat   = propensities.ravel(order='F')
            cumsum              = propensities_flat.cumsum()
            alpha               = propensities_flat.sum()

            #~~~~~~~~~~~~~~~~~~~~~~~~~~~~~~~~~~~~~~~~
            # Compute the time until the next event takes place
            #~~~~~~~~~~~~~~~~~~~~~~~~~~~~~~~~~~~~~~~~
            tau = (1/alpha)*numpy.log(float(1/r1))
            self.t += tau
            self.timer_state += tau

            #~~~~~~~~~~~~~~~~~~~~~~~~~~~~~~~~~~~~~~~~
            # Compute which event takes place
            #~~~~~~~~~~~~~~~~~~~~~~~~~~~~~~~~~~~~~~~~
            transitionIdx   = numpy.searchsorted(cumsum,r2*alpha)
            transitionNode  = transitionIdx % self.numNodes
            transitionType  = transitionTypes[ int(transitionIdx/self.numNodes) ]

            #~~~~~~~~~~~~~~~~~~~~~~~~~~~~~~~~~~~~~~~~
            # Perform updates triggered by rate propensities:
            #~~~~~~~~~~~~~~~~~~~~~~~~~~~~~~~~~~~~~~~~
            assert(self.X[transitionNode] == self.transitions[transitionType]['currentState'] and self.X[transitionNode]!=self.F), "Assertion error: Node "+str(transitionNode)+" has unexpected current state "+str(self.X[transitionNode])+" given the intended transition of "+str(transitionType)+"."
            self.X[transitionNode] = self.transitions[transitionType]['newState']

            self.testedInCurrentState[transitionNode] = False

            self.timer_state[transitionNode] = 0.0

            #~~~~~~~~~~~~~~~~~~~~~~~~~~~~~~~~~~~~~~~~

            # Save information about infection events when they occur:
            if(transitionType == 'StoE'):
                transitionNode_GNbrs  = list(self.G[transitionNode].keys())
                transitionNode_GQNbrs = list(self.G_Q[transitionNode].keys())
                self.infectionsLog.append({ 't':                            self.t,
                                            'infected_node':                transitionNode,
                                            'infection_type':               transitionType,
                                            'infected_node_degree':         self.degree[transitionNode],
                                            'local_contact_nodes':          transitionNode_GNbrs,
                                            'local_contact_node_states':    self.X[transitionNode_GNbrs].flatten(),
                                            'isolation_contact_nodes':      transitionNode_GQNbrs,
                                            'isolation_contact_node_states':self.X[transitionNode_GQNbrs].flatten() })

            #~~~~~~~~~~~~~~~~~~~~~~~~~~~~~~~~~~~~~~~~

            if(transitionType in ['EtoQE', 'ItoQI']):
                self.set_positive(node=transitionNode, positive=True)

        #~~~~~~~~~~~~~~~~~~~~~~~~~~~~~~~~~~~~~~~~

        else:

            tau = 0.01
            self.t += tau
            self.timer_state += tau

        #~~~~~~~~~~~~~~~~~~~~~~~~~~~~~~~~~~~~~~~~

        self.tidx += 1
        
        self.tseries[self.tidx]     = self.t
        self.numS[self.tidx]        = numpy.clip(numpy.count_nonzero(self.X==self.S), a_min=0, a_max=self.numNodes)
        self.numE[self.tidx]        = numpy.clip(numpy.count_nonzero(self.X==self.E), a_min=0, a_max=self.numNodes)
        self.numI[self.tidx]        = numpy.clip(numpy.count_nonzero(self.X==self.I), a_min=0, a_max=self.numNodes)
        self.numR[self.tidx]        = numpy.clip(numpy.count_nonzero(self.X==self.R), a_min=0, a_max=self.numNodes)
        self.numF[self.tidx]        = numpy.clip(numpy.count_nonzero(self.X==self.F), a_min=0, a_max=self.numNodes)
        self.numQ_E[self.tidx]      = numpy.clip(numpy.count_nonzero(self.X==self.Q_E), a_min=0, a_max=self.numNodes)
        self.numQ_I[self.tidx]      = numpy.clip(numpy.count_nonzero(self.X==self.Q_I), a_min=0, a_max=self.numNodes)
        self.numTested[self.tidx]   = numpy.clip(numpy.count_nonzero(self.tested), a_min=0, a_max=self.numNodes)
        self.numPositive[self.tidx] = numpy.clip(numpy.count_nonzero(self.positive), a_min=0, a_max=self.numNodes)
        
        self.N[self.tidx]           = numpy.clip((self.numNodes - self.numF[self.tidx]), a_min=0, a_max=self.numNodes)

        #~~~~~~~~~~~~~~~~~~~~~~~~~~~~~~~~~~~~~~~~
        # Update testing and isolation statuses
        #~~~~~~~~~~~~~~~~~~~~~~~~~~~~~~~~~~~~~~~~

        isolatedNodes = numpy.argwhere((self.X==self.Q_E)|(self.X==self.Q_I))[:,0].flatten()
        self.timer_isolation[isolatedNodes] = self.timer_isolation[isolatedNodes] + tau

        nodesExitingIsolation = numpy.argwhere((self.timer_isolation >= self.isolationTime) & (self.positive[:,0]==False))
        for isoNode in nodesExitingIsolation:
            self.set_isolation(node=isoNode, isolate=False)

        #~~~~~~~~~~~~~~~~~~~~~~~~~~~~~~~~~~~~~~~~
        # Store system states
        #~~~~~~~~~~~~~~~~~~~~~~~~~~~~~~~~~~~~~~~~
        if(self.store_Xseries):
            self.Xseries[self.tidx,:] = self.X.T

        if(self.nodeGroupData):
            for groupName in self.nodeGroupData:
                self.nodeGroupData[groupName]['numS'][self.tidx]        = numpy.count_nonzero(self.nodeGroupData[groupName]['mask']*self.X==self.S)
                self.nodeGroupData[groupName]['numE'][self.tidx]        = numpy.count_nonzero(self.nodeGroupData[groupName]['mask']*self.X==self.E)
                self.nodeGroupData[groupName]['numI'][self.tidx]        = numpy.count_nonzero(self.nodeGroupData[groupName]['mask']*self.X==self.I)
                self.nodeGroupData[groupName]['numR'][self.tidx]        = numpy.count_nonzero(self.nodeGroupData[groupName]['mask']*self.X==self.R)
                self.nodeGroupData[groupName]['numF'][self.tidx]        = numpy.count_nonzero(self.nodeGroupData[groupName]['mask']*self.X==self.F)
                self.nodeGroupData[groupName]['numQ_E'][self.tidx]      = numpy.count_nonzero(self.nodeGroupData[groupName]['mask']*self.X==self.Q_E)
                self.nodeGroupData[groupName]['numQ_I'][self.tidx]      = numpy.count_nonzero(self.nodeGroupData[groupName]['mask']*self.X==self.Q_I)
                self.nodeGroupData[groupName]['N'][self.tidx]           = numpy.clip((self.nodeGroupData[groupName]['numS'][0] + self.nodeGroupData[groupName]['numE'][0] + self.nodeGroupData[groupName]['numI'][0] + self.nodeGroupData[groupName]['numQ_E'][0] + self.nodeGroupData[groupName]['numQ_I'][0] + self.nodeGroupData[groupName]['numR'][0]), a_min=0, a_max=self.numNodes)
                self.nodeGroupData[groupName]['numTested'][self.tidx]   = numpy.count_nonzero(self.nodeGroupData[groupName]['mask']*self.tested)
                self.nodeGroupData[groupName]['numPositive'][self.tidx] = numpy.count_nonzero(self.nodeGroupData[groupName]['mask']*self.positive)

        #~~~~~~~~~~~~~~~~~~~~~~~~~~~~~~~~~~~~~~~~
        # Terminate if tmax reached or num infections is 0:
        #~~~~~~~~~~~~~~~~~~~~~~~~~~~~~~~~~~~~~~~~
        if(self.t >= self.tmax or (self.total_num_infected(self.tidx) < 1 and self.total_num_isolated(self.tidx) < 1)):
            self.finalize_data_series()
            return False

        #~~~~~~~~~~~~~~~~~~~~~~~~~~~~~~~~~~~~~~~~

        return True


#^^^^^^^^^^^^^^^^^^^^^^^^^^^^^^^^^^^^^^^^
#^^^^^^^^^^^^^^^^^^^^^^^^^^^^^^^^^^^^^^^^

    def run(self, T, checkpoints=None, print_interval=10, verbose='t'):
        if(T>0):
            self.tmax += T
        else:
            return False

        #~~~~~~~~~~~~~~~~~~~~~~~~~~~~~~~~~~~~~~~~
        # Pre-process checkpoint values:
        #~~~~~~~~~~~~~~~~~~~~~~~~~~~~~~~~~~~~~~~~
        if(checkpoints):
            numCheckpoints = len(checkpoints['t'])
            for chkpt_param, chkpt_values in checkpoints.items():
                assert(isinstance(chkpt_values, (list, numpy.ndarray)) and len(chkpt_values)==numCheckpoints), "Expecting a list of values with length equal to number of checkpoint times ("+str(numCheckpoints)+") for each checkpoint parameter."
            checkpointIdx  = numpy.searchsorted(checkpoints['t'], self.t) # Finds 1st index in list greater than given val
            if(checkpointIdx >= numCheckpoints):
                # We are out of checkpoints, stop checking them:
                checkpoints = None 
            else:
                checkpointTime = checkpoints['t'][checkpointIdx]

        #%%%%%%%%%%%%%%%%%%%%%%%%%%%%%%%%%%%%%%%%
        # Run the simulation loop:
        #%%%%%%%%%%%%%%%%%%%%%%%%%%%%%%%%%%%%%%%%
        print_reset = True
        running     = True
        while running:

            running = self.run_iteration()

            #~~~~~~~~~~~~~~~~~~~~~~~~~~~~~~~~~~~~~~~~
            # Handle checkpoints if applicable:
            if(checkpoints):
                if(self.t >= checkpointTime):
                    if(verbose is not False):
                        print("[Checkpoint: Updating parameters]")
                    # A checkpoint has been reached, update param values:
                    for param in list(self.parameters.keys()):
                        if(param in list(checkpoints.keys())):
                            self.parameters.update({param: checkpoints[param][checkpointIdx]})
                    # Update parameter data structures and scenario flags:
                    self.update_parameters()
                    # Update the next checkpoint time:
                    checkpointIdx  = numpy.searchsorted(checkpoints['t'], self.t) # Finds 1st index in list greater than given val
                    if(checkpointIdx >= numCheckpoints):
                        # We are out of checkpoints, stop checking them:
                        checkpoints = None 
                    else:
                        checkpointTime = checkpoints['t'][checkpointIdx]
            #~~~~~~~~~~~~~~~~~~~~~~~~~~~~~~~~~~~~~~~~

            #~~~~~~~~~~~~~~~~~~~~~~~~~~~~~~~~~~~~~~~~

            if(print_interval):
                if(print_reset and (int(self.t) % print_interval == 0)):
                    if(verbose=="t"):
                        print("t = %.2f" % self.t)
                    if(verbose==True):
                        print("t = %.2f" % self.t)
                        print("\t S      = " + str(self.numS[self.tidx]))
                        print("\t E      = " + str(self.numE[self.tidx]))
                        print("\t I  = " + str(self.numI[self.tidx]))
                        print("\t R      = " + str(self.numR[self.tidx]))
                        print("\t F      = " + str(self.numF[self.tidx]))
                        print("\t Q_E    = " + str(self.numQ_E[self.tidx]))
                        print("\t Q_I  = " + str(self.numQ_I[self.tidx]))
                    print_reset = False
                elif(not print_reset and (int(self.t) % 10 != 0)):
                    print_reset = True

        return True


#^^^^^^^^^^^^^^^^^^^^^^^^^^^^^^^^^^^^^^^^
#^^^^^^^^^^^^^^^^^^^^^^^^^^^^^^^^^^^^^^^^

    def plot(self, ax=None,  plot_S='line', plot_E='line', plot_I='line',plot_R='line', plot_F='line',
                            plot_Q_E='line', plot_Q_I='line', combine_D=True,
                            color_S='tab:green', color_E='orange', color_I='crimson', color_R='tab:blue', color_F='black',
                            color_Q_E='mediumorchid', color_Q_I='mediumorchid', color_reference='#E0E0E0',
                            dashed_reference_results=None, dashed_reference_label='reference', 
                            shaded_reference_results=None, shaded_reference_label='reference', 
                            vlines=[], vline_colors=[], vline_styles=[], vline_labels=[],
                            ylim=None, xlim=None, legend=True, title=None, side_title=None, plot_percentages=True):

        import matplotlib.pyplot as pyplot

        #~~~~~~~~~~~~~~~~~~~~~~~~~~~~~~~~~~~~~~~~
        # Create an Axes object if None provided:
        #~~~~~~~~~~~~~~~~~~~~~~~~~~~~~~~~~~~~~~~~
        if(not ax):
            fig, ax = pyplot.subplots()

        #~~~~~~~~~~~~~~~~~~~~~~~~~~~~~~~~~~~~~~~~
        # Prepare data series to be plotted:
        #~~~~~~~~~~~~~~~~~~~~~~~~~~~~~~~~~~~~~~~~
        Fseries     = self.numF/self.numNodes if plot_percentages else self.numF
        Eseries     = self.numE/self.numNodes if plot_percentages else self.numE
        Dseries     = (self.numQ_E+self.numQ_I)/self.numNodes if plot_percentages else (self.numQ_E+self.numQ_I)
        Q_Eseries   = self.numQ_E/self.numNodes if plot_percentages else self.numQ_E
        Q_Iseries   = self.numQ_I/self.numNodes if plot_percentages else self.numQ_I
        Iseries     = self.numI/self.numNodes if plot_percentages else self.numI
        Rseries     = self.numR/self.numNodes if plot_percentages else self.numR
        Sseries     = self.numS/self.numNodes if plot_percentages else self.numS 

        #~~~~~~~~~~~~~~~~~~~~~~~~~~~~~~~~~~~~~~~~
        # Draw the reference data:      
        #~~~~~~~~~~~~~~~~~~~~~~~~~~~~~~~~~~~~~~~~
        if(dashed_reference_results):
            dashedReference_tseries  = dashed_reference_results.tseries[::int(self.numNodes/100)]
            dashedReference_IDEstack = (dashed_reference_results.numI + dashed_reference_results.numQ_I + dashed_reference_results.numQ_E + dashed_reference_results.numE)[::int(self.numNodes/100)] / (self.numNodes if plot_percentages else 1)
            ax.plot(dashedReference_tseries, dashedReference_IDEstack, color='#E0E0E0', linestyle='--', label='$I+D+E$ ('+dashed_reference_label+')', zorder=0)
        if(shaded_reference_results):
            shadedReference_tseries  = shaded_reference_results.tseries
            shadedReference_IDEstack = (shaded_reference_results.numI + shaded_reference_results.numQ_I + shaded_reference_results.numQ_E + shaded_reference_results.numE) / (self.numNodes if plot_percentages else 1)
            ax.fill_between(shaded_reference_results.tseries, shadedReference_IDEstack, 0, color='#EFEFEF', label='$I+D+E$ ('+shaded_reference_label+')', zorder=0)
            ax.plot(shaded_reference_results.tseries, shadedReference_IDEstack, color='#E0E0E0', zorder=1)

        #~~~~~~~~~~~~~~~~~~~~~~~~~~~~~~~~~~~~~~~~
        # Draw the stacked variables:
        #~~~~~~~~~~~~~~~~~~~~~~~~~~~~~~~~~~~~~~~~
        topstack = numpy.zeros_like(self.tseries)
        if(any(Fseries) and plot_F=='stacked'):
            ax.fill_between(numpy.ma.masked_where(Fseries<=0, self.tseries), numpy.ma.masked_where(Fseries<=0, topstack+Fseries), topstack, color=color_F, alpha=0.5, label='$F$', zorder=2)
            ax.plot(        numpy.ma.masked_where(Fseries<=0, self.tseries), numpy.ma.masked_where(Fseries<=0, topstack+Fseries),           color=color_F, zorder=3)
            topstack = topstack+Fseries
        if(any(Eseries) and plot_E=='stacked'):
            ax.fill_between(numpy.ma.masked_where(Eseries<=0, self.tseries), numpy.ma.masked_where(Eseries<=0, topstack+Eseries), topstack, color=color_E, alpha=0.5, label='$E$', zorder=2)
            ax.plot(        numpy.ma.masked_where(Eseries<=0, self.tseries), numpy.ma.masked_where(Eseries<=0, topstack+Eseries),           color=color_E, zorder=3)
            topstack = topstack+Eseries
        if(combine_D and plot_Q_E=='stacked' and plot_Q_I=='stacked'):
            ax.fill_between(numpy.ma.masked_where(Dseries<=0, self.tseries), numpy.ma.masked_where(Dseries<=0, topstack+Dseries), topstack, color=color_Q_E, alpha=0.5, label='$Q_{all}$', zorder=2)
            ax.plot(        numpy.ma.masked_where(Dseries<=0, self.tseries), numpy.ma.masked_where(Dseries<=0, topstack+Dseries),           color=color_Q_E, zorder=3)
            topstack = topstack+Dseries
        else:
            if(any(Q_Eseries) and plot_Q_E=='stacked'):
                ax.fill_between(numpy.ma.masked_where(Q_Eseries<=0, self.tseries), numpy.ma.masked_where(Q_Eseries<=0, topstack+Q_Eseries), topstack, color=color_Q_E, alpha=0.5, label='$Q_E$', zorder=2)
                ax.plot(        numpy.ma.masked_where(Q_Eseries<=0, self.tseries), numpy.ma.masked_where(Q_Eseries<=0, topstack+Q_Eseries),           color=color_Q_E, zorder=3)
                topstack = topstack+Q_Eseries
            if(any(Q_Iseries) and plot_Q_I=='stacked'):
                ax.fill_between(numpy.ma.masked_where(Q_Iseries<=0, self.tseries), numpy.ma.masked_where(Q_Iseries<=0, topstack+Q_Iseries), topstack, color=color_Q_I, alpha=0.5, label='$Q_I$', zorder=2)
                ax.plot(        numpy.ma.masked_where(Q_Iseries<=0, self.tseries), numpy.ma.masked_where(Q_Iseries<=0, topstack+Q_Iseries),           color=color_Q_I, zorder=3)
                topstack = topstack+Q_Iseries
        if(any(Iseries) and plot_I=='stacked'):
            ax.fill_between(numpy.ma.masked_where(Iseries<=0, self.tseries), numpy.ma.masked_where(Iseries<=0, topstack+Iseries), topstack, color=color_I, alpha=0.5, label='$I$', zorder=2)
            ax.plot(        numpy.ma.masked_where(Iseries<=0, self.tseries), numpy.ma.masked_where(Iseries<=0, topstack+Iseries),           color=color_I, zorder=3)
            topstack = topstack+Iseries
        if(any(Rseries) and plot_R=='stacked'):
            ax.fill_between(numpy.ma.masked_where(Rseries<=0, self.tseries), numpy.ma.masked_where(Rseries<=0, topstack+Rseries), topstack, color=color_R, alpha=0.5, label='$R$', zorder=2)
            ax.plot(        numpy.ma.masked_where(Rseries<=0, self.tseries), numpy.ma.masked_where(Rseries<=0, topstack+Rseries),           color=color_R, zorder=3)
            topstack = topstack+Rseries
        if(any(Sseries) and plot_S=='stacked'):
            ax.fill_between(numpy.ma.masked_where(Sseries<=0, self.tseries), numpy.ma.masked_where(Sseries<=0, topstack+Sseries), topstack, color=color_S, alpha=0.5, label='$S$', zorder=2)
            ax.plot(        numpy.ma.masked_where(Sseries<=0, self.tseries), numpy.ma.masked_where(Sseries<=0, topstack+Sseries),           color=color_S, zorder=3)
            topstack = topstack+Sseries
        

        #~~~~~~~~~~~~~~~~~~~~~~~~~~~~~~~~~~~~~~~~
        # Draw the shaded variables:
        #~~~~~~~~~~~~~~~~~~~~~~~~~~~~~~~~~~~~~~~~
        if(any(Fseries) and plot_F=='shaded'):
            ax.fill_between(numpy.ma.masked_where(Fseries<=0, self.tseries), numpy.ma.masked_where(Fseries<=0, Fseries), 0, color=color_F, alpha=0.5, label='$F$', zorder=4)
            ax.plot(        numpy.ma.masked_where(Fseries<=0, self.tseries), numpy.ma.masked_where(Fseries<=0, Fseries),    color=color_F, zorder=5)
        if(any(Eseries) and plot_E=='shaded'):
            ax.fill_between(numpy.ma.masked_where(Eseries<=0, self.tseries), numpy.ma.masked_where(Eseries<=0, Eseries), 0, color=color_E, alpha=0.5, label='$E$', zorder=4)
            ax.plot(        numpy.ma.masked_where(Eseries<=0, self.tseries), numpy.ma.masked_where(Eseries<=0, Eseries),    color=color_E, zorder=5)
        if(combine_D and (any(Dseries) and plot_Q_E=='shaded' and plot_Q_I=='shaded')):
            ax.fill_between(numpy.ma.masked_where(Dseries<=0, self.tseries), numpy.ma.masked_where(Dseries<=0, Dseries), 0, color=color_Q_E, alpha=0.5, label='$Q_{all}$', zorder=4)
            ax.plot(        numpy.ma.masked_where(Dseries<=0, self.tseries), numpy.ma.masked_where(Dseries<=0, Dseries),    color=color_Q_E, zorder=5)
        else:
            if(any(Q_Eseries) and plot_Q_E=='shaded'):
                ax.fill_between(numpy.ma.masked_where(Q_Eseries<=0, self.tseries), numpy.ma.masked_where(Q_Eseries<=0, Q_Eseries), 0, color=color_Q_E, alpha=0.5, label='$Q_E$', zorder=4)
                ax.plot(        numpy.ma.masked_where(Q_Eseries<=0, self.tseries), numpy.ma.masked_where(Q_Eseries<=0, Q_Eseries),    color=color_Q_E, zorder=5)
            if(any(Q_Iseries) and plot_Q_I=='shaded'):
                ax.fill_between(numpy.ma.masked_where(Q_Iseries<=0, self.tseries), numpy.ma.masked_where(Q_Iseries<=0, Q_Iseries), 0, color=color_Q_I, alpha=0.5, label='$Q_I$', zorder=4)
                ax.plot(        numpy.ma.masked_where(Q_Iseries<=0, self.tseries), numpy.ma.masked_where(Q_Iseries<=0, Q_Iseries),    color=color_Q_I, zorder=5)
        if(any(Iseries) and plot_I=='shaded'):
            ax.fill_between(numpy.ma.masked_where(Iseries<=0, self.tseries), numpy.ma.masked_where(Iseries<=0, Iseries), 0, color=color_I, alpha=0.5, label='$I$', zorder=4)
            ax.plot(        numpy.ma.masked_where(Iseries<=0, self.tseries), numpy.ma.masked_where(Iseries<=0, Iseries),    color=color_I, zorder=5)
        if(any(Sseries) and plot_S=='shaded'):
            ax.fill_between(numpy.ma.masked_where(Sseries<=0, self.tseries), numpy.ma.masked_where(Sseries<=0, Sseries), 0, color=color_S, alpha=0.5, label='$S$', zorder=4)
            ax.plot(        numpy.ma.masked_where(Sseries<=0, self.tseries), numpy.ma.masked_where(Sseries<=0, Sseries),    color=color_S, zorder=5)
        if(any(Rseries) and plot_R=='shaded'):
            ax.fill_between(numpy.ma.masked_where(Rseries<=0, self.tseries), numpy.ma.masked_where(Rseries<=0, Rseries), 0, color=color_R, alpha=0.5, label='$R$', zorder=4)
            ax.plot(        numpy.ma.masked_where(Rseries<=0, self.tseries), numpy.ma.masked_where(Rseries<=0, Rseries),    color=color_R, zorder=5)

        #~~~~~~~~~~~~~~~~~~~~~~~~~~~~~~~~~~~~~~~~
        # Draw the line variables:
        #~~~~~~~~~~~~~~~~~~~~~~~~~~~~~~~~~~~~~~~~        
        if(any(Fseries) and plot_F=='line'):
            ax.plot(numpy.ma.masked_where(Fseries<=0, self.tseries), numpy.ma.masked_where(Fseries<=0, Fseries), color=color_F, label='$F$', zorder=6)
        if(any(Eseries) and plot_E=='line'):
            ax.plot(numpy.ma.masked_where(Eseries<=0, self.tseries), numpy.ma.masked_where(Eseries<=0, Eseries), color=color_E, label='$E$', zorder=6)
        if(combine_D and (any(Dseries) and plot_Q_E=='line' and plot_Q_I=='line')):
            ax.plot(numpy.ma.masked_where(Dseries<=0, self.tseries), numpy.ma.masked_where(Dseries<=0, Dseries), color=color_Q_E, label='$Q_{all}$', zorder=6)
        else:
            if(any(Q_Eseries) and plot_Q_E=='line'):
                ax.plot(numpy.ma.masked_where(Q_Eseries<=0, self.tseries), numpy.ma.masked_where(Q_Eseries<=0, Q_Eseries), color=color_Q_E, label='$Q_E$', zorder=6)
            if(any(Q_Iseries) and plot_Q_I=='line'):
                ax.plot(numpy.ma.masked_where(Q_Iseries<=0, self.tseries), numpy.ma.masked_where(Q_Iseries<=0, Q_Iseries), color=color_Q_I, label='$Q_I$', zorder=6)
        if(any(Iseries) and plot_I=='line'):
            ax.plot(numpy.ma.masked_where(Iseries<=0, self.tseries), numpy.ma.masked_where(Iseries<=0, Iseries), color=color_I, label='$I$', zorder=6)
        if(any(Sseries) and plot_S=='line'):
            ax.plot(numpy.ma.masked_where(Sseries<=0, self.tseries), numpy.ma.masked_where(Sseries<=0, Sseries), color=color_S, label='$S$', zorder=6)
        if(any(Rseries) and plot_R=='line'):
            ax.plot(numpy.ma.masked_where(Rseries<=0, self.tseries), numpy.ma.masked_where(Rseries<=0, Rseries), color=color_R, label='$R$', zorder=6)

        #~~~~~~~~~~~~~~~~~~~~~~~~~~~~~~~~~~~~~~~~
        # Draw the vertical line annotations:
        #~~~~~~~~~~~~~~~~~~~~~~~~~~~~~~~~~~~~~~~~
        if(len(vlines)>0 and len(vline_colors)==0):
            vline_colors = ['gray']*len(vlines)
        if(len(vlines)>0 and len(vline_labels)==0):
            vline_labels = [None]*len(vlines)
        if(len(vlines)>0 and len(vline_styles)==0):
            vline_styles = [':']*len(vlines)
        for vline_x, vline_color, vline_style, vline_label in zip(vlines, vline_colors, vline_styles, vline_labels):
            if(vline_x is not None):
                ax.axvline(x=vline_x, color=vline_color, linestyle=vline_style, alpha=1, label=vline_label)

        #~~~~~~~~~~~~~~~~~~~~~~~~~~~~~~~~~~~~~~~~
        # Draw the plot labels:
        #~~~~~~~~~~~~~~~~~~~~~~~~~~~~~~~~~~~~~~~~
        ax.set_xlabel('days')
        ax.set_ylabel('percent of population' if plot_percentages else 'number of individuals')
        ax.set_xlim(0, (max(self.tseries) if not xlim else xlim))
        ax.set_ylim(0, ylim)
        if(plot_percentages):
            ax.set_yticklabels(['{:,.0%}'.format(y) for y in ax.get_yticks()])
        if(legend):
            legend_handles, legend_labels = ax.get_legend_handles_labels()
            ax.legend(legend_handles[::-1], legend_labels[::-1], loc='upper right', facecolor='white', edgecolor='none', framealpha=0.9, prop={'size': 8})
        if(title):
            ax.set_title(title, size=12)
        if(side_title):
            ax.annotate(side_title, (0, 0.5), xytext=(-45, 0), ha='right', va='center',
                size=12, rotation=90, xycoords='axes fraction', textcoords='offset points')
       
        return ax


#^^^^^^^^^^^^^^^^^^^^^^^^^^^^^^^^^^^^^^^^
#^^^^^^^^^^^^^^^^^^^^^^^^^^^^^^^^^^^^^^^^

    def figure_basic(self, plot_S='line', plot_E='line', plot_I='line',plot_R='line', plot_F='line',
                        plot_Q_E='line', plot_Q_I='line', combine_D=True,
                        color_S='tab:green', color_E='orange', color_I='crimson', color_R='tab:blue', color_F='black',
                        color_Q_E='mediumorchid', color_Q_I='mediumorchid', color_reference='#E0E0E0',
                        dashed_reference_results=None, dashed_reference_label='reference', 
                        shaded_reference_results=None, shaded_reference_label='reference', 
                        vlines=[], vline_colors=[], vline_styles=[], vline_labels=[],
                        ylim=None, xlim=None, legend=True, title=None, side_title=None, plot_percentages=True,
                        figsize=(12,8), use_seaborn=True, show=True):

        import matplotlib.pyplot as pyplot

        fig, ax = pyplot.subplots(figsize=figsize)

        if(use_seaborn):
            import seaborn
            seaborn.set_style('ticks')
            seaborn.despine()

        self.plot(ax=ax, plot_S=plot_S, plot_E=plot_E, plot_I=plot_I,plot_R=plot_R, plot_F=plot_F,
                        plot_Q_E=plot_Q_E, plot_Q_I=plot_Q_I, combine_D=combine_D,
                        color_S=color_S, color_E=color_E, color_I=color_I, color_R=color_R, color_F=color_F,
                        color_Q_E=color_Q_E, color_Q_I=color_Q_I, color_reference=color_reference,
                        dashed_reference_results=dashed_reference_results, dashed_reference_label=dashed_reference_label, 
                        shaded_reference_results=shaded_reference_results, shaded_reference_label=shaded_reference_label, 
                        vlines=vlines, vline_colors=vline_colors, vline_styles=vline_styles, vline_labels=vline_labels,
                        ylim=ylim, xlim=xlim, legend=legend, title=title, side_title=side_title, plot_percentages=plot_percentages)

        if(show):
            pyplot.show()

        return fig, ax


#^^^^^^^^^^^^^^^^^^^^^^^^^^^^^^^^^^^^^^^^
#^^^^^^^^^^^^^^^^^^^^^^^^^^^^^^^^^^^^^^^^

    def figure_infections(self, plot_S=False, plot_E='stacked', plot_I='stacked',plot_R=False, plot_F=False,
                            plot_Q_E='stacked', plot_Q_I='stacked', combine_D=True,
                            color_S='tab:green', color_E='orange', color_I='crimson', color_R='tab:blue', color_F='black',
                            color_Q_E='mediumorchid', color_Q_I='mediumorchid', color_reference='#E0E0E0',
                            dashed_reference_results=None, dashed_reference_label='reference', 
                            shaded_reference_results=None, shaded_reference_label='reference', 
                            vlines=[], vline_colors=[], vline_styles=[], vline_labels=[],
                            ylim=None, xlim=None, legend=True, title=None, side_title=None, plot_percentages=True,
                            figsize=(12,8), use_seaborn=True, show=True):

        import matplotlib.pyplot as pyplot

        fig, ax = pyplot.subplots(figsize=figsize)

        if(use_seaborn):
            import seaborn
            seaborn.set_style('ticks')
            seaborn.despine()

        self.plot(ax=ax, plot_S=plot_S, plot_E=plot_E, plot_I=plot_I,plot_R=plot_R, plot_F=plot_F,
                        plot_Q_E=plot_Q_E, plot_Q_I=plot_Q_I, combine_D=combine_D,
                        color_S=color_S, color_E=color_E, color_I=color_I, color_R=color_R, color_F=color_F,
                        color_Q_E=color_Q_E, color_Q_I=color_Q_I, color_reference=color_reference,
                        dashed_reference_results=dashed_reference_results, dashed_reference_label=dashed_reference_label, 
                        shaded_reference_results=shaded_reference_results, shaded_reference_label=shaded_reference_label, 
                        vlines=vlines, vline_colors=vline_colors, vline_styles=vline_styles, vline_labels=vline_labels, 
                        ylim=ylim, xlim=xlim, legend=legend, title=title, side_title=side_title, plot_percentages=plot_percentages)

        if(show):
            pyplot.show()

        return fig, ax


#%%%%%%%%%%%%%%%%%%%%%%%%%%%%%%%%%%%%%%%%%%%%%%%%%%
#%%%%%%%%%%%%%%%%%%%%%%%%%%%%%%%%%%%%%%%%%%%%%%%%%%
#%%%%%%%%%%%%%%%%%%%%%%%%%%%%%%%%%%%%%%%%%%%%%%%%%%
#%%%%%%%%%%%%%%%%%%%%%%%%%%%%%%%%%%%%%%%%%%%%%%%%%%


########################################################
#@@@@@@@@@@@@@@@@@@@@@@@@@@@@@@@@@@@@@@@@@@@@@@@@@@@@@@#
#@                                                    @#
#@  EXTENDED SEIRS MODELS                             @#
#@                                                    @#
#@@@@@@@@@@@@@@@@@@@@@@@@@@@@@@@@@@@@@@@@@@@@@@@@@@@@@@#
########################################################


class ExtSEIRSNetworkModel():
    """
    A class to simulate the Extended SEIRS Stochastic Network Model
    ===================================================
    Params: 
            G               Network adjacency matrix (numpy array) or Networkx graph object.
            beta            Rate of transmission (global interactions)
            beta_local      Rate(s) of transmission between adjacent individuals (optional)
            beta_asym       Rate of transmission (global interactions)
            beta_asym_local Rate(s) of transmission between adjacent individuals (optional)
            sigma           Rate of progression to infectious state (inverse of latent period)             
            lamda           Rate of progression to infectious (a)symptomatic state (inverse of prodromal period)               
            eta             Rate of progression to hospitalized state (inverse of onset-to-admission period)           
            gamma           Rate of recovery for non-hospitalized symptomatic individuals (inverse of symptomatic infectious period)           
            gamma_asym      Rate of recovery for asymptomatic individuals (inverse of asymptomatic infectious period)              
            gamma_H         Rate of recovery for hospitalized symptomatic individuals (inverse of hospitalized infectious period)              
            mu_H            Rate of death for hospitalized individuals (inverse of admission-to-death period)              
            xi              Rate of re-susceptibility (upon recovery)
            mu_0            Rate of baseline death
            nu              Rate of baseline birth
            a               Probability of an infected individual remaining asymptomatic
            h               Probability of a symptomatic individual being hospitalized  
            f               Probability of death for hospitalized individuals (case fatality rate)                         
            p               Probability of individuals interacting with global population              
            
            G_Q             Quarantine adjacency matrix (numpy array) or Networkx graph object.
            beta_Q          Rate of transmission for isolated individuals (global interactions)
            beta_Q_local    Rate(s) of transmission (exposure) for adjacent isolated individuals (optional)              
            sigma_Q         Rate of progression to infectious state for isolated individuals           
            lamda_Q         Rate of progression to infectious (a)symptomatic state for isolated individuals            
            eta_Q           Rate of progression to hospitalized state for isolated individuals             
            gamma_Q_sym     Rate of recovery for non-hospitalized symptomatic individuals for isolated individuals             
            gamma_Q_asym    Rate of recovery for asymptomatic individuals for isolated individuals             
            theta_E         Rate of random testing for exposed individuals             
            theta_pre       Rate of random testing for infectious pre-symptomatic individuals              
            theta_sym       Rate of random testing for infectious symptomatic individuals              
            theta_asym      Rate of random testing for infectious asymptomatic individuals             
            phi_E           Rate of testing when a close contact has tested positive for exposed individuals               
            phi_pre         Rate of testing when a close contact has tested positive for infectious pre-symptomatic individuals                
            phi_sym         Rate of testing when a close contact has tested positive for infectious symptomatic individuals                
            phi_asym        Rate of testing when a close contact has tested positive for infectious asymptomatic individuals               
            psi_E           Probability of positive test for exposed individuals               
            psi_pre         Probability of positive test for infectious pre-symptomatic individuals                
            psi_sym         Probability of positive test for infectious symptomatic individuals                
            psi_asym        Probability of positive test for infectious asymptomatic individuals               
            q               Probability of isolated individuals interacting with global population
            isolation_time  Time to remain in isolation upon positive test, self-isolation, etc.            
            
            initE           Initial number of exposed individuals
            initI_pre       Initial number of infectious pre-symptomatic individuals
            initI_sym       Initial number of infectious symptomatic individuals
            initI_asym      Initial number of infectious asymptomatic individuals
            initH           Initial number of hospitalized individuals
            initR           Initial number of recovered individuals     
            initF           Initial number of infection-related fatalities
            initQ_S         Initial number of isolated susceptible individuals
            initQ_E         Initial number of isolated exposed individuals
            initQ_pre       Initial number of isolated infectious pre-symptomatic individuals
            initQ_sym       Initial number of isolated infectious symptomatic individuals
            initQ_asym      Initial number of isolated infectious asymptomatic individuals
            initQ_R         Initial number of isolated recovered individuals
                            (all remaining nodes initialized susceptible)   
    """
    def __init__(self, G, beta, sigma, lamda, gamma, 
                    gamma_asym=None, eta=0, gamma_H=None, mu_H=0, alpha=1.0, xi=0, mu_0=0, nu=0, a=0, h=0, f=0, p=0,             
                    beta_local=None, beta_asym=None, beta_asym_local=None, beta_pairwise_mode='infected', delta=None, delta_pairwise_mode=None,
                    G_Q=None, beta_Q=None, beta_Q_local=None, sigma_Q=None, lamda_Q=None, eta_Q=None, gamma_Q_sym=None, gamma_Q_asym=None, alpha_Q=None, delta_Q=None,
                    theta_S=0, theta_E=0, theta_pre=0, theta_sym=0, theta_asym=0, phi_S=0, phi_E=0, phi_pre=0, phi_sym=0, phi_asym=0,    
                    psi_S=0, psi_E=1, psi_pre=1, psi_sym=1, psi_asym=1, q=0, isolation_time=14,
                    initE=0, initI_pre=0, initI_sym=0, initI_asym=0, initH=0, initR=0, initF=0,        
                    initQ_S=0, initQ_E=0, initQ_pre=0, initQ_sym=0, initQ_asym=0, initQ_R=0,
                    o=0, prevalence_ext=0,
                    transition_mode='exponential_rates', node_groups=None, store_Xseries=False, seed=None):

        if(seed is not None):
            numpy.random.seed(seed)
            self.seed = seed

        #~~~~~~~~~~~~~~~~~~~~~~~~~~~~~~~~~~~~~~~~
        # Model Parameters:
        #~~~~~~~~~~~~~~~~~~~~~~~~~~~~~~~~~~~~~~~~
        self.parameters = { 'G':G, 'G_Q':G_Q,
                            'beta':beta, 'sigma':sigma, 'lamda':lamda, 'gamma':gamma, 
                            'eta':eta, 'gamma_asym':gamma_asym, 'gamma_H':gamma_H, 'mu_H':mu_H, 
                            'xi':xi, 'mu_0':mu_0, 'nu':nu, 'a':a, 'h':h, 'f':f, 'p':p, 
                            'beta_local':beta_local, 'beta_asym':beta_asym, 'beta_asym_local':beta_asym_local, 'beta_pairwise_mode':beta_pairwise_mode,
                            'alpha':alpha, 'delta':delta, 'delta_pairwise_mode':delta_pairwise_mode,
                            'lamda_Q':lamda_Q, 'beta_Q':beta_Q, 'beta_Q_local':beta_Q_local, 'alpha_Q':alpha_Q, 'sigma_Q':sigma_Q, 
                            'eta_Q':eta_Q, 'gamma_Q_sym':gamma_Q_sym, 'gamma_Q_asym':gamma_Q_asym, 'delta_Q':delta_Q,
                            'theta_S':theta_S, 'theta_E':theta_E, 'theta_pre':theta_pre, 'theta_sym':theta_sym, 'theta_asym':theta_asym, 
                            'phi_S':phi_S, 'phi_E':phi_E, 'phi_pre':phi_pre, 'phi_sym':phi_sym, 'phi_asym':phi_asym, 
                            'psi_S':psi_S, 'psi_E':psi_E, 'psi_pre':psi_pre, 'psi_sym':psi_sym, 'psi_asym':psi_asym, 'q':q, 'isolation_time':isolation_time,
                            'initE':initE, 'initI_pre':initI_pre, 'initI_sym':initI_sym, 'initI_asym':initI_asym, 
                            'initH':initH, 'initR':initR, 'initF':initF, 
                            'initQ_S':initQ_S, 'initQ_E':initQ_E, 'initQ_pre':initQ_pre, 
                            'initQ_sym':initQ_sym, 'initQ_asym':initQ_asym, 'initQ_R':initQ_R,
                            'o':o, 'prevalence_ext':prevalence_ext}
        self.update_parameters()

        #~~~~~~~~~~~~~~~~~~~~~~~~~~~~~~~~~~~~~~~~
        # Each node can undergo 4-6 transitions (sans vitality/re-susceptibility returns to S state),
        # so there are ~numNodes*6 events/timesteps expected; initialize numNodes*6 timestep slots to start 
        # (will be expanded during run if needed for some reason)
        self.tseries    = numpy.zeros(6*self.numNodes)
        self.numS       = numpy.zeros(6*self.numNodes)
        self.numE       = numpy.zeros(6*self.numNodes)
        self.numI_pre   = numpy.zeros(6*self.numNodes)
        self.numI_sym   = numpy.zeros(6*self.numNodes)
        self.numI_asym  = numpy.zeros(6*self.numNodes)
        self.numH       = numpy.zeros(6*self.numNodes)
        self.numR       = numpy.zeros(6*self.numNodes)
        self.numF       = numpy.zeros(6*self.numNodes)
        self.numQ_S     = numpy.zeros(6*self.numNodes)
        self.numQ_E     = numpy.zeros(6*self.numNodes)
        self.numQ_pre   = numpy.zeros(6*self.numNodes)
        self.numQ_sym   = numpy.zeros(6*self.numNodes)
        self.numQ_asym  = numpy.zeros(6*self.numNodes)
        self.numQ_R     = numpy.zeros(6*self.numNodes)
        self.N          = numpy.zeros(6*self.numNodes)

        #~~~~~~~~~~~~~~~~~~~~~~~~~~~~~~~~~~~~~~~~
        # Initialize Timekeeping:
        #~~~~~~~~~~~~~~~~~~~~~~~~~~~~~~~~~~~~~~~~
        self.t          = 0
        self.tmax       = 0 # will be set when run() is called
        self.tidx       = 0
        self.tseries[0] = 0

        # Vectors holding the time that each node has been in a given state or in isolation:
        self.timer_state     = numpy.zeros((self.numNodes,1))
        self.timer_isolation = numpy.zeros(self.numNodes)
        self.isolationTime   = isolation_time
        
        #~~~~~~~~~~~~~~~~~~~~~~~~~~~~~~~~~~~~~~~~
        # Initialize Counts of inidividuals with each state:
        #~~~~~~~~~~~~~~~~~~~~~~~~~~~~~~~~~~~~~~~~
        self.numE[0]        = int(initE)
        self.numI_pre[0]    = int(initI_pre)
        self.numI_sym[0]    = int(initI_sym)
        self.numI_asym[0]   = int(initI_asym)
        self.numH[0]        = int(initH)
        self.numR[0]        = int(initR)
        self.numF[0]        = int(initF)
        self.numQ_S[0]      = int(initQ_S)
        self.numQ_E[0]      = int(initQ_E)
        self.numQ_pre[0]    = int(initQ_pre)
        self.numQ_sym[0]    = int(initQ_sym)
        self.numQ_asym[0]   = int(initQ_asym)
        self.numQ_R[0]      = int(initQ_R)
        self.numS[0]        = (self.numNodes - self.numE[0] - self.numI_pre[0] - self.numI_sym[0] - self.numI_asym[0] - self.numH[0] - self.numR[0] 
                                             - self.numQ_S[0] - self.numQ_E[0] - self.numQ_pre[0] - self.numQ_sym[0] - self.numQ_asym[0] - self.numQ_R[0] - self.numF[0])
        self.N[0]           = self.numNodes - self.numF[0]

        #~~~~~~~~~~~~~~~~~~~~~~~~~~~~~~~~~~~~~~~~
        # Node states:
        #~~~~~~~~~~~~~~~~~~~~~~~~~~~~~~~~~~~~~~~~
        self.S          = 1
        self.E          = 2
        self.I_pre      = 3
        self.I_sym      = 4
        self.I_asym     = 5
        self.H          = 6
        self.R          = 7
        self.F          = 8
        self.Q_S        = 11
        self.Q_E        = 12
        self.Q_pre      = 13
        self.Q_sym      = 14
        self.Q_asym     = 15
        self.Q_R        = 17
        
        self.X = numpy.array( [self.S]*int(self.numS[0]) + [self.E]*int(self.numE[0]) 
                               + [self.I_pre]*int(self.numI_pre[0]) + [self.I_sym]*int(self.numI_sym[0]) + [self.I_asym]*int(self.numI_asym[0]) 
                               + [self.H]*int(self.numH[0]) + [self.R]*int(self.numR[0]) + [self.F]*int(self.numF[0])
                               + [self.Q_S]*int(self.numQ_S[0]) + [self.Q_E]*int(self.numQ_E[0]) 
                               + [self.Q_pre]*int(self.numQ_pre[0]) + [self.Q_sym]*int(self.numQ_sym[0]) + [self.Q_asym]*int(self.numQ_asym[0]) 
                               + [self.Q_R]*int(self.numQ_R[0])
                            ).reshape((self.numNodes,1))
        numpy.random.shuffle(self.X)

        self.store_Xseries = store_Xseries
        if(store_Xseries):
            self.Xseries        = numpy.zeros(shape=(6*self.numNodes, self.numNodes), dtype='uint8')
            self.Xseries[0,:]   = self.X.T

        self.transitions =  { 
                                'StoE':         {'currentState':self.S,       'newState':self.E},
                                'StoQS':        {'currentState':self.S,       'newState':self.Q_S},
                                'EtoIPRE':      {'currentState':self.E,       'newState':self.I_pre},
                                'EtoQE':        {'currentState':self.E,       'newState':self.Q_E},
                                'IPREtoISYM':   {'currentState':self.I_pre,   'newState':self.I_sym},
                                'IPREtoIASYM':  {'currentState':self.I_pre,   'newState':self.I_asym},
                                'IPREtoQPRE':   {'currentState':self.I_pre,   'newState':self.Q_pre},
                                'ISYMtoH':      {'currentState':self.I_sym,   'newState':self.H},
                                'ISYMtoR':      {'currentState':self.I_sym,   'newState':self.R},
                                'ISYMtoQSYM':   {'currentState':self.I_sym,   'newState':self.Q_sym},
                                'IASYMtoR':     {'currentState':self.I_asym,  'newState':self.R},
                                'IASYMtoQASYM': {'currentState':self.I_asym,  'newState':self.Q_asym},
                                'HtoR':         {'currentState':self.H,       'newState':self.R},
                                'HtoF':         {'currentState':self.H,       'newState':self.F},
                                'RtoS':         {'currentState':self.R,       'newState':self.S},
                                'QStoQE':       {'currentState':self.Q_S,     'newState':self.Q_E},
                                'QEtoQPRE':     {'currentState':self.Q_E,     'newState':self.Q_pre},
                                'QPREtoQSYM':   {'currentState':self.Q_pre,   'newState':self.Q_sym},
                                'QPREtoQASYM':  {'currentState':self.Q_pre,   'newState':self.Q_asym},
                                'QSYMtoH':      {'currentState':self.Q_sym,   'newState':self.H},
                                'QSYMtoQR':     {'currentState':self.Q_sym,   'newState':self.Q_R},
                                'QASYMtoQR':    {'currentState':self.Q_asym,  'newState':self.Q_R},
                                '_toS':         {'currentState':True,         'newState':self.S},
                            }

        self.transition_mode = transition_mode

        #~~~~~~~~~~~~~~~~~~~~~~~~~~~~~~~~~~~~~~~~
        # Initialize other node metadata:
        #~~~~~~~~~~~~~~~~~~~~~~~~~~~~~~~~~~~~~~~~
        self.tested      = numpy.array([False]*self.numNodes).reshape((self.numNodes,1))
        self.positive    = numpy.array([False]*self.numNodes).reshape((self.numNodes,1))
        self.numTested   = numpy.zeros(6*self.numNodes) 
        self.numPositive = numpy.zeros(6*self.numNodes) 

        self.testedInCurrentState = numpy.array([False]*self.numNodes).reshape((self.numNodes,1))

        self.infectionsLog = []

        #~~~~~~~~~~~~~~~~~~~~~~~~~~~~~~~~~~~~~~~~
        # Initialize node subgroup data series:
        #~~~~~~~~~~~~~~~~~~~~~~~~~~~~~~~~~~~~~~~~
        self.nodeGroupData = None
        if(node_groups):
            self.nodeGroupData = {}
            for groupName, nodeList in node_groups.items():
                self.nodeGroupData[groupName] = {'nodes':   numpy.array(nodeList),
                                                 'mask':    numpy.isin(range(self.numNodes), nodeList).reshape((self.numNodes,1))}
                self.nodeGroupData[groupName]['numS']           = numpy.zeros(6*self.numNodes)
                self.nodeGroupData[groupName]['numE']           = numpy.zeros(6*self.numNodes)
                self.nodeGroupData[groupName]['numI_pre']       = numpy.zeros(6*self.numNodes)
                self.nodeGroupData[groupName]['numI_sym']       = numpy.zeros(6*self.numNodes)
                self.nodeGroupData[groupName]['numI_asym']      = numpy.zeros(6*self.numNodes)
                self.nodeGroupData[groupName]['numH']           = numpy.zeros(6*self.numNodes)
                self.nodeGroupData[groupName]['numR']           = numpy.zeros(6*self.numNodes)
                self.nodeGroupData[groupName]['numF']           = numpy.zeros(6*self.numNodes)
                self.nodeGroupData[groupName]['numQ_S']         = numpy.zeros(6*self.numNodes)
                self.nodeGroupData[groupName]['numQ_E']         = numpy.zeros(6*self.numNodes)
                self.nodeGroupData[groupName]['numQ_pre']       = numpy.zeros(6*self.numNodes)
                self.nodeGroupData[groupName]['numQ_sym']       = numpy.zeros(6*self.numNodes)
                self.nodeGroupData[groupName]['numQ_asym']      = numpy.zeros(6*self.numNodes)
                self.nodeGroupData[groupName]['numQ_R']         = numpy.zeros(6*self.numNodes)
                self.nodeGroupData[groupName]['N']              = numpy.zeros(6*self.numNodes)
                self.nodeGroupData[groupName]['numPositive']    = numpy.zeros(6*self.numNodes)
                self.nodeGroupData[groupName]['numTested']      = numpy.zeros(6*self.numNodes)
                self.nodeGroupData[groupName]['numS'][0]        = numpy.count_nonzero(self.nodeGroupData[groupName]['mask']*self.X==self.S)
                self.nodeGroupData[groupName]['numE'][0]        = numpy.count_nonzero(self.nodeGroupData[groupName]['mask']*self.X==self.E)
                self.nodeGroupData[groupName]['numI_pre'][0]    = numpy.count_nonzero(self.nodeGroupData[groupName]['mask']*self.X==self.I_pre)
                self.nodeGroupData[groupName]['numI_sym'][0]    = numpy.count_nonzero(self.nodeGroupData[groupName]['mask']*self.X==self.I_sym)
                self.nodeGroupData[groupName]['numI_asym'][0]   = numpy.count_nonzero(self.nodeGroupData[groupName]['mask']*self.X==self.I_asym)
                self.nodeGroupData[groupName]['numH'][0]        = numpy.count_nonzero(self.nodeGroupData[groupName]['mask']*self.X==self.H)
                self.nodeGroupData[groupName]['numR'][0]        = numpy.count_nonzero(self.nodeGroupData[groupName]['mask']*self.X==self.R)
                self.nodeGroupData[groupName]['numF'][0]        = numpy.count_nonzero(self.nodeGroupData[groupName]['mask']*self.X==self.F)
                self.nodeGroupData[groupName]['numQ_S'][0]      = numpy.count_nonzero(self.nodeGroupData[groupName]['mask']*self.X==self.Q_E)
                self.nodeGroupData[groupName]['numQ_E'][0]      = numpy.count_nonzero(self.nodeGroupData[groupName]['mask']*self.X==self.Q_E)
                self.nodeGroupData[groupName]['numQ_pre'][0]    = numpy.count_nonzero(self.nodeGroupData[groupName]['mask']*self.X==self.Q_pre)
                self.nodeGroupData[groupName]['numQ_I_sym'][0]  = numpy.count_nonzero(self.nodeGroupData[groupName]['mask']*self.X==self.Q_I_sym)
                self.nodeGroupData[groupName]['numQ_I_asym'][0] = numpy.count_nonzero(self.nodeGroupData[groupName]['mask']*self.X==self.Q_I_asym)
                self.nodeGroupData[groupName]['numQ_R'][0]      = numpy.count_nonzero(self.nodeGroupData[groupName]['mask']*self.X==self.Q_E)
                self.nodeGroupData[groupName]['N'][0]           = self.numNodes - self.numF[0]

         
#^^^^^^^^^^^^^^^^^^^^^^^^^^^^^^^^^^^^^^^^
#^^^^^^^^^^^^^^^^^^^^^^^^^^^^^^^^^^^^^^^^

    def update_parameters(self):

        #~~~~~~~~~~~~~~~~~~~~~~~~~~~~~~~~~~~~~~~~
        # Model graphs:
        #~~~~~~~~~~~~~~~~~~~~~~~~~~~~~~~~~~~~~~~~
        self.G = self.parameters['G']
        # Adjacency matrix:
        if type(self.G)==numpy.ndarray:
            self.A = scipy.sparse.csr_matrix(self.G)
        elif type(self.G)==networkx.classes.graph.Graph:
            self.A = networkx.adj_matrix(self.G) # adj_matrix gives scipy.sparse csr_matrix
        else:
            raise BaseException("Input an adjacency matrix or networkx object only.")
        self.numNodes   = int(self.A.shape[1])
        self.degree     = numpy.asarray(self.node_degrees(self.A)).astype(float)
        #----------------------------------------
        if(self.parameters['G_Q'] is None):
            self.G_Q = self.G # If no Q graph is provided, use G in its place
        else:
            self.G_Q = self.parameters['G_Q']
        # Quarantine Adjacency matrix:
        if type(self.G_Q)==numpy.ndarray:
            self.A_Q = scipy.sparse.csr_matrix(self.G_Q)
        elif type(self.G_Q)==networkx.classes.graph.Graph:
            self.A_Q = networkx.adj_matrix(self.G_Q) # adj_matrix gives scipy.sparse csr_matrix
        else:
            raise BaseException("Input an adjacency matrix or networkx object only.")
        self.numNodes_Q   = int(self.A_Q.shape[1])
        self.degree_Q     = numpy.asarray(self.node_degrees(self.A_Q)).astype(float)
        #----------------------------------------
        assert(self.numNodes == self.numNodes_Q), "The normal and quarantine adjacency graphs must be of the same size."

        #~~~~~~~~~~~~~~~~~~~~~~~~~~~~~~~~~~~~~~~~
        # Model parameters:
        #~~~~~~~~~~~~~~~~~~~~~~~~~~~~~~~~~~~~~~~~
        self.beta           = numpy.array(self.parameters['beta']).reshape((self.numNodes, 1))          if isinstance(self.parameters['beta'], (list, numpy.ndarray)) else numpy.full(fill_value=self.parameters['beta'], shape=(self.numNodes,1))
        self.beta_asym      = (numpy.array(self.parameters['beta_asym']).reshape((self.numNodes, 1))    if isinstance(self.parameters['beta_asym'], (list, numpy.ndarray)) else numpy.full(fill_value=self.parameters['beta_asym'], shape=(self.numNodes,1))) if self.parameters['beta_asym'] is not None else self.beta
        self.sigma          = numpy.array(self.parameters['sigma']).reshape((self.numNodes, 1))         if isinstance(self.parameters['sigma'], (list, numpy.ndarray)) else numpy.full(fill_value=self.parameters['sigma'], shape=(self.numNodes,1))
        self.lamda          = numpy.array(self.parameters['lamda']).reshape((self.numNodes, 1))         if isinstance(self.parameters['lamda'], (list, numpy.ndarray)) else numpy.full(fill_value=self.parameters['lamda'], shape=(self.numNodes,1))
        self.gamma          = numpy.array(self.parameters['gamma']).reshape((self.numNodes, 1))         if isinstance(self.parameters['gamma'], (list, numpy.ndarray)) else numpy.full(fill_value=self.parameters['gamma'], shape=(self.numNodes,1))
        self.eta            = numpy.array(self.parameters['eta']).reshape((self.numNodes, 1))           if isinstance(self.parameters['eta'], (list, numpy.ndarray)) else numpy.full(fill_value=self.parameters['eta'], shape=(self.numNodes,1))
        self.gamma_asym     = (numpy.array(self.parameters['gamma_asym']).reshape((self.numNodes, 1))   if isinstance(self.parameters['gamma_asym'], (list, numpy.ndarray)) else numpy.full(fill_value=self.parameters['gamma_asym'], shape=(self.numNodes,1))) if self.parameters['gamma_asym'] is not None else self.gamma
        self.gamma_H        = (numpy.array(self.parameters['gamma_H']).reshape((self.numNodes, 1))      if isinstance(self.parameters['gamma_H'], (list, numpy.ndarray)) else numpy.full(fill_value=self.parameters['gamma_H'], shape=(self.numNodes,1))) if self.parameters['gamma_H'] is not None else self.gamma
        self.mu_H           = numpy.array(self.parameters['mu_H']).reshape((self.numNodes, 1))          if isinstance(self.parameters['mu_H'], (list, numpy.ndarray)) else numpy.full(fill_value=self.parameters['mu_H'], shape=(self.numNodes,1))
        self.alpha          = numpy.array(self.parameters['alpha']).reshape((self.numNodes, 1))         if isinstance(self.parameters['alpha'], (list, numpy.ndarray)) else numpy.full(fill_value=self.parameters['alpha'], shape=(self.numNodes,1))
        self.xi             = numpy.array(self.parameters['xi']).reshape((self.numNodes, 1))            if isinstance(self.parameters['xi'], (list, numpy.ndarray)) else numpy.full(fill_value=self.parameters['xi'], shape=(self.numNodes,1))
        self.mu_0           = numpy.array(self.parameters['mu_0']).reshape((self.numNodes, 1))          if isinstance(self.parameters['mu_0'], (list, numpy.ndarray)) else numpy.full(fill_value=self.parameters['mu_0'], shape=(self.numNodes,1))
        self.nu             = numpy.array(self.parameters['nu']).reshape((self.numNodes, 1))            if isinstance(self.parameters['nu'], (list, numpy.ndarray)) else numpy.full(fill_value=self.parameters['nu'], shape=(self.numNodes,1))
        self.a              = numpy.array(self.parameters['a']).reshape((self.numNodes, 1))             if isinstance(self.parameters['a'], (list, numpy.ndarray)) else numpy.full(fill_value=self.parameters['a'], shape=(self.numNodes,1))
        self.h              = numpy.array(self.parameters['h']).reshape((self.numNodes, 1))             if isinstance(self.parameters['h'], (list, numpy.ndarray)) else numpy.full(fill_value=self.parameters['h'], shape=(self.numNodes,1))
        self.f              = numpy.array(self.parameters['f']).reshape((self.numNodes, 1))             if isinstance(self.parameters['f'], (list, numpy.ndarray)) else numpy.full(fill_value=self.parameters['f'], shape=(self.numNodes,1))
        self.p              = numpy.array(self.parameters['p']).reshape((self.numNodes, 1))             if isinstance(self.parameters['p'], (list, numpy.ndarray)) else numpy.full(fill_value=self.parameters['p'], shape=(self.numNodes,1))
        self.o              = numpy.array(self.parameters['o']).reshape((self.numNodes, 1))             if isinstance(self.parameters['o'], (list, numpy.ndarray)) else numpy.full(fill_value=self.parameters['o'], shape=(self.numNodes,1))

        self.rand_a = numpy.random.rand(self.a.shape[0], self.a.shape[1])
        self.rand_h = numpy.random.rand(self.h.shape[0], self.h.shape[1])
        self.rand_f = numpy.random.rand(self.f.shape[0], self.f.shape[1])

        #~~~~~~~~~~~~~~~~~~~~~~~~~~~~~~~~~~~~~~~~
        # External infection introduction variables:
        #~~~~~~~~~~~~~~~~~~~~~~~~~~~~~~~~~~~~~~~~
        self.prevalence_ext     = numpy.array(self.parameters['prevalence_ext']).reshape((self.numNodes, 1)) if isinstance(self.parameters['prevalence_ext'], (list, numpy.ndarray)) else numpy.full(fill_value=self.parameters['prevalence_ext'], shape=(self.numNodes,1))
       
        #----------------------------------------
        # Testing-related parameters:
        #----------------------------------------
        self.beta_Q         = (numpy.array(self.parameters['beta_Q']).reshape((self.numNodes, 1))       if isinstance(self.parameters['beta_Q'], (list, numpy.ndarray)) else numpy.full(fill_value=self.parameters['beta_Q'], shape=(self.numNodes,1))) if self.parameters['beta_Q'] is not None else self.beta
        self.sigma_Q        = (numpy.array(self.parameters['sigma_Q']).reshape((self.numNodes, 1))      if isinstance(self.parameters['sigma_Q'], (list, numpy.ndarray)) else numpy.full(fill_value=self.parameters['sigma_Q'], shape=(self.numNodes,1))) if self.parameters['sigma_Q'] is not None else self.sigma
        self.lamda_Q        = (numpy.array(self.parameters['lamda_Q']).reshape((self.numNodes, 1))      if isinstance(self.parameters['lamda_Q'], (list, numpy.ndarray)) else numpy.full(fill_value=self.parameters['lamda_Q'], shape=(self.numNodes,1))) if self.parameters['lamda_Q'] is not None else self.lamda
        self.gamma_Q_sym    = (numpy.array(self.parameters['gamma_Q_sym']).reshape((self.numNodes, 1))  if isinstance(self.parameters['gamma_Q_sym'], (list, numpy.ndarray)) else numpy.full(fill_value=self.parameters['gamma_Q_sym'], shape=(self.numNodes,1))) if self.parameters['gamma_Q_sym'] is not None else self.gamma
        self.gamma_Q_asym   = (numpy.array(self.parameters['gamma_Q_asym']).reshape((self.numNodes, 1)) if isinstance(self.parameters['gamma_Q_asym'], (list, numpy.ndarray)) else numpy.full(fill_value=self.parameters['gamma_Q_asym'], shape=(self.numNodes,1))) if self.parameters['gamma_Q_asym'] is not None else self.gamma
        self.eta_Q          = (numpy.array(self.parameters['eta_Q']).reshape((self.numNodes, 1))        if isinstance(self.parameters['eta_Q'], (list, numpy.ndarray)) else numpy.full(fill_value=self.parameters['eta_Q'], shape=(self.numNodes,1))) if self.parameters['eta_Q'] is not None else self.eta
        self.alpha_Q        = (numpy.array(self.parameters['alpha_Q']).reshape((self.numNodes, 1))      if isinstance(self.parameters['alpha_Q'], (list, numpy.ndarray)) else numpy.full(fill_value=self.parameters['alpha_Q'], shape=(self.numNodes,1))) if self.parameters['alpha_Q'] is not None else self.alpha
        self.theta_S        = numpy.array(self.parameters['theta_S']).reshape((self.numNodes, 1))       if isinstance(self.parameters['theta_S'], (list, numpy.ndarray)) else numpy.full(fill_value=self.parameters['theta_S'], shape=(self.numNodes,1))
        self.theta_E        = numpy.array(self.parameters['theta_E']).reshape((self.numNodes, 1))       if isinstance(self.parameters['theta_E'], (list, numpy.ndarray)) else numpy.full(fill_value=self.parameters['theta_E'], shape=(self.numNodes,1))
        self.theta_pre      = numpy.array(self.parameters['theta_pre']).reshape((self.numNodes, 1))     if isinstance(self.parameters['theta_pre'], (list, numpy.ndarray)) else numpy.full(fill_value=self.parameters['theta_pre'], shape=(self.numNodes,1))
        self.theta_sym      = numpy.array(self.parameters['theta_sym']).reshape((self.numNodes, 1))     if isinstance(self.parameters['theta_sym'], (list, numpy.ndarray)) else numpy.full(fill_value=self.parameters['theta_sym'], shape=(self.numNodes,1))
        self.theta_asym     = numpy.array(self.parameters['theta_asym']).reshape((self.numNodes, 1))    if isinstance(self.parameters['theta_asym'], (list, numpy.ndarray)) else numpy.full(fill_value=self.parameters['theta_asym'], shape=(self.numNodes,1))
        self.phi_S          = numpy.array(self.parameters['phi_S']).reshape((self.numNodes, 1))         if isinstance(self.parameters['phi_S'], (list, numpy.ndarray)) else numpy.full(fill_value=self.parameters['phi_S'], shape=(self.numNodes,1))
        self.phi_E          = numpy.array(self.parameters['phi_E']).reshape((self.numNodes, 1))         if isinstance(self.parameters['phi_E'], (list, numpy.ndarray)) else numpy.full(fill_value=self.parameters['phi_E'], shape=(self.numNodes,1))
        self.phi_pre        = numpy.array(self.parameters['phi_pre']).reshape((self.numNodes, 1))       if isinstance(self.parameters['phi_pre'], (list, numpy.ndarray)) else numpy.full(fill_value=self.parameters['phi_pre'], shape=(self.numNodes,1))
        self.phi_sym        = numpy.array(self.parameters['phi_sym']).reshape((self.numNodes, 1))       if isinstance(self.parameters['phi_sym'], (list, numpy.ndarray)) else numpy.full(fill_value=self.parameters['phi_sym'], shape=(self.numNodes,1))
        self.phi_asym       = numpy.array(self.parameters['phi_asym']).reshape((self.numNodes, 1))      if isinstance(self.parameters['phi_asym'], (list, numpy.ndarray)) else numpy.full(fill_value=self.parameters['phi_asym'], shape=(self.numNodes,1))
        self.psi_S            = numpy.array(self.parameters['psi_S']).reshape((self.numNodes, 1))           if isinstance(self.parameters['psi_S'], (list, numpy.ndarray)) else numpy.full(fill_value=self.parameters['psi_S'], shape=(self.numNodes,1))
        self.psi_E            = numpy.array(self.parameters['psi_E']).reshape((self.numNodes, 1))           if isinstance(self.parameters['psi_E'], (list, numpy.ndarray)) else numpy.full(fill_value=self.parameters['psi_E'], shape=(self.numNodes,1))
        self.psi_pre          = numpy.array(self.parameters['psi_pre']).reshape((self.numNodes, 1))         if isinstance(self.parameters['psi_pre'], (list, numpy.ndarray)) else numpy.full(fill_value=self.parameters['psi_pre'], shape=(self.numNodes,1))
        self.psi_sym          = numpy.array(self.parameters['psi_sym']).reshape((self.numNodes, 1))         if isinstance(self.parameters['psi_sym'], (list, numpy.ndarray)) else numpy.full(fill_value=self.parameters['psi_sym'], shape=(self.numNodes,1))
        self.psi_asym         = numpy.array(self.parameters['psi_asym']).reshape((self.numNodes, 1))        if isinstance(self.parameters['psi_asym'], (list, numpy.ndarray)) else numpy.full(fill_value=self.parameters['psi_asym'], shape=(self.numNodes,1))
        self.q              = numpy.array(self.parameters['q']).reshape((self.numNodes, 1))             if isinstance(self.parameters['q'], (list, numpy.ndarray)) else numpy.full(fill_value=self.parameters['q'], shape=(self.numNodes,1))

        #----------------------------------------

        self.beta_pairwise_mode = self.parameters['beta_pairwise_mode']

        #----------------------------------------
        # Global transmission parameters:
        #----------------------------------------
        if(self.beta_pairwise_mode == 'infected' or self.beta_pairwise_mode is None):
            self.beta_global         = numpy.full_like(self.beta, fill_value=numpy.mean(self.beta))
            self.beta_Q_global       = numpy.full_like(self.beta_Q, fill_value=numpy.mean(self.beta_Q))
            self.beta_asym_global    = numpy.full_like(self.beta_asym, fill_value=numpy.mean(self.beta_asym))
        elif(self.beta_pairwise_mode == 'infectee'):
            self.beta_global         = self.beta      
            self.beta_Q_global       = self.beta_Q    
            self.beta_asym_global    = self.beta_asym
        elif(self.beta_pairwise_mode == 'min'):
            self.beta_global         = numpy.minimum(self.beta, numpy.mean(beta)) 
            self.beta_Q_global       = numpy.minimum(self.beta_Q, numpy.mean(beta_Q)) 
            self.beta_asym_global    = numpy.minimum(self.beta_asym, numpy.mean(beta_asym))
        elif(self.beta_pairwise_mode == 'max'):
            self.beta_global         = numpy.maximum(self.beta, numpy.mean(beta)) 
            self.beta_Q_global       = numpy.maximum(self.beta_Q, numpy.mean(beta_Q)) 
            self.beta_asym_global    = numpy.maximum(self.beta_asym, numpy.mean(beta_asym))
        elif(self.beta_pairwise_mode == 'mean'):
            self.beta_global         = (self.beta + numpy.full_like(self.beta, fill_value=numpy.mean(self.beta)))/2
            self.beta_Q_global       = (self.beta_Q + numpy.full_like(self.beta_Q, fill_value=numpy.mean(self.beta_Q)))/2
            self.beta_asym_global    = (self.beta_asym + numpy.full_like(self.beta_asym, fill_value=numpy.mean(self.beta_asym)))/2
            
        #----------------------------------------
        # Local transmission parameters:
        #----------------------------------------
        self.beta_local         = self.beta      if self.parameters['beta_local'] is None      else numpy.array(self.parameters['beta_local'])      if isinstance(self.parameters['beta_local'], (list, numpy.ndarray))      else numpy.full(fill_value=self.parameters['beta_local'], shape=(self.numNodes,1))
        self.beta_Q_local       = self.beta_Q    if self.parameters['beta_Q_local'] is None    else numpy.array(self.parameters['beta_Q_local'])    if isinstance(self.parameters['beta_Q_local'], (list, numpy.ndarray))    else numpy.full(fill_value=self.parameters['beta_Q_local'], shape=(self.numNodes,1))
        self.beta_asym_local    = None           if self.parameters['beta_asym_local'] is None else numpy.array(self.parameters['beta_asym_local']) if isinstance(self.parameters['beta_asym_local'], (list, numpy.ndarray)) else numpy.full(fill_value=self.parameters['beta_asym_local'], shape=(self.numNodes,1))
        #----------------------------------------
        if(self.beta_local.ndim == 2 and self.beta_local.shape[0] == self.numNodes and self.beta_local.shape[1] == self.numNodes):
            self.A_beta_pairwise = self.beta_local
        elif((self.beta_local.ndim == 1 and self.beta_local.shape[0] == self.numNodes) or (self.beta_local.ndim == 2 and (self.beta_local.shape[0] == self.numNodes or self.beta_local.shape[1] == self.numNodes))):
            self.beta_local = self.beta_local.reshape((self.numNodes,1))
            # Pre-multiply beta values by the adjacency matrix ("transmission weight connections")
            A_beta_pairwise_byInfected = scipy.sparse.csr_matrix.multiply(self.A, self.beta_local.T).tocsr()
            A_beta_pairwise_byInfectee = scipy.sparse.csr_matrix.multiply(self.A, self.beta_local).tocsr()    
            #------------------------------
            # Compute the effective pairwise beta values as a function of the infected/infectee pair:
            if(self.beta_pairwise_mode == 'infected'):
                self.A_beta_pairwise = A_beta_pairwise_byInfected
            elif(self.beta_pairwise_mode == 'infectee'):
                self.A_beta_pairwise = A_beta_pairwise_byInfectee
            elif(self.beta_pairwise_mode == 'min'):
                self.A_beta_pairwise = scipy.sparse.csr_matrix.minimum(A_beta_pairwise_byInfected, A_beta_pairwise_byInfectee)
            elif(self.beta_pairwise_mode == 'max'):
                self.A_beta_pairwise = scipy.sparse.csr_matrix.maximum(A_beta_pairwise_byInfected, A_beta_pairwise_byInfectee)
            elif(self.beta_pairwise_mode == 'mean' or self.beta_pairwise_mode is None):
                self.A_beta_pairwise = (A_beta_pairwise_byInfected + A_beta_pairwise_byInfectee)/2
            else:
                print("Unrecognized beta_pairwise_mode value (support for 'infected', 'infectee', 'min', 'max', and 'mean').")
        else:
            print("Invalid values given for beta_local (expected 1xN list/array or NxN 2d array)")
        #----------------------------------------
        if(self.beta_Q_local.ndim == 2 and self.beta_Q_local.shape[0] == self.numNodes and self.beta_Q_local.shape[1] == self.numNodes):
            self.A_Q_beta_Q_pairwise = self.beta_Q_local
        elif((self.beta_Q_local.ndim == 1 and self.beta_Q_local.shape[0] == self.numNodes) or (self.beta_Q_local.ndim == 2 and (self.beta_Q_local.shape[0] == self.numNodes or self.beta_Q_local.shape[1] == self.numNodes))):
            self.beta_Q_local = self.beta_Q_local.reshape((self.numNodes,1))
            # Pre-multiply beta_Q values by the isolation adjacency matrix ("transmission weight connections")
            A_Q_beta_Q_pairwise_byInfected      = scipy.sparse.csr_matrix.multiply(self.A_Q, self.beta_Q_local.T).tocsr()
            A_Q_beta_Q_pairwise_byInfectee      = scipy.sparse.csr_matrix.multiply(self.A_Q, self.beta_Q_local).tocsr()
            #------------------------------
            # Compute the effective pairwise beta values as a function of the infected/infectee pair:
            if(self.beta_pairwise_mode == 'infected'):
                self.A_Q_beta_Q_pairwise = A_Q_beta_Q_pairwise_byInfected
            elif(self.beta_pairwise_mode == 'infectee'):
                self.A_Q_beta_Q_pairwise = A_Q_beta_Q_pairwise_byInfectee
            elif(self.beta_pairwise_mode == 'min'):
                self.A_Q_beta_Q_pairwise = scipy.sparse.csr_matrix.minimum(A_Q_beta_Q_pairwise_byInfected, A_Q_beta_Q_pairwise_byInfectee)
            elif(self.beta_pairwise_mode == 'max'):
                self.A_Q_beta_Q_pairwise = scipy.sparse.csr_matrix.maximum(A_Q_beta_Q_pairwise_byInfected, A_Q_beta_Q_pairwise_byInfectee)
            elif(self.beta_pairwise_mode == 'mean' or self.beta_pairwise_mode is None):
                self.A_Q_beta_Q_pairwise = (A_Q_beta_Q_pairwise_byInfected + A_Q_beta_Q_pairwise_byInfectee)/2
            else:
                print("Unrecognized beta_pairwise_mode value (support for 'infected', 'infectee', 'min', 'max', and 'mean').")
        else:
            print("Invalid values given for beta_Q_local (expected 1xN list/array or NxN 2d array)")
        #----------------------------------------
        if(self.beta_asym_local is None):
            self.A_beta_asym_pairwise = None
        elif(self.beta_asym_local.ndim == 2 and self.beta_asym_local.shape[0] == self.numNodes and self.beta_asym_local.shape[1] == self.numNodes):
            self.A_beta_asym_pairwise = self.beta_asym_local
        elif((self.beta_asym_local.ndim == 1 and self.beta_asym_local.shape[0] == self.numNodes) or (self.beta_asym_local.ndim == 2 and (self.beta_asym_local.shape[0] == self.numNodes or self.beta_asym_local.shape[1] == self.numNodes))):
            self.beta_asym_local = self.beta_asym_local.reshape((self.numNodes,1))
            # Pre-multiply beta_asym values by the adjacency matrix ("transmission weight connections")
            A_beta_asym_pairwise_byInfected      = scipy.sparse.csr_matrix.multiply(self.A, self.beta_asym_local.T).tocsr()
            A_beta_asym_pairwise_byInfectee      = scipy.sparse.csr_matrix.multiply(self.A, self.beta_asym_local).tocsr()
            #------------------------------
            # Compute the effective pairwise beta values as a function of the infected/infectee pair:
            if(self.beta_pairwise_mode == 'infected'):
                self.A_beta_asym_pairwise = A_beta_asym_pairwise_byInfected
            elif(self.beta_pairwise_mode == 'infectee'):
                self.A_beta_asym_pairwise = A_beta_asym_pairwise_byInfectee
            elif(self.beta_pairwise_mode == 'min'):
                self.A_beta_asym_pairwise = scipy.sparse.csr_matrix.minimum(A_beta_asym_pairwise_byInfected, A_beta_asym_pairwise_byInfectee)
            elif(self.beta_pairwise_mode == 'max'):
                self.A_beta_asym_pairwise = scipy.sparse.csr_matrix.maximum(A_beta_asym_pairwise_byInfected, A_beta_asym_pairwise_byInfectee)
            elif(self.beta_pairwise_mode == 'mean' or self.beta_pairwise_mode is None):
                self.A_beta_asym_pairwise = (A_beta_asym_pairwise_byInfected + A_beta_asym_pairwise_byInfectee)/2
            else:
                print("Unrecognized beta_pairwise_mode value (support for 'infected', 'infectee', 'min', 'max', and 'mean').")
        else:
            print("Invalid values given for beta_asym_local (expected 1xN list/array or NxN 2d array)")

        #----------------------------------------
        # Degree-based transmission scaling parameters:
        #----------------------------------------
        self.delta_pairwise_mode = self.parameters['delta_pairwise_mode']
        with numpy.errstate(divide='ignore'): # ignore log(0) warning, then convert log(0) = -inf -> 0.0
            self.delta               = numpy.log(self.degree)/numpy.log(numpy.mean(self.degree))     if self.parameters['delta'] is None   else numpy.array(self.parameters['delta'])   if isinstance(self.parameters['delta'], (list, numpy.ndarray))   else numpy.full(fill_value=self.parameters['delta'], shape=(self.numNodes,1))
            self.delta_Q             = numpy.log(self.degree_Q)/numpy.log(numpy.mean(self.degree_Q)) if self.parameters['delta_Q'] is None else numpy.array(self.parameters['delta_Q']) if isinstance(self.parameters['delta_Q'], (list, numpy.ndarray)) else numpy.full(fill_value=self.parameters['delta_Q'], shape=(self.numNodes,1))
        self.delta[numpy.isneginf(self.delta)] = 0.0
        self.delta_Q[numpy.isneginf(self.delta_Q)] = 0.0
        #----------------------------------------
        if(self.delta.ndim == 2 and self.delta.shape[0] == self.numNodes and self.delta.shape[1] == self.numNodes):
            self.A_delta_pairwise = self.delta
        elif((self.delta.ndim == 1 and self.delta.shape[0] == self.numNodes) or (self.delta.ndim == 2 and (self.delta.shape[0] == self.numNodes or self.delta.shape[1] == self.numNodes))):
            self.delta = self.delta.reshape((self.numNodes,1))
            # Pre-multiply delta values by the adjacency matrix ("transmission weight connections")
            A_delta_pairwise_byInfected = scipy.sparse.csr_matrix.multiply(self.A, self.delta.T).tocsr()
            A_delta_pairwise_byInfectee = scipy.sparse.csr_matrix.multiply(self.A, self.delta).tocsr()    
            #------------------------------
            # Compute the effective pairwise delta values as a function of the infected/infectee pair:
            if(self.delta_pairwise_mode == 'infected'):
                self.A_delta_pairwise = A_delta_pairwise_byInfected
            elif(self.delta_pairwise_mode == 'infectee'):
                self.A_delta_pairwise = A_delta_pairwise_byInfectee
            elif(self.delta_pairwise_mode == 'min'):
                self.A_delta_pairwise = scipy.sparse.csr_matrix.minimum(A_delta_pairwise_byInfected, A_delta_pairwise_byInfectee)
            elif(self.delta_pairwise_mode == 'max'):
                self.A_delta_pairwise = scipy.sparse.csr_matrix.maximum(A_delta_pairwise_byInfected, A_delta_pairwise_byInfectee)
            elif(self.delta_pairwise_mode == 'mean'):
                self.A_delta_pairwise = (A_delta_pairwise_byInfected + A_delta_pairwise_byInfectee)/2
            elif(self.delta_pairwise_mode is None):
                self.A_delta_pairwise = self.A
            else:
                print("Unrecognized delta_pairwise_mode value (support for 'infected', 'infectee', 'min', 'max', and 'mean').")
        else:
            print("Invalid values given for delta (expected 1xN list/array or NxN 2d array)")
        #----------------------------------------
        if(self.delta_Q.ndim == 2 and self.delta_Q.shape[0] == self.numNodes and self.delta_Q.shape[1] == self.numNodes):
            self.A_Q_delta_Q_pairwise = self.delta_Q
        elif((self.delta_Q.ndim == 1 and self.delta_Q.shape[0] == self.numNodes) or (self.delta_Q.ndim == 2 and (self.delta_Q.shape[0] == self.numNodes or self.delta_Q.shape[1] == self.numNodes))):
            self.delta_Q = self.delta_Q.reshape((self.numNodes,1))
            # Pre-multiply delta_Q values by the isolation adjacency matrix ("transmission weight connections")
            A_Q_delta_Q_pairwise_byInfected      = scipy.sparse.csr_matrix.multiply(self.A_Q, self.delta_Q).tocsr()
            A_Q_delta_Q_pairwise_byInfectee      = scipy.sparse.csr_matrix.multiply(self.A_Q, self.delta_Q.T).tocsr()
            #------------------------------
            # Compute the effective pairwise delta values as a function of the infected/infectee pair:
            if(self.delta_pairwise_mode == 'infected'):
                self.A_Q_delta_Q_pairwise = A_Q_delta_Q_pairwise_byInfected
            elif(self.delta_pairwise_mode == 'infectee'):
                self.A_Q_delta_Q_pairwise = A_Q_delta_Q_pairwise_byInfectee
            elif(self.delta_pairwise_mode == 'min'):
                self.A_Q_delta_Q_pairwise = scipy.sparse.csr_matrix.minimum(A_Q_delta_Q_pairwise_byInfected, A_Q_delta_Q_pairwise_byInfectee)
            elif(self.delta_pairwise_mode == 'max'):
                self.A_Q_delta_Q_pairwise = scipy.sparse.csr_matrix.maximum(A_Q_delta_Q_pairwise_byInfected, A_Q_delta_Q_pairwise_byInfectee)
            elif(self.delta_pairwise_mode == 'mean'):
                self.A_Q_delta_Q_pairwise = (A_Q_delta_Q_pairwise_byInfected + A_Q_delta_Q_pairwise_byInfectee)/2
            elif(self.delta_pairwise_mode is None):
                self.A_Q_delta_Q_pairwise = self.A
            else:
                print("Unrecognized delta_pairwise_mode value (support for 'infected', 'infectee', 'min', 'max', and 'mean').")
        else:
            print("Invalid values given for delta_Q (expected 1xN list/array or NxN 2d array)")

        #----------------------------------------
        # Pre-calculate the pairwise delta*beta values:
        #----------------------------------------
        self.A_deltabeta          = scipy.sparse.csr_matrix.multiply(self.A_delta_pairwise, self.A_beta_pairwise)
        self.A_Q_deltabeta_Q      = scipy.sparse.csr_matrix.multiply(self.A_Q_delta_Q_pairwise, self.A_Q_beta_Q_pairwise)
        if(self.A_beta_asym_pairwise is not None):
            self.A_deltabeta_asym = scipy.sparse.csr_matrix.multiply(self.A_delta_pairwise, self.A_beta_asym_pairwise)
        else:
            self.A_deltabeta_asym = None
    

#^^^^^^^^^^^^^^^^^^^^^^^^^^^^^^^^^^^^^^^^
#^^^^^^^^^^^^^^^^^^^^^^^^^^^^^^^^^^^^^^^^

    def node_degrees(self, Amat):
        return Amat.sum(axis=0).reshape(self.numNodes,1)   # sums of adj matrix cols


#^^^^^^^^^^^^^^^^^^^^^^^^^^^^^^^^^^^^^^^^
#^^^^^^^^^^^^^^^^^^^^^^^^^^^^^^^^^^^^^^^^

    def total_num_susceptible(self, t_idx=None):
        if(t_idx is None):
            return (self.numS[:] + self.numQ_S[:])            
        else:
            return (self.numS[t_idx] + self.numQ_S[t_idx])

#^^^^^^^^^^^^^^^^^^^^^^^^^^^^^^^^^^^^^^^^

    def total_num_infected(self, t_idx=None):
        if(t_idx is None):
            return (self.numE[:] + self.numI_pre[:] + self.numI_sym[:] + self.numI_asym[:] + self.numH[:]
                    + self.numQ_E[:] + self.numQ_pre[:] + self.numQ_sym[:] + self.numQ_asym[:])            
        else:
            return (self.numE[t_idx] + self.numI_pre[t_idx] + self.numI_sym[t_idx] + self.numI_asym[t_idx] + self.numH[t_idx]
                    + self.numQ_E[t_idx] + self.numQ_pre[t_idx] + self.numQ_sym[t_idx] + self.numQ_asym[t_idx])

#^^^^^^^^^^^^^^^^^^^^^^^^^^^^^^^^^^^^^^^^

    def total_num_isolated(self, t_idx=None):
        if(t_idx is None):
            return (self.numQ_S[:] + self.numQ_E[:] + self.numQ_pre[:] + self.numQ_sym[:] + self.numQ_asym[:] + self.numQ_R[:])            
        else:
            return (self.numQ_S[t_idx] + self.numQ_E[t_idx] + self.numQ_pre[t_idx] + self.numQ_sym[t_idx] + self.numQ_asym[t_idx] + self.numQ_R[t_idx])            

#^^^^^^^^^^^^^^^^^^^^^^^^^^^^^^^^^^^^^^^^

    def total_num_tested(self, t_idx=None):
        if(t_idx is None):
            return (self.numTested[:])            
        else:
            return (self.numTested[t_idx])

#^^^^^^^^^^^^^^^^^^^^^^^^^^^^^^^^^^^^^^^^

    def total_num_positive(self, t_idx=None):
        if(t_idx is None):
            return (self.numPositive[:])            
        else:
            return (self.numPositive[t_idx])

#^^^^^^^^^^^^^^^^^^^^^^^^^^^^^^^^^^^^^^^^

    def total_num_recovered(self, t_idx=None):
        if(t_idx is None):
            return (self.numR[:] + self.numQ_R[:])            
        else:
            return (self.numR[t_idx] + self.numQ_R[t_idx])


#^^^^^^^^^^^^^^^^^^^^^^^^^^^^^^^^^^^^^^^^
#^^^^^^^^^^^^^^^^^^^^^^^^^^^^^^^^^^^^^^^^
        
    def calc_propensities(self):

        #~~~~~~~~~~~~~~~~~~~~~~~~~~~~~~~~~~~~~~~~
        # Pre-calculate matrix multiplication terms that may be used in multiple propensity calculations,
        # and check to see if their computation is necessary before doing the multiplication
        #------------------------------------

        self.transmissionTerms_I = numpy.zeros(shape=(self.numNodes,1))      
        if(numpy.any(self.numI_sym[self.tidx]) or numpy.any(self.numI_asym[self.tidx]) or numpy.any(self.numI_pre[self.tidx])):
            if(self.A_deltabeta_asym is not None):
                self.transmissionTerms_sym  = numpy.asarray(scipy.sparse.csr_matrix.dot(self.A_deltabeta, self.X==self.I_sym))
                self.transmissionTerms_asym = numpy.asarray(scipy.sparse.csr_matrix.dot(self.A_deltabeta_asym, ((self.X==self.I_pre)|(self.X==self.I_asym))))
                self.transmissionTerms_I    = self.transmissionTerms_sym+self.transmissionTerms_asym
            else:
                self.transmissionTerms_I    = numpy.asarray(scipy.sparse.csr_matrix.dot(self.A_deltabeta, ((self.X==self.I_sym)|(self.X==self.I_pre)|(self.X==self.I_asym))))

        #------------------------------------

        self.transmissionTerms_Q = numpy.zeros(shape=(self.numNodes,1))
        if(numpy.any(self.numQ_pre[self.tidx]) or numpy.any(self.numQ_sym[self.tidx]) or numpy.any(self.numQ_asym[self.tidx])):
            self.transmissionTerms_Q = numpy.asarray(scipy.sparse.csr_matrix.dot(self.A_Q_deltabeta_Q, ((self.X==self.Q_pre)|(self.X==self.Q_sym)|(self.X==self.Q_asym))))

        #------------------------------------

        self.transmissionTerms_IQ = numpy.zeros(shape=(self.numNodes,1))      
        if(numpy.any(self.numQ_S[self.tidx]) and (numpy.any(self.numI_sym[self.tidx]) or numpy.any(self.numI_asym[self.tidx]) or numpy.any(self.numI_pre[self.tidx]))):
            self.transmissionTerms_IQ = numpy.asarray(scipy.sparse.csr_matrix.dot(self.A_Q_deltabeta_Q, ((self.X==self.I_sym)|(self.X==self.I_pre)|(self.X==self.I_asym))))

        #------------------------------------

        numContacts_Q = numpy.zeros(shape=(self.numNodes,1))
        if(numpy.any(self.positive) and (numpy.any(self.phi_S) or numpy.any(self.phi_E) or numpy.any(self.phi_pre) or numpy.any(self.phi_sym) or numpy.any(self.phi_asym))):
            numContacts_Q = numpy.asarray(scipy.sparse.csr_matrix.dot(self.A, ((self.positive)&(self.X!=self.R)&(self.X!=self.Q_R)&(self.X!=self.F))))

        #~~~~~~~~~~~~~~~~~~~~~~~~~~~~~~~~~~~~~~~~

        propensities_StoE       = ( self.alpha *
                                        (self.o*(self.beta_global*self.prevalence_ext)
                                        + (1-self.o)*(
                                            self.p*((self.beta_global*self.numI_sym[self.tidx] + self.beta_asym_global*(self.numI_pre[self.tidx] + self.numI_asym[self.tidx])
                                            + self.q*self.beta_Q_global*(self.numQ_pre[self.tidx] + self.numQ_sym[self.tidx] + self.numQ_asym[self.tidx]))/self.N[self.tidx])
                                            + (1-self.p)*(numpy.divide(self.transmissionTerms_I, self.degree, out=numpy.zeros_like(self.degree), where=self.degree!=0)
                                                          + numpy.divide(self.transmissionTerms_Q, self.degree_Q, out=numpy.zeros_like(self.degree_Q), where=self.degree_Q!=0))))
                                  )*(self.X==self.S)

        propensities_QStoQE = numpy.zeros_like(propensities_StoE)
        if(numpy.any(self.X==self.Q_S)):
            propensities_QStoQE = ( self.alpha_Q * 
                                        #(self.o*(self.q*self.beta_global*self.prevalence_ext)
                                        (1*(self.beta_global*self.prevalence_ext)
                                        #+ (1-self.o)*(
                                        + (0)*(
                                            self.p*(self.q*(self.beta_global*self.numI_sym[self.tidx] + self.beta_asym_global*(self.numI_pre[self.tidx] + self.numI_asym[self.tidx])
                                            + self.beta_Q_global*(self.numQ_pre[self.tidx] + self.numQ_sym[self.tidx] + self.numQ_asym[self.tidx]))/self.N[self.tidx])
                                            + (1-self.p)*(numpy.divide(self.transmissionTerms_IQ+self.transmissionTerms_Q, self.degree_Q, out=numpy.zeros_like(self.degree_Q), where=self.degree_Q!=0))))
                                   )*(self.X==self.Q_S)

        #~~~~~~~~~~~~~~~~~~~~~~~~~~~~~~~~~~~~~~~~

        if(self.transition_mode == 'time_in_state'):

            propensities_EtoIPRE     = 1e6 * ((self.X==self.E) & numpy.greater(self.timer_state, 1/self.sigma))

            propensities_IPREtoISYM  = 1e6 * ((self.X==self.I_pre) & numpy.greater(self.timer_state, 1/self.lamda) & numpy.greater_equal(self.rand_a, self.a))

            propensities_IPREtoIASYM = 1e6 * ((self.X==self.I_pre) & numpy.greater(self.timer_state, 1/self.lamda) & numpy.less(self.rand_a, self.a))

            propensities_ISYMtoR     = 1e6 * ((self.X==self.I_sym) & numpy.greater(self.timer_state, 1/self.gamma) & numpy.greater_equal(self.rand_h, self.h))

            propensities_ISYMtoH     = 1e6 * ((self.X==self.I_sym) & numpy.greater(self.timer_state, 1/self.eta) & numpy.less(self.rand_h, self.h))

            propensities_IASYMtoR    = 1e6 * ((self.X==self.I_asym) & numpy.greater(self.timer_state, 1/self.gamma))

            #propensities_HtoR        = 1e6 * ((self.X==self.H) & numpy.greater(self.timer_state, 1/self.gamma_H) & numpy.greater_equal(self.rand_f, self.f))
            propensities_HtoR        = 1e6 * ((self.X==self.H) & numpy.greater(self.timer_state, 1e6) & numpy.greater_equal(self.rand_f, self.f))

            #propensities_HtoF        = 1e6 * ((self.X==self.H) & numpy.greater(self.timer_state, 1/self.mu_H) & numpy.less(self.rand_f, self.f))
            propensities_HtoF        = 1e6 * ((self.X==self.H) & numpy.greater(self.timer_state, 1e6) & numpy.less(self.rand_f, self.f))

            propensities_StoQS       = numpy.zeros_like(propensities_StoE)

            propensities_EtoQE       = numpy.zeros_like(propensities_StoE)

            propensities_IPREtoQPRE  = numpy.zeros_like(propensities_StoE)

            propensities_ISYMtoQSYM  = numpy.zeros_like(propensities_StoE)

            propensities_IASYMtoQASYM = numpy.zeros_like(propensities_StoE)

            propensities_QEtoQPRE    = 1e6 * ((self.X==self.Q_E) & numpy.greater(self.timer_state, 1/self.sigma_Q))

            propensities_QPREtoQSYM  = 1e6 * ((self.X==self.Q_pre) & numpy.greater(self.timer_state, 1/self.lamda_Q) & numpy.greater_equal(self.rand_a, self.a))

            propensities_QPREtoQASYM = 1e6 * ((self.X==self.Q_pre) & numpy.greater(self.timer_state, 1/self.lamda_Q) & numpy.less(self.rand_a, self.a))

            propensities_QSYMtoQR    = 1e6 * ((self.X==self.Q_sym) & numpy.greater(self.timer_state, 1/self.gamma_Q_sym) & numpy.greater_equal(self.rand_h, self.h))

            #propensities_QSYMtoH     = 1e6 * ((self.X==self.Q_sym) & numpy.greater(self.timer_state, 1/self.eta_Q) & numpy.less(self.rand_h, self.h))
            propensities_QSYMtoH     = 1e6 * ((self.X==self.Q_sym) & numpy.greater(self.timer_state, 1e6) & numpy.less(self.rand_h, self.h))

            propensities_QASYMtoQR   = 1e6 * ((self.X==self.Q_asym) & numpy.greater(self.timer_state, 1/self.gamma_Q_asym))

            #propensities_RtoS        = 1e6 * ((self.X==self.R) & numpy.greater(self.timer_state, 1/self.xi))
            propensities_RtoS        = 1e6 * ((self.X==self.R) & numpy.greater(self.timer_state, 1e6))

            #propensities__toS        = 1e6 * ((self.X!=self.F) & numpy.greater(self.timer_state, 1/self.nu))
            propensities__toS        = 1e6 * ((self.X!=self.F) & numpy.greater(self.timer_state, 1e6))

        #~~~~~~~~~~~~~~~~~~~~~~~~~~~~~~~~~~~~~~~~

        else: # exponential_rates

            propensities_EtoIPRE     = self.sigma * (self.X==self.E)

            propensities_IPREtoISYM  = self.lamda * ((self.X==self.I_pre) & (numpy.greater_equal(self.rand_a, self.a)))

            propensities_IPREtoIASYM = self.lamda * ((self.X==self.I_pre) & (numpy.less(self.rand_a, self.a)))

            propensities_ISYMtoR     = self.gamma * ((self.X==self.I_sym) & (numpy.greater_equal(self.rand_h, self.h)))

            propensities_ISYMtoH     = self.eta * ((self.X==self.I_sym) & (numpy.less(self.rand_h, self.h)))

            propensities_IASYMtoR    = self.gamma_asym * (self.X==self.I_asym)

            propensities_HtoR        = self.gamma_H * ((self.X==self.H) & (numpy.greater_equal(self.rand_f, self.f)))

            propensities_HtoF        = self.mu_H * ((self.X==self.H) & (numpy.less(self.rand_f, self.f)))

            propensities_StoQS       = (self.theta_S + self.phi_S*numContacts_Q)*self.psi_S * (self.X==self.S)

            propensities_EtoQE       = (self.theta_E + self.phi_E*numContacts_Q)*self.psi_E * (self.X==self.E)

            propensities_IPREtoQPRE  = (self.theta_pre + self.phi_pre*numContacts_Q)*self.psi_pre * (self.X==self.I_pre)

            propensities_ISYMtoQSYM  = (self.theta_sym + self.phi_sym*numContacts_Q)*self.psi_sym * (self.X==self.I_sym)

            propensities_IASYMtoQASYM = (self.theta_asym + self.phi_asym*numContacts_Q)*self.psi_asym * (self.X==self.I_asym)

            propensities_QEtoQPRE    = self.sigma_Q * (self.X==self.Q_E)

            propensities_QPREtoQSYM  = self.lamda_Q * ((self.X==self.Q_pre) & (numpy.greater_equal(self.rand_a, self.a)))

            propensities_QPREtoQASYM = self.lamda_Q * ((self.X==self.Q_pre) & (numpy.less(self.rand_a, self.a)))

            propensities_QSYMtoQR    = self.gamma_Q_sym * ((self.X==self.Q_sym) & (numpy.greater_equal(self.rand_h, self.h)))

            propensities_QSYMtoH     = self.eta_Q * ((self.X==self.Q_sym) & (numpy.less(self.rand_h, self.h)))

            propensities_QASYMtoQR   = self.gamma_Q_asym * (self.X==self.Q_asym)

            propensities_RtoS        = self.xi * (self.X==self.R)

            propensities__toS        = self.nu * (self.X!=self.F)

        #~~~~~~~~~~~~~~~~~~~~~~~~~~~~~~~~~~~~~~~~

        propensities = numpy.hstack([propensities_StoE, propensities_EtoIPRE, propensities_IPREtoISYM, propensities_IPREtoIASYM,
                                     propensities_ISYMtoR, propensities_ISYMtoH, propensities_IASYMtoR, propensities_HtoR, propensities_HtoF, 
                                     propensities_StoQS, propensities_EtoQE, propensities_IPREtoQPRE, propensities_ISYMtoQSYM, propensities_IASYMtoQASYM, 
                                     propensities_QStoQE, propensities_QEtoQPRE, propensities_QPREtoQSYM, propensities_QPREtoQASYM, 
                                     propensities_QSYMtoQR, propensities_QSYMtoH, propensities_QASYMtoQR, propensities_RtoS, propensities__toS])

        columns = [ 'StoE', 'EtoIPRE', 'IPREtoISYM', 'IPREtoIASYM',
                    'ISYMtoR', 'ISYMtoH', 'IASYMtoR', 'HtoR', 'HtoF', 
                    'StoQS', 'EtoQE', 'IPREtoQPRE', 'ISYMtoQSYM', 'IASYMtoQASYM', 
                    'QStoQE', 'QEtoQPRE', 'QPREtoQSYM', 'QPREtoQASYM', 
                    'QSYMtoQR', 'QSYMtoH', 'QASYMtoQR', 'RtoS', '_toS' ]

        return propensities, columns


#^^^^^^^^^^^^^^^^^^^^^^^^^^^^^^^^^^^^^^^^
#^^^^^^^^^^^^^^^^^^^^^^^^^^^^^^^^^^^^^^^^

    def set_isolation(self, node, isolate):
        # Move this node in/out of the appropriate isolation state:
        if(isolate == True):
            if(self.X[node] == self.S):
                self.X[node] = self.Q_S
            elif(self.X[node] == self.E):
                self.X[node] = self.Q_E
            elif(self.X[node] == self.I_pre):
                self.X[node] = self.Q_pre
            elif(self.X[node] == self.I_sym):
                self.X[node] = self.Q_sym
            elif(self.X[node] == self.I_asym):
                self.X[node] = self.Q_asym
            elif(self.X[node] == self.R):
                self.X[node] = self.Q_R
        elif(isolate == False):
            if(self.X[node] == self.Q_S):
                self.X[node] = self.S
            elif(self.X[node] == self.Q_E):
                self.X[node] = self.E
            elif(self.X[node] == self.Q_pre):
                self.X[node] = self.I_pre
            elif(self.X[node] == self.Q_sym):
                self.X[node] = self.I_sym
            elif(self.X[node] == self.Q_asym):
                self.X[node] = self.I_asym
            elif(self.X[node] == self.Q_R):
                self.X[node] = self.R
        # Reset the isolation timer:
        self.timer_isolation[node] = 0

#^^^^^^^^^^^^^^^^^^^^^^^^^^^^^^^^^^^^^^^^

    def set_tested(self, node, tested):
        self.tested[node] = tested
        self.testedInCurrentState[node] = tested

#^^^^^^^^^^^^^^^^^^^^^^^^^^^^^^^^^^^^^^^^

    def set_positive(self, node, positive):
        self.positive[node] = positive

#^^^^^^^^^^^^^^^^^^^^^^^^^^^^^^^^^^^^^^^^

    def introduce_exposures(self, num_new_exposures):
        exposedNodes = numpy.random.choice(range(self.numNodes), size=num_new_exposures, replace=False)
        for exposedNode in exposedNodes:
            if(self.X[exposedNode]==self.S):
                self.X[exposedNode] = self.E
            elif(self.X[exposedNode]==self.Q_S):
                self.X[exposedNode] = self.Q_E


#^^^^^^^^^^^^^^^^^^^^^^^^^^^^^^^^^^^^^^^^
#^^^^^^^^^^^^^^^^^^^^^^^^^^^^^^^^^^^^^^^^   

    def increase_data_series_length(self):
        self.tseries     = numpy.pad(self.tseries, [(0, 6*self.numNodes)], mode='constant', constant_values=0)
        self.numS        = numpy.pad(self.numS, [(0, 6*self.numNodes)], mode='constant', constant_values=0)
        self.numE        = numpy.pad(self.numE, [(0, 6*self.numNodes)], mode='constant', constant_values=0)
        self.numI_pre    = numpy.pad(self.numI_pre, [(0, 6*self.numNodes)], mode='constant', constant_values=0)
        self.numI_sym    = numpy.pad(self.numI_sym, [(0, 6*self.numNodes)], mode='constant', constant_values=0)
        self.numI_asym   = numpy.pad(self.numI_asym, [(0, 6*self.numNodes)], mode='constant', constant_values=0)
        self.numH        = numpy.pad(self.numH, [(0, 6*self.numNodes)], mode='constant', constant_values=0)
        self.numR        = numpy.pad(self.numR, [(0, 6*self.numNodes)], mode='constant', constant_values=0)
        self.numF        = numpy.pad(self.numF, [(0, 6*self.numNodes)], mode='constant', constant_values=0)
        self.numQ_S      = numpy.pad(self.numQ_S, [(0, 6*self.numNodes)], mode='constant', constant_values=0)
        self.numQ_E      = numpy.pad(self.numQ_E, [(0, 6*self.numNodes)], mode='constant', constant_values=0)
        self.numQ_pre    = numpy.pad(self.numQ_pre, [(0, 6*self.numNodes)], mode='constant', constant_values=0)
        self.numQ_sym    = numpy.pad(self.numQ_sym, [(0, 6*self.numNodes)], mode='constant', constant_values=0)
        self.numQ_asym   = numpy.pad(self.numQ_asym, [(0, 6*self.numNodes)], mode='constant', constant_values=0)
        self.numQ_R      = numpy.pad(self.numQ_R, [(0, 6*self.numNodes)], mode='constant', constant_values=0)
        self.N           = numpy.pad(self.N, [(0, 6*self.numNodes)], mode='constant', constant_values=0)
        self.numTested   = numpy.pad(self.numTested, [(0, 6*self.numNodes)], mode='constant', constant_values=0)
        self.numPositive = numpy.pad(self.numPositive, [(0, 6*self.numNodes)], mode='constant', constant_values=0)

        if(self.store_Xseries):
            self.Xseries = numpy.pad(self.Xseries, [(0, 6*self.numNodes), (0,0)], mode='constant', constant_values=0)

        if(self.nodeGroupData):
            for groupName in self.nodeGroupData:
                self.nodeGroupData[groupName]['numS']        = numpy.pad(self.nodeGroupData[groupName]['numS'], [(0, 6*self.numNodes)], mode='constant', constant_values=0)
                self.nodeGroupData[groupName]['numE']        = numpy.pad(self.nodeGroupData[groupName]['numE'], [(0, 6*self.numNodes)], mode='constant', constant_values=0)
                self.nodeGroupData[groupName]['numI_pre']    = numpy.pad(self.nodeGroupData[groupName]['numI_pre'], [(0, 6*self.numNodes)], mode='constant', constant_values=0)
                self.nodeGroupData[groupName]['numI_sym']    = numpy.pad(self.nodeGroupData[groupName]['numI_sym'], [(0, 6*self.numNodes)], mode='constant', constant_values=0)
                self.nodeGroupData[groupName]['numI_asym']   = numpy.pad(self.nodeGroupData[groupName]['numI_asym'], [(0, 6*self.numNodes)], mode='constant', constant_values=0)
                self.nodeGroupData[groupName]['numH']        = numpy.pad(self.nodeGroupData[groupName]['numH'], [(0, 6*self.numNodes)], mode='constant', constant_values=0)
                self.nodeGroupData[groupName]['numR']        = numpy.pad(self.nodeGroupData[groupName]['numR'], [(0, 6*self.numNodes)], mode='constant', constant_values=0)
                self.nodeGroupData[groupName]['numF']        = numpy.pad(self.nodeGroupData[groupName]['numF'], [(0, 6*self.numNodes)], mode='constant', constant_values=0)
                self.nodeGroupData[groupName]['numQ_S']      = numpy.pad(self.nodeGroupData[groupName]['numQ_S'], [(0, 6*self.numNodes)], mode='constant', constant_values=0)
                self.nodeGroupData[groupName]['numQ_E']      = numpy.pad(self.nodeGroupData[groupName]['numQ_E'], [(0, 6*self.numNodes)], mode='constant', constant_values=0)
                self.nodeGroupData[groupName]['numQ_pre']    = numpy.pad(self.nodeGroupData[groupName]['numQ_pre'], [(0, 6*self.numNodes)], mode='constant', constant_values=0)
                self.nodeGroupData[groupName]['numQ_sym']    = numpy.pad(self.nodeGroupData[groupName]['numQ_sym'], [(0, 6*self.numNodes)], mode='constant', constant_values=0)
                self.nodeGroupData[groupName]['numQ_asym']   = numpy.pad(self.nodeGroupData[groupName]['numQ_asym'], [(0, 6*self.numNodes)], mode='constant', constant_values=0)
                self.nodeGroupData[groupName]['numQ_R']      = numpy.pad(self.nodeGroupData[groupName]['numQ_R'], [(0, 6*self.numNodes)], mode='constant', constant_values=0)
                self.nodeGroupData[groupName]['N']           = numpy.pad(self.nodeGroupData[groupName]['N'], [(0, 6*self.numNodes)], mode='constant', constant_values=0)
                self.nodeGroupData[groupName]['numTested']   = numpy.pad(self.nodeGroupData[groupName]['numTested'], [(0, 6*self.numNodes)], mode='constant', constant_values=0)
                self.nodeGroupData[groupName]['numPositive'] = numpy.pad(self.nodeGroupData[groupName]['numPositive'], [(0, 6*self.numNodes)], mode='constant', constant_values=0)

        return None

#^^^^^^^^^^^^^^^^^^^^^^^^^^^^^^^^^^^^^^^^ 

    def finalize_data_series(self):
        self.tseries     = numpy.array(self.tseries, dtype=float)[:self.tidx+1]
        self.numS        = numpy.array(self.numS, dtype=float)[:self.tidx+1]
        self.numE        = numpy.array(self.numE, dtype=float)[:self.tidx+1]
        self.numI_pre    = numpy.array(self.numI_pre, dtype=float)[:self.tidx+1]
        self.numI_sym    = numpy.array(self.numI_sym, dtype=float)[:self.tidx+1]
        self.numI_asym   = numpy.array(self.numI_asym, dtype=float)[:self.tidx+1]
        self.numH        = numpy.array(self.numH, dtype=float)[:self.tidx+1]
        self.numR        = numpy.array(self.numR, dtype=float)[:self.tidx+1]
        self.numF        = numpy.array(self.numF, dtype=float)[:self.tidx+1]
        self.numQ_S      = numpy.array(self.numQ_S, dtype=float)[:self.tidx+1]
        self.numQ_E      = numpy.array(self.numQ_E, dtype=float)[:self.tidx+1]
        self.numQ_pre    = numpy.array(self.numQ_pre, dtype=float)[:self.tidx+1]
        self.numQ_sym    = numpy.array(self.numQ_sym, dtype=float)[:self.tidx+1]
        self.numQ_asym   = numpy.array(self.numQ_asym, dtype=float)[:self.tidx+1]
        self.numQ_R      = numpy.array(self.numQ_R, dtype=float)[:self.tidx+1]
        self.N           = numpy.array(self.N, dtype=float)[:self.tidx+1]
        self.numTested   = numpy.array(self.numTested, dtype=float)[:self.tidx+1]
        self.numPositive = numpy.array(self.numPositive, dtype=float)[:self.tidx+1]

        if(self.store_Xseries):
            self.Xseries = self.Xseries[:self.tidx+1, :]

        if(self.nodeGroupData):
            for groupName in self.nodeGroupData:
                self.nodeGroupData[groupName]['numS']        = numpy.array(self.nodeGroupData[groupName]['numS'], dtype=float)[:self.tidx+1]
                self.nodeGroupData[groupName]['numE']        = numpy.array(self.nodeGroupData[groupName]['numE'], dtype=float)[:self.tidx+1]
                self.nodeGroupData[groupName]['numI_pre']    = numpy.array(self.nodeGroupData[groupName]['numI_pre'], dtype=float)[:self.tidx+1]
                self.nodeGroupData[groupName]['numI_sym']    = numpy.array(self.nodeGroupData[groupName]['numI_sym'], dtype=float)[:self.tidx+1]
                self.nodeGroupData[groupName]['numI_asym']   = numpy.array(self.nodeGroupData[groupName]['numI_asym'], dtype=float)[:self.tidx+1]
                self.nodeGroupData[groupName]['numR']        = numpy.array(self.nodeGroupData[groupName]['numR'], dtype=float)[:self.tidx+1]
                self.nodeGroupData[groupName]['numF']        = numpy.array(self.nodeGroupData[groupName]['numF'], dtype=float)[:self.tidx+1]
                self.nodeGroupData[groupName]['numQ_S']      = numpy.array(self.nodeGroupData[groupName]['numQ_S'], dtype=float)[:self.tidx+1]
                self.nodeGroupData[groupName]['numQ_E']      = numpy.array(self.nodeGroupData[groupName]['numQ_E'], dtype=float)[:self.tidx+1]
                self.nodeGroupData[groupName]['numQ_pre']    = numpy.array(self.nodeGroupData[groupName]['numQ_pre'], dtype=float)[:self.tidx+1]
                self.nodeGroupData[groupName]['numQ_sym']    = numpy.array(self.nodeGroupData[groupName]['numQ_sym'], dtype=float)[:self.tidx+1]
                self.nodeGroupData[groupName]['numQ_asym']   = numpy.array(self.nodeGroupData[groupName]['numQ_asym'], dtype=float)[:self.tidx+1]
                self.nodeGroupData[groupName]['numQ_R']      = numpy.array(self.nodeGroupData[groupName]['numQ_R'], dtype=float)[:self.tidx+1]
                self.nodeGroupData[groupName]['N']           = numpy.array(self.nodeGroupData[groupName]['N'], dtype=float)[:self.tidx+1]
                self.nodeGroupData[groupName]['numTested']   = numpy.array(self.nodeGroupData[groupName]['numTested'], dtype=float)[:self.tidx+1]
                self.nodeGroupData[groupName]['numPositive'] = numpy.array(self.nodeGroupData[groupName]['numPositive'], dtype=float)[:self.tidx+1]

        return None

#^^^^^^^^^^^^^^^^^^^^^^^^^^^^^^^^^^^^^^^^
#^^^^^^^^^^^^^^^^^^^^^^^^^^^^^^^^^^^^^^^^     

    def run_iteration(self, max_dt=None):

        max_dt = self.tmax if max_dt is None else max_dt

        if(self.tidx >= len(self.tseries)-1):
            # Room has run out in the timeseries storage arrays; double the size of these arrays:
            self.increase_data_series_length()

        #~~~~~~~~~~~~~~~~~~~~~~~~~~~~~~~~~~~~~~~~
        # Generate 2 random numbers uniformly distributed in (0,1)
        #~~~~~~~~~~~~~~~~~~~~~~~~~~~~~~~~~~~~~~~~
        r1 = numpy.random.rand()
        r2 = numpy.random.rand()

        #~~~~~~~~~~~~~~~~~~~~~~~~~~~~~~~~~~~~~~~~
        # Calculate propensities
        #~~~~~~~~~~~~~~~~~~~~~~~~~~~~~~~~~~~~~~~~
        propensities, transitionTypes = self.calc_propensities()

        if(propensities.sum() > 0):

            #~~~~~~~~~~~~~~~~~~~~~~~~~~~~~~~~~~~~~~~~
            # Calculate alpha
            #~~~~~~~~~~~~~~~~~~~~~~~~~~~~~~~~~~~~~~~~
            propensities_flat   = propensities.ravel(order='F')
            cumsum              = propensities_flat.cumsum()
            alpha               = propensities_flat.sum()

            #~~~~~~~~~~~~~~~~~~~~~~~~~~~~~~~~~~~~~~~~
            # Compute the time until the next event takes place
            #~~~~~~~~~~~~~~~~~~~~~~~~~~~~~~~~~~~~~~~~
            tau = (1/alpha)*numpy.log(float(1/r1))

            if(tau > max_dt):
                # If the time to next event exceeds the max allowed interval,
                # advance the system time by the max allowed interval,
                # but do not execute any events (recalculate Gillespie interval/event next iteration)
                self.t += max_dt
                self.timer_state += max_dt
                # Update testing and isolation timers/statuses
                isolatedNodes = numpy.argwhere((self.X==self.Q_S)|(self.X==self.Q_E)|(self.X==self.Q_pre)|(self.X==self.Q_sym)|(self.X==self.Q_asym)|(self.X==self.Q_R))[:,0].flatten()
                self.timer_isolation[isolatedNodes] = self.timer_isolation[isolatedNodes] + max_dt
                nodesExitingIsolation = numpy.argwhere((self.timer_isolation >= self.isolationTime) & (self.positive[:,0]==False))
                for isoNode in nodesExitingIsolation:
                    self.set_isolation(node=isoNode, isolate=False)
                # return without any further event execution
                return True
            else:
                self.t += tau
                self.timer_state += tau

            #~~~~~~~~~~~~~~~~~~~~~~~~~~~~~~~~~~~~~~~~
            # Compute which event takes place
            #~~~~~~~~~~~~~~~~~~~~~~~~~~~~~~~~~~~~~~~~
            transitionIdx   = numpy.searchsorted(cumsum,r2*alpha)
            transitionNode  = transitionIdx % self.numNodes
            transitionType  = transitionTypes[ int(transitionIdx/self.numNodes) ]

            #~~~~~~~~~~~~~~~~~~~~~~~~~~~~~~~~~~~~~~~~
            # Perform updates triggered by rate propensities:
            #~~~~~~~~~~~~~~~~~~~~~~~~~~~~~~~~~~~~~~~~
            assert(self.X[transitionNode] == self.transitions[transitionType]['currentState'] and self.X[transitionNode]!=self.F), "Assertion error: Node "+str(transitionNode)+" has unexpected current state "+str(self.X[transitionNode])+" given the intended transition of "+str(transitionType)+"."
            self.X[transitionNode] = self.transitions[transitionType]['newState']

            self.testedInCurrentState[transitionNode] = False

            self.timer_state[transitionNode] = 0.0

            #~~~~~~~~~~~~~~~~~~~~~~~~~~~~~~~~~~~~~~~~

            # Save information about infection events when they occur:
            if(transitionType == 'StoE' or transitionType == 'QStoQE'):
                transitionNode_GNbrs  = list(self.G[transitionNode].keys())
                transitionNode_GQNbrs = list(self.G_Q[transitionNode].keys())
                self.infectionsLog.append({ 't':                            self.t,
                                            'infected_node':                transitionNode,
                                            'infection_type':               transitionType,
                                            'infected_node_degree':         self.degree[transitionNode],
                                            'local_contact_nodes':          transitionNode_GNbrs,
                                            'local_contact_node_states':    self.X[transitionNode_GNbrs].flatten(),
                                            'isolation_contact_nodes':      transitionNode_GQNbrs,
                                            'isolation_contact_node_states':self.X[transitionNode_GQNbrs].flatten() })

            #~~~~~~~~~~~~~~~~~~~~~~~~~~~~~~~~~~~~~~~~

            if(transitionType in ['EtoQE', 'IPREtoQPRE', 'ISYMtoQSYM', 'IASYMtoQASYM', 'ISYMtoH']):
                self.set_positive(node=transitionNode, positive=True)

        #~~~~~~~~~~~~~~~~~~~~~~~~~~~~~~~~~~~~~~~~

        else:

            tau = 0.01
            self.t += tau
            self.timer_state += tau

        #~~~~~~~~~~~~~~~~~~~~~~~~~~~~~~~~~~~~~~~~

        self.tidx += 1
        
        self.tseries[self.tidx]     = self.t
        self.numS[self.tidx]        = numpy.clip(numpy.count_nonzero(self.X==self.S), a_min=0, a_max=self.numNodes)
        self.numE[self.tidx]        = numpy.clip(numpy.count_nonzero(self.X==self.E), a_min=0, a_max=self.numNodes)
        self.numI_pre[self.tidx]    = numpy.clip(numpy.count_nonzero(self.X==self.I_pre), a_min=0, a_max=self.numNodes)
        self.numI_sym[self.tidx]    = numpy.clip(numpy.count_nonzero(self.X==self.I_sym), a_min=0, a_max=self.numNodes)
        self.numI_asym[self.tidx]   = numpy.clip(numpy.count_nonzero(self.X==self.I_asym), a_min=0, a_max=self.numNodes)
        self.numH[self.tidx]        = numpy.clip(numpy.count_nonzero(self.X==self.H), a_min=0, a_max=self.numNodes)
        self.numR[self.tidx]        = numpy.clip(numpy.count_nonzero(self.X==self.R), a_min=0, a_max=self.numNodes)
        self.numF[self.tidx]        = numpy.clip(numpy.count_nonzero(self.X==self.F), a_min=0, a_max=self.numNodes)
        self.numQ_S[self.tidx]      = numpy.clip(numpy.count_nonzero(self.X==self.Q_S), a_min=0, a_max=self.numNodes)
        self.numQ_E[self.tidx]      = numpy.clip(numpy.count_nonzero(self.X==self.Q_E), a_min=0, a_max=self.numNodes)
        self.numQ_pre[self.tidx]    = numpy.clip(numpy.count_nonzero(self.X==self.Q_pre), a_min=0, a_max=self.numNodes)
        self.numQ_sym[self.tidx]    = numpy.clip(numpy.count_nonzero(self.X==self.Q_sym), a_min=0, a_max=self.numNodes)
        self.numQ_asym[self.tidx]   = numpy.clip(numpy.count_nonzero(self.X==self.Q_asym), a_min=0, a_max=self.numNodes)
        self.numQ_R[self.tidx]      = numpy.clip(numpy.count_nonzero(self.X==self.Q_R), a_min=0, a_max=self.numNodes)
        self.numTested[self.tidx]   = numpy.clip(numpy.count_nonzero(self.tested), a_min=0, a_max=self.numNodes)
        self.numPositive[self.tidx] = numpy.clip(numpy.count_nonzero(self.positive), a_min=0, a_max=self.numNodes)
        
        self.N[self.tidx]           = numpy.clip((self.numNodes - self.numF[self.tidx]), a_min=0, a_max=self.numNodes)

        #~~~~~~~~~~~~~~~~~~~~~~~~~~~~~~~~~~~~~~~~
        # Update testing and isolation statuses
        #~~~~~~~~~~~~~~~~~~~~~~~~~~~~~~~~~~~~~~~~

        isolatedNodes = numpy.argwhere((self.X==self.Q_S)|(self.X==self.Q_E)|(self.X==self.Q_pre)|(self.X==self.Q_sym)|(self.X==self.Q_asym)|(self.X==self.Q_R))[:,0].flatten()
        self.timer_isolation[isolatedNodes] = self.timer_isolation[isolatedNodes] + tau

        nodesExitingIsolation = numpy.argwhere((self.timer_isolation >= self.isolationTime) & (self.positive[:,0]==False))
        #print(self.timer_isolation, self.positive[:,0])
        for isoNode in nodesExitingIsolation:
            self.set_isolation(node=isoNode, isolate=False)

        #~~~~~~~~~~~~~~~~~~~~~~~~~~~~~~~~~~~~~~~~
        # Store system states
        #~~~~~~~~~~~~~~~~~~~~~~~~~~~~~~~~~~~~~~~~
        if(self.store_Xseries):
            self.Xseries[self.tidx,:] = self.X.T

        if(self.nodeGroupData):
            for groupName in self.nodeGroupData:
                self.nodeGroupData[groupName]['numS'][self.tidx]        = numpy.count_nonzero(self.nodeGroupData[groupName]['mask']*self.X==self.S)
                self.nodeGroupData[groupName]['numE'][self.tidx]        = numpy.count_nonzero(self.nodeGroupData[groupName]['mask']*self.X==self.E)
                self.nodeGroupData[groupName]['numI_pre'][self.tidx]    = numpy.count_nonzero(self.nodeGroupData[groupName]['mask']*self.X==self.I_pre)
                self.nodeGroupData[groupName]['numI_sym'][self.tidx]    = numpy.count_nonzero(self.nodeGroupData[groupName]['mask']*self.X==self.I_sym)
                self.nodeGroupData[groupName]['numI_asym'][self.tidx]   = numpy.count_nonzero(self.nodeGroupData[groupName]['mask']*self.X==self.I_asym)
                self.nodeGroupData[groupName]['numH'][self.tidx]        = numpy.count_nonzero(self.nodeGroupData[groupName]['mask']*self.X==self.H)
                self.nodeGroupData[groupName]['numR'][self.tidx]        = numpy.count_nonzero(self.nodeGroupData[groupName]['mask']*self.X==self.R)
                self.nodeGroupData[groupName]['numF'][self.tidx]        = numpy.count_nonzero(self.nodeGroupData[groupName]['mask']*self.X==self.F)
                self.nodeGroupData[groupName]['numQ_S'][self.tidx]      = numpy.count_nonzero(self.nodeGroupData[groupName]['mask']*self.X==self.Q_S)
                self.nodeGroupData[groupName]['numQ_E'][self.tidx]      = numpy.count_nonzero(self.nodeGroupData[groupName]['mask']*self.X==self.Q_E)
                self.nodeGroupData[groupName]['numQ_pre'][self.tidx]    = numpy.count_nonzero(self.nodeGroupData[groupName]['mask']*self.X==self.Q_pre)
                self.nodeGroupData[groupName]['numQ_sym'][self.tidx]    = numpy.count_nonzero(self.nodeGroupData[groupName]['mask']*self.X==self.Q_sym)
                self.nodeGroupData[groupName]['numQ_asym'][self.tidx]   = numpy.count_nonzero(self.nodeGroupData[groupName]['mask']*self.X==self.Q_asym)
                self.nodeGroupData[groupName]['numQ_R'][self.tidx]      = numpy.count_nonzero(self.nodeGroupData[groupName]['mask']*self.X==self.Q_R)
                self.nodeGroupData[groupName]['N'][self.tidx]           = numpy.clip((self.nodeGroupData[groupName]['numS'][0] + self.nodeGroupData[groupName]['numE'][0] + self.nodeGroupData[groupName]['numI'][0] + self.nodeGroupData[groupName]['numQ_E'][0] + self.nodeGroupData[groupName]['numQ_I'][0] + self.nodeGroupData[groupName]['numR'][0]), a_min=0, a_max=self.numNodes)
                self.nodeGroupData[groupName]['numTested'][self.tidx]   = numpy.count_nonzero(self.nodeGroupData[groupName]['mask']*self.tested)
                self.nodeGroupData[groupName]['numPositive'][self.tidx] = numpy.count_nonzero(self.nodeGroupData[groupName]['mask']*self.positive)

        #~~~~~~~~~~~~~~~~~~~~~~~~~~~~~~~~~~~~~~~~
        # Terminate if tmax reached or num infections is 0:
        #~~~~~~~~~~~~~~~~~~~~~~~~~~~~~~~~~~~~~~~~
        #if(self.t >= self.tmax or (self.total_num_infected(self.tidx) < 1 and self.total_num_isolated(self.tidx) < 1)):
        if(self.t >= self.tmax):
            self.finalize_data_series()
            return False

        #~~~~~~~~~~~~~~~~~~~~~~~~~~~~~~~~~~~~~~~~

        return True


#^^^^^^^^^^^^^^^^^^^^^^^^^^^^^^^^^^^^^^^^
#^^^^^^^^^^^^^^^^^^^^^^^^^^^^^^^^^^^^^^^^

    def run(self, T, checkpoints=None, max_dt=None, min_dt=None, print_interval=10, verbose='t'):
        if(T>0):
            self.tmax += T
        else:
            return False

        #~~~~~~~~~~~~~~~~~~~~~~~~~~~~~~~~~~~~~~~~
        # Pre-process checkpoint values:
        #~~~~~~~~~~~~~~~~~~~~~~~~~~~~~~~~~~~~~~~~
        if(checkpoints):
            numCheckpoints = len(checkpoints['t'])
            for chkpt_param, chkpt_values in checkpoints.items():
                assert(isinstance(chkpt_values, (list, numpy.ndarray)) and len(chkpt_values)==numCheckpoints), "Expecting a list of values with length equal to number of checkpoint times ("+str(numCheckpoints)+") for each checkpoint parameter."
            checkpointIdx  = numpy.searchsorted(checkpoints['t'], self.t) # Finds 1st index in list greater than given val
            if(checkpointIdx >= numCheckpoints):
                # We are out of checkpoints, stop checking them:
                checkpoints = None 
            else:
                checkpointTime = checkpoints['t'][checkpointIdx]

        #%%%%%%%%%%%%%%%%%%%%%%%%%%%%%%%%%%%%%%%%
        # Run the simulation loop:
        #%%%%%%%%%%%%%%%%%%%%%%%%%%%%%%%%%%%%%%%%
        print_reset = True
        running     = True
        while running:

            running = self.run_iteration()

            #~~~~~~~~~~~~~~~~~~~~~~~~~~~~~~~~~~~~~~~~
            # Handle checkpoints if applicable:
            if(checkpoints):
                if(self.t >= checkpointTime):
                    if(verbose is not False):
                        print("[Checkpoint: Updating parameters]")
                    # A checkpoint has been reached, update param values:
                    for param in list(self.parameters.keys()):
                        if(param in list(checkpoints.keys())):
                            self.parameters.update({param: checkpoints[param][checkpointIdx]})
                    # Update parameter data structures and scenario flags:
                    self.update_parameters()
                    # Update the next checkpoint time:
                    checkpointIdx  = numpy.searchsorted(checkpoints['t'], self.t) # Finds 1st index in list greater than given val
                    if(checkpointIdx >= numCheckpoints):
                        # We are out of checkpoints, stop checking them:
                        checkpoints = None 
                    else:
                        checkpointTime = checkpoints['t'][checkpointIdx]
            #~~~~~~~~~~~~~~~~~~~~~~~~~~~~~~~~~~~~~~~~

            #~~~~~~~~~~~~~~~~~~~~~~~~~~~~~~~~~~~~~~~~

            if(print_interval):
                if(print_reset and (int(self.t) % print_interval == 0)):
                    if(verbose=="t"):
                        print("t = %.2f" % self.t)
                    if(verbose==True):
                        print("t = %.2f" % self.t)
                        print("\t S      = " + str(self.numS[self.tidx]))
                        print("\t E      = " + str(self.numE[self.tidx]))
                        print("\t I_pre  = " + str(self.numI_pre[self.tidx]))
                        print("\t I_sym  = " + str(self.numI_sym[self.tidx]))
                        print("\t I_asym = " + str(self.numI_asym[self.tidx]))
                        print("\t H      = " + str(self.numH[self.tidx]))
                        print("\t R      = " + str(self.numR[self.tidx]))
                        print("\t F      = " + str(self.numF[self.tidx]))
                        print("\t Q_S    = " + str(self.numQ_S[self.tidx]))
                        print("\t Q_E    = " + str(self.numQ_E[self.tidx]))
                        print("\t Q_pre  = " + str(self.numQ_pre[self.tidx]))
                        print("\t Q_sym  = " + str(self.numQ_sym[self.tidx]))
                        print("\t Q_asym = " + str(self.numQ_asym[self.tidx]))
                        print("\t Q_R    = " + str(self.numQ_R[self.tidx]))
                        
                    print_reset = False
                elif(not print_reset and (int(self.t) % 10 != 0)):
                    print_reset = True

        return True


#^^^^^^^^^^^^^^^^^^^^^^^^^^^^^^^^^^^^^^^^
#^^^^^^^^^^^^^^^^^^^^^^^^^^^^^^^^^^^^^^^^

    def plot(self, ax=None, plot_S='line', plot_E='line', plot_I_pre='line', plot_I_sym='line', plot_I_asym='line',
                            plot_H='line', plot_R='line', plot_F='line',
                            plot_Q_E='line', plot_Q_pre='line', plot_Q_sym='line', plot_Q_asym='line', 
                            plot_Q_S='line', plot_Q_R='line', combine_Q_infected=True,
                            color_S='tab:green', color_E='orange', color_I_pre='tomato', color_I_sym='crimson', color_I_asym='#F0909B', 
                            color_H='violet', color_R='tab:blue', color_F='black',
                            color_Q_E='orange', color_Q_pre='tomato', color_Q_sym='crimson', color_Q_asym='#F0909B', 
                            color_Q_S='tab:green',  color_Q_R='tab:blue', color_Q_infected='tab:purple',
                            color_reference='#E0E0E0',
                            dashed_reference_results=None, dashed_reference_label='reference', 
                            shaded_reference_results=None, shaded_reference_label='reference', 
                            vlines=[], vline_colors=[], vline_styles=[], vline_labels=[],
                            ylim=None, xlim=None, legend=True, title=None, side_title=None, plot_percentages=True):

        import matplotlib.pyplot as pyplot

        #~~~~~~~~~~~~~~~~~~~~~~~~~~~~~~~~~~~~~~~~
        # Create an Axes object if None provided:
        #~~~~~~~~~~~~~~~~~~~~~~~~~~~~~~~~~~~~~~~~
        if(not ax):
            fig, ax = pyplot.subplots()

        #~~~~~~~~~~~~~~~~~~~~~~~~~~~~~~~~~~~~~~~~
        # Prepare data series to be plotted:
        #~~~~~~~~~~~~~~~~~~~~~~~~~~~~~~~~~~~~~~~~
        Sseries      = self.numS/self.numNodes if plot_percentages else self.numS
        Eseries      = self.numE/self.numNodes if plot_percentages else self.numE
        I_preseries  = self.numI_pre/self.numNodes if plot_percentages else self.numI_pre 
        I_symseries  = self.numI_sym/self.numNodes if plot_percentages else self.numI_sym
        I_asymseries = self.numI_asym/self.numNodes if plot_percentages else self.numI_asym
        Rseries      = self.numR/self.numNodes if plot_percentages else self.numR
        Hseries      = self.numH/self.numNodes if plot_percentages else self.numH
        Fseries      = self.numF/self.numNodes if plot_percentages else self.numF
        Q_Sseries    = self.numQ_S/self.numNodes if plot_percentages else self.numQ_S 
        Q_Eseries    = self.numQ_E/self.numNodes if plot_percentages else self.numQ_E 
        Q_preseries  = self.numQ_pre/self.numNodes if plot_percentages else self.numQ_pre 
        Q_asymseries = self.numQ_asym/self.numNodes if plot_percentages else self.numQ_asym 
        Q_symseries  = self.numQ_sym/self.numNodes if plot_percentages else self.numQ_sym 
        Q_Rseries    = self.numQ_R/self.numNodes if plot_percentages else self.numQ_R 
        Q_infectedseries = (Q_Eseries + Q_preseries + Q_asymseries + Q_symseries) 

        #~~~~~~~~~~~~~~~~~~~~~~~~~~~~~~~~~~~~~~~~
        # Draw the reference data:      
        #~~~~~~~~~~~~~~~~~~~~~~~~~~~~~~~~~~~~~~~~
        if(dashed_reference_results):
            dashedReference_tseries       = dashed_reference_results.tseries[::int(self.numNodes/100)]
            dashedReference_infectedStack = dashed_reference_results.total_num_infected()[::int(self.numNodes/100)] / (self.numNodes if plot_percentages else 1)
            ax.plot(dashedReference_tseries, dashedReference_infectedStack, color='#E0E0E0', linestyle='--', label='Total infections ('+dashed_reference_label+')', zorder=0)
        if(shaded_reference_results):
            shadedReference_tseries       = shaded_reference_results.tseries
            shadedReference_infectedStack = shaded_reference_results.total_num_infected() / (self.numNodes if plot_percentages else 1)
            ax.fill_between(shaded_reference_results.tseries, shadedReference_infectedStack, 0, color='#EFEFEF', label='Total infections ('+shaded_reference_label+')', zorder=0)
            ax.plot(shaded_reference_results.tseries, shadedReference_infectedStack, color='#E0E0E0', zorder=1)

        #~~~~~~~~~~~~~~~~~~~~~~~~~~~~~~~~~~~~~~~~
        # Draw the stacked variables:
        #~~~~~~~~~~~~~~~~~~~~~~~~~~~~~~~~~~~~~~~~
        topstack = numpy.zeros_like(self.tseries)
        if(any(Fseries) and plot_F=='stacked'):
            ax.fill_between(numpy.ma.masked_where(Fseries<=0, self.tseries), numpy.ma.masked_where(Fseries<=0, topstack+Fseries), topstack, color=color_F, alpha=0.75, label='$F$', zorder=2)
            ax.plot(        numpy.ma.masked_where(Fseries<=0, self.tseries), numpy.ma.masked_where(Fseries<=0, topstack+Fseries),           color=color_F, zorder=3)
            topstack = topstack+Fseries
        
        if(any(Hseries) and plot_H=='stacked'):
            ax.fill_between(numpy.ma.masked_where(Hseries<=0, self.tseries), numpy.ma.masked_where(Hseries<=0, topstack+Hseries), topstack, color=color_H, alpha=0.75, label='$H$', zorder=2)
            ax.plot(        numpy.ma.masked_where(Hseries<=0, self.tseries), numpy.ma.masked_where(Hseries<=0, topstack+Hseries),           color=color_H, zorder=3)
            topstack = topstack+Hseries
        
        if(combine_Q_infected and any(Q_infectedseries) and plot_Q_E=='stacked' and plot_Q_pre=='stacked' and plot_Q_sym=='stacked' and plot_Q_asym=='stacked'):
            ax.fill_between(numpy.ma.masked_where(Q_infectedseries<=0, self.tseries), numpy.ma.masked_where(Q_infectedseries<=0, topstack+Q_infectedseries), topstack, facecolor=color_Q_infected, alpha=0.75, hatch='//////', edgecolor='white', linewidth=0.0, label='$Q_{infected}$', zorder=2)
            ax.plot(        numpy.ma.masked_where(Q_infectedseries<=0, self.tseries), numpy.ma.masked_where(Q_infectedseries<=0, topstack+Q_infectedseries),           color=color_Q_infected, zorder=3)
            topstack = topstack+Q_infectedseries

        if(not combine_Q_infected and any(Q_Eseries) and plot_Q_E=='stacked'):
            ax.fill_between(numpy.ma.masked_where(Q_Eseries<=0, self.tseries), numpy.ma.masked_where(Q_Eseries<=0, topstack+Q_Eseries), topstack, facecolor=color_Q_E, alpha=0.75, hatch='//////', edgecolor='white', linewidth=0.0, label='$Q_E$', zorder=2)
            ax.plot(        numpy.ma.masked_where(Q_Eseries<=0, self.tseries), numpy.ma.masked_where(Q_Eseries<=0, topstack+Q_Eseries),           color=color_Q_E, zorder=3)
            topstack = topstack+Q_Eseries
        
        if(any(Eseries) and plot_E=='stacked'):
            ax.fill_between(numpy.ma.masked_where(Eseries<=0, self.tseries), numpy.ma.masked_where(Eseries<=0, topstack+Eseries), topstack, color=color_E, alpha=0.75, label='$E$', zorder=2)
            ax.plot(        numpy.ma.masked_where(Eseries<=0, self.tseries), numpy.ma.masked_where(Eseries<=0, topstack+Eseries),           color=color_E, zorder=3)
            topstack = topstack+Eseries
        
        if(not combine_Q_infected and any(Q_preseries) and plot_Q_pre=='stacked'):
            ax.fill_between(numpy.ma.masked_where(Q_preseries<=0, self.tseries), numpy.ma.masked_where(Q_preseries<=0, topstack+Q_preseries), topstack, facecolor=color_Q_pre, alpha=0.75,  hatch='//////', edgecolor='white', linewidth=0.0, label='$Q_{pre}$', zorder=2)
            ax.plot(        numpy.ma.masked_where(Q_preseries<=0, self.tseries), numpy.ma.masked_where(Q_preseries<=0, topstack+Q_preseries),           color=color_Q_pre, zorder=3)
            topstack = topstack+Q_preseries

        if(any(I_preseries) and plot_I_pre=='stacked'):
            ax.fill_between(numpy.ma.masked_where(I_preseries<=0, self.tseries), numpy.ma.masked_where(I_preseries<=0, topstack+I_preseries), topstack, color=color_I_pre, alpha=0.75, label='$I_{pre}$', zorder=2)
            ax.plot(        numpy.ma.masked_where(I_preseries<=0, self.tseries), numpy.ma.masked_where(I_preseries<=0, topstack+I_preseries),           color=color_I_pre, zorder=3)
            topstack = topstack+I_preseries
        
        if(not combine_Q_infected and any(Q_symseries) and plot_Q_sym=='stacked'):
            ax.fill_between(numpy.ma.masked_where(Q_symseries<=0, self.tseries), numpy.ma.masked_where(Q_symseries<=0, topstack+Q_symseries), topstack, facecolor=color_Q_sym, alpha=0.75,  hatch='//////', edgecolor='white', linewidth=0.0, label='$Q_{sym}$', zorder=2)
            ax.plot(        numpy.ma.masked_where(Q_symseries<=0, self.tseries), numpy.ma.masked_where(Q_symseries<=0, topstack+Q_symseries),           color=color_Q_sym, zorder=3)
            topstack = topstack+Q_symseries

        if(any(I_symseries) and plot_I_sym=='stacked'):
            ax.fill_between(numpy.ma.masked_where(I_symseries<=0, self.tseries), numpy.ma.masked_where(I_symseries<=0, topstack+I_symseries), topstack, color=color_I_sym, alpha=0.75, label='$I_{sym}$', zorder=2)
            ax.plot(        numpy.ma.masked_where(I_symseries<=0, self.tseries), numpy.ma.masked_where(I_symseries<=0, topstack+I_symseries),           color=color_I_sym, zorder=3)
            topstack = topstack+I_symseries
        
        if(not combine_Q_infected and any(Q_asymseries) and plot_Q_asym=='stacked'):
            ax.fill_between(numpy.ma.masked_where(Q_asymseries<=0, self.tseries), numpy.ma.masked_where(Q_asymseries<=0, topstack+Q_asymseries), topstack, facecolor=color_Q_asym, alpha=0.75,  hatch='//////', edgecolor='white', linewidth=0.0, label='$Q_{asym}$', zorder=2)
            ax.plot(        numpy.ma.masked_where(Q_asymseries<=0, self.tseries), numpy.ma.masked_where(Q_asymseries<=0, topstack+Q_asymseries),           color=color_Q_asym, zorder=3)
            topstack = topstack+Q_asymseries

        if(any(I_asymseries) and plot_I_asym=='stacked'):
            ax.fill_between(numpy.ma.masked_where(I_asymseries<=0, self.tseries), numpy.ma.masked_where(I_asymseries<=0, topstack+I_asymseries), topstack, color=color_I_asym, alpha=0.75, label='$I_{asym}$', zorder=2)
            ax.plot(        numpy.ma.masked_where(I_asymseries<=0, self.tseries), numpy.ma.masked_where(I_asymseries<=0, topstack+I_asymseries),           color=color_I_asym, zorder=3)
            topstack = topstack+I_asymseries
        
        if(any(Q_Rseries) and plot_Q_R=='stacked'):
            ax.fill_between(numpy.ma.masked_where(Q_Rseries<=0, self.tseries), numpy.ma.masked_where(Q_Rseries<=0, topstack+Q_Rseries), topstack, facecolor=color_Q_R, alpha=0.75, hatch='//////', edgecolor='white', linewidth=0.0, label='$Q_R$', zorder=2)
            ax.plot(        numpy.ma.masked_where(Q_Rseries<=0, self.tseries), numpy.ma.masked_where(Q_Rseries<=0, topstack+Q_Rseries),           color=color_Q_R, zorder=3)
            topstack = topstack+Q_Rseries

        if(any(Rseries) and plot_R=='stacked'):
            ax.fill_between(numpy.ma.masked_where(Rseries<=0, self.tseries), numpy.ma.masked_where(Rseries<=0, topstack+Rseries), topstack, color=color_R, alpha=0.75, label='$R$', zorder=2)
            ax.plot(        numpy.ma.masked_where(Rseries<=0, self.tseries), numpy.ma.masked_where(Rseries<=0, topstack+Rseries),           color=color_R, zorder=3)
            topstack = topstack+Rseries
        
        if(any(Q_Sseries) and plot_Q_S=='stacked'):
            ax.fill_between(numpy.ma.masked_where(Q_Sseries<=0, self.tseries), numpy.ma.masked_where(Q_Sseries<=0, topstack+Q_Sseries), topstack, facecolor=color_Q_S, alpha=0.75, hatch='//////', edgecolor='white', linewidth=0.0, label='$Q_S$', zorder=2)
            ax.plot(        numpy.ma.masked_where(Q_Sseries<=0, self.tseries), numpy.ma.masked_where(Q_Sseries<=0, topstack+Q_Sseries),           color=color_Q_S, zorder=3)
            topstack = topstack+Q_Sseries

        if(any(Sseries) and plot_S=='stacked'):
            ax.fill_between(numpy.ma.masked_where(Sseries<=0, self.tseries), numpy.ma.masked_where(Sseries<=0, topstack+Sseries), topstack, color=color_S, alpha=0.75, label='$S$', zorder=2)
            ax.plot(        numpy.ma.masked_where(Sseries<=0, self.tseries), numpy.ma.masked_where(Sseries<=0, topstack+Sseries),           color=color_S, zorder=3)
            topstack = topstack+Sseries
        
        
        #~~~~~~~~~~~~~~~~~~~~~~~~~~~~~~~~~~~~~~~~
        # Draw the shaded variables:
        #~~~~~~~~~~~~~~~~~~~~~~~~~~~~~~~~~~~~~~~~
        if(any(Fseries) and plot_F=='shaded'):
            ax.fill_between(numpy.ma.masked_where(Fseries<=0, self.tseries), numpy.ma.masked_where(Fseries<=0, Fseries), 0, color=color_F, alpha=0.75, label='$F$', zorder=4)
            ax.plot(        numpy.ma.masked_where(Fseries<=0, self.tseries), numpy.ma.masked_where(Fseries<=0, Fseries),    color=color_F, zorder=5)
        
        if(any(Hseries) and plot_H=='shaded'):
            ax.fill_between(numpy.ma.masked_where(Hseries<=0, self.tseries), numpy.ma.masked_where(Hseries<=0, Hseries), 0, color=color_H, alpha=0.75, label='$H$', zorder=4)
            ax.plot(        numpy.ma.masked_where(Hseries<=0, self.tseries), numpy.ma.masked_where(Hseries<=0, Hseries),    color=color_H, zorder=5)
        
        if(combine_Q_infected and any(Q_infectedseries) and plot_Q_E=='shaded' and plot_Q_pre=='shaded' and plot_Q_sym=='shaded' and plot_Q_asym=='shaded'):
            ax.fill_between(numpy.ma.masked_where(Q_infectedseries<=0, self.tseries), numpy.ma.masked_where(Q_infectedseries<=0, Q_infectedseries), 0, color=color_Q_infected, alpha=0.75, hatch='//////', edgecolor='white', linewidth=0.0, label='$Q_{infected}$', zorder=4)
            ax.plot(        numpy.ma.masked_where(Q_infectedseries<=0, self.tseries), numpy.ma.masked_where(Q_infectedseries<=0, Q_infectedseries),    color=color_Q_infected, zorder=5)

        if(not combine_Q_infected and any(Q_Eseries) and plot_Q_E=='shaded'):
            ax.fill_between(numpy.ma.masked_where(Q_Eseries<=0, self.tseries), numpy.ma.masked_where(Q_Eseries<=0, Q_Eseries), 0, facecolor=color_Q_E, alpha=0.75, hatch='//////', edgecolor='white', linewidth=0.0, label='$Q_E$', zorder=4)
            ax.plot(        numpy.ma.masked_where(Q_Eseries<=0, self.tseries), numpy.ma.masked_where(Q_Eseries<=0, Q_Eseries),    color=color_Q_E, zorder=5)
        
        if(any(Eseries) and plot_E=='shaded'):
            ax.fill_between(numpy.ma.masked_where(Eseries<=0, self.tseries), numpy.ma.masked_where(Eseries<=0, Eseries), 0, color=color_E, alpha=0.75, label='$E$', zorder=4)
            ax.plot(        numpy.ma.masked_where(Eseries<=0, self.tseries), numpy.ma.masked_where(Eseries<=0, Eseries),    color=color_E, zorder=5)
        
        if(not combine_Q_infected and any(Q_preseries) and plot_Q_pre=='shaded'):
            ax.fill_between(numpy.ma.masked_where(Q_preseries<=0, self.tseries), numpy.ma.masked_where(Q_preseries<=0, Q_preseries), 0, facecolor=color_Q_pre, alpha=0.75,  hatch='//////', edgecolor='white', linewidth=0.0, label='$Q_{pre}$', zorder=4)
            ax.plot(        numpy.ma.masked_where(Q_preseries<=0, self.tseries), numpy.ma.masked_where(Q_preseries<=0, Q_preseries),    color=color_Q_pre, zorder=5)

        if(any(I_preseries) and plot_I_pre=='shaded'):
            ax.fill_between(numpy.ma.masked_where(I_preseries<=0, self.tseries), numpy.ma.masked_where(I_preseries<=0, I_preseries), 0, color=color_I_pre, alpha=0.75, label='$I_{pre}$', zorder=4)
            ax.plot(        numpy.ma.masked_where(I_preseries<=0, self.tseries), numpy.ma.masked_where(I_preseries<=0, I_preseries),    color=color_I_pre, zorder=5)
        
        if(not combine_Q_infected and any(Q_symseries) and plot_Q_sym=='shaded'):
            ax.fill_between(numpy.ma.masked_where(Q_symseries<=0, self.tseries), numpy.ma.masked_where(Q_symseries<=0, Q_symseries), 0, facecolor=color_Q_sym, alpha=0.75,  hatch='//////', edgecolor='white', linewidth=0.0, label='$Q_{sym}$', zorder=4)
            ax.plot(        numpy.ma.masked_where(Q_symseries<=0, self.tseries), numpy.ma.masked_where(Q_symseries<=0, Q_symseries),    color=color_Q_sym, zorder=5)

        if(any(I_symseries) and plot_I_sym=='shaded'):
            ax.fill_between(numpy.ma.masked_where(I_symseries<=0, self.tseries), numpy.ma.masked_where(I_symseries<=0, I_symseries), 0, color=color_I_sym, alpha=0.75, label='$I_{sym}$', zorder=4)
            ax.plot(        numpy.ma.masked_where(I_symseries<=0, self.tseries), numpy.ma.masked_where(I_symseries<=0, I_symseries),    color=color_I_sym, zorder=5)
        
        if(not combine_Q_infected and any(Q_asymseries) and plot_Q_asym=='shaded'):
            ax.fill_between(numpy.ma.masked_where(Q_asymseries<=0, self.tseries), numpy.ma.masked_where(Q_asymseries<=0, Q_asymseries), 0, facecolor=color_Q_asym, alpha=0.75,  hatch='//////', edgecolor='white', linewidth=0.0, label='$Q_{asym}$', zorder=4)
            ax.plot(        numpy.ma.masked_where(Q_asymseries<=0, self.tseries), numpy.ma.masked_where(Q_asymseries<=0, Q_asymseries),    color=color_Q_asym, zorder=5)

        if(any(I_asymseries) and plot_I_asym=='shaded'):
            ax.fill_between(numpy.ma.masked_where(I_asymseries<=0, self.tseries), numpy.ma.masked_where(I_asymseries<=0, I_asymseries), 0, color=color_I_asym, alpha=0.75, label='$I_{asym}$', zorder=4)
            ax.plot(        numpy.ma.masked_where(I_asymseries<=0, self.tseries), numpy.ma.masked_where(I_asymseries<=0, I_asymseries),    color=color_I_asym, zorder=5)
        
        if(any(Q_Rseries) and plot_Q_R=='shaded'):
            ax.fill_between(numpy.ma.masked_where(Q_Rseries<=0, self.tseries), numpy.ma.masked_where(Q_Rseries<=0, Q_Rseries), 0, facecolor=color_Q_R, alpha=0.75, hatch='//////', edgecolor='white', linewidth=0.0, label='$Q_R$', zorder=4)
            ax.plot(        numpy.ma.masked_where(Q_Rseries<=0, self.tseries), numpy.ma.masked_where(Q_Rseries<=0, Q_Rseries),    color=color_Q_R, zorder=5)

        if(any(Rseries) and plot_R=='shaded'):
            ax.fill_between(numpy.ma.masked_where(Rseries<=0, self.tseries), numpy.ma.masked_where(Rseries<=0, Rseries), 0, color=color_R, alpha=0.75, label='$R$', zorder=4)
            ax.plot(        numpy.ma.masked_where(Rseries<=0, self.tseries), numpy.ma.masked_where(Rseries<=0, Rseries),    color=color_R, zorder=5)

        if(any(Q_Sseries) and plot_Q_S=='shaded'):
            ax.fill_between(numpy.ma.masked_where(Q_Sseries<=0, self.tseries), numpy.ma.masked_where(Q_Sseries<=0, Q_Sseries), 0, facecolor=color_Q_S, alpha=0.75, hatch='//////', edgecolor='white', linewidth=0.0, label='$Q_S$', zorder=4)
            ax.plot(        numpy.ma.masked_where(Q_Sseries<=0, self.tseries), numpy.ma.masked_where(Q_Sseries<=0, Q_Sseries),    color=color_Q_S, zorder=5)
        
        if(any(Sseries) and plot_S=='shaded'):
            ax.fill_between(numpy.ma.masked_where(Sseries<=0, self.tseries), numpy.ma.masked_where(Sseries<=0, Sseries), 0, color=color_S, alpha=0.75, label='$S$', zorder=4)
            ax.plot(        numpy.ma.masked_where(Sseries<=0, self.tseries), numpy.ma.masked_where(Sseries<=0, Sseries),    color=color_S, zorder=5)

        #~~~~~~~~~~~~~~~~~~~~~~~~~~~~~~~~~~~~~~~~
        # Draw the line variables:
        #~~~~~~~~~~~~~~~~~~~~~~~~~~~~~~~~~~~~~~~~
        if(any(Fseries) and plot_F=='line'):
            ax.plot(numpy.ma.masked_where(Fseries<=0, self.tseries), numpy.ma.masked_where(Fseries<=0, Fseries), color=color_F, label='$F$', zorder=6)
        
        if(any(Hseries) and plot_H=='line'):
            ax.plot(numpy.ma.masked_where(Hseries<=0, self.tseries), numpy.ma.masked_where(Hseries<=0, Hseries), color=color_H, label='$H$', zorder=6)
        
        if(combine_Q_infected and any(Q_infectedseries) and plot_Q_E=='line' and plot_Q_pre=='line' and plot_Q_sym=='line' and plot_Q_asym=='line'):
            ax.plot(numpy.ma.masked_where(Q_infectedseries<=0, self.tseries), numpy.ma.masked_where(Q_infectedseries<=0, Q_infectedseries), color=color_Q_infected, label='$Q_{infected}$', zorder=6)

        if(not combine_Q_infected and any(Q_Eseries) and plot_Q_E=='line'):
            ax.plot(numpy.ma.masked_where(Q_Eseries<=0, self.tseries), numpy.ma.masked_where(Q_Eseries<=0, Q_Eseries), color=color_Q_E, label='$Q_E$', zorder=6)
        
        if(any(Eseries) and plot_E=='line'):
            ax.plot(numpy.ma.masked_where(Eseries<=0, self.tseries), numpy.ma.masked_where(Eseries<=0, Eseries), color=color_E, label='$E$', zorder=6)
        
        if(not combine_Q_infected and any(Q_preseries) and plot_Q_pre=='line'):
            ax.plot(numpy.ma.masked_where(Q_preseries<=0, self.tseries), numpy.ma.masked_where(Q_preseries<=0, Q_preseries), color=color_Q_pre, label='$Q_{pre}$', zorder=6)

        if(any(I_preseries) and plot_I_pre=='line'):
            ax.plot(numpy.ma.masked_where(I_preseries<=0, self.tseries), numpy.ma.masked_where(I_preseries<=0, I_preseries), color=color_I_pre, label='$I_{pre}$', zorder=6)
        
        if(not combine_Q_infected and any(Q_symseries) and plot_Q_sym=='line'):
            ax.plot(numpy.ma.masked_where(Q_symseries<=0, self.tseries), numpy.ma.masked_where(Q_symseries<=0, Q_symseries), color=color_Q_sym, label='$Q_{sym}$', zorder=6)

        if(any(I_symseries) and plot_I_sym=='line'):
            ax.plot(numpy.ma.masked_where(I_symseries<=0, self.tseries), numpy.ma.masked_where(I_symseries<=0, I_symseries), color=color_I_sym, label='$I_{sym}$', zorder=6)
        
        if(not combine_Q_infected and any(Q_asymseries) and plot_Q_asym=='line'):
            ax.plot(numpy.ma.masked_where(Q_asymseries<=0, self.tseries), numpy.ma.masked_where(Q_asymseries<=0, Q_asymseries), color=color_Q_asym, label='$Q_{asym}$', zorder=6)

        if(any(I_asymseries) and plot_I_asym=='line'):
            ax.plot(numpy.ma.masked_where(I_asymseries<=0, self.tseries), numpy.ma.masked_where(I_asymseries<=0, I_asymseries), color=color_I_asym, label='$I_{asym}$', zorder=6)
        
        if(any(Q_Rseries) and plot_Q_R=='line'):
            ax.plot(numpy.ma.masked_where(Q_Rseries<=0, self.tseries), numpy.ma.masked_where(Q_Rseries<=0, Q_Rseries), color=color_Q_R, linestyle='--', label='$Q_R$', zorder=6)

        if(any(Rseries) and plot_R=='line'):
            ax.plot(numpy.ma.masked_where(Rseries<=0, self.tseries), numpy.ma.masked_where(Rseries<=0, Rseries), color=color_R, label='$R$', zorder=6)

        if(any(Q_Sseries) and plot_Q_S=='line'):
            ax.plot(numpy.ma.masked_where(Q_Sseries<=0, self.tseries), numpy.ma.masked_where(Q_Sseries<=0, Q_Sseries), color=color_Q_S, linestyle='--', label='$Q_S$', zorder=6)
        
        if(any(Sseries) and plot_S=='line'):
            ax.plot(numpy.ma.masked_where(Sseries<=0, self.tseries), numpy.ma.masked_where(Sseries<=0, Sseries), color=color_S, label='$S$', zorder=6)

        #~~~~~~~~~~~~~~~~~~~~~~~~~~~~~~~~~~~~~~~~
        # Draw the vertical line annotations:
        #~~~~~~~~~~~~~~~~~~~~~~~~~~~~~~~~~~~~~~~~
        if(len(vlines)>0 and len(vline_colors)==0):
            vline_colors = ['gray']*len(vlines)
        if(len(vlines)>0 and len(vline_labels)==0):
            vline_labels = [None]*len(vlines)
        if(len(vlines)>0 and len(vline_styles)==0):
            vline_styles = [':']*len(vlines)
        for vline_x, vline_color, vline_style, vline_label in zip(vlines, vline_colors, vline_styles, vline_labels):
            if(vline_x is not None):
                ax.axvline(x=vline_x, color=vline_color, linestyle=vline_style, alpha=1, label=vline_label)

        #~~~~~~~~~~~~~~~~~~~~~~~~~~~~~~~~~~~~~~~~
        # Draw the plot labels:
        #~~~~~~~~~~~~~~~~~~~~~~~~~~~~~~~~~~~~~~~~
        ax.set_xlabel('days')
        ax.set_ylabel('percent of population' if plot_percentages else 'number of individuals')
        ax.set_xlim(0, (max(self.tseries) if not xlim else xlim))
        ax.set_ylim(0, ylim)
        if(plot_percentages):
            ax.set_yticklabels(['{:,.0%}'.format(y) for y in ax.get_yticks()])
        if(legend):
            legend_handles, legend_labels = ax.get_legend_handles_labels()
            ax.legend(legend_handles[::-1], legend_labels[::-1], loc='upper right', facecolor='white', edgecolor='none', framealpha=0.9, prop={'size': 8})
        if(title):
            ax.set_title(title, size=12)
        if(side_title):
            ax.annotate(side_title, (0, 0.5), xytext=(-45, 0), ha='right', va='center',
                size=12, rotation=90, xycoords='axes fraction', textcoords='offset points')
       
        return ax


#^^^^^^^^^^^^^^^^^^^^^^^^^^^^^^^^^^^^^^^^
#^^^^^^^^^^^^^^^^^^^^^^^^^^^^^^^^^^^^^^^^

    def figure_basic(self,  plot_S='line', plot_E='line', plot_I_pre='line', plot_I_sym='line', plot_I_asym='line',
                            plot_H='line', plot_R='line', plot_F='line',
                            plot_Q_E='line', plot_Q_pre='line', plot_Q_sym='line', plot_Q_asym='line', 
                            plot_Q_S=False, plot_Q_R=False, combine_Q_infected=True,
                            color_S='tab:green', color_E='orange', color_I_pre='tomato', color_I_sym='crimson', color_I_asym='#F0909B', 
                            color_H='violet', color_R='tab:blue', color_F='black',
                            color_Q_E='orange', color_Q_pre='tomato', color_Q_sym='crimson', color_Q_asym='#F0909B', 
                            color_Q_S='tab:green',  color_Q_R='tab:blue', color_Q_infected='tab:purple',
                            color_reference='#E0E0E0',
                            dashed_reference_results=None, dashed_reference_label='reference', 
                            shaded_reference_results=None, shaded_reference_label='reference', 
                            vlines=[], vline_colors=[], vline_styles=[], vline_labels=[],
                            ylim=None, xlim=None, legend=True, title=None, side_title=None, plot_percentages=True,
                            figsize=(12,8), use_seaborn=True, show=True):

        import matplotlib.pyplot as pyplot

        fig, ax = pyplot.subplots(figsize=figsize)

        if(use_seaborn):
            import seaborn
            seaborn.set_style('ticks')
            seaborn.despine()

        self.plot(ax=ax, plot_S=plot_S, plot_E=plot_E, plot_I_pre=plot_I_pre, plot_I_sym=plot_I_sym, plot_I_asym=plot_I_asym,
                        plot_H=plot_H, plot_R=plot_R, plot_F=plot_F,
                        plot_Q_E=plot_Q_E, plot_Q_pre=plot_Q_pre, plot_Q_sym=plot_Q_sym, plot_Q_asym=plot_Q_asym, 
                        plot_Q_S=plot_Q_S, plot_Q_R=plot_Q_R, combine_Q_infected=combine_Q_infected,
                        color_S=color_S, color_E=color_E, color_I_pre=color_I_pre, color_I_sym=color_I_sym, color_I_asym=color_I_asym, 
                        color_H=color_H, color_R=color_R, color_F=color_F,
                        color_Q_E=color_Q_E, color_Q_pre=color_Q_pre, color_Q_sym=color_Q_sym, color_Q_asym=color_Q_asym, 
                        color_Q_S=color_Q_S,  color_Q_R=color_Q_R, color_Q_infected=color_Q_infected,
                        color_reference=color_reference,
                        dashed_reference_results=dashed_reference_results, dashed_reference_label=dashed_reference_label, 
                        shaded_reference_results=shaded_reference_results, shaded_reference_label=shaded_reference_label, 
                        vlines=vlines, vline_colors=vline_colors, vline_styles=vline_styles, vline_labels=vline_labels,
                        ylim=ylim, xlim=xlim, legend=legend, title=title, side_title=side_title, plot_percentages=plot_percentages)

        if(show):
            pyplot.show()

        return fig, ax


#^^^^^^^^^^^^^^^^^^^^^^^^^^^^^^^^^^^^^^^^
#^^^^^^^^^^^^^^^^^^^^^^^^^^^^^^^^^^^^^^^^

    def figure_infections(self, plot_S=False, plot_E='stacked', plot_I_pre='stacked', plot_I_sym='stacked', plot_I_asym='stacked',
                                plot_H='stacked', plot_R=False, plot_F='stacked',
                                plot_Q_E='stacked', plot_Q_pre='stacked', plot_Q_sym='stacked', plot_Q_asym='stacked', 
                                plot_Q_S=False, plot_Q_R=False, combine_Q_infected=True,
                                color_S='tab:green', color_E='orange', color_I_pre='tomato', color_I_sym='crimson', color_I_asym='#F0909B', 
                                color_H='violet', color_R='tab:blue', color_F='black',
                                color_Q_E='orange', color_Q_pre='tomato', color_Q_sym='crimson', color_Q_asym='#F0909B', 
                                color_Q_S='tab:green',  color_Q_R='tab:blue', color_Q_infected='tab:purple',
                                color_reference='#E0E0E0',
                                dashed_reference_results=None, dashed_reference_label='reference', 
                                shaded_reference_results=None, shaded_reference_label='reference', 
                                vlines=[], vline_colors=[], vline_styles=[], vline_labels=[],
                                ylim=None, xlim=None, legend=True, title=None, side_title=None, plot_percentages=True,
                                figsize=(12,8), use_seaborn=True, show=True):

        import matplotlib.pyplot as pyplot

        fig, ax = pyplot.subplots(figsize=figsize)

        if(use_seaborn):
            import seaborn
            seaborn.set_style('ticks')
            seaborn.despine()

        self.plot(ax=ax, plot_S=plot_S, plot_E=plot_E, plot_I_pre=plot_I_pre, plot_I_sym=plot_I_sym, plot_I_asym=plot_I_asym,
                        plot_H=plot_H, plot_R=plot_R, plot_F=plot_F,
                        plot_Q_E=plot_Q_E, plot_Q_pre=plot_Q_pre, plot_Q_sym=plot_Q_sym, plot_Q_asym=plot_Q_asym, 
                        plot_Q_S=plot_Q_S, plot_Q_R=plot_Q_R, combine_Q_infected=combine_Q_infected,
                        color_S=color_S, color_E=color_E, color_I_pre=color_I_pre, color_I_sym=color_I_sym, color_I_asym=color_I_asym, 
                        color_H=color_H, color_R=color_R, color_F=color_F,
                        color_Q_E=color_Q_E, color_Q_pre=color_Q_pre, color_Q_sym=color_Q_sym, color_Q_asym=color_Q_asym, 
                        color_Q_S=color_Q_S,  color_Q_R=color_Q_R, color_Q_infected=color_Q_infected,
                        color_reference=color_reference,
                        dashed_reference_results=dashed_reference_results, dashed_reference_label=dashed_reference_label, 
                        shaded_reference_results=shaded_reference_results, shaded_reference_label=shaded_reference_label, 
                        vlines=vlines, vline_colors=vline_colors, vline_styles=vline_styles, vline_labels=vline_labels, 
                        ylim=ylim, xlim=xlim, legend=legend, title=title, side_title=side_title, plot_percentages=plot_percentages)

        if(show):
            pyplot.show()

        return fig, ax
        

#%%%%%%%%%%%%%%%%%%%%%%%%%%%%%%%%%%%%%%%%
#%%%%%%%%%%%%%%%%%%%%%%%%%%%%%%%%%%%%%%%%
```
