## Supplementary Data 7 for "Effective high-throughput RT-qPCR screening for SARS-CoV-2 infections in children": networks.py

```
from __future__ import division
import numpy
import scipy
import networkx
import FARZ
from models import *

import matplotlib.pyplot as pyplot


def generate_workplace_contact_network(num_cohorts=1, num_nodes_per_cohort=100, num_teams_per_cohort=10,
                                        mean_intracohort_degree=6, pct_contacts_intercohort=0.2,
                                        farz_params={'alpha':5.0, 'gamma':5.0, 'beta':0.5, 'r':1, 'q':0.0, 'phi':10, 
                                                     'b':0, 'epsilon':1e-6, 'directed': False, 'weighted': False},
                                        distancing_scales=[]):

    #~~~~~~~~~~~~~~~~~~~~~~~~~~~~~~~~~~~~~~~~~~~~~~~~~~
    # Generate FARZ networks of intra-cohort contacts:
    #~~~~~~~~~~~~~~~~~~~~~~~~~~~~~~~~~~~~~~~~~~~~~~~~~~

    cohortNetworks = []

    teams_indices = {}

    for i in range(num_cohorts):

        numNodes            = num_nodes_per_cohort[i] if isinstance(num_nodes_per_cohort, list) else num_nodes_per_cohort
        numTeams            = num_teams_per_cohort[i] if isinstance(num_teams_per_cohort, list) else num_teams_per_cohort
        cohortMeanDegree    = mean_intracohort_degree[i] if isinstance(mean_intracohort_degree, list) else mean_intracohort_degree

        farz_params.update({'n':numNodes, 'k':numTeams, 'm':cohortMeanDegree})

        cohortNetwork, cohortTeamLabels = FARZ.generate(farz_params=farz_params)

        cohortNetworks.append(cohortNetwork)

        for node, teams in cohortTeamLabels.items():
            for team in teams:
                try:
                    teams_indices['c'+str(i)+'-t'+str(team)].append(node)
                except KeyError:
                    teams_indices['c'+str(i)+'-t'+str(team)] = [node]    

    #~~~~~~~~~~~~~~~~~~~~~~~~~~~~~~~~~~~~~~~~~~~~~~~~~~
    # Establish inter-cohort contacts:
    #~~~~~~~~~~~~~~~~~~~~~~~~~~~~~~~~~~~~~~~~~~~~~~~~~~

    cohortsAdjMatrices = [networkx.adj_matrix(cohortNetwork) for cohortNetwork in cohortNetworks]

    workplaceAdjMatrix = scipy.sparse.block_diag(cohortsAdjMatrices)
    workplaceNetwork   = networkx.from_scipy_sparse_matrix(workplaceAdjMatrix)

    N = workplaceNetwork.number_of_nodes()

    cohorts_indices = {}
    cohortStartIdx  = -1
    cohortFinalIdx  = -1
    for c, cohortNetwork in enumerate(cohortNetworks):

        cohortStartIdx = cohortFinalIdx + 1
        cohortFinalIdx = cohortStartIdx + cohortNetwork.number_of_nodes() - 1
        cohorts_indices['c'+str(c)] = list(range(cohortStartIdx, cohortFinalIdx))

        for team, indices in teams_indices.items():
            if('c'+str(c) in team):
                teams_indices[team] = [idx+cohortStartIdx for idx in indices]

        for i in list(range(cohortNetwork.number_of_nodes())):
            i_intraCohortDegree = cohortNetwork.degree[i]
            i_interCohortDegree = int( ((1/(1-pct_contacts_intercohort))*i_intraCohortDegree)-i_intraCohortDegree )
            # Add intercohort edges:
            if(len(cohortNetworks) > 1):
                for d in list(range(i_interCohortDegree)):
                    j = numpy.random.choice(list(range(0, cohortStartIdx))+list(range(cohortFinalIdx+1, N)))
                    workplaceNetwork.add_edge(i, j)

    return workplaceNetwork, cohorts_indices, teams_indices


# %%%%%%%%%%%%%%%%%%%%%%%%%%%%%%%%%%%%%%%%
# %%%%%%%%%%%%%%%%%%%%%%%%%%%%%%%%%%%%%%%%


def generate_demographic_contact_network(N, demographic_data, layer_generator='FARZ', layer_info=None, distancing_scales=[], isolation_groups=[], verbose=False):

    graphs = {}

    age_distn               = demographic_data['age_distn']
    household_size_distn    = demographic_data['household_size_distn']
    household_stats         = demographic_data['household_stats']

    #########################################
    # Preprocess Demographic Statistics:
    #########################################
    meanHouseholdSize = numpy.average(list(household_size_distn.keys()), weights=list(household_size_distn.values()))
    # print("mean household size: " + str(meanHouseholdSize))

    # Calculate the distribution of household sizes given that the household has more than 1 member:
    household_size_distn_givenGT1 = {key: value/(1-household_size_distn[1]) for key, value in household_size_distn.items()}
    household_size_distn_givenGT1[1] = 0

    # Percent of households with at least one member under 20:
    pctHouseholdsWithMember_U20          = household_stats['pct_with_under20']
    # Percent of households with at least one member over 60:
    pctHouseholdsWithMember_O60          = household_stats['pct_with_over60']
    # Percent of households with at least one member under 20 AND at least one over 60:
    pctHouseholdsWithMember_U20andO60    = household_stats['pct_with_under20_over60']
    # Percent of SINGLE OCCUPANT households where the occupant is over 60:
    pctHouseholdsWithMember_O60_givenEq1 = household_stats['pct_with_over60_givenSingleOccupant']
    # Average number of members Under 20 in households with at least one member Under 20:
    meanNumU20PerHousehold_givenU20      = household_stats['mean_num_under20_givenAtLeastOneUnder20']

    #~~~~~~~~~~~~~~~~~~~~~~~~~~~~~~~~~~~~~~~~
    # Define major age groups (under 20, between 20-60, over 60), 
    # and calculate age distributions conditional on belonging (or not) to one of these groups:
    #~~~~~~~~~~~~~~~~~~~~~~~~~~~~~~~~~~~~~~~~
    ageBrackets_U20              = ['0-9', '10-19']
    totalPctU20                  = numpy.sum([age_distn[bracket] for bracket in ageBrackets_U20])
    age_distn_givenU20    = {bracket: pct/totalPctU20 for bracket, pct in age_distn.items() if bracket in ageBrackets_U20}

    ageBrackets_20to60           = ['20-29', '30-39', '40-49', '50-59']
    totalPct20to60               = numpy.sum([age_distn[bracket] for bracket in ageBrackets_20to60])
    age_distn_given20to60 = {bracket: pct/totalPct20to60 for bracket, pct in age_distn.items() if bracket in ageBrackets_20to60}

    ageBrackets_O60              = ['60-69', '70-79', '80+']
    totalPctO60                  = numpy.sum([age_distn[bracket] for bracket in ageBrackets_O60])
    age_distn_givenO60    = {bracket: pct/totalPctO60 for bracket, pct in age_distn.items() if bracket in ageBrackets_O60}

    ageBrackets_NOTU20           = ['20-29', '30-39', '40-49', '50-59', '60-69', '70-79', '80+']
    totalPctNOTU20               = numpy.sum([age_distn[bracket] for bracket in ageBrackets_NOTU20])
    age_distn_givenNOTU20 = {bracket: pct/totalPctNOTU20 for bracket, pct in age_distn.items() if bracket in ageBrackets_NOTU20}

    ageBrackets_NOTO60           = ['0-9', '10-19', '20-29', '30-39', '40-49', '50-59']
    totalPctNOTO60               = numpy.sum([age_distn[bracket] for bracket in ageBrackets_NOTO60])
    age_distn_givenNOTO60 = {bracket: pct/totalPctNOTO60 for bracket, pct in age_distn.items() if bracket in ageBrackets_NOTO60}


    #~~~~~~~~~~~~~~~~~~~~~~~~~~~~~~~~~~~~~~~~
    # Calculate the probabilities of a household having members in the major age groups, 
    # conditional on single/multi-occupancy:
    #~~~~~~~~~~~~~~~~~~~~~~~~~~~~~~~~~~~~~~~~
    prob_u20 = pctHouseholdsWithMember_U20    # probability of household having at least 1 member under 20
    prob_o60 = pctHouseholdsWithMember_O60    # probability of household having at least 1 member over 60
    prob_eq1 = household_size_distn[1]         # probability of household having 1 member
    prob_gt1 = 1 - prob_eq1                   # probability of household having greater than 1 member
    householdSituations_prob = {}    
    householdSituations_prob['u20_o60_eq1']       = 0     # can't have both someone under 20 and over 60 in a household with 1 member
    householdSituations_prob['u20_NOTo60_eq1']    = 0     # assume no one under 20 lives on their own (data suggests <1% actually do)
    householdSituations_prob['NOTu20_o60_eq1']    = pctHouseholdsWithMember_O60_givenEq1*prob_eq1
    householdSituations_prob['NOTu20_NOTo60_eq1'] = (1 - pctHouseholdsWithMember_O60_givenEq1)*prob_eq1
    householdSituations_prob['u20_o60_gt1']       = pctHouseholdsWithMember_U20andO60
    householdSituations_prob['u20_NOTo60_gt1']    = prob_u20 - householdSituations_prob['u20_o60_gt1'] - householdSituations_prob['u20_NOTo60_eq1'] - householdSituations_prob['u20_o60_eq1']
    householdSituations_prob['NOTu20_o60_gt1']    = prob_o60 - householdSituations_prob['u20_o60_gt1'] - householdSituations_prob['NOTu20_o60_eq1'] - householdSituations_prob['u20_o60_eq1']
    householdSituations_prob['NOTu20_NOTo60_gt1'] = prob_gt1 - householdSituations_prob['u20_o60_gt1'] - householdSituations_prob['NOTu20_o60_gt1'] - householdSituations_prob['u20_NOTo60_gt1']
    assert(numpy.sum(list(householdSituations_prob.values())) == 1.0), "Household situation probabilities must do not sum to 1"


    #########################################
    #########################################
    # Randomly construct households following the size and age distributions defined above:
    #########################################
    #########################################
    households     = []    # List of dicts storing household data structures and metadata
    homelessNodes  = N     # Number of individuals to place in households
    curMemberIndex = 0
    while(homelessNodes > 0):
        
        household = {}

        household['situation'] = numpy.random.choice(list(householdSituations_prob.keys()), p=list(householdSituations_prob.values()))

        household['ageBrackets'] = []

        if(household['situation'] == 'NOTu20_o60_eq1'): 
            
            #~~~~~~~~~~~~~~~~~~~~~~~~~~~~~~
            # Household size is definitely 1
            household['size'] = 1

            #~~~~~~~~~~~~~~~~~~~~~~~~~~~~~~
            # There is only 1 member in this household, and they are OVER 60; add them:
            household['ageBrackets'].append( numpy.random.choice(list(age_distn_givenO60.keys()), p=list(age_distn_givenO60.values())) )
            
        elif(household['situation'] == 'NOTu20_NOTo60_eq1'):
            
            #~~~~~~~~~~~~~~~~~~~~~~~~~~~~~~
            # Household size is definitely 1
            household['size'] = 1
            
            # There is only 1 member in this household, and they are BETWEEN 20-60; add them:
            household['ageBrackets'].append( numpy.random.choice(list(age_distn_given20to60.keys()), p=list(age_distn_given20to60.values())) )
        
        elif(household['situation'] == 'u20_o60_gt1'):

            #~~~~~~~~~~~~~~~~~~~~~~~~~~~~~~
            # Draw a household size (given the situation, there's at least 2 members):
            household['size'] = min(homelessNodes, max(2, numpy.random.choice(list(household_size_distn_givenGT1), p=list(household_size_distn_givenGT1.values()))) )

            #~~~~~~~~~~~~~~~~~~~~~~~~~~~~~~
            # There's definitely at least one UNDER 20 in this household, add an appropriate age bracket:
            household['ageBrackets'].append( numpy.random.choice(list(age_distn_givenU20.keys()), p=list(age_distn_givenU20.values())) )
            # Figure out how many additional Under 20 to add given there is at least one U20; add them:
            # > Must leave room for at least one Over 60 (see minmax terms)
            numAdditionalU20_givenAtLeastOneU20 = min(max(0, numpy.random.poisson(meanNumU20PerHousehold_givenU20-1)), household['size']-len(household['ageBrackets'])-1)
            for k in range(numAdditionalU20_givenAtLeastOneU20):
                household['ageBrackets'].append( numpy.random.choice(list(age_distn_givenU20.keys()), p=list(age_distn_givenU20.values())) )

            #~~~~~~~~~~~~~~~~~~~~~~~~~~~~~~
            # There's definitely one OVER 60 in this household, add an appropriate age bracket:
            household['ageBrackets'].append( numpy.random.choice(list(age_distn_givenO60.keys()), p=list(age_distn_givenO60.values())) )
            
            #~~~~~~~~~~~~~~~~~~~~~~~~~~~~~~
            # Any remaining members can be any age EXCLUDING Under 20 (all U20s already added):
            for m in range(household['size'] - len(household['ageBrackets'])):
                household['ageBrackets'].append( numpy.random.choice(list(age_distn_givenNOTU20.keys()), p=list(age_distn_givenNOTU20.values())) )

        elif(household['situation'] == 'u20_NOTo60_gt1'): 

            #~~~~~~~~~~~~~~~~~~~~~~~~~~~~~~
            # Draw a household size (given the situation, there's at least 2 members):
            household['size'] = min(homelessNodes, max(2, numpy.random.choice(list(household_size_distn_givenGT1), p=list(household_size_distn_givenGT1.values()))) )

            #~~~~~~~~~~~~~~~~~~~~~~~~~~~~~~
            # There's definitely at least one UNDER 20 in this household, add an appropriate age bracket:
            household['ageBrackets'].append( numpy.random.choice(list(age_distn_givenU20.keys()), p=list(age_distn_givenU20.values())) )
            # Figure out how many additional Under 20 to add given there is at least one U20; add them:
            # > NOT CURRENTLY ASSUMING that there must be at least one non-Under20 member in every household (doing so makes total % U20 in households too low)

            numAdditionalU20_givenAtLeastOneU20 = min(max(0, numpy.random.poisson(meanNumU20PerHousehold_givenU20-1)), household['size']-len(household['ageBrackets']))
            for k in range(numAdditionalU20_givenAtLeastOneU20):
                household['ageBrackets'].append( numpy.random.choice(list(age_distn_givenU20.keys()), p=list(age_distn_givenU20.values())) )

            #~~~~~~~~~~~~~~~~~~~~~~~~~~~~~~
            # There are no OVER 60 in this household.

            #~~~~~~~~~~~~~~~~~~~~~~~~~~~~~~
            # Remaining members can be any age EXCLUDING OVER 60 and EXCLUDING UNDER 20 (all U20s already added):
            for m in range(household['size'] - len(household['ageBrackets'])):
                household['ageBrackets'].append( numpy.random.choice(list(age_distn_given20to60.keys()), p=list(age_distn_given20to60.values())) )

        elif(household['situation'] == 'NOTu20_o60_gt1'): 

            #~~~~~~~~~~~~~~~~~~~~~~~~~~~~~~
            # Draw a household size (given the situation, there's at least 2 members):
            household['size'] = min(homelessNodes, max(2, numpy.random.choice(list(household_size_distn_givenGT1), p=list(household_size_distn_givenGT1.values()))) )

            #~~~~~~~~~~~~~~~~~~~~~~~~~~~~~~
            # There are no UNDER 20 in this household.

            #~~~~~~~~~~~~~~~~~~~~~~~~~~~~~~
            # There's definitely one OVER 60 in this household, add an appropriate age bracket:
            household['ageBrackets'].append( numpy.random.choice(list(age_distn_givenO60.keys()), p=list(age_distn_givenO60.values())) )
            
            #~~~~~~~~~~~~~~~~~~~~~~~~~~~~~~
            # Any remaining members can be any age EXCLUDING UNDER 20:
            for m in range(household['size'] - len(household['ageBrackets'])):
                household['ageBrackets'].append( numpy.random.choice(list(age_distn_givenNOTU20.keys()), p=list(age_distn_givenNOTU20.values())) )

        elif(household['situation'] == 'NOTu20_NOTo60_gt1'):        
        
            #~~~~~~~~~~~~~~~~~~~~~~~~~~~~~~
            # Draw a household size (given the situation, there's at least 2 members):
            household['size'] = min(homelessNodes, max(2, numpy.random.choice(list(household_size_distn_givenGT1), p=list(household_size_distn_givenGT1.values()))) )

            #~~~~~~~~~~~~~~~~~~~~~~~~~~~~~~
            # There are no UNDER 20 in this household.

            #~~~~~~~~~~~~~~~~~~~~~~~~~~~~~~
            # There are no OVER 60 in this household.

            #~~~~~~~~~~~~~~~~~~~~~~~~~~~~~~
            # Remaining household members can be any age BETWEEN 20 TO 60, add as many as needed to meet the household size:
            for m in range(household['size'] - len(household['ageBrackets'])):
                household['ageBrackets'].append( numpy.random.choice(list(age_distn_given20to60.keys()), p=list(age_distn_given20to60.values())) )

        # elif(household['situation'] == 'u20_NOTo60_eq1'): 
        #    impossible by assumption
        # elif(household['situation'] == 'u20_o60_eq1'):    
        #    impossible

        if(len(household['ageBrackets']) == household['size']):

            homelessNodes -= household['size']

            households.append(household)

        else:
            print("Household size does not match number of age brackets assigned. "+household['situation'])


    numHouseholds = len(households)


    #~~~~~~~~~~~~~~~~~~~~~~~~~~~~~~~~~~~~~~~~
    # Check the frequencies of constructed households against the target distributions:
    #~~~~~~~~~~~~~~~~~~~~~~~~~~~~~~~~~~~~~~~~
    print("Generated overall age distribution:")
    for ageBracket in sorted(age_distn):
        age_freq = numpy.sum([len([age for age in household['ageBrackets'] if age==ageBracket]) for household in households])/N
        print(str(ageBracket)+": %.4f\t(%.4f from target)" % (age_freq, (age_freq - age_distn[ageBracket])) )
    print()

    print("Generated household size distribution:")
    for size in sorted(household_size_distn):
        size_freq = numpy.sum([1 for household in households if household['size']==size])/numHouseholds
        print(str(size)+": %.4f\t(%.4f from target)" % (size_freq, (size_freq - household_size_distn[size])) )
    print("Num households: " +str(numHouseholds))
    print("mean household size: " + str(meanHouseholdSize))
    print()
    
    if(verbose):
        print("Generated percent households with at least one member Under 20:")
        checkval = len([household for household in households if not set(household['ageBrackets']).isdisjoint(ageBrackets_U20)])/numHouseholds
        target   = pctHouseholdsWithMember_U20
        print("%.4f\t\t(%.4f from target)" % (checkval, checkval - target))
        
        print("Generated percent households with at least one Over 60")
        checkval = len([household for household in households if not set(household['ageBrackets']).isdisjoint(ageBrackets_O60)])/numHouseholds
        target   = pctHouseholdsWithMember_O60
        print("%.4f\t\t(%.4f from target)" % (checkval, checkval - target))
        
        print("Generated percent households with at least one Under 20 AND Over 60")
        checkval = len([household for household in households if not set(household['ageBrackets']).isdisjoint(ageBrackets_O60) and not set(household['ageBrackets']).isdisjoint(ageBrackets_U20)])/numHouseholds
        target   = pctHouseholdsWithMember_U20andO60
        print("%.4f\t\t(%.4f from target)" % (checkval, checkval - target))
        
        print("Generated percent households with 1 total member who is Over 60")
        checkval = numpy.sum([1 for household in households if household['size']==1 and not set(household['ageBrackets']).isdisjoint(ageBrackets_O60)])/numHouseholds
        target   = pctHouseholdsWithMember_O60_givenEq1*prob_eq1
        print("%.4f\t\t(%.4f from target)" % (checkval, checkval - target))

        print("Generated mean num members Under 20 given at least one member is Under 20")
        checkval = numpy.mean([numpy.in1d(household['ageBrackets'], ageBrackets_U20).sum() for household in households if not set(household['ageBrackets']).isdisjoint(ageBrackets_U20)])
        target   = meanNumU20PerHousehold_givenU20
        print("%.4f\t\t(%.4f from target)" % (checkval, checkval - target))

    #

    #########################################
    #########################################
    # Generate Contact Networks
    #########################################
    #########################################
    
    #########################################
    # Generate baseline (no intervention) contact network:
    #########################################

    #~~~~~~~~~~~~~~~~~~~~~~~~~~~~~~~~~~~~~~~~
    # Define the age groups and desired mean degree for each graph layer:
    #~~~~~~~~~~~~~~~~~~~~~~~~~~~~~~~~~~~~~~~~
    if(layer_info is None):
        # Use the following default data if none is provided:
        # Data source: https://www.medrxiv.org/content/10.1101/2020.03.19.20039107v1
        layer_info  = { '0-9':   {'ageBrackets': ['0-9'],                   'meanDegree': 8.6, 'meanDegree_CI': (0.0, 17.7) },
                        '10-19': {'ageBrackets': ['10-19'],                 'meanDegree': 16.2, 'meanDegree_CI': (12.5, 19.8) },
                        '20-59': {'ageBrackets': ['20-29', '30-39', '40-49', '50-59'], 
                                                    'meanDegree': ((age_distn_given20to60['20-29']+age_distn_given20to60['30-39'])*15.3 + (age_distn_given20to60['40-49']+age_distn_given20to60['50-59'])*13.8), 
                                                    'meanDegree_CI': ( ((age_distn_given20to60['20-29']+age_distn_given20to60['30-39'])*12.6 + (age_distn_given20to60['40-49']+age_distn_given20to60['50-59'])*11.0), ((age_distn_given20to60['20-29']+age_distn_given20to60['30-39'])*17.9 + (age_distn_given20to60['40-49']+age_distn_given20to60['50-59'])*16.6) ) },
                        # '20-39': {'ageBrackets': ['20-29', '30-39'],        'meanDegree': 15.3, 'meanDegree_CI': (12.6, 17.9) },
                        # '40-59': {'ageBrackets': ['40-49', '50-59'],        'meanDegree': 13.8, 'meanDegree_CI': (11.0, 16.6) },
                        '60+':   {'ageBrackets': ['60-69', '70-79', '80+'], 'meanDegree': 13.9, 'meanDegree_CI': (7.3, 20.5) } }

    # Count the number of individuals in each age bracket in the generated households:
    ageBrackets_numInPop = {ageBracket: numpy.sum([len([age for age in household['ageBrackets'] if age==ageBracket]) for household in households])
                            for ageBracket, __ in age_distn.items()}

    #~~~~~~~~~~~~~~~~~~~~~~~~~~~~~~~~~~~~~~~~
    # Generate a graph layer for each age group, representing the public contacts for each age group:
    #~~~~~~~~~~~~~~~~~~~~~~~~~~~~~~~~~~~~~~~~
    adjMatrices           = []
    adjMatrices_isolation_mask = []

    individualAgeGroupLabels = []

    curidx = 0
    for layerGroup, layerInfo in layer_info.items():
        print("Generating graph for "+layerGroup+"...")
        
        layerInfo['numIndividuals'] = numpy.sum([ageBrackets_numInPop[ageBracket] for ageBracket in layerInfo['ageBrackets']])

        layerInfo['indices']        = range(curidx, curidx+layerInfo['numIndividuals'])
        curidx                      += layerInfo['numIndividuals']

        individualAgeGroupLabels[min(layerInfo['indices']):max(layerInfo['indices'])] = [layerGroup]*layerInfo['numIndividuals']

        graph_generated = False
        graph_gen_attempts = 0

        # Note, we generate a graph with average_degree parameter = target mean degree - meanHousehold size
        # so that when in-household edges are added each graph's mean degree will be close to the target mean
        targetMeanDegree = layerInfo['meanDegree']-int(meanHouseholdSize)

        targetMeanDegreeRange = (targetMeanDegree+meanHouseholdSize-0.75, targetMeanDegree+meanHouseholdSize+0.75) if layer_generator=='FARZ' else layerInfo['meanDegree_CI']
        # targetMeanDegreeRange = (targetMeanDegree+meanHouseholdSize-1, targetMeanDegree+meanHouseholdSize+1)

        while(not graph_generated):
            try:
                if(layer_generator == 'LFR'):

                    # print "TARGET MEAN DEGREE     = " + str(targetMeanDegree)
                
                    layerInfo['graph'] = networkx.generators.community.LFR_benchmark_graph(
                                            n=layerInfo['numIndividuals'], 
                                            tau1=3, tau2=2, mu=0.5, 
                                            average_degree=int(targetMeanDegree), 
                                            tol=1e-01, max_iters=200, seed=(None if graph_gen_attempts<10 else int(numpy.random.rand()*1000)))

                elif(layer_generator == 'FARZ'):

                    # https://github.com/rabbanyk/FARZ
                    layerInfo['graph'], layerInfo['communities'] = FARZ.generate(farz_params={ 
                                                                    'n': layerInfo['numIndividuals'], 
                                                                    'm': int(targetMeanDegree/2), # mean degree / 2
                                                                    'k': int(layerInfo['numIndividuals']/50), # num communities
                                                                    'alpha': 2.0,                 # clustering param
                                                                    'gamma': -0.6,                 # assortativity param
                                                                    'beta':  0.6,                 # prob within community edges
                                                                    'r':     1,                  # max num communities node can be part of
                                                                    'q':     0.5,                 # probability of multi-community membership
                                                                    'phi': 1, 'b': 0.0, 'epsilon': 0.0000001, 
                                                                    'directed': False, 'weighted': False})

                elif(layer_generator == 'BA'):
                    pass

                else:
                    print("Layer generator \""+layer_generator+"\" is not recognized (support for 'LFR', 'FARZ', 'BA'")
                
                nodeDegrees = [d[1] for d in layerInfo['graph'].degree()]
                meanDegree  = numpy.mean(nodeDegrees)
                maxDegree   = numpy.max(nodeDegrees)

                # Enforce that the generated graph has mean degree within the 95% CI of the mean for this group in the data:
                if(meanDegree+meanHouseholdSize >= targetMeanDegreeRange[0] and meanDegree+meanHouseholdSize <= targetMeanDegreeRange[1]):
                # if(meanDegree+meanHouseholdSize >= targetMeanDegree+meanHouseholdSize-1 and meanDegree+meanHouseholdSize <= targetMeanDegree+meanHouseholdSize+1):
            
                    if(verbose):
                        print(layerGroup+" public mean degree = "+str((meanDegree)))
                        print(layerGroup+" public max degree  = "+str((maxDegree)))

                    adjMatrices.append(networkx.adj_matrix(layerInfo['graph']))

                    # Create an adjacency matrix mask that will zero out all public edges
                    # for any isolation groups but allow all public edges for other groups:
                    if(layerGroup in isolation_groups):
                        adjMatrices_isolation_mask.append(numpy.zeros(shape=networkx.adj_matrix(layerInfo['graph']).shape))
                    else:
                        # adjMatrices_isolation_mask.append(numpy.ones(shape=networkx.adj_matrix(layerInfo['graph']).shape))
                        # The graph layer we just created represents the baseline (no dist) public connections;
                        # this should be the superset of all connections that exist in any modification of the network,
                        # therefore it should work to use this baseline adj matrix as the mask instead of a block of 1s 
                        # (which uses unnecessary memory to store a whole block of 1s, ie not sparse)
                        adjMatrices_isolation_mask.append(networkx.adj_matrix(layerInfo['graph']))

                    graph_generated = True

                else:
                    graph_gen_attempts += 1
                    if(graph_gen_attempts >= 1):# and graph_gen_attempts % 2):
                        if(meanDegree+meanHouseholdSize < targetMeanDegreeRange[0]):
                            targetMeanDegree += 1 if layer_generator=='FARZ' else 0.05
                        elif(meanDegree+meanHouseholdSize > targetMeanDegreeRange[1]):
                            targetMeanDegree -= 1 if layer_generator=='FARZ' else 0.05
                        # reload(networkx)
                    if(verbose):
                        # print("Try again... (mean degree = "+str(meanDegree)+"+"+str(meanHouseholdSize)+" is outside the target range for mean degree "+str(targetMeanDegreeRange)+")")
                        print("\tTry again... (mean degree = %.2f+%.2f=%.2f is outside the target range for mean degree (%.2f, %.2f)" % (meanDegree, meanHouseholdSize, meanDegree+meanHouseholdSize, targetMeanDegreeRange[0], targetMeanDegreeRange[1]))
            
            # The networks LFR graph generator function has unreliable convergence.
            # If it fails to converge in allotted iterations, try again to generate.
            # If it is stuck (for some reason) and failing many times, reload networkx.
            except networkx.exception.ExceededMaxIterations:
                graph_gen_attempts += 1
                # if(graph_gen_attempts >= 10 and graph_gen_attempts % 10):
                #     reload(networkx)
                if(verbose):
                    print("\tTry again... (networkx failed to converge on a graph)")

    #~~~~~~~~~~~~~~~~~~~~~~~~~~~~~~~~~~~~~~~~
    # Assemble an graph for the full population out of the adjacencies generated for each layer:
    #~~~~~~~~~~~~~~~~~~~~~~~~~~~~~~~~~~~~~~~~
    A_baseline = scipy.sparse.lil_matrix(scipy.sparse.block_diag(adjMatrices))
    # Create a networkx Graph object from the adjacency matrix:
    G_baseline = networkx.from_scipy_sparse_matrix(A_baseline)
    graphs['baseline'] = G_baseline


    #########################################
    # Generate social distancing modifications to the baseline *public* contact network:
    #########################################
    # In-household connections are assumed to be unaffected by social distancing, 
    # and edges will be added to strongly connect households below.
    
    #~~~~~~~~~~~~~~~~~~~~~~~~~~~~~~~~~~~~~~~~
    # Social distancing graphs are generated by randomly drawing (from an exponential distribution) 
    # a number of edges for each node to *keep*, and other edges are removed.
    #~~~~~~~~~~~~~~~~~~~~~~~~~~~~~~~~~~~~~~~~
    G_baseline_NODIST   = graphs['baseline'].copy()
    # Social distancing interactions:
    for dist_scale in distancing_scales:
        graphs['distancingScale'+str(dist_scale)] = custom_exponential_graph(G_baseline_NODIST, scale=dist_scale)

        if(verbose):
            nodeDegrees_baseline_public_DIST    = [d[1] for d in graphs['distancingScale'+str(dist_scale)].degree()]
            print("Distancing Public Degree Pcts:")
            (unique, counts) = numpy.unique(nodeDegrees_baseline_public_DIST, return_counts=True)
            print([str(unique)+": "+str(count/N) for (unique, count) in zip(unique, counts)])
            # pyplot.hist(nodeDegrees_baseline_public_NODIST, bins=range(int(max(nodeDegrees_baseline_public_NODIST))), alpha=0.5, color='tab:blue', label='Public Contacts (no dist)')
            pyplot.hist(nodeDegrees_baseline_public_DIST, bins=range(int(max(nodeDegrees_baseline_public_DIST))), alpha=0.5, color='tab:purple', label='Public Contacts (distancingScale'+str(dist_scale)+')')
            pyplot.xlim(0,40)
            pyplot.xlabel('degree')
            pyplot.ylabel('num nodes')
            pyplot.legend(loc='upper right')
            pyplot.show()


    #########################################
    # Generate modifications to the contact network representing isolation of individuals in specified groups:
    #########################################
    if(len(isolation_groups) > 0):
    
        #~~~~~~~~~~~~~~~~~~~~~~~~~~~~~~~~~~~~~~~~
        # Assemble an adjacency matrix mask (from layer generation step) that will zero out 
        # all public contact edges for the isolation groups but allow all public edges for other groups. 
        #~~~~~~~~~~~~~~~~~~~~~~~~~~~~~~~~~~~~~~~~
        A_isolation_mask    = scipy.sparse.lil_matrix(scipy.sparse.block_diag(adjMatrices_isolation_mask))

        #~~~~~~~~~~~~~~~~~~~~~~~~~~~~~~~~~~~~~~~~
        # Then multiply each distancing graph by this mask to generate the corresponding 
        # distancing adjacency matrices where the isolation groups are isolated (no public edges),
        # and create graphs corresponding to the isolation intervention for each distancing level:
        #~~~~~~~~~~~~~~~~~~~~~~~~~~~~~~~~~~~~~~~~
        for graphName, graph in graphs.items():
            A_withIsolation = scipy.sparse.csr_matrix.multiply( networkx.adj_matrix(graph), A_isolation_mask )
            graphs[graphName+'_isolation'] = networkx.from_scipy_sparse_matrix(A_withIsolation)

    

    #########################################
    #########################################
    # Add edges between housemates to strongly connect households:
    #########################################
    #########################################
    # Apply to all distancing graphs

    # Create a copy of the list of node indices for each age group (graph layer) to draw from:
    for layerGroup, layerInfo in layer_info.items():
        layerInfo['selection_indices'] = list(layerInfo['indices'])

    individualAgeBracketLabels = [None]*N

    # Go through each household, look up what the age brackets of the members should be,
    # and randomly select nodes from corresponding age groups (graph layers) to place in the given household.
    # Strongly connect the nodes selected for each household by adding edges to the adjacency matrix.
    for household in households:
        household['indices'] = []
        for ageBracket in household['ageBrackets']:
            ageGroupIndices = next(layer_info[item]['selection_indices'] for item in layer_info if ageBracket in layer_info[item]["ageBrackets"])
            memberIndex     = ageGroupIndices.pop()
            household['indices'].append(memberIndex)

            individualAgeBracketLabels[memberIndex] = ageBracket

        for memberIdx in household['indices']:
            nonselfIndices = [i for i in household['indices'] if memberIdx!=i]
            for housemateIdx in nonselfIndices:
                # Apply to all distancing graphs
                for graphName, graph in graphs.items():
                    graph.add_edge(memberIdx, housemateIdx)


    #########################################
    # Check the connectivity of the fully constructed contacts graphs for each age group's layer:
    #########################################
    if(verbose):
        for graphName, graph in graphs.items():
            nodeDegrees    = [d[1] for d in graph.degree()]
            meanDegree= numpy.mean(nodeDegrees)
            maxDegree= numpy.max(nodeDegrees)
            components = sorted(networkx.connected_components(graph), key=len, reverse=True)
            numConnectedComps = len(components)
            largestConnectedComp = graph.subgraph(components[0])
            print(graphName+": Overall mean degree = "+str((meanDegree)))
            print(graphName+": Overall max degree = "+str((maxDegree)))
            print(graphName+": number of connected components = {0:d}".format(numConnectedComps))
            print(graphName+": largest connected component = {0:d}".format(len(largestConnectedComp)))
            for layerGroup, layerInfo in layer_info.items():
                nodeDegrees_group = networkx.adj_matrix(graph)[min(layerInfo['indices']):max(layerInfo['indices']), :].sum(axis=1)
                print("\t"+graphName+": "+layerGroup+" final graph mean degree = "+str(numpy.mean(nodeDegrees_group)))
                print("\t"+graphName+": "+layerGroup+" final graph max degree  = "+str(numpy.max(nodeDegrees_group)))
                pyplot.hist(nodeDegrees_group, bins=range(int(max(nodeDegrees_group))), alpha=0.5, label=layerGroup)
            # pyplot.hist(nodeDegrees, bins=range(int(max(nodeDegrees))), alpha=0.5, color='black', label=graphName)
            pyplot.xlim(0,40)
            pyplot.xlabel('degree')
            pyplot.ylabel('num nodes')
            pyplot.legend(loc='upper right')
            pyplot.show()

    #########################################

    return graphs, individualAgeBracketLabels, households


def household_country_data(country):

    if(country=='US'):
        household_data = {
                            'household_size_distn':{ 1: 0.283708848, 
                                                    2: 0.345103011, 
                                                    3: 0.150677793,
                                                    4: 0.127649150, 
                                                    5: 0.057777709, 
                                                    6: 0.022624223, 
                                                    7: 0.012459266  },

                            'age_distn':{'0-9':   0.121,
                                                '10-19': 0.131,
                                                '20-29': 0.137,
                                                '30-39': 0.133,
                                                '40-49': 0.124,
                                                '50-59': 0.131,
                                                '60-69': 0.115,
                                                '70-79': 0.070,
                                                '80+'  : 0.038  },

                            'household_stats':{ 'pct_with_under20':          0.3368, 
                                                'pct_with_over60':           0.3801,
                                                'pct_with_under20_over60':  0.0341,
                                                'pct_with_over60_givenSingleOccupant':       0.110,
                                                'mean_num_under20_givenAtLeastOneUnder20':  1.91 }
                        }

    return household_data

#%%%%%%%%%%%%%%%%%%%%%%%%%%%%%%%%%%%%%%%%
#%%%%%%%%%%%%%%%%%%%%%%%%%%%%%%%%%%%%%%%%


#~~~~~~~~~~~~~~~~~~~~~~~~~~~~~~~~~~~~~~~~~~~~~~~~~~
# Defines a random exponential edge pruning mechanism   
# where the mean degree be easily down-shifted
#~~~~~~~~~~~~~~~~~~~~~~~~~~~~~~~~~~~~~~~~~~~~~~~~~~
def custom_exponential_graph(base_graph=None, scale=100, min_num_edges=0, m=9, n=None):
    # If no base graph is provided, generate a random preferential attachment power law graph as a starting point.
    if(base_graph):
        graph = base_graph.copy()
    else:
        assert(n is not None), "Argument n (number of nodes) must be provided when no base graph is given."
        graph = networkx.barabasi_albert_graph(n=n, m=m)

    # We modify the graph by probabilistically dropping some edges from each node. 
    for node in graph:
        neighbors = list(graph[node].keys())
        if(len(neighbors) > 0):
            quarantineEdgeNum = int( max(min(numpy.random.exponential(scale=scale, size=1), len(neighbors)), min_num_edges) )
            quarantineKeepNeighbors = numpy.random.choice(neighbors, size=quarantineEdgeNum, replace=False)
            for neighbor in neighbors:
                if(neighbor not in quarantineKeepNeighbors):
                    graph.remove_edge(node, neighbor)
    
    return graph

#^^^^^^^^^^^^^^^^^^^^^^^^^^^^^^^^^^^^^^^^^^^^^^^^^^
#^^^^^^^^^^^^^^^^^^^^^^^^^^^^^^^^^^^^^^^^^^^^^^^^^^

def plot_degree_distn(graph, max_degree=None, show=True, use_seaborn=True):
    import matplotlib.pyplot as pyplot
    if(use_seaborn):
        import seaborn
        seaborn.set_style('ticks')
        seaborn.despine()
    # Get a list of the node degrees:
    if type(graph)==numpy.ndarray:
        nodeDegrees = graph.sum(axis=0).reshape((graph.shape[0],1))   # sums of adj matrix cols
    elif type(graph)==networkx.classes.graph.Graph:
        nodeDegrees = [d[1] for d in graph.degree()]
    else:
        raise BaseException("Input an adjacency matrix or networkx object only.")
    # Calculate the mean degree:
    meanDegree = numpy.mean(nodeDegrees)
    # Generate a histogram of the node degrees:
    pyplot.hist(nodeDegrees, bins=range(max(nodeDegrees)), alpha=0.75, color='tab:blue', label=('mean degree = %.1f' % meanDegree))
    pyplot.xlim(0, max(nodeDegrees) if not max_degree else max_degree)
    pyplot.xlabel('degree')
    pyplot.ylabel('num nodes')
    pyplot.legend(loc='upper right')
    if(show):
        pyplot.show()
```
