## Supplementary Data 7 for "Effective high-throughput RT-qPCR screening for SARS-CoV-2 infections in children": sim_loops.py

```
from __future__ import division
import pickle
import numpy

import time


def run_tti_sim(model, T, max_dt=None,
                intervention_start_pct_infected=0, average_introductions_per_day=0,
                testing_cadence='everyday', pct_tested_per_day=1.0, test_falseneg_rate='temporal', 
                testing_compliance_symptomatic=[None], max_pct_tests_for_symptomatics=1.0,
                testing_compliance_traced=[None], max_pct_tests_for_traces=1.0,
                testing_compliance_random=[None], random_testing_degree_bias=0,
                tracing_compliance=[None], num_contacts_to_trace=None, pct_contacts_to_trace=1.0, tracing_lag=1,
                isolation_compliance_symptomatic_individual=[None], isolation_compliance_symptomatic_groupmate=[None], 
                isolation_compliance_positive_individual=[None], isolation_compliance_positive_groupmate=[None],
                isolation_compliance_positive_contact=[None], isolation_compliance_positive_contactgroupmate=[None],
                isolation_lag_symptomatic=1, isolation_lag_positive=1, isolation_lag_contact=0, isolation_groups=None,
                cadence_testing_days=None, cadence_cycle_length=28, temporal_falseneg_rates=None, backlog_skipped_intervals=False,
                print_report = True
                ):

    #~~~~~~~~~~~~~~~~~~~~~~~~~~~~~~~~~~~~~~~~

    # Testing cadences involve a repeating 28 day cycle starting on a Monday
    # (0:Mon, 1:Tue, 2:Wed, 3:Thu, 4:Fri, 5:Sat, 6:Sun, 7:Mon, 8:Tues, ...)
    # For each cadence, testing is done on the day numbers included in the associated list.

    if(cadence_testing_days is None):
        cadence_testing_days    = {
                                    'everyday':     [0, 1, 2, 3, 4, 5, 6, 7, 8, 9, 10, 11, 12, 13, 14, 15, 16, 17, 18, 19, 20, 21, 22, 23, 24, 25, 26, 27],
                                    'workday':      [0, 1, 2, 3, 4, 7, 8, 9, 10, 11, 14, 15, 16, 17, 18, 21, 22, 23, 24, 25],
                                    'triweekly':    [0, 2, 4, 7, 9, 11, 14, 16, 18, 21, 23, 25, 28, 30, 32, 35, 37, 39, 43, 44, 46, 49, 51, 53],
                                    'semiweekly':   [0, 3, 7, 10, 14, 17, 21, 24, 28, 31, 35, 38, 42, 45, 49, 52],
                                    'semiweekly2':  [1, 3, 8, 10, 15, 17, 22, 24, 29, 31, 36, 38, 43, 45, 50, 52],
                                    'weekly':       [0, 7, 14, 21, 28, 35, 42, 49],
                                    'weekly2':      [2, 9, 16, 23, 30, 37, 44, 51],
                                    'biweekly':     [0, 14],
                                    'monthly':      [0],
                                    'cycle_start':  [0],
                                    'never':        [60]
                                }

    #~~~~~~~~~~~~~~~~~~~~~~~~~~~~~~~~~~~~~~~~

    if(temporal_falseneg_rates is None):
        temporal_falseneg_rates = { 
                                    model.E:        {0: 1.00, 1: 0.30, 2: 0.10, 3: 0.10},
                                    model.I_pre:    {0: 0.10, 1: 0.10, 2: 0.10},
                                    model.I_sym:    {0: 0.10, 1: 0.08, 2: 0.08, 3: 0.10, 4: 0.15, 5: 0.22, 6: 0.26, 7: 0.29, 8: 0.34, 9: 0.38, 10: 0.43, 11: 0.48, 12: 0.52, 13: 0.57, 14: 0.62, 15: 0.66, 16: 0.70, 17: 0.76, 18: 0.79, 19: 0.82, 20: 0.85, 21: 0.88, 22: 0.90, 23: 0.92, 24: 0.93, 25: 0.95, 26: 0.96, 27: 0.97, 28: 0.97, 29: 0.98, 30: 0.98, 31: 0.99},
                                    model.I_asym:   {0: 0.10, 1: 0.08, 2: 0.08, 3: 0.10, 4: 0.15, 5: 0.22, 6: 0.26, 7: 0.29, 8: 0.34, 9: 0.38, 10: 0.43, 11: 0.48, 12: 0.52, 13: 0.57, 14: 0.62, 15: 0.66, 16: 0.70, 17: 0.76, 18: 0.79, 19: 0.82, 20: 0.85, 21: 0.88, 22: 0.90, 23: 0.92, 24: 0.93, 25: 0.95, 26: 0.96, 27: 0.97, 28: 0.97, 29: 0.98, 30: 0.98, 31: 0.99},
                                    model.Q_E:      {0: 1.00, 1: 1.00, 2: 1.00, 3: 1.00},
                                    model.Q_pre:    {0: 0.25, 1: 0.25, 2: 0.22},
                                    model.Q_sym:    {0: 0.19, 1: 0.16, 2: 0.16, 3: 0.17, 4: 0.19, 5: 0.22, 6: 0.26, 7: 0.29, 8: 0.34, 9: 0.38, 10: 0.43, 11: 0.48, 12: 0.52, 13: 0.57, 14: 0.62, 15: 0.66, 16: 0.70, 17: 0.76, 18: 0.79, 19: 0.82, 20: 0.85, 21: 0.88, 22: 0.90, 23: 0.92, 24: 0.93, 25: 0.95, 26: 0.96, 27: 0.97, 28: 0.97, 29: 0.98, 30: 0.98, 31: 0.99},
                                    model.Q_asym:   {0: 0.19, 1: 0.16, 2: 0.16, 3: 0.17, 4: 0.19, 5: 0.22, 6: 0.26, 7: 0.29, 8: 0.34, 9: 0.38, 10: 0.43, 11: 0.48, 12: 0.52, 13: 0.57, 14: 0.62, 15: 0.66, 16: 0.70, 17: 0.76, 18: 0.79, 19: 0.82, 20: 0.85, 21: 0.88, 22: 0.90, 23: 0.92, 24: 0.93, 25: 0.95, 26: 0.96, 27: 0.97, 28: 0.97, 29: 0.98, 30: 0.98, 31: 0.99},
                                  }

    #%%%%%%%%%%%%%%%%%%%%%%%%%%%%%%%%%%%%%%%%
    # Custom simulation loop:
    #%%%%%%%%%%%%%%%%%%%%%%%%%%%%%%%%%%%%%%%%

    interventionOn         = False
    interventionStartTime  = None

    timeOfLastIntervention = -1
    timeOfLastIntroduction = -1

    testingDays            = cadence_testing_days[testing_cadence]
    cadenceDayNumber       = 0

    tests_per_day                 = int(model.numNodes * pct_tested_per_day)
    max_tracing_tests_per_day     = int(tests_per_day * max_pct_tests_for_traces)
    max_symptomatic_tests_per_day = int(tests_per_day * max_pct_tests_for_symptomatics)

    tracingPoolQueue              = [[] for i in range(tracing_lag)]
    isolationQueue_symptomatic    = [[] for i in range(isolation_lag_symptomatic)]
    isolationQueue_positive       = [[] for i in range(isolation_lag_positive)]
    isolationQueue_contact        = [[] for i in range(isolation_lag_contact)]

    model.tmax  = T
    running     = True
    while running:

        running = model.run_iteration(max_dt=max_dt)

        #~~~~~~~~~~~~~~~~~~~~~~~~~~~~~~~~~~~~~~~~~~~~~~~~~
        # Introduce exogenous exposures randomly:
        #~~~~~~~~~~~~~~~~~~~~~~~~~~~~~~~~~~~~~~~~~~~~~~~~~
        if(int(model.t)!=int(timeOfLastIntroduction)):

            timeOfLastIntroduction = model.t

            numNewExposures = numpy.random.poisson(lam=average_introductions_per_day)
            
            model.introduce_exposures(num_new_exposures=numNewExposures)

            if(numNewExposures > 0):
                print("[NEW EXPOSURE @ t = %.2f (%d exposed)]" % (model.t, numNewExposures))

        #~~~~~~~~~~~~~~~~~~~~~~~~~~~~~~~~~~~~~~~~~~~~~~~~~
        # Execute testing policy at designated intervals:
        #~~~~~~~~~~~~~~~~~~~~~~~~~~~~~~~~~~~~~~~~~~~~~~~~~

        if(int(model.t)!=int(timeOfLastIntervention)):
        
            cadenceDayNumbers = [int(model.t % cadence_cycle_length)]

            if(backlog_skipped_intervals):
                cadenceDayNumbers = [int(i % cadence_cycle_length) for i in numpy.arange(start=timeOfLastIntervention, stop=int(model.t), step=1.0)[1:]] + cadenceDayNumbers

            timeOfLastIntervention = model.t

            for cadenceDayNumber in cadenceDayNumbers:

                currentNumInfected = model.total_num_infected()[model.tidx]
                currentPctInfected = model.total_num_infected()[model.tidx]/model.numNodes

                if(currentPctInfected >= intervention_start_pct_infected and not interventionOn):
                    interventionOn        = True
                    interventionStartTime = model.t
                
                if(interventionOn):
                    if(print_report):
                        print("[INTERVENTIONS @ t = %.2f (%d (%.2f%%) infected)]" % (model.t, currentNumInfected, currentPctInfected*100))
                    
                    nodeStates                       = model.X.flatten()
                    nodeTestedStatuses               = model.tested.flatten()
                    nodeTestedInCurrentStateStatuses = model.testedInCurrentState.flatten()
                    nodePositiveStatuses             = model.positive.flatten()

                    #~~~~~~~~~~~~~~~~~~~~~~~~~~~~~~~~~~~~~~~~~~~~~~~~~

                    # tracingPoolQueue[0] = tracingPoolQueue[0]Queue.pop(0)

                    #~~~~~~~~~~~~~~~~~~~~~~~~~~~~~~~~~~~~~~~~~~~~~~~~~
                    
                    newIsolationGroup_symptomatic = []
                    newIsolationGroup_contact     = []

                    #----------------------------------------
                    # Isolate SYMPTOMATIC cases without a test:
                    #----------------------------------------
                    numSelfIsolated_symptoms = 0
                    numSelfIsolated_symptomaticGroupmate = 0

                    if(any(isolation_compliance_symptomatic_individual)):
                        symptomaticNodes = numpy.argwhere((nodeStates==model.I_sym)).flatten()
                        for symptomaticNode in symptomaticNodes:
                            if(isolation_compliance_symptomatic_individual[symptomaticNode]):
                                if(model.X[symptomaticNode] == model.I_sym):
                                    numSelfIsolated_symptoms += 1   
                                    newIsolationGroup_symptomatic.append(symptomaticNode)

                                #----------------------------------------
                                # Isolate the GROUPMATES of this SYMPTOMATIC node without a test:
                                #----------------------------------------
                                if(isolation_groups is not None and any(isolation_compliance_symptomatic_groupmate)):
                                    isolationGroupmates = next((group for group in isolation_groups if symptomaticNode in group), None)
                                    for isolationGroupmate in isolationGroupmates:
                                        if(isolationGroupmate != symptomaticNode):
                                            if(isolation_compliance_symptomatic_groupmate[isolationGroupmate]):
                                                numSelfIsolated_symptomaticGroupmate += 1
                                                newIsolationGroup_symptomatic.append(isolationGroupmate)


                    #----------------------------------------
                    # Isolate the CONTACTS of detected POSITIVE cases without a test:
                    #----------------------------------------
                    numSelfIsolated_positiveContact = 0
                    numSelfIsolated_positiveContactGroupmate = 0

                    if(any(isolation_compliance_positive_contact) or any(isolation_compliance_positive_contactgroupmate)):
                        for contactNode in tracingPoolQueue[0]:
                            if(isolation_compliance_positive_contact[contactNode]):
                                newIsolationGroup_contact.append(contactNode)
                                numSelfIsolated_positiveContact += 1 

                            #----------------------------------------
                            # Isolate the GROUPMATES of this self-isolating CONTACT without a test:
                            #----------------------------------------
                            if(isolation_groups is not None and any(isolation_compliance_positive_contactgroupmate)):
                                isolationGroupmates = next((group for group in isolation_groups if contactNode in group), None)
                                for isolationGroupmate in isolationGroupmates:
                                    # if(isolationGroupmate != contactNode):
                                    if(isolation_compliance_positive_contactgroupmate[isolationGroupmate]):
                                        newIsolationGroup_contact.append(isolationGroupmate)
                                        numSelfIsolated_positiveContactGroupmate += 1
                                        

                    #----------------------------------------
                    # Update the nodeStates list after self-isolation updates to model.X:
                    #----------------------------------------
                    nodeStates = model.X.flatten()


                    #~~~~~~~~~~~~~~~~~~~~~~~~~~~~~~~~~~~~~~~~~~~~~~~~~


                    #----------------------------------------
                    # Allow SYMPTOMATIC individuals to self-seek tests
                    # regardless of cadence testing days
                    #----------------------------------------
                    symptomaticSelection = []

                    if(any(testing_compliance_symptomatic)):
                        
                        symptomaticPool = numpy.argwhere((testing_compliance_symptomatic==True)
                                                         & (nodeTestedInCurrentStateStatuses==False)
                                                         & (nodePositiveStatuses==False)
                                                         & ((nodeStates==model.I_sym)|(nodeStates==model.Q_sym))
                                                        ).flatten()

                        numSymptomaticTests  = min(len(symptomaticPool), max_symptomatic_tests_per_day)
                        
                        if(len(symptomaticPool) > 0):
                            symptomaticSelection = symptomaticPool[numpy.random.choice(len(symptomaticPool), min(numSymptomaticTests, len(symptomaticPool)), replace=False)]


                    #----------------------------------------
                    # Test individuals randomly and via contact tracing
                    # on cadence testing days:
                    #----------------------------------------

                    tracingSelection = []
                    randomSelection = []

                    if(cadenceDayNumber in testingDays):

                        #----------------------------------------
                        # Apply a designated portion of this day's tests 
                        # to individuals identified by CONTACT TRACING:
                        #----------------------------------------

                        tracingPool = tracingPoolQueue.pop(0)

                        if(any(testing_compliance_traced)):

                            numTracingTests = min(len(tracingPool), min(tests_per_day-len(symptomaticSelection), max_tracing_tests_per_day))

                            for trace in range(numTracingTests):
                                traceNode = tracingPool.pop()
                                if((nodePositiveStatuses[traceNode]==False)
                                    and (testing_compliance_traced[traceNode]==True)
                                    and (model.X[traceNode] != model.R)
                                    and (model.X[traceNode] != model.Q_R) 
                                    and (model.X[traceNode] != model.H)
                                    and (model.X[traceNode] != model.F)):
                                    tracingSelection.append(traceNode)

                        #----------------------------------------
                        # Apply the remainder of this day's tests to random testing:
                        #----------------------------------------

                        if(any(testing_compliance_random)):
                            
                            testingPool = numpy.argwhere((testing_compliance_random==True)
                                                         & (nodePositiveStatuses==False)
                                                         & (nodeStates != model.Q_S) 
                                                         & (nodeStates != model.Q_E)
                                                         & (nodeStates != model.Q_pre)
                                                         & (nodeStates != model.Q_sym)
                                                         & (nodeStates != model.Q_asym)
                                                         & (nodeStates != model.Q_R)
                                                         & (nodeStates != model.F)
                                                        ).flatten()

                            numRandomTests = max(min(tests_per_day-len(tracingSelection)-len(symptomaticSelection), len(testingPool)), 0)
                            
                            testingPool_degrees       = model.degree.flatten()[testingPool]
                            testingPool_degreeWeights = numpy.power(testingPool_degrees,random_testing_degree_bias)/numpy.sum(numpy.power(testingPool_degrees,random_testing_degree_bias))

                            if(len(testingPool) > 0):
                                randomSelection = testingPool[numpy.random.choice(len(testingPool), numRandomTests, p=testingPool_degreeWeights, replace=False)]

                    
                    #~~~~~~~~~~~~~~~~~~~~~~~~~~~~~~~~~~~~~~~~~~~~~~~~~


                    #----------------------------------------
                    # Perform the tests on the selected individuals:
                    #----------------------------------------

                    selectedToTest = numpy.concatenate((symptomaticSelection, tracingSelection, randomSelection)).astype(int)

                    numTested                     = 0
                    numTested_random              = 0
                    numTested_tracing             = 0
                    numTested_symptomatic         = 0
                    numPositive                   = 0
                    numPositive_random            = 0
                    numPositive_tracing           = 0
                    numPositive_symptomatic       = 0 
                    numIsolated_positiveGroupmate = 0
                    
                    newTracingPool = []

                    newIsolationGroup_positive = []

                    for i, testNode in enumerate(selectedToTest):

                        model.set_tested(testNode, True)

                        numTested += 1
                        if(i < len(symptomaticSelection)):
                            numTested_symptomatic  += 1
                        elif(i < len(symptomaticSelection)+len(tracingSelection)):
                            numTested_tracing += 1
                        else:
                            numTested_random += 1                  

                        # If the node to be tested is not infected, then the test is guaranteed negative, 
                        # so don't bother going through with doing the test:
                        if(model.X[testNode] == model.S or model.X[testNode] == model.Q_S):
                            pass
                        # Also assume that latent infections are not picked up by tests:
                        elif(model.X[testNode] == model.E or model.X[testNode] == model.Q_E):
                            pass
                        elif(model.X[testNode] == model.I_pre or model.X[testNode] == model.Q_pre 
                             or model.X[testNode] == model.I_sym or model.X[testNode] == model.Q_sym 
                             or model.X[testNode] == model.I_asym or model.X[testNode] == model.Q_asym):
                            
                            if(test_falseneg_rate == 'temporal'):
                                testNodeState       = model.X[testNode][0]
                                testNodeTimeInState = model.timer_state[testNode][0]
                                if(testNodeState in list(temporal_falseneg_rates.keys())):
                                    falseneg_prob = temporal_falseneg_rates[testNodeState][ int(min(testNodeTimeInState, max(list(temporal_falseneg_rates[testNodeState].keys())))) ]
                                else:
                                    falseneg_prob = 1.00
                            else:
                                falseneg_prob = test_falseneg_rate

                            if(numpy.random.rand() < (1-falseneg_prob)):
                                # +++++++++++++++++++++++++++++++++++++++++++++
                                # The tested node has returned a positive test
                                # +++++++++++++++++++++++++++++++++++++++++++++
                                numPositive += 1
                                if(i < len(symptomaticSelection)):
                                    numPositive_symptomatic  += 1
                                elif(i < len(symptomaticSelection)+len(tracingSelection)):
                                    numPositive_tracing += 1
                                else:
                                    numPositive_random += 1 
                                
                                # Update the node's state to the appropriate detected case state:
                                model.set_positive(testNode, True)

                                #----------------------------------------
                                # Add this positive node to the isolation group:
                                #----------------------------------------
                                if(isolation_compliance_positive_individual[testNode]):
                                    newIsolationGroup_positive.append(testNode)

                                #----------------------------------------
                                # Add the groupmates of this positive node to the isolation group:
                                #----------------------------------------  
                                if(isolation_groups is not None and any(isolation_compliance_positive_groupmate)):
                                    isolationGroupmates = next((group for group in isolation_groups if testNode in group), None)
                                    for isolationGroupmate in isolationGroupmates:
                                        if(isolationGroupmate != testNode):
                                            if(isolation_compliance_positive_groupmate[isolationGroupmate]):
                                                numIsolated_positiveGroupmate += 1
                                                newIsolationGroup_positive.append(isolationGroupmate)

                                #----------------------------------------  
                                # Add this node's neighbors to the contact tracing pool:
                                #----------------------------------------  
                                if(any(tracing_compliance) or any(isolation_compliance_positive_contact) or any(isolation_compliance_positive_contactgroupmate)):
                                    if(tracing_compliance[testNode]):
                                        testNodeContacts = list(model.G[testNode].keys())
                                        numpy.random.shuffle(testNodeContacts)
                                        if(num_contacts_to_trace is None):
                                            numContactsToTrace = int(pct_contacts_to_trace*len(testNodeContacts))
                                        else:
                                            numContactsToTrace = num_contacts_to_trace
                                        newTracingPool.extend(testNodeContacts[0:numContactsToTrace])

            
                    # Add the nodes to be isolated to the isolation queue:
                    isolationQueue_positive.append(newIsolationGroup_positive)
                    isolationQueue_symptomatic.append(newIsolationGroup_symptomatic)
                    isolationQueue_contact.append(newIsolationGroup_contact)

                    # Add the nodes to be traced to the tracing queue:
                    tracingPoolQueue.append(newTracingPool)

                    if(print_report):
                        print("\t"+str(numTested_symptomatic) +"\ttested due to symptoms  [+ "+str(numPositive_symptomatic)+" positive (%.2f %%) +]" % (numPositive_symptomatic/numTested_symptomatic*100 if numTested_symptomatic>0 else 0))
                        print("\t"+str(numTested_tracing)     +"\ttested as traces        [+ "+str(numPositive_tracing)+" positive (%.2f %%) +]" % (numPositive_tracing/numTested_tracing*100 if numTested_tracing>0 else 0))            
                        print("\t"+str(numTested_random)      +"\ttested randomly         [+ "+str(numPositive_random)+" positive (%.2f %%) +]" % (numPositive_random/numTested_random*100 if numTested_random>0 else 0))            
                        print("\t"+str(numTested)             +"\ttested TOTAL            [+ "+str(numPositive)+" positive (%.2f %%) +]" % (numPositive/numTested*100 if numTested>0 else 0))           

                        print("\t"+str(numSelfIsolated_symptoms)        +" will isolate due to symptoms         ("+str(numSelfIsolated_symptomaticGroupmate)+" as groupmates of symptomatic)")
                        print("\t"+str(numPositive)                     +" will isolate due to positive test    ("+str(numIsolated_positiveGroupmate)+" as groupmates of positive)")
                        print("\t"+str(numSelfIsolated_positiveContact) +" will isolate due to positive contact ("+str(numSelfIsolated_positiveContactGroupmate)+" as groupmates of contact)")

                    #----------------------------------------
                    # Update the status of nodes who are to be isolated:
                    #----------------------------------------

                    numIsolated = 0

                    isolationGroup_symptomatic = isolationQueue_symptomatic.pop(0)
                    for isolationNode in isolationGroup_symptomatic:
                        model.set_isolation(isolationNode, True)
                        numIsolated += 1

                    isolationGroup_contact = isolationQueue_contact.pop(0)
                    for isolationNode in isolationGroup_contact:
                        model.set_isolation(isolationNode, True)
                        numIsolated += 1

                    isolationGroup_positive = isolationQueue_positive.pop(0)
                    for isolationNode in isolationGroup_positive:
                        model.set_isolation(isolationNode, True)
                        numIsolated += 1

                    if(print_report):
                        print("\t"+str(numIsolated)+" entered isolation")
                    
                #~~~~~~~~~~~~~~~~~~~~~~~~~~~~~~~~~~~~~~~~

        #~~~~~~~~~~~~~~~~~~~~~~~~~~~~~~~~~~~~~~~~

    interventionInterval = (interventionStartTime, model.t)

    return interventionInterval


#%%%%%%%%%%%%%%%%%%%%%%%%%%%%%%%%%%%%%%%%%%%%%%%%%%
#%%%%%%%%%%%%%%%%%%%%%%%%%%%%%%%%%%%%%%%%%%%%%%%%%%
```
