## Supplementary Data 7 for "Effective high-throughput RT-qPCR screening for SARS-CoV-2 infections in children": utilities.py

```
import numpy
import matplotlib.pyplot as pyplot

def gamma_dist(mean, coeffvar, N):
	scale = mean*coeffvar**2
	shape = mean/scale
	return numpy.random.gamma(scale=scale, shape=shape, size=N)


def dist_info(dists, names=None, plot=False, bin_size=1, colors=None, reverse_plot=False):
    dists  = [dists] if not isinstance(dists, list) else dists
    names  = [names] if(names is not None and not isinstance(names, list)) else (names if names is not None else [None]*len(dists))
    colors = [colors] if(colors is not None and not isinstance(colors, list)) else (colors if colors is not None else pyplot.rcParams['axes.prop_cycle'].by_key()['color'])
    
    for i, (dist, name) in enumerate(zip(dists, names)):
        print((name+": " if name else "")+" mean = %.2f, std = %.2f, 95%% CI = (%.2f, %.2f)" % (numpy.mean(dist), numpy.std(dist), numpy.percentile(dist, 2.5), numpy.percentile(dist, 97.5)))
        print()
    
        if(plot):
            pyplot.hist(dist, bins=numpy.arange(0, int(max(dist)+1), step=bin_size), label=(name if name else False), color=colors[i], edgecolor='white', alpha=0.6, zorder=(-1*i if reverse_plot else i))
            
    if(plot):
        pyplot.ylabel('num nodes')
        pyplot.legend(loc='upper right')
        pyplot.show()


def network_info(networks, names=None, plot=False, bin_size=1, colors=None, reverse_plot=False):
    import networkx
    networks = [networks] if not isinstance(networks, list) else networks
    names    = [names] if not isinstance(names, list) else names
    colors = [colors] if(colors is not None and not isinstance(colors, list)) else (colors if colors is not None else pyplot.rcParams['axes.prop_cycle'].by_key()['color'])
    
    for i, (network, name) in enumerate(zip(networks, names)):
    
        degree        = [d[1] for d in network.degree()]

        if(name):
            print(name+":")
        print("Degree: mean = %.2f, std = %.2f, 95%% CI = (%.2f, %.2f)\n        coeff var = %.2f" 
              % (numpy.mean(degree), numpy.std(degree), numpy.percentile(degree, 2.5), numpy.percentile(degree, 97.5), 
                 numpy.std(degree)/numpy.mean(degree)))
        r = networkx.degree_assortativity_coefficient(network)
        print("Assortativity:    %.2f" % (r))
        c = networkx.average_clustering(network)
        print("Clustering coeff: %.2f" % (c))
        print()
    
        if(plot):
            pyplot.hist(degree, bins=numpy.arange(0, int(max(degree)+1), step=bin_size), label=(name+" degree" if name else False), color=colors[i], edgecolor='white', alpha=0.6, zorder=(-1*i if reverse_plot else i))
    
    if(plot):
        pyplot.ylabel('num nodes')
        pyplot.legend(loc='upper right')
        pyplot.show()


def results_summary(model):
    print("total percent infected: %0.2f%%" % ((model.total_num_infected()[-1]+model.total_num_recovered()[-1])/model.numNodes * 100) )
    print("total percent fatality: %0.2f%%" % (model.numF[-1]/model.numNodes * 100) )
    print("peak  pct hospitalized: %0.2f%%" % (numpy.max(model.numH)/model.numNodes * 100) )
```
