## Supplementary Data 7 for "Effective high-throughput RT-qPCR screening for SARS-CoV-2 infections in children": run_seir.py

```
#---------Import---------
import sys
import time
import numpy as np
import matplotlib.pyplot as plt
import pandas as pd
sys.path.append('../lib/')
from models import *
from networks import *
from sim_loops import *
from utilities import *
from Epi_models import *
from matplotlib.ticker import FormatStrFormatter
import matplotlib.ticker as mtick

print('Hola mundo \n')


colors = ['tab:green', 'orange', 'orange', 'darkred', 'darkred', 'white', 'indigo', 'black', 'black', 'black', 'silver', 'silver', 'silver', 'silver', 'silver', 'silver', 'indigo']
N=20
NN = N*3
numNodes = NN

# numNodes = 50
# baseGraph    = networkx.barabasi_albert_graph(n=numNodes, m=9)
# G_normal     = custom_exponential_graph(baseGraph, scale=100)
# # Social distancing interactions:
# G_distancing = custom_exponential_graph(baseGraph, scale=10)
# # Quarantine interactions:
# G_quarantine = custom_exponential_graph(baseGraph, scale=5)

G = np.zeros(shape=(NN,NN), dtype = int)
for i in range(NN):
    for j in range(int(i/N)*N, int(i/N)*N+N):
        G[i, j] = 1
G = G-np.identity(NN, dtype = int)
print(G)
G = networkx.convert_matrix.from_numpy_matrix(G)


testing_frequencies = ['weekly2', 'semiweekly', 'semiweekly2', 'triweekly', 'never']
cadence_testing_days    = {
                                    'everyday':     [0, 1, 2, 3, 4, 5, 6, 7, 8, 9, 10, 11, 12, 13, 14, 15, 16, 17, 18, 19, 20, 21, 22, 23, 24, 25, 26, 27],
                                    'workday':      [0, 1, 2, 3, 4, 7, 8, 9, 10, 11, 14, 15, 16, 17, 18, 21, 22, 23, 24, 25],
                                    'triweekly':    [0, 2, 4, 7, 9, 11, 14, 16, 18, 21, 23, 25, 28, 30, 32, 35, 37, 39, 43, 44, 46, 49, 51, 53],
                                    'semiweekly':   [0, 3, 7, 10, 14, 17, 21, 24, 28, 31, 35, 38, 42, 45, 49, 52],
                                    'semiweekly2':  [1, 3, 8, 10, 15, 17, 22, 24, 29, 31, 36, 38, 43, 45, 50, 52],
                                    'weekly':       [0, 7, 14, 21, 28, 35, 42, 49],
                                    'weekly2':      [2, 9, 16, 23, 30, 37, 44, 51],
                                    'biweekly':     [0, 14],
                                    'monthly':      [0],
                                    'cycle_start':  [0],
                                    'never':        []
                                }
frequency = testing_frequencies[0]
p_exts = [1e-2]
R0s = np.array([4.5])
sigma = 1/3
lamda = 1e6
gamma = 1/6

Q_days = np.array([14])

T = 4*7
rates = ['exponential_rates']
#rates = ['time_in_state']

Interactions = [1, 0]

for interaction in Interactions:
	if(interaction):
		p=0
		beta_local=None
	if not(interaction):
		#rate=1
		p=0
		beta_local=0

	for R0 in R0s:
		beta = R0*gamma
		for p_ext in p_exts:
			for i, d in enumerate(Q_days):

				model = ExtSEIRSNetworkModel(G=G, beta=beta, beta_local = beta_local, beta_asym_local=beta_local,  beta_Q = 0, sigma=sigma, lamda = lamda, gamma = gamma,
				 gamma_asym=gamma, a = 1,  p=0, q=0, G_Q=G, initE=0, initI_pre=0, initI_asym=0, isolation_time = d, sigma_Q = sigma, lamda_Q = lamda, 
				 gamma_Q_asym = gamma, gamma_Q_sym = gamma, transition_mode = rates[0], prevalence_ext = p_ext,
				 o = 0.7, store_Xseries=True)
				
				#checkpoints = {'t': [20, 100], 'G': [G_distancing, G_normal], 'p': [0.1, 0.5], 'theta_E': [0.02, 0.02], 'theta_I': [0.02, 0.02], 'phi_E':   [0.2, 0.2], 'phi_I':   [0.2, 0.2]}
				#model.run(T=7*4, checkpoints=None)

				run_tti_sim(model = model, T=T, max_dt=None, testing_cadence = frequency, testing_compliance_random = np.array([True]*NN), 
					isolation_compliance_positive_individual = [True]*NN, isolation_compliance_positive_groupmate = np.array([True]*NN), 
					isolation_groups = np.array([[np.arange(i*N, (i+1)*N)]*N for i in np.arange(NN/N)], dtype = int).reshape(NN, N), 
					isolation_lag_positive = 1, isolation_lag_contact = 1)
				numQ = model.numQ_S + model.numQ_E + model.numQ_pre + model.numQ_asym + model.numQ_R

				#-----------------------------------------------------
				fig1, ax1 = plt.subplots(figsize = (12,8), gridspec_kw={'bottom': 0.15, 'left': 0.1, 'right':.9})
				#ax2.stackplot(model.tseries, [model.numS ,model.numE, model.numI_pre, model.numI_asym, model.numR, model.numQ_S, model.numQ_E, model.numQ_pre, model.numQ_asym, model.numQ_R], labels = ['S', 'E', 'pre', 'I', 'R', 'Q_S', 'Q_E', 'Q_pre', 'Q_I', 'Q_R'], colors = ['darkblue', 'darkred', 'darkgreen', 'indigo', 'darkgoldenrod', 'blue', 'red', 'green', 'blueviolet', 'orange'])
				ax1.stackplot(model.tseries, [model.numS ,model.numE, model.numI_pre, model.numI_asym, model.numR, numQ], labels = ['S', 'E', 'pre', 'I', 'R', 'Q'], colors = ['tab:green', 'orange', 'orange', 'darkred', 'indigo', 'silver'])
				ax1.vlines(cadence_testing_days[frequency], 0, NN, color = 'black', linestyle = '--')
				my_plot_layout(ax = ax1, xlabel = 'Time', ylabel = 'Individuals')
				ax1.set_yticks([])
				#ax1.set_yticks([20*i + 10 for i in np.arange(NN/N)])
				#ax1.set_yticklabels(FormatStrFormatter('%d').format_ticks(np.arange(1, NN/N + 1)))
				lines_symbols = [Line2D([0], [0], color=colors[0], linestyle='', marker = 's', ms = 10, alpha = 1), 
				Line2D([0], [0], color=colors[1], linestyle='', marker = 's', ms = 10, alpha = 1), 
				Line2D([0], [0], color=colors[3], linestyle='', marker = 's', ms = 10, alpha = 1), 
				Line2D([0], [0], color=colors[6], linestyle='', marker = 's', ms = 10, alpha = 1), 
				Line2D([0], [0], color=colors[10], linestyle='', marker = 's', ms = 10, alpha = 1)]
				labels_symbols = ['S', 'E', 'I', 'R', 'Q']
				ax1.legend(lines_symbols, labels_symbols, fontsize = 22, loc = 4)
				ax1.set_title('%d days of Quarantine for non-positive individuals'%(d), fontsize = 24)
				ax1.set_xlim(left = 0, right=T)
				fig1.savefig('../../Figures/1_Extended_Model/examples/population_R0-%.1f_d-%d_prev-%.1e_interactions-%d.png'%(R0, d, p_ext, interaction))
				plt.close(fig1)
				#-----------------------------------------------------
				reg_dt = 1e-3
				T_max = model.tseries[-1]
				reg_time = np.linspace(0, T_max, int((T_max)/reg_dt))
				reg_Xseries = np.ones(shape=(NN, len(reg_time)))

				j=0
				for i in np.arange(len(reg_time)):
					if(reg_time[i]<model.tseries[j]):
						reg_Xseries[:, i] = np.copy(model.Xseries[j, :])
					else:
						j+=1

				fig2, ax2 = plt.subplots(figsize = (12,8), gridspec_kw={'bottom': 0.15, 'left': 0.1, 'right':.9})
				sns.heatmap(reg_Xseries[:60, :], cmap = colors,  ax = ax2, cbar = False)
				ax2.hlines([i*N for i in np.arange(NN/N)], 0, ax2.get_xlim()[1], color = 'black', linewidth = .5)
				ax2.vlines(np.array(cadence_testing_days[frequency], dtype=int)/reg_dt, 0, NN, color = 'black', linestyle = '--')

				my_plot_layout(ax = ax2, xlabel = 'Time', ylabel = 'Groups')
				ax2.set_yticks([])
				#ax2.set_yticks([20*i + 10 for i in np.arange(NN/N)])
				#ax2.set_yticklabels(FormatStrFormatter('%d').format_ticks(np.arange(1, NN/N + 1)))
				ax2.set_xticks(np.arange(0, len(reg_time), int(2/reg_dt)))
				ax2.set_xticklabels(FormatStrFormatter('%.f').format_ticks(reg_time[::int(2/reg_dt)]))
				ax2.set_xlim(left = 0, right=T/reg_dt)
				lines_symbols = [Line2D([0], [0], color=colors[0], linestyle='', marker = 's', ms = 10, alpha = 1), 
				Line2D([0], [0], color=colors[1], linestyle='', marker = 's', ms = 10, alpha = 1), 
				Line2D([0], [0], color=colors[3], linestyle='', marker = 's', ms = 10, alpha = 1), 
				Line2D([0], [0], color=colors[6], linestyle='', marker = 's', ms = 10, alpha = 1), 
				Line2D([0], [0], color=colors[10], linestyle='', marker = 's', ms = 10, alpha = 1)]
				labels_symbols = ['S', 'E', 'I', 'R', 'Q']
				ax2.legend(lines_symbols, labels_symbols, fontsize = 22, bbox_to_anchor=(1.2, .8))
				ax2.set_title('%d days of Quarantine for non-positive individuals'%(d), fontsize = 24)
				fig2.savefig('../../Figures/1_Extended_Model/examples/groups_R0-%.1f_d-%d_prev-%.1e_interactions-%d.png'%(R0, d, p_ext, interaction))
				plt.close(fig2)
```
