## Supplementary Data 7 for "Effective high-throughput RT-qPCR screening for SARS-CoV-2 infections in children": run_seir_ensemble.py

```
#---------Import---------
import sys
import time
from tqdm import tqdm
import numpy as np
import matplotlib.pyplot as plt
import pandas as pd
sys.path.append('../lib/')
from models import *
from networks import *
from sim_loops import *
from utilities import *
from functions import *

Text_files_path = '../../Text_files/SEIR_ensemble/'

print('Hola mundo \n')

N=20
N_groups = 12
NN = N*N_groups

#----- Interaction in groups
G = np.zeros(shape=(NN,NN), dtype = int)
for i in range(NN):
    for j in range(int(i/N)*N, int(i/N)*N+N):
        G[i, j] = 1
G = G-np.identity(NN, dtype = int)
G = networkx.convert_matrix.from_numpy_matrix(G)

#----- Random interactions
#numNodes = NN
#baseGraph    = networkx.barabasi_albert_graph(n=numNodes, m=2)
#G_normal     = custom_exponential_graph(baseGraph, scale=100)
# Social distancing interactions:
#G_distancing = custom_exponential_graph(baseGraph, scale=10)
# Quarantine interactions:
#G_quarantine = custom_exponential_graph(baseGraph, scale=5)

# ----- MODEL PARAMATERS -----
N_ensemble = 10000
testing_frequencies = ['weekly2', 'semiweekly2', 'triweekly', 'never']
#testing_frequencies = ['weekly2']
p_exts = [1e-4, 1e-3]
R0s = np.array([7.5, 4.5, 2.5])
sigma = 1/3
lamda = 1e6
gamma = 1/6
T = 8*7
Q_days = np.array([0, 14])

#rates = ['time_in_state']
rates = ['exponential_rates']

Interactions = [1, 0]


for rate in np.arange(len(rates)):
	for interaction in Interactions:
		if(interaction):
			p=0
			beta_local=None
		if not(interaction):
			#rate=1
			p=0
			beta_local=0
		for testing_frequency in testing_frequencies:
			#if(testing_frequency=='never'):
				#Q_days = np.array([14])
			for R0 in R0s:
				beta = R0*gamma
				#beta = gamma-(((1-R0**2)*(sigma+gamma)**2)/(4*sigma))
				for p_ext in p_exts:
					for i, d in enumerate(Q_days):
						I_cum = np.array([])
						S_cum = np.array([])
						print('RUNNING ENSEMBLE \n Interactions= %d;  d = %d days ; R0 = %.1f ; Prevalence = %.e \n'%(interaction, d, R0, p_ext) + 'Testing frequency: '+testing_frequency+'\n')
						for n_ensemble in tqdm(np.arange(N_ensemble)):

							model = ExtSEIRSNetworkModel(G=G, beta=beta, beta_local = beta_local, beta_asym_local=beta_local,  beta_Q = 0, sigma=sigma, lamda = lamda, 
								gamma = gamma, gamma_asym=gamma, a = 1,  p=p, q=0, G_Q=G, initI_asym=0, isolation_time = d, sigma_Q = sigma, 
								lamda_Q = lamda, gamma_Q_asym = gamma, gamma_Q_sym = gamma, transition_mode = rates[rate], prevalence_ext = p_ext, 
								o = 0.7)

							run_tti_sim(model = model, T=T, max_dt=None, testing_cadence = testing_frequency, testing_compliance_random = np.array([True]*NN), isolation_compliance_positive_individual = [True]*NN,
								isolation_compliance_positive_groupmate = np.array([True]*NN), isolation_groups = np.array([[np.arange(i*N, (i+1)*N)]*N for i in np.arange(NN/N)], dtype = int).reshape(NN, N), 
								isolation_lag_positive = 1, isolation_lag_contact = 1, print_report = False)

							I_cum = np.append(I_cum, (model.numE[-1] + model.numI_pre[-1] + model.numI_sym[-1] + model.numI_asym[-1] + model.numR[-1] + model.numQ_E[-1] + model.numQ_pre[-1] + model.numQ_sym[-1] + model.numQ_asym[-1] + model.numQ_R[-1]))
							S_cum = np.append(S_cum, (model.numS[-1] + model.numQ_S[-1]))

						outfile = open(Text_files_path+'I_cummulative_pop_R0-%.1f_p_ext-%.2e_d-%.d_'%(R0, p_ext, d)+'testing_frequency-'+testing_frequency+'_interactions-%d_%d.pkl'%(interaction, rate),'wb')
						pickle.dump(I_cum, outfile)
						outfile.close()

						outfile2 = open(Text_files_path+'S_cummulative_pop_R0-%.1f_p_ext-%.2e_d-%.d_'%(R0, p_ext, d)+'testing_frequency-'+testing_frequency+'_interactions-%d_%d.pkl'%(interaction, rate),'wb')
						pickle.dump(S_cum, outfile2)
						outfile2.close()

	
```
