## Supplementary Data 7 for "Effective high-throughput RT-qPCR screening for SARS-CoV-2 infections in children": 0_Sensitivity.ipynb

```
# %% [code]
import sys
sys.path.append('../Codes/lib/')
from functions import *
import numpy as np
import matplotlib.pyplot as plt
import pandas as pd
from io import StringIO
from matplotlib.lines import Line2D
from datetime import datetime, timedelta
import scipy.special as sc
import os.path
import pickle
import networkx
from matplotlib import style
from scipy.interpolate import interp1d
from scipy.optimize import curve_fit
from IPython.display import display, Math
import matplotlib.ticker as mtick
from matplotlib.ticker import PercentFormatter


Text_files_path = '../Text_files/'
print(style.available)
#style.use('seaborn-paper')
%autosave 60

# %% [code]
g = 4.5
time = np.linspace(0, 5, 100)
virus = np.exp(g * time)
sensitivity = (1/(1+(np.log10(virus[1:])/(1.66))**(-7.49)))
t_inc = 3
tau = 0
fig, ax  = plt.subplots(figsize = (10,10), gridspec_kw={'bottom':0.1, 'left':0.2})
ax.plot(virus[1:], sensitivity, color = 'darkred')
#ax.fill_between(time[1:], 1e4*(1/(1+(time[1:]/5)**(-6))), np.ones_like(time[1:])*1, alpha = .8)
#ax.vlines([t_inc, t_inc-tau], 1, [np.exp(g * t_inc), np.exp(g * (t_inc-tau))], linestyle = '--', color = ['grey', 'darkgrey'])
#ax.hlines([np.exp(g * t_inc), np.exp(g * (t_inc-tau))], 0, [t_inc, t_inc-tau], linestyle = '--', color = ['grey', 'darkgrey'])
ax.set_ylim(0, 1.01)
my_plot_layout(ax, xscale = 'log', yscale = 'linear', xlabel = 'VL', ylabel = 'sensitivity',
              title = 'Exponential growth')
ax.yaxis.set_major_formatter(PercentFormatter(1))
fig.savefig('../Figures/0_Sensitivity/sensitivity_fit.pdf')


# %% [code]
data = pd.read_excel(Text_files_path+'data/Sensitivität.xlsx', header = 1)
data

# %% [code]
fig, ax  = plt.subplots(figsize = (12,10), gridspec_kw={'bottom':0.1, 'left':0.2, 'right':.8})

ax.plot(data['from'][1:], data['Sensitivity'][1:]/100, marker = 'o', ms = 10, linestyle = '', color = 'darkblue', label = 'Lollipop morning', linewidth = 3)
ax.plot(data['from'][1:], data['Sensitivity.1'][1:]/100, marker = '.', ms = 14, linestyle = '', color = 'lightblue', alpha = .6, label = 'Lollipop random')

x1 = np.log(np.log10(data['from'][4:-1]))
y1 = np.log((1/(abs(data['Sensitivity'][4:-1])/100.0))-1)

x2 = np.log(np.log10(data['from'][1:-2]))
y2 = np.log((1/(abs(data['Sensitivity.1'][1:-2])/100.0))-1)

popt1, pcov1 = curve_fit(my_linear_func, xdata=x1, ydata=y1, p0 = (2, -6))
popt2, pcov2 = curve_fit(my_linear_func, xdata=x2, ydata=y2, p0 = (1, -4))

print((np.exp(-popt1[0]/popt1[1])), (np.exp(-popt2[0]/popt2[1])))
print(popt1[1], popt2[1])

S1 = 1/(np.exp(my_linear_func(np.log(np.log10(virus[1:])), *popt1))+1)
S2 = 1/(np.exp(my_linear_func(np.log(np.log10(virus[1:])), *popt2))+1)

S10 = 1/(np.exp(my_linear_func(x1, *popt1))+1)
S20 = 1/(np.exp(my_linear_func(x2, *popt2))+1)

ax.plot(virus[1:], S1, color = 'darkblue', linewidth = 3)
ax.plot(virus[1:], S2, color = 'lightblue', linewidth = 3, alpha = .6)

X1 = np.log(np.log10(virus[1:]))
Y1 = np.log(1/S1-1)
S1_delta = (np.exp(Y1)/(np.exp(Y1)+1)**2)*np.sqrt(pcov1[0,0] + X1**2*pcov1[1,1])

X2 = np.log(np.log10(virus[1:]))
Y2 = np.log(1/S2-1)
S2_delta = (np.exp(Y2)/(np.exp(Y2)+1)**2)*np.sqrt(pcov2[0,0] + X2**2*pcov2[1,1])

ax.fill_between(virus[1:], S1-1.96*S1_delta, S1+1.96*S1_delta, alpha = .15, color = 'darkblue')
ax.fill_between(virus[1:], S2-1.96*S2_delta, S2+1.96*S2_delta, alpha = .15, color = 'lightblue')

ax.vlines([10**(np.exp(-popt1[0]/popt1[1])), 10**(np.exp(-popt2[0]/popt2[1]))], 0, 1, colors = ['darkblue', 'lightblue', 'darkorange'], linestyle = '--', alpha = .6)
ax.hlines([.5], 1, 1e9, alpha = .2, color = 'silver')

my_plot_layout(ax, xscale = 'log', xlabel = 'Viral load [copies/mL]', ylabel = 'Detection rate')
ax.yaxis.set_major_formatter(PercentFormatter(1))
#ax.set_ylim(0, 1)
ax.legend(fontsize = 24, loc=4)
fig.savefig('../Figures/0_Sensitivity/fit.pdf')

pcov1

# %% [code]
fit_data = pd.DataFrame(data=np.transpose(np.vstack((virus[1:], S1, S1-1.96*S1_delta, S1+1.96*S1_delta, S2, S2-1.96*S2_delta, S2+1.96*S2_delta  ))), columns = ['Viral load','Morning','Morning lower','Morning upper','Random','Random lower','Random upper'])
fit_data.to_excel(Text_files_path+ 'data/Sensitivity_fit.xlsx')

# %% [code]
fig, ax  = plt.subplots(figsize = (15,10), gridspec_kw={'bottom':0.1, 'left':0.2, 'right':.8})

ax.plot(time[1:], S1 , color = 'darkblue', label = 'lollipop', linewidth = 3)
#ax.plot(time[1:], sensitivity2 , color = 'lightblue')
#ax.plot(time[1:], sensitivity3 , color = 'darkorange', label = 'antigen', linewidth = 3)
#ax.plot(time, np.heaviside(time - (t_inc-tau), 0), color = 'silver', linewidth = 3, label  = 'control')

#ax.vlines([np.log(10**(np.exp(-popt1[0]/popt1[1])))/g, np.log(10**(np.exp(-popt5[0]/popt5[1])))/g], [0, 0], [.5, .5], colors = ['darkblue', 'darkorange'], linestyle = '--', alpha = .6)
#ax.hlines([.5, .5], [0, 0], [np.log(10**(np.exp(-popt5[0]/popt5[1])))/g, np.log(10**(np.exp(-popt1[0]/popt1[1])))/g], color = ['darkorange', 'darkblue'], linestyle = '--', alpha = .6)
#ax.fill_between(time, np.heaviside(time - (t_inc-tau), 0), 0, alpha = .1, color = 'silver')
ax.fill_between(time[1:], S1, 0, alpha = .05, color = 'darkblue')
#ax.fill_between(time[1:], sensitivity3, 0, alpha = .1, color = 'darkorange')

ax.set_ylim(0, 1.02)
ax.set_xlim(0, 5)
my_plot_layout(ax, ylabel = 'Detection rate', xlabel = 'Time (days)')
ax.yaxis.set_major_formatter(PercentFormatter(1))
ax.yaxis.label.set_color('darkblue')
ax.tick_params('y', colors='darkblue')
#ax.legend(fontsize = 22, loc = 2)

ax2 = ax.twinx() 
ax2.plot(time, virus, color = 'darkred', linewidth = 4)
ax2.vlines([t_inc], 1, [np.exp(g * t_inc),], linestyle = '--', color = ['k', 'k'], linewidth = 3, label = r'$\tau_{p}$',alpha = .6)
ax2.hlines([np.exp(g * t_inc)], [t_inc], 7, linestyle = '--', color = ['k', 'k'], linewidth = 3, alpha = .3)
ax2.set_ylim(1, 1e9)
ax2.legend(fontsize = 28, loc = 4)
my_plot_layout(ax2, yscale = 'log',  xlabel = 'Time', ylabel = 'Viral load (copies/mL)')
ax2.yaxis.label.set_color('darkred')
ax2.tick_params('y', colors='darkred')

fig.savefig('../Figures/0_Sensitivity/detection_rate_1.pdf')


# %% [code]


# %% [code]


# %% [code]
```
