## Supplementary Data 7 for "Effective high-throughput RT-qPCR screening for SARS-CoV-2 infections in children": README.md

# lolli_testing
This repository contains the code used to simulate an epidemiological scenario where the Lolli-testing method is used. 

The Python code is a modified version of the publicly available SEIRS+ code at https://github.com/ryansmcgee/seirsplus. We thank the Ryan Seamus McGee (ryansmcgee) for making this code available.

It contains 4 main folders: /Text_files, /Notebooks, /Figures and /Codes

/Text_files/SEIR_ensemble contains output of simulations in the form of text files. 
/Text_files/Data contains data of sensibility of Lolli test and output of analysis of sensitivity and simulations.

Analysis of sensitivity data is done in the jupyter notebook at /Notebooks/0_Sensitivity.ipynb

The main analysis of simulation outputs if done with /Codes/Python/analysis_ensemble1.py
/Codes/lib contains the important classes and fucntions to run the model
/Codes/Python/run_seir.py performs the simulation of one population for given time T and outputs two figures in /Figures/1_Extended_Model/examples
/Codes/Python/run_seir_ensemble.py performs the simulation of an ensemble of populations for a given time T and outputs total number of infected and susceptible individuals at the end of the time in *.pkl files at /Textfiles/SEIR_ensemble.

All the codes run stably under Python 3.8.5 and require the following libraries: numpy, matplotlib, networkx, tqdm, pandas, sys, time, random, bisect, math, os, scipy, FARZ and pickle.
