## Supplementary figures and images for "Effective high-throughput RT-qPCR screening for SARS-CoV-2 infections in children"

### detection_rate_1.pdf

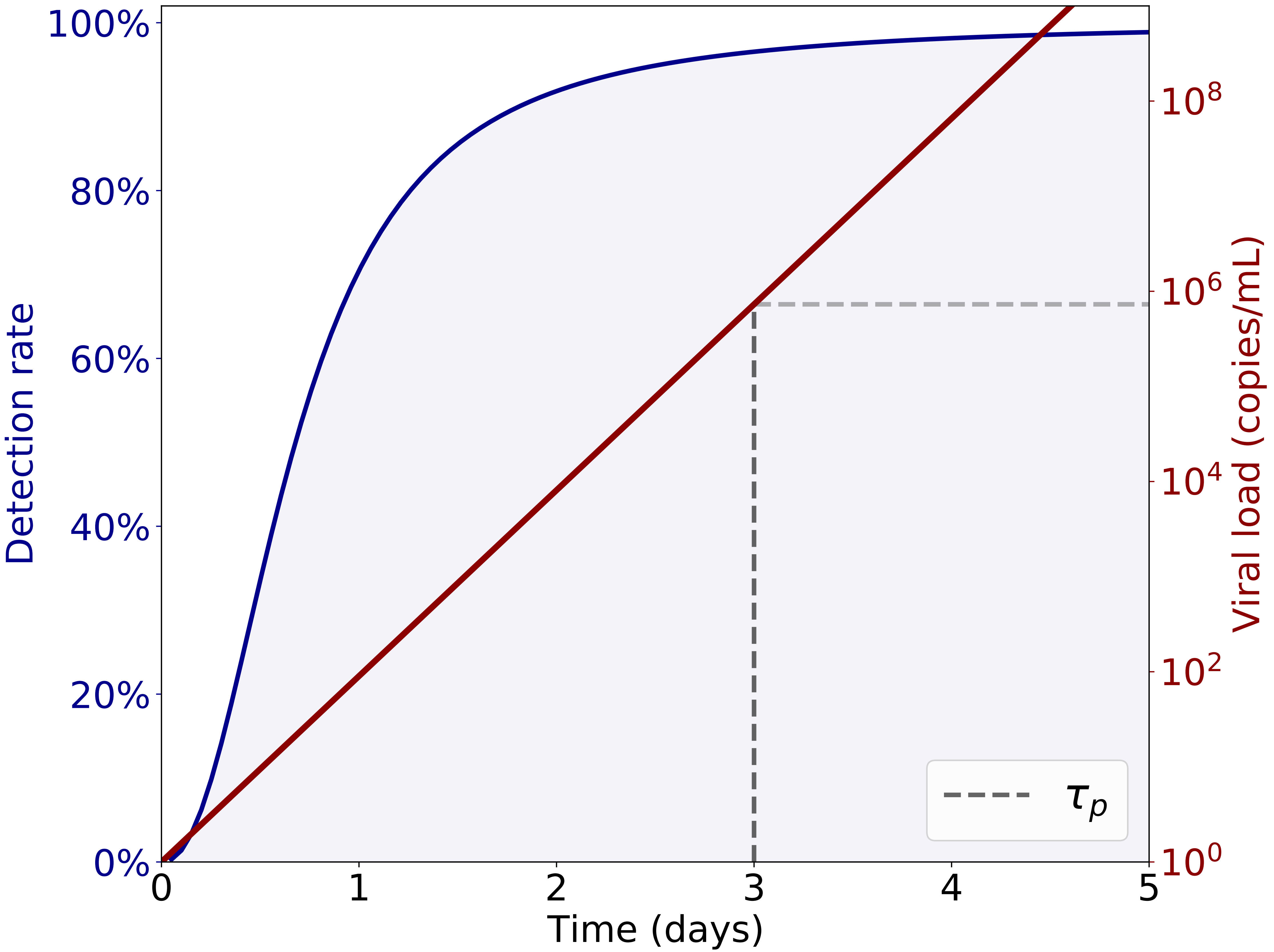

### example.png

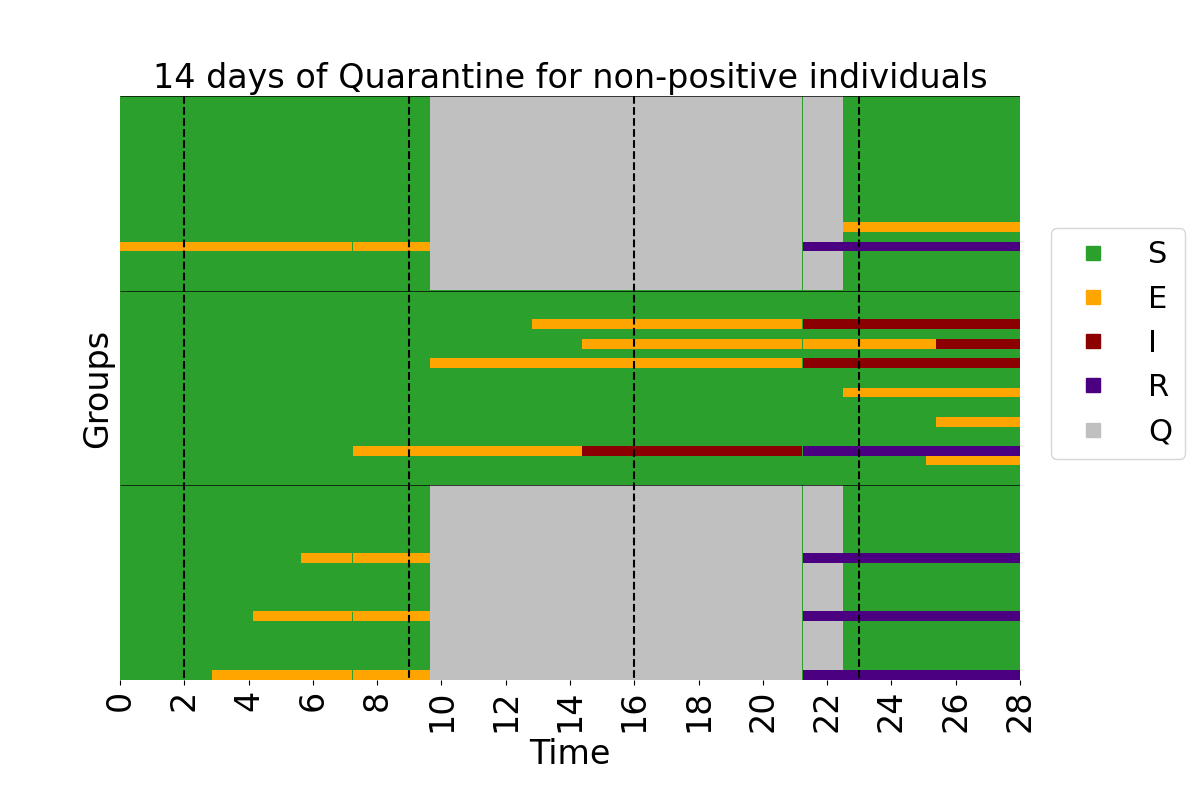

### fit.pdf

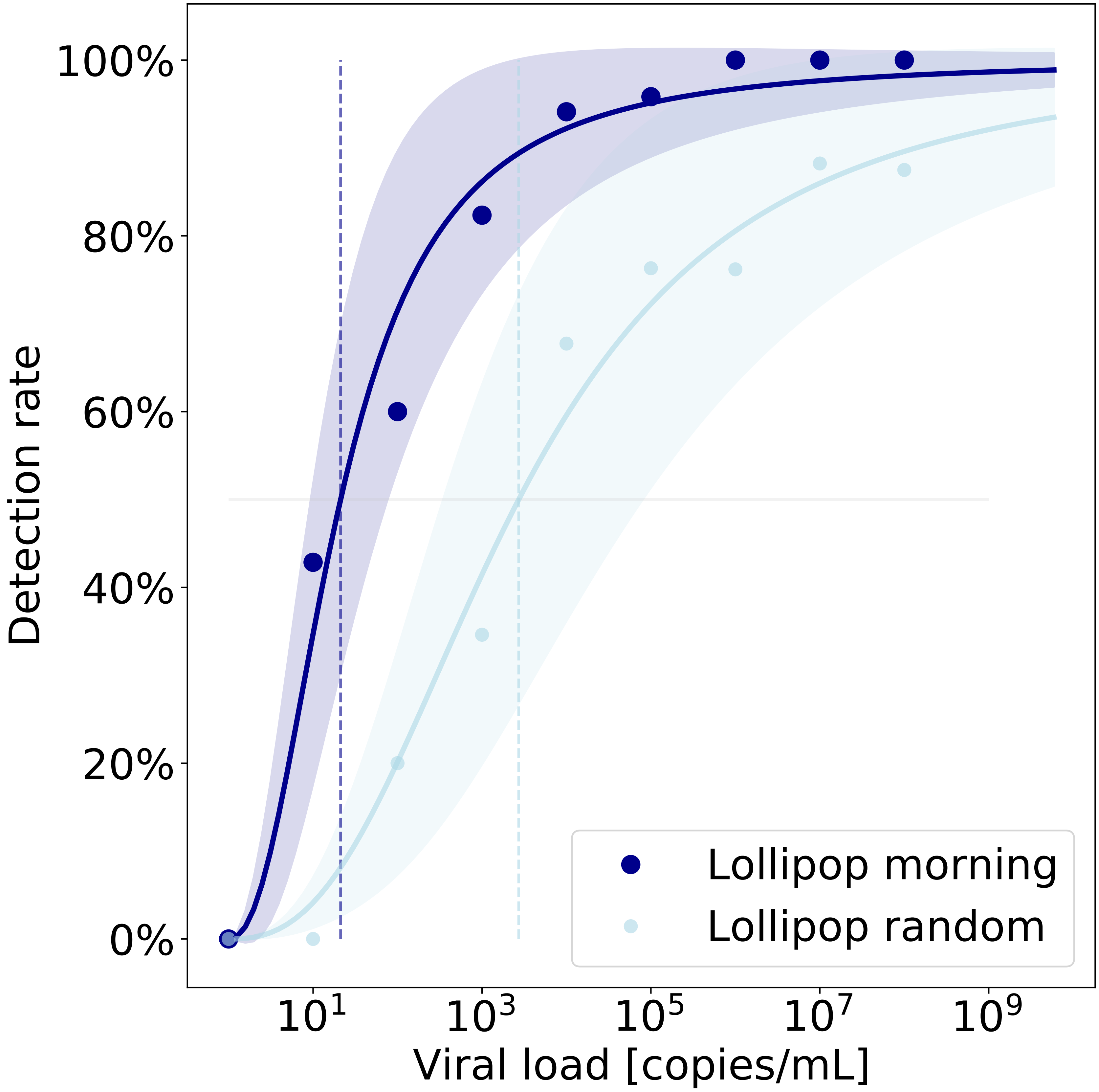

### groups_R0-4.5_d-0_prev-1.0e-02.png

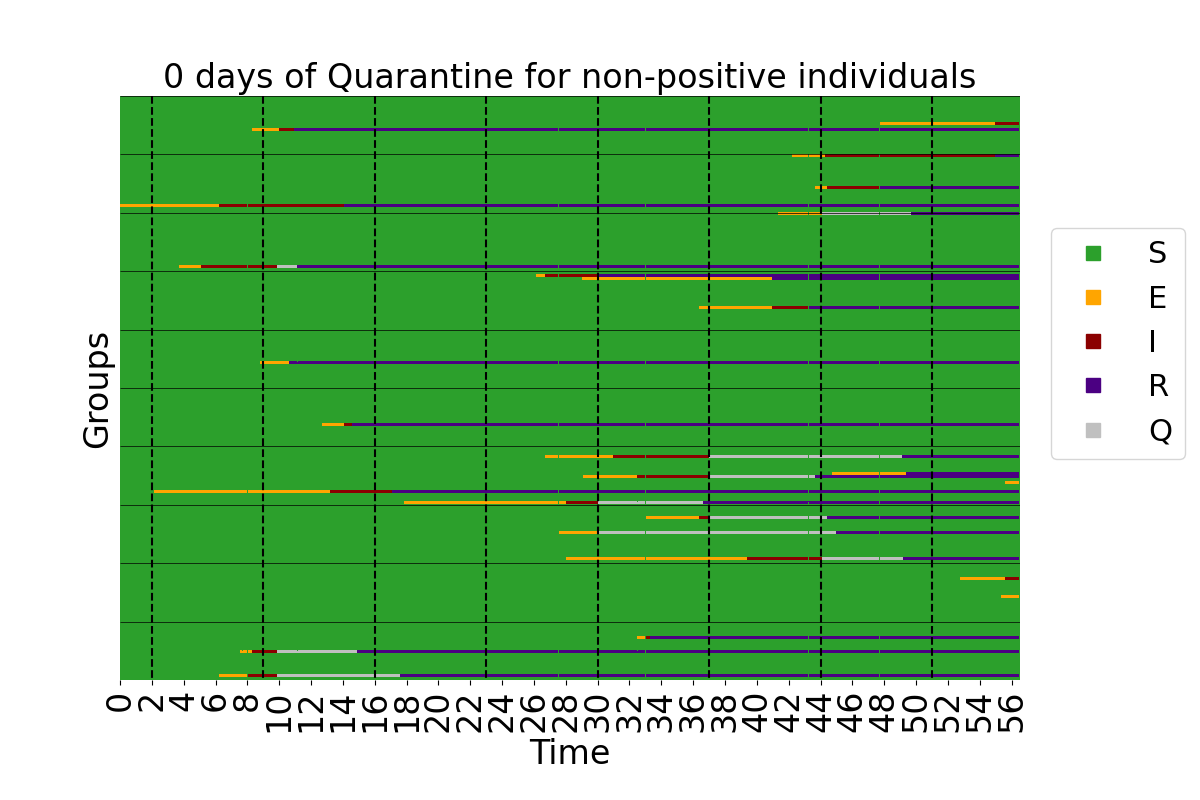

### groups_R0-4.5_d-0_prev-1.0e-03.png

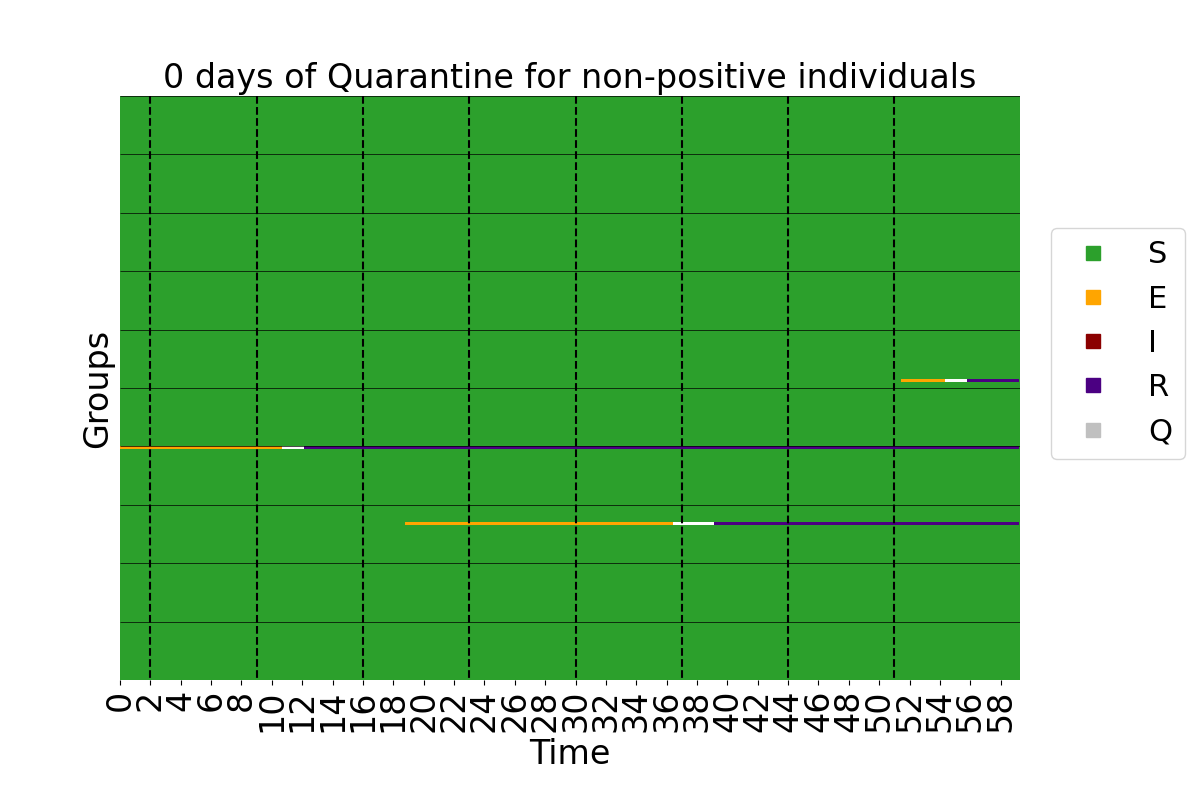

### groups_R0-4.5_d-14_prev-1.0e-02_interactions-0.png

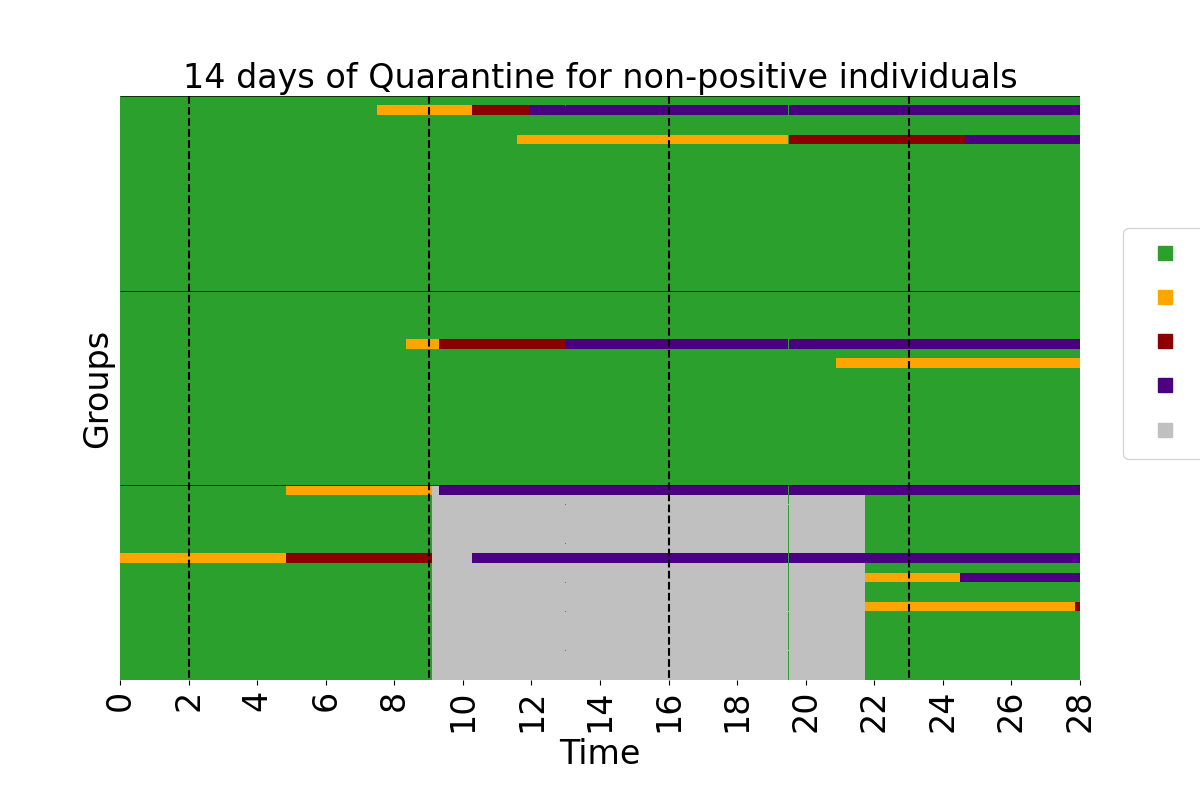

### groups_R0-4.5_d-14_prev-1.0e-02_interactions-1.png

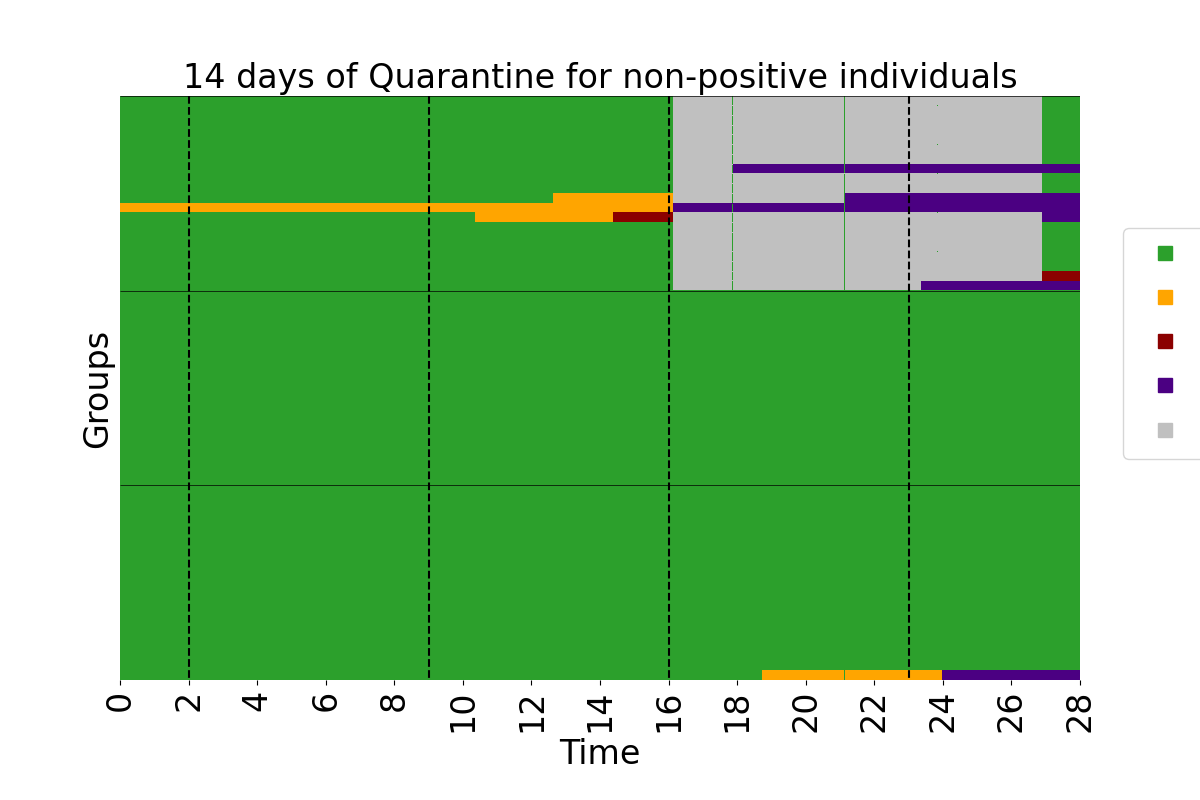

### groups_R0-4.5_d-14_prev-1.0e-03.png

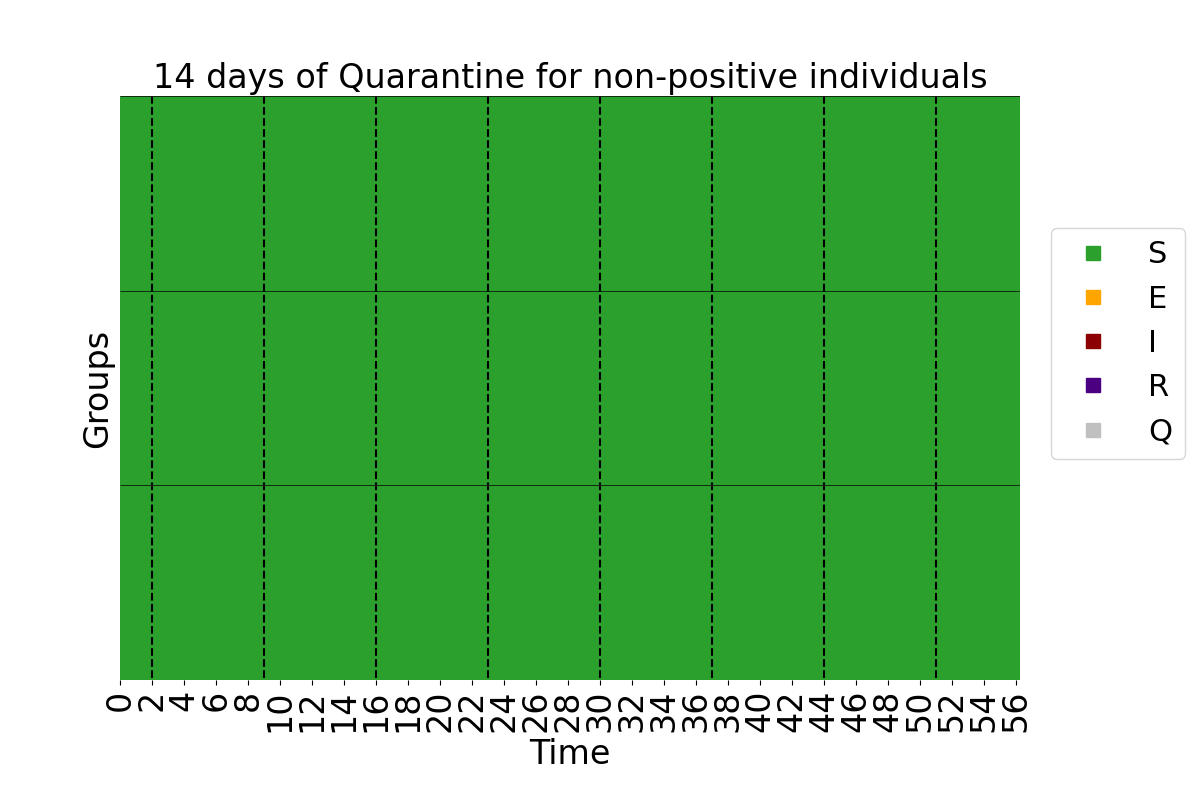

### population_R0-4.5_d-0_prev-1.0e-02.png

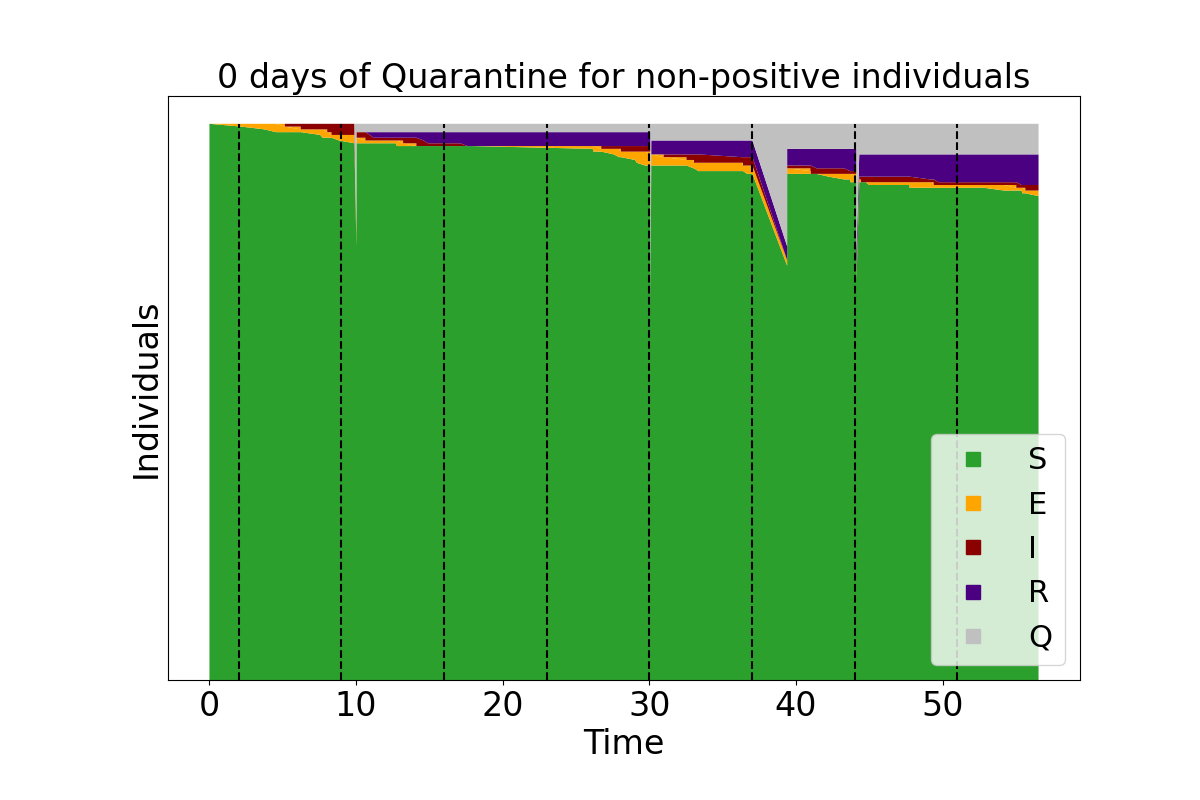

### population_R0-4.5_d-0_prev-1.0e-03.png

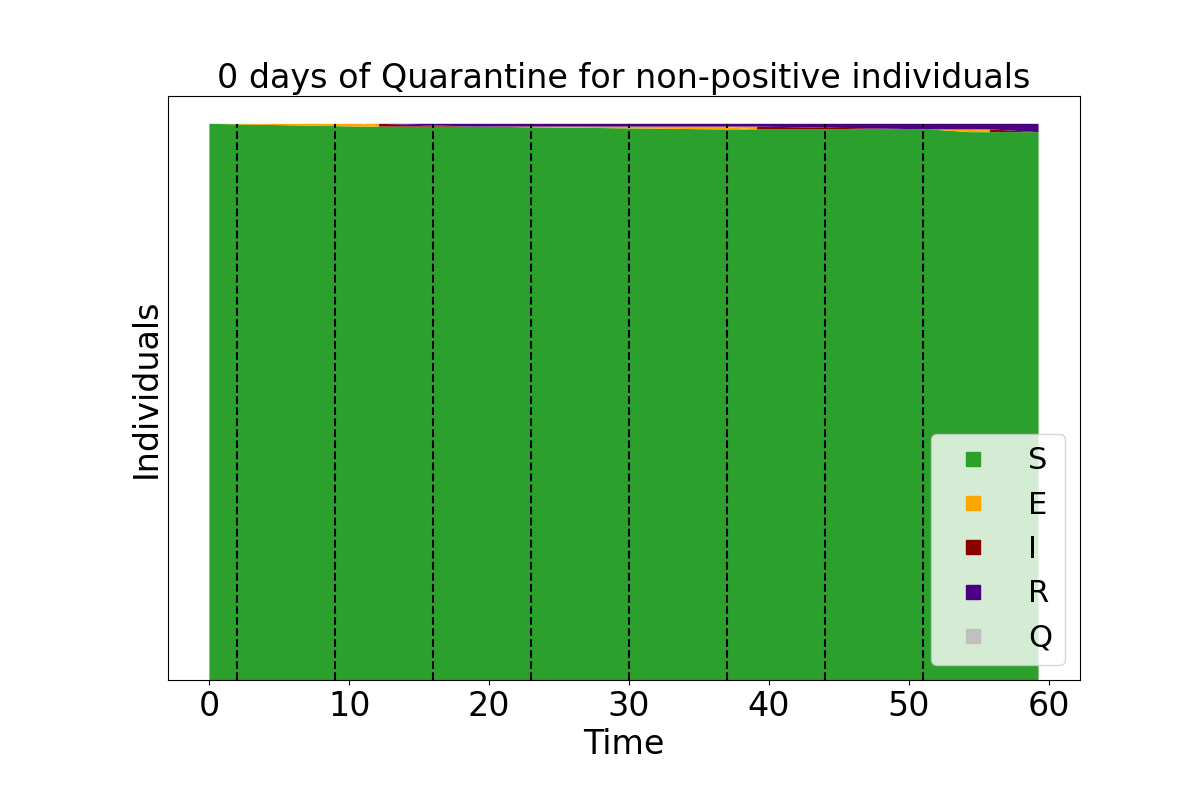

### population_R0-4.5_d-14_prev-1.0e-02.png

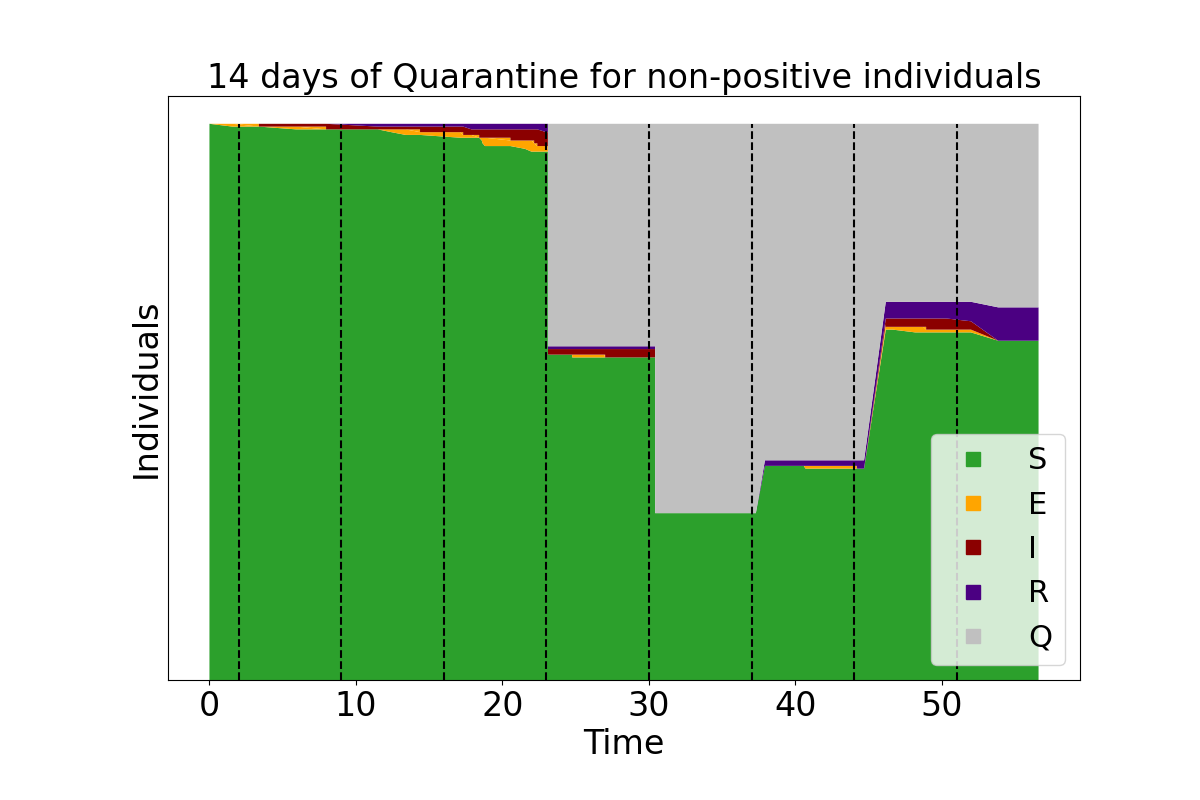

### population_R0-4.5_d-14_prev-1.0e-02_interactions-0.png

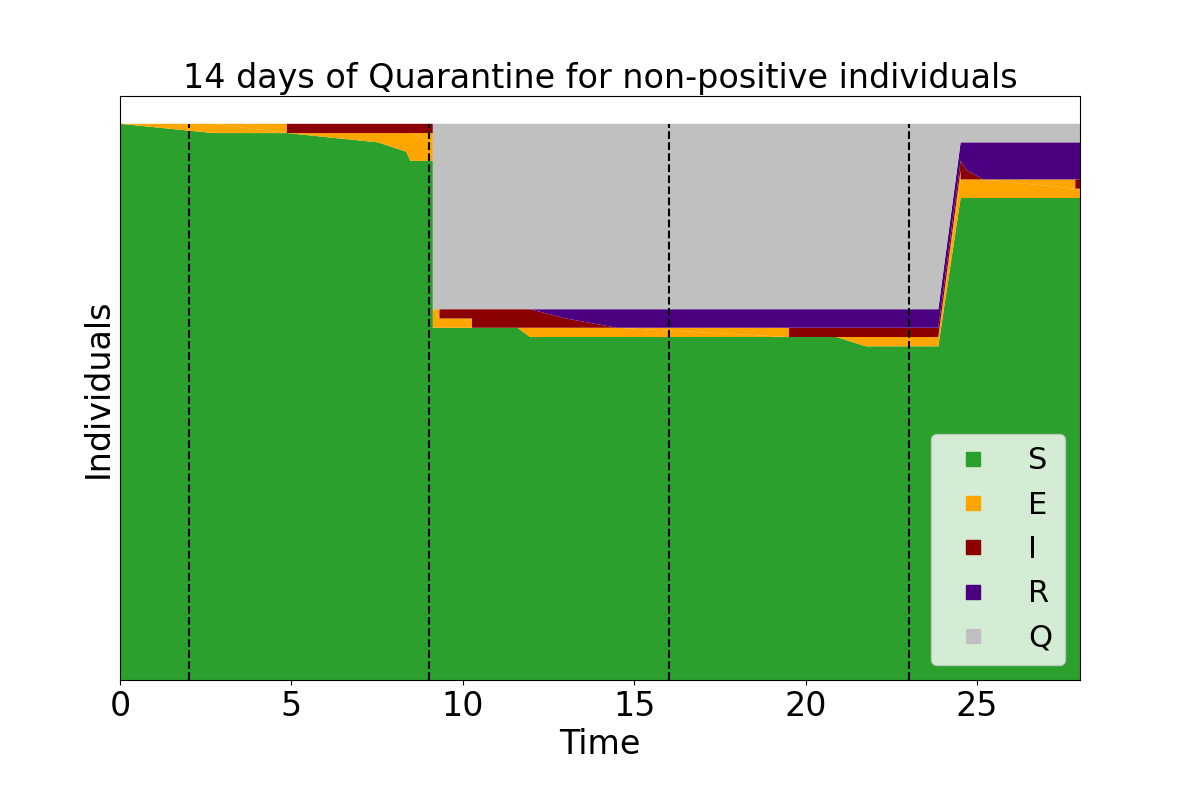

### population_R0-4.5_d-14_prev-1.0e-02_interactions-1.png

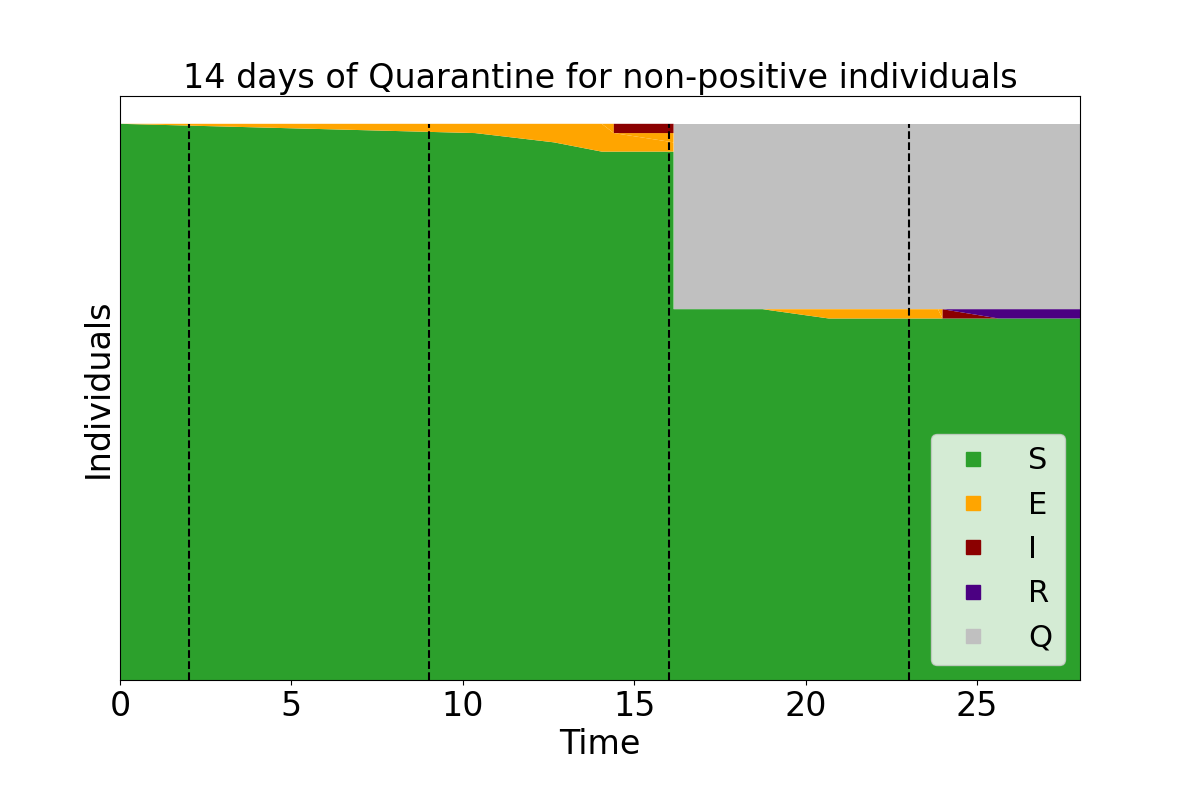

### population_R0-4.5_d-14_prev-1.0e-03.png

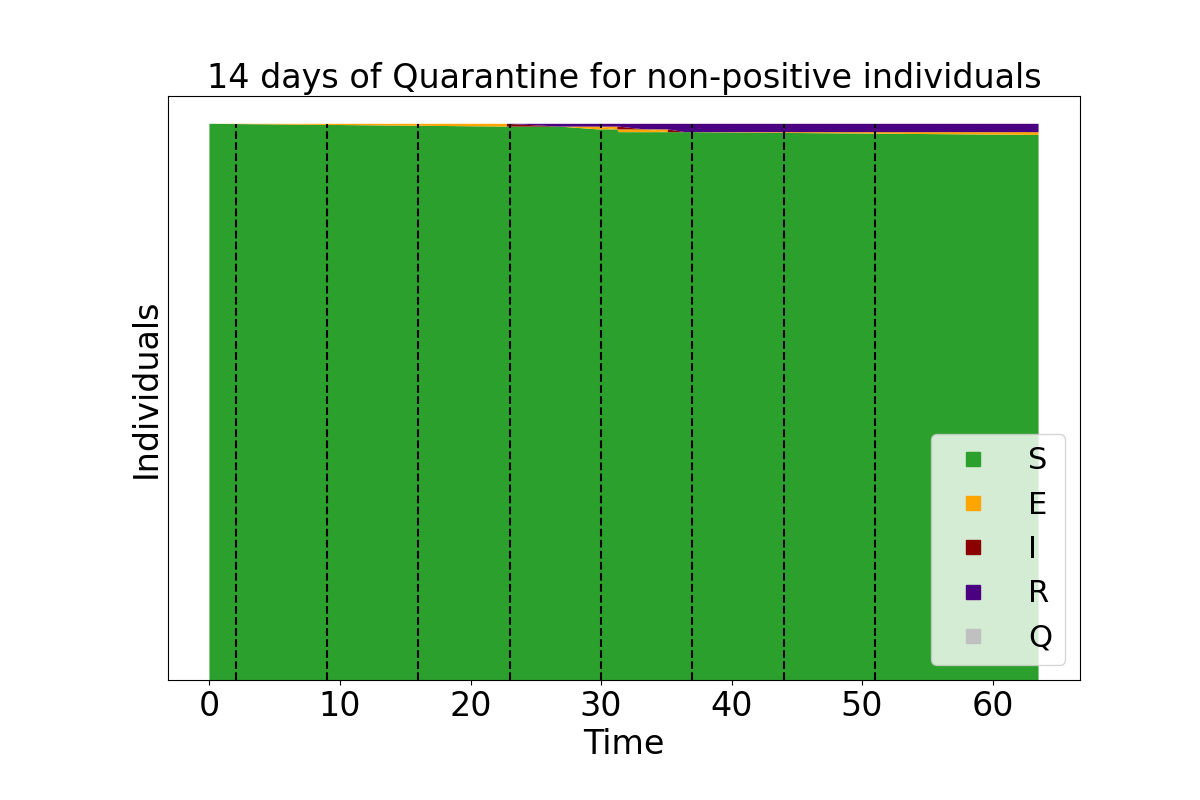

### population_R0-4.5_d-14_prev-1.0e-03_interactions-0.png

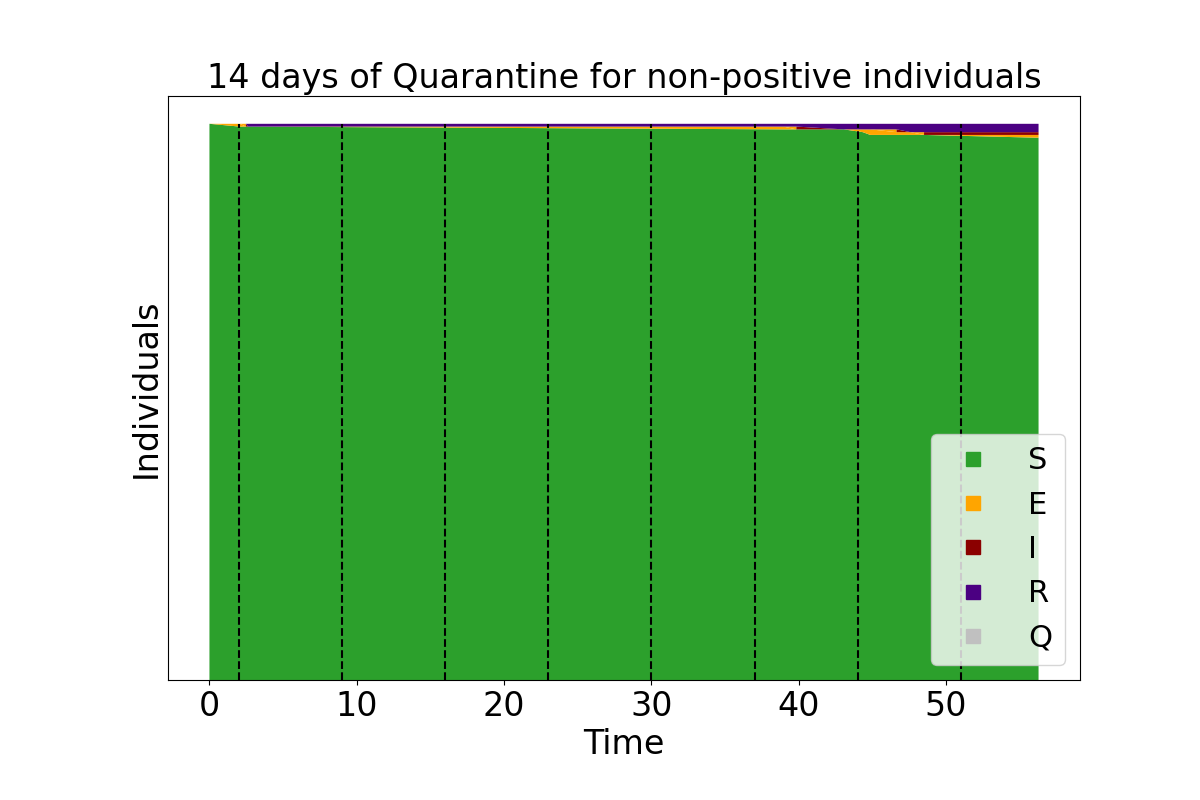

### population_R0-4.5_d-14_prev-1.0e-03_interactions-1.png

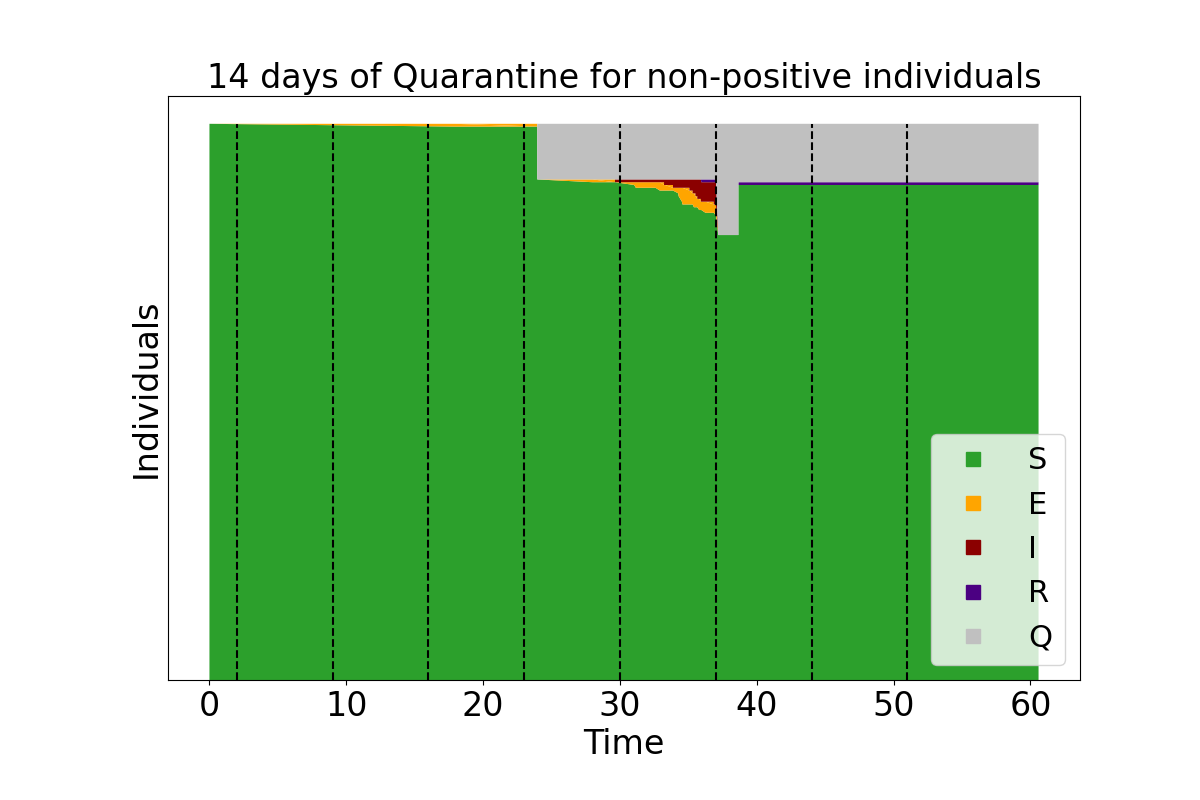

### PrevTrans_d-0.pdf

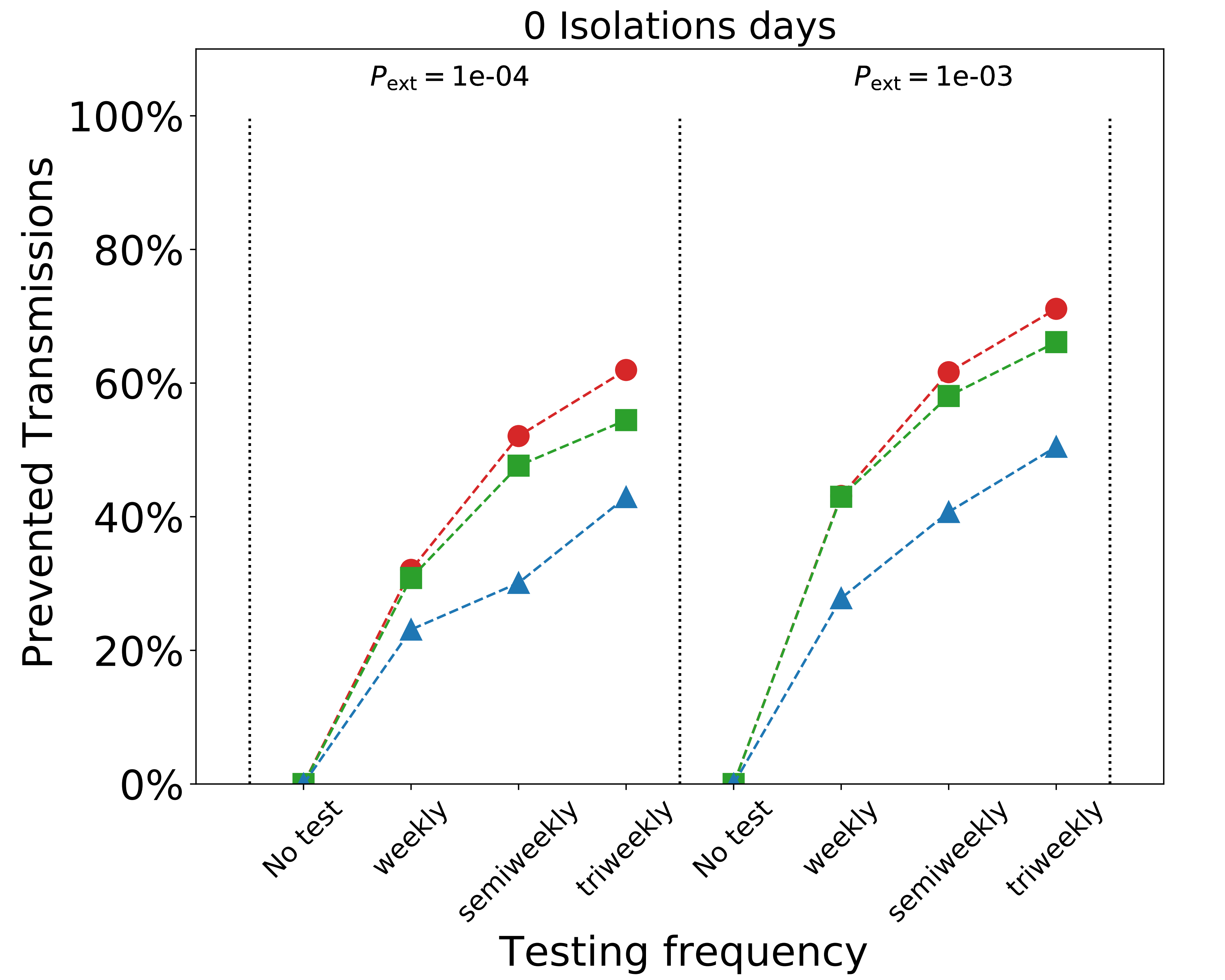

### PrevTrans_d-14.pdf

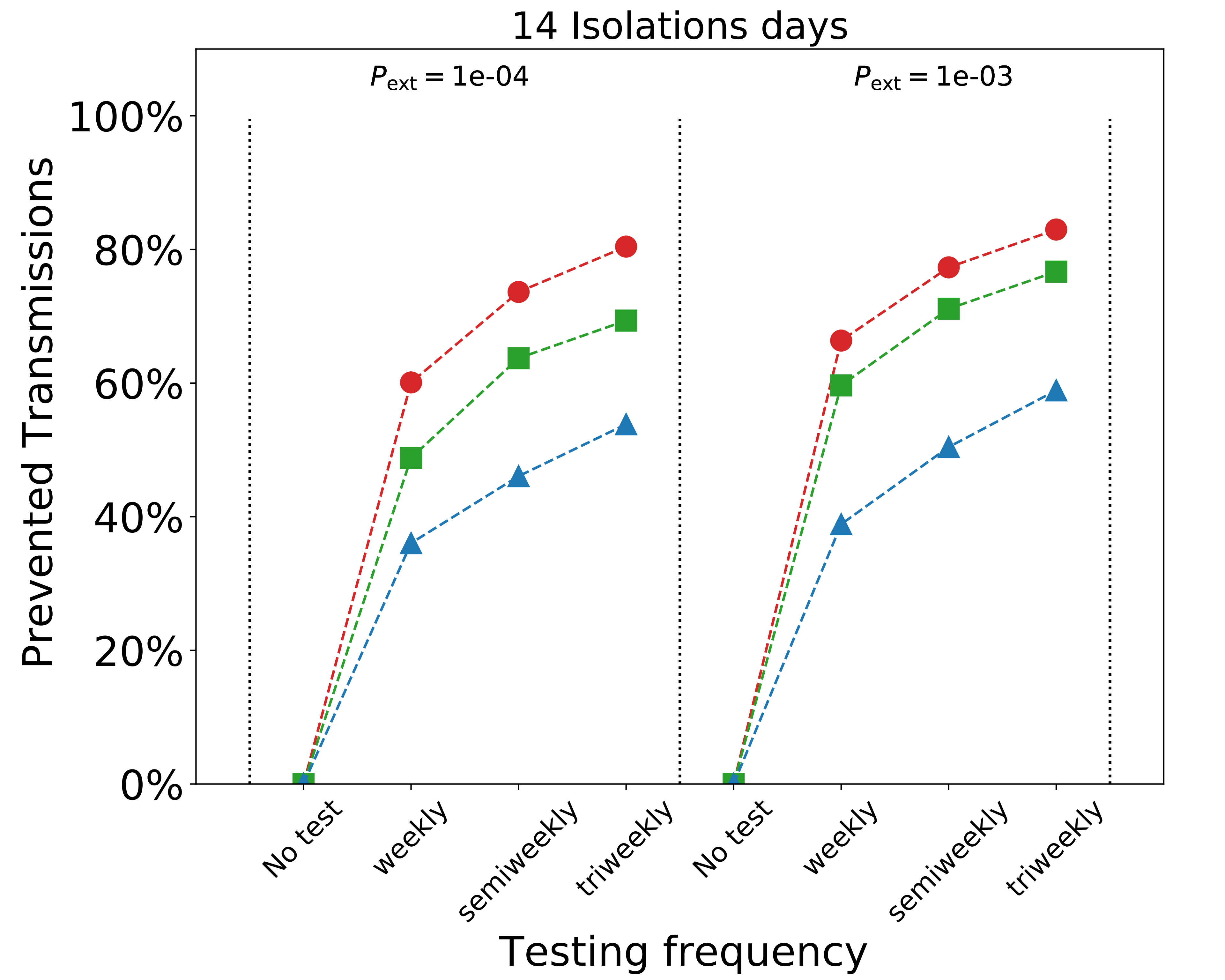

### sensitivity_fit.pdf

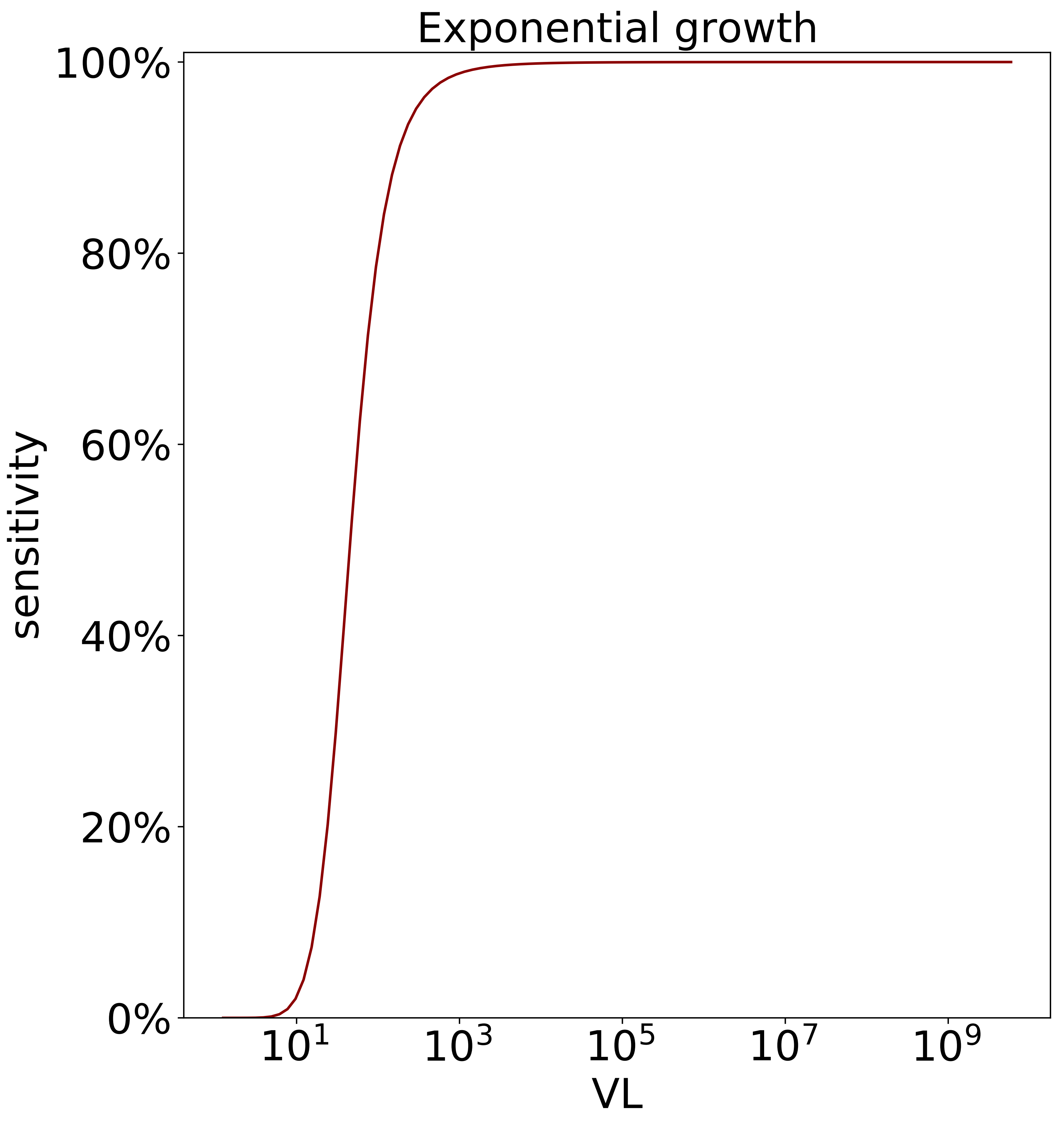

### tau_distributions.pdf

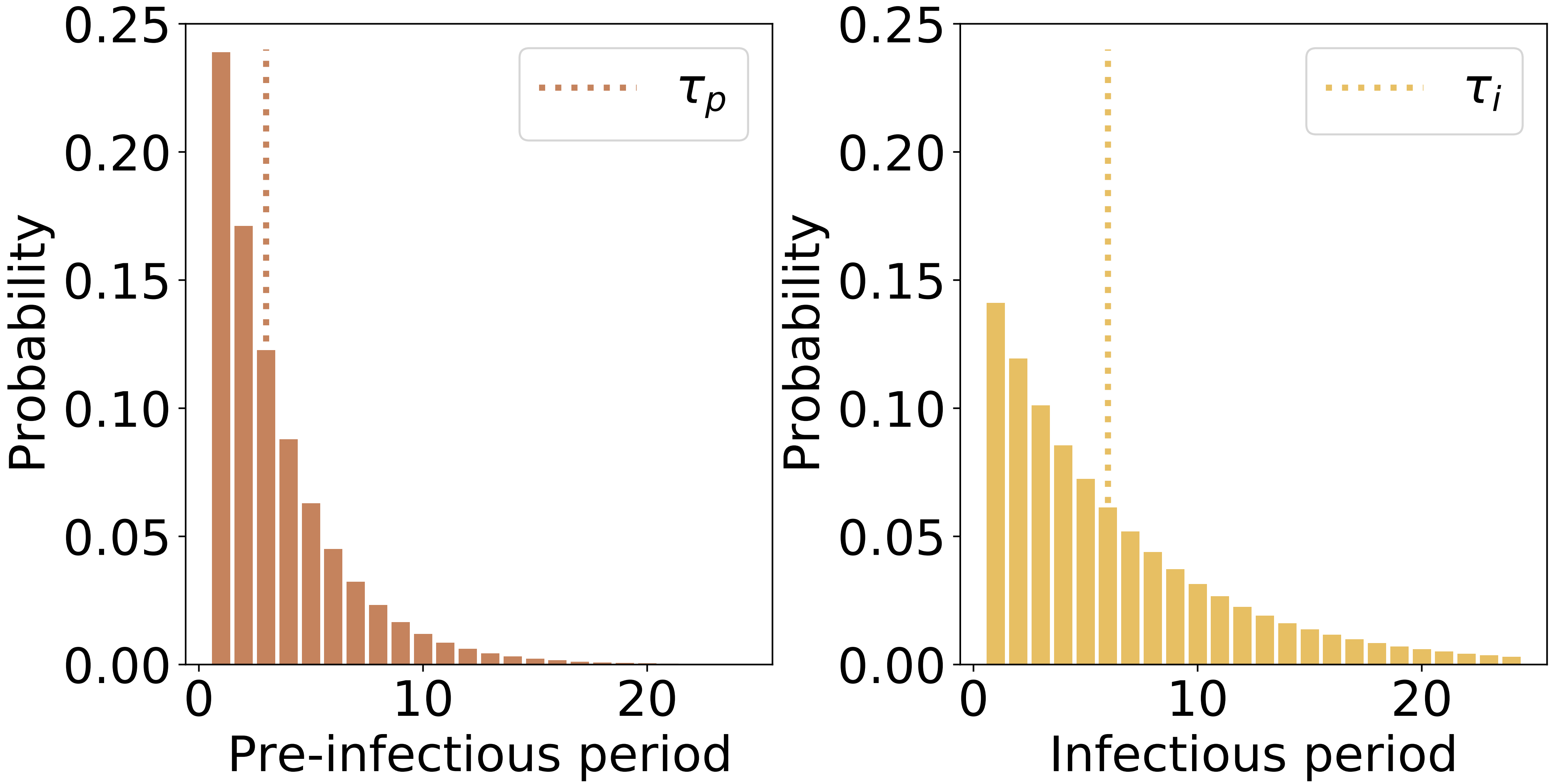
