## Supplementary Data 8 for "Effective high-throughput RT-qPCR screening for SARS-CoV-2 infections in children": README.txt

# Lolli-Test-NRW

## SARS-CoV-2 infection dynamics in schools

### Input Data

- "df_list_clean$tbl_pools":
"poolsId" - unique identifier 
"clientId" - unique identifier for every school 
"datum" - date on which the PCR was performed 
"ergebnis" - PCR result (1 for positive and 2 for negative)

- "df_list_clean$tbl_singles":
"singleId" - unique identifier 
"clientId" - unique identifier for every school
"datum" - date on which the PCR was performed  
"ergebnis" - PCR result (1 for positive and 2 for negative) 

- "inzidenz_nrw": 
downloaded from RKI https://survstat.rki.de/Content/Query/Create.aspx 
rows - districts 
columns - calendar weeks 
entries - 7-day-incidence for 4-10 year olds 

- "SSIperSchool":
"clientId" - unique identifier for every school 
"Kreisname" - district 
"Sozialindexstufe" - school social deprivation index

- "poolAufloesung":
based on script "poolResolution.R" 
"poolId" - unique identifier for pool-PCRs 
"singleId" - unique identifier for corresponding single-PCRs 
"Begruendung" - description if a positive pool-PCR for a positive single-PCR was found

- "pool_infos":
based on script "poolResolution.R" 
"poolsId" - unique identifier for pool-PCRs 
"clientId" - unique identifier for every school 
"nPos" - number of positive single-PCRs for the corresponding positive pool-PCR 
"datum" - date on which the pool-PCR was performed 

- "Soll_daten":
based on script "classSize.R"
"clientId" - unique identifier for every school 
"avgPoolGroesseVorJuni" - pool size when alternate classes were held 
"avgPoolGroesseAbJuni" - pool size for full-time schooling

- "districtPerSchool":
"clientId" - unique identifier for every school 
"district" - district 

### Output Figures 
Figure 4e,f and Extended Figure 8

## Class Size

### Input Data

- "df_list_clean$tbl_pools":
"poolsId" - unique identifier 
"clientId" - unique identifier for every school 
"datum" - date on which the PCR was performed 
"ergebnis" - PCR result (1 for positive and 2 for negative)

- "df_list_clean$tbl_singles":
"singleId" - unique identifier 
"clientId" - unique identifier for every school
"datum" - date on which the PCR was performed
"ergebnis" - PCR result (1 for positive and 2 for negative) 

- "df_list_clean$sys_clients"
"clientId" - unique identifier for every school
"clientName" - school name
"street" - school address
"zipcode" - zipcode where the school is located
"city" - city where the school is located


- "20210505_Pooltesting_Schulliste_vUpdate_small":
"Schulen" - school name 
"Addresse der Schule" - school address 
"PLZ" -  zipcode where the school is located 
"Stadt" - city where the school is located 
"Klassenname"	- class name 
"Klassenstaerke" - class size

### Output Data

- Soll_daten 
poolSollVorJuni : "avgPoolGroesseVorJuni" - pool size when alternate classes were held 
poolSollAbJuni: "avgPoolGroesseAbJuni" - pool size for full-time schooling

## Pool resolution

### Input Data

- "df_list_clean$tbl_pools":
"poolsId" - unique identifier 
"clientId" - unique identifier for every school 
"datum" - date on which the PCR was performed 
"ergebnis" - PCR result (1 for positive and 2 for negative) 
"klasseLabor" - class name

- "df_list_clean$tbl_singles":
"singleId" - unique identifier 
"clientId" - unique identifier for every school
"datum" - date on which the PCR was performed 
"ergebnis" - PCR result (1 for positive and 2 for negative)
"klasseLabor" - class name

- "df_list_clean$sys_clients"
"clientId" - unique identifier for every school 
"clientName" - school name 
"street" - school address 
"zipcode" - zipcode where the school is located 
"city" - city where the school is located 
"description" - another unique identifier for every school 

- "df_list_clean$tbl_stammdaten"
"clientId" - unique identifier for every school 
"avgClassSize" - average size of the classes in one school 

- "Altersstruktur 20210630_SuS_Schulen mit Primarstufe.xlsx"
"SCHULNR"- unique identifier for every school 
"Schulform" - type of school 
"SuS insgesamt" - Number of pupils per grade


### Output Data
- "pool_infos" 
"poolsId" - unique identifier for pool-PCRs 
"clientId" - unique identifier for every school 
"nPos" - number of positive single-PCRs for the corresponding positive pool-PCR 
"datum" - date on which the pool-PCR was performed 

- "poolAufloesung" 
"poolId" - unique identifier for pool-PCRs 
"singleId" - unique identifier for corresponding single-PCRs 
"Begruendung" - description if a positive pool-PCR for a positive single-PCR was found

## 1. System requirements
The code runs on Linux operating systems (Fedora 27) and has not been tested on other versions. 

The code was developed on R-version 3.5.4 including following packages:
```
reshape 0.8.8
ggplot2 3.3.5
ggplus 0.1
readxl 1.3.1
writexl 1.4.0
xlsx 0.6.5
```

The code requires only a standard computer.

## 2. Installation Guide
There is no installation process needed and run time is negligible. 

## 3. Demo
Run the script "SARS-CoV-2_infection_dynamics_in_schools.R" in folder "scripts/" that loads the demo data in folder "demo_data". The output is the Figure 4e,f and Extended Figure 8. The run time is negligible. 

## 4. Instructions for use
Scripts are available in folder scripts/.
