## Supplementary Data 8 for "Effective high-throughput RT-qPCR screening for SARS-CoV-2 infections in children": classSize.R

```
###set global options###
Sys.setlocale("LC_TIME", "en_US.UTF-8")
setwd("/cellnet/Ronja/LolliTest/Analysis/jansAnalysen")
###load libraries###
library(writexl)
library(xlsx)
###load data###
datum_heute <- as.Date('2021-09-23')
df_list_cleaned <- readRDS(file=paste0("/cellnet/Ronja/LolliTest/Data/",datum_heute,"/",datum_heute,"_df_list_clean.rds"))
df_list_cleaned$tbl_pools <- subset(df_list_cleaned$tbl_pools, strftime(df_list_cleaned$tbl_pools$datum, '%V') %in% c(19:26,33:37))
df_list_cleaned$tbl_single <- subset(df_list_cleaned$tbl_single, strftime(df_list_cleaned$tbl_single$datum, '%V') %in% c(19:26,33:37))

gc()
klassen <- read.xlsx(file = "../../Analysis/paper/data/20210505_Pooltesting_Schulliste_vUpdate_small.xlsx",startRow = 8,sheetIndex = 2,colIndex = 1:7,header = T)
colnames(klassen)[c(3,4,5)] <- c("PLZ","Stadt","leer")
###prepare and process data###
p <- df_list_cleaned$tbl_pools
s <- df_list_cleaned$tbl_single
cl <- df_list_cleaned$sys_clients
for(clname in colnames(cl)){
  if(is.character(cl[,clname])){
    Encoding(cl[,clname]) <- "latin1"
  }
}
# asd <- df_list_cleaned$tbl_stammdaten
# rownames(asd) <- asd$clientId
#match classes from state survey data and clients (schools)
klassenProClient <- lapply(1:nrow(cl),FUN=function(i){
  street <- cl[i,"street"]
  zip <- cl[i,"zipcode"]
  clname <- gsub(pattern = paste0(" [(]",street,".*"),"",cl[i,"clientName"])
  klassenGroessen <- klassen[klassen$Schulen==clname&klassen$PLZ==zip&klassen$Addresse.der.Schule==street,"Klassenstärke"]
  if(length(klassenGroessen)==0){return(NA)}
  return(klassenGroessen)
})
names(klassenProClient) <- cl$clientId

#before june, classes were half sized
i <- 0
poolSollVorJuni <- sapply(klassenProClient,FUN=function(kl){
  i<<-i+1
  if(all(is.na(kl))){return(NA)}
  kl <- kl/2
  kleineKlassen <- kl[kl<=25] #maximum pool size is 25
  grosseKlassen <- kl[kl>25]
  grosseKlassen <- unlist(lapply(grosseKlassen,FUN=function(n){
    fac <- ceiling(n/25)
    return(rep(floor(n/fac),fac))
  })) #classes that had to be split up into two pools anyways, dont have to be split further before june
  korrigierteKlassen <- c(kleineKlassen,grosseKlassen)
  return(mean(korrigierteKlassen))
})
i <- 0
poolSollAbJuni <- sapply(klassenProClient,FUN=function(kl){
  i<<-i+1
  if(all(is.na(kl))){return(NA)}
  #kl <- kl/2
  kleineKlassen <- kl[kl<=25]
  grosseKlassen <- kl[kl>25]
  grosseKlassen <- unlist(lapply(grosseKlassen,FUN=function(n){
    fac <- ceiling(n/25)
    return(rep(floor(n/fac),fac))
  }))
  korrigierteKlassen <- c(kleineKlassen,grosseKlassen)
  return(mean(korrigierteKlassen))
})  
###compare data on class size transmitted through the medeora database with that provided by the state government###
p_sub_vor_juni <- p[p$KinderImPool%in%(1:50)&p$datum<as.Date("2021-05-31"),]

#cor(asd[as.character(p_sub_vor_juni$clientId),"avgClassSize"],p_sub_vor_juni$KinderImPool,use="pair")
p_sub_ab_juni <- p[p$KinderImPool%in%(1:50)&!p$datum<as.Date("2021-05-31"),]

#cor(asd[as.character(p_sub_ab_juni$clientId),"avgClassSize"],p_sub_ab_juni$KinderImPool,use="pair")

medianVorJuni <- sapply(unique(p_sub_vor_juni$clientId),FUN=function(id){
  median(p_sub_vor_juni$KinderImPool[p_sub_vor_juni$clientId==id],na.rm=T)
})
names(medianVorJuni) <- unique(p_sub_vor_juni$clientId)

medianAbJuni <- sapply(unique(p_sub_ab_juni$clientId),FUN=function(id){
  median(p_sub_ab_juni$KinderImPool[p_sub_ab_juni$clientId==id],na.rm=T)
})
names(medianAbJuni) <- unique(p_sub_ab_juni$clientId)

meanVorJuni <- sapply(unique(p_sub_vor_juni$clientId),FUN=function(id){
  mean(p_sub_vor_juni$KinderImPool[p_sub_vor_juni$clientId==id],na.rm=T)
})
names(meanVorJuni) <- unique(p_sub_vor_juni$clientId)

meanAbJuni <- sapply(unique(p_sub_ab_juni$clientId),FUN=function(id){
  mean(p_sub_ab_juni$KinderImPool[p_sub_ab_juni$clientId==id],na.rm=T)
})
names(meanAbJuni) <- unique(p_sub_ab_juni$clientId)


###plots###
png(filename = "plots/klassenGroessKeinMedian_20210923.png",width=800)
par(mfrow=c(1,2))
plot(p_sub_vor_juni$KinderImPool,poolSollVorJuni[as.character(p_sub_vor_juni$clientId)],pch=19,col=rgb(0,0,0,0.1),xlab="Angabe ueber das Medeora Tool (vor Juni)",ylab="Durchschnittliche Klassengroesse (ASD)")
#abline(a=0,b=1,col="red")
#abline(a=0,b=2,col="green")

plot(p_sub_ab_juni$KinderImPool,poolSollAbJuni[as.character(p_sub_ab_juni$clientId)],pch=19,col=rgb(0,0,0,0.1),xlab="Angabe ueber das Medeora Tool (ab Juni)",ylab="Durchschnittliche Klassengroesse (ASD)")
#abline(a=0,b=1,col="red")
#abline(a=0,b=2,col="green")
dev.off()

pdf("plots/klassengroesseMedian_20210923.pdf",width = 12)
par(mfrow=c(1,2))

plot(medianVorJuni,poolSollVorJuni[names(medianVorJuni)],xlab="Median der Angabe ueber das Medeora Tool (vor Juni)",ylab="Durchschnittliche Klassengroesse (ASD)",pch=19,col=rgb(0,0,0,0.2))
abline(a=0,b=1,col="red")
#abline(a=0,b=2,col="green")

plot(medianAbJuni,poolSollAbJuni[names(medianAbJuni)],xlab="Median der Angabe ueber das Medeora Tool (ab Juni)",ylab="Durchschnittliche Klassengroesse (ASD)",pch=19,col=rgb(0,0,0,0.2))
abline(a=0,b=1,col="red")
#abline(a=0,b=2,col="green")
dev.off()

asdClassSizeVorJuni <- poolSollVorJuni[names(meanVorJuni)]
lmVorJuni <- lm(meanVorJuni~asdClassSizeVorJuni+0) #Relationship of expected to observed class sizes before june
asdClassSizeAbJuni <- poolSollAbJuni[names(meanAbJuni)]
#asdClassSizeAbJuni <- asdClassSizeAbJuni*ifelse(asdClassSizeAbJuni>25,0.5,1)
lmAbJuni <- lm(meanAbJuni~asdClassSizeAbJuni+0) #Relationship of expected to observed class sizes after june
pdf("plots/klassengroesseMean_20210923.pdf",width = 12)
par(mfrow=c(1,2))

plot(asdClassSizeVorJuni,meanVorJuni,ylab="Mittelwert der Angabe ueber das Medeora Tool (vor Juni)",xlab="erwartete Poolgroesse (ASD)",main=paste0("m=",round(lmVorJuni$coefficients,digits=2)),pch=19,col=rgb(0,0,0,0.2))
abline(lmVorJuni,col="red")
plot(asdClassSizeAbJuni,meanAbJuni,ylab="Mittelwert der Angabe ueber das Medeora Tool (ab Juni)",xlab="erwartete Poolgroesse (ASD)",main=paste0("m=",round(lmAbJuni$coefficients,digits=2)),pch=19,col=rgb(0,0,0,0.2))
abline(lmAbJuni,col="red")
dev.off()

###write Excel###
write_xlsx(path = "out/klassenGroessenMedeoraASD_20210923.xlsx",col_names = T,
           x = list(vorJuni=data.frame(clientId=names(medianVorJuni),medeoraMedian=medianVorJuni,medeoraMean=meanVorJuni,AsdAngepasst=asdClassSizeVorJuni),abJuni=data.frame(clientId=names(medianAbJuni),medeoraMedian=medianAbJuni,medeoraMean=meanAbJuni,AsdAngepasst=asdClassSizeAbJuni)))

###save R objects###
save(medianVorJuni,medianAbJuni,meanVorJuni,meanAbJuni,lmVorJuni,lmAbJuni,poolSollVorJuni,poolSollAbJuni,file = "out/medeoraKlassengroesseProSchule_20210923.RData")
```
