## Supplementary Data 8 for "Effective high-throughput RT-qPCR screening for SARS-CoV-2 infections in children": compute_number_children_adjusted.R

```
compute_number_children_adjusted <- function(df, Soll_daten, list_dates){
  
  dates_in_df <- unique(df$datum)
  
  if(is.null(list_dates)){
    Factor <- 1
    Modell <- 'Wechsel' 
    print('adjust ASD for all weeks')
  }
  else{
    if( length(unique(dates_in_df)) > 1 ){
      
      if(unique(list_dates$KW) == 19 & unique(strftime(list_dates$datum, '%Y')) == '2021'){
        Weekdays <- c('Mo','Di','Mi')
        Factor <- 1.5
      }else if(unique(list_dates$KW ) == 21 & unique(strftime(list_dates$datum, '%Y')) == '2021'){
        Weekdays <- c('Mi','Do')
        Factor <- 1
      }else if((unique(list_dates$KW) == 20 | unique(list_dates$KW) == 23 | unique(list_dates$KW) == 24) & unique(strftime(list_dates$datum, '%Y')) == '2021'){
        Weekdays <- c('Mo','Di','Mi','Do')
        Factor <- 2
      }else if(unique(list_dates$KW) == 22 & unique(strftime(list_dates$datum, '%Y')) == '2021'){
        Weekdays <- c('Mo','Di','Mi')
        Factor <- 2
      }else if(unique(list_dates$KW) == 33 & unique(strftime(list_dates$datum, '%Y')) == '2021'){
        Weekdays <- c('Mi','Do')
        Factor <- 1
      }else if(unique(list_dates$KW) >= 23 & unique(strftime(list_dates$datum, '%Y')) == '2021'){
        Weekdays <- c('Mo','Di','Mi','Do')
        Factor <- 2
      }else if(unique(list_dates$KW) >= 1 & unique(strftime(list_dates$datum, '%Y')) == '2022'){
        Weekdays <- c('Mo','Di','Mi','Do')
        Factor <- 2
      }else(print('KW?'))
    }else{Factor <- 1}
    if(unique(list_dates$KW) <= 21){
      Modell <- 'Wechsel'
    }else if(unique(list_dates$KW) >= 22){
      Modell <- 'Voll'
    }
  }
  
  if(unique(list_dates$KW) <= 26){
    Soll <- Soll_daten$VorFerien[[2]]
  }else if(unique(list_dates$KW) >= 33){
    Soll <- Soll_daten$NachFerien[[2]]
  }else if(unique(strftime(list_dates$datum, '%Y')) == '2022'){
    Soll <- Soll$NeuesJahr[[2]]
  }
  
  ids <- df$clientId
  # Number of tested schools
  uniqueClientId <- unique(ids)
  number_schools <- length(uniqueClientId)
  
  # compute number pools per school
  df_per_school <- split(df, df$clientId)
  number_pools <- lapply(df_per_school, function(school) nrow(school) )
  number_pools <- do.call(rbind,number_pools)
  number_pools <- data.frame('clientId'=rownames(number_pools),'NumberPools'=number_pools)
  
  # define pool size
  if(Modell == 'Wechsel'){PoolSize <- Soll[,c('clientId','avgPoolGroesseVorJuni')]}else if(Modell == 'Voll'){PoolSize <- Soll[,c('clientId','avgPoolGroesseAbJuni')]}
  PoolSize <- PoolSize[which(PoolSize[,'clientId'] %in% as.character(uniqueClientId)),,drop=F]
  PoolSize <- merge(number_pools, PoolSize, by='clientId',all.x=T)
  
  #compute number tested children per school
  number_tested_child_per_school <- data.frame('clientId'=PoolSize[,'clientId'],
                                               'ChildrenPerSchool'=((as.numeric(PoolSize[,3])*as.numeric(PoolSize$NumberPools))/Factor))
  number_tested_child <- sum(number_tested_child_per_school$ChildrenPerSchool, na.rm = TRUE)

  number_tested_child_per_school<- data.frame('clientId'=PoolSize$clientId,'Number tested children'=number_tested_child_per_school )
  df_general_values <- data.frame(number_schools, number_tested_child )
  colnames(df_general_values) <- c("Number tested schools", "Number tested children")
  
  return(list(df_general_values,number_tested_child_per_school))
}
```
