## Supplementary Data 8 for "Effective high-throughput RT-qPCR screening for SARS-CoV-2 infections in children": compute_number_pools.R

```
compute_number_pools <- function(data_list, list_dates, districtPerSchool){
  #select data based on date
  date <- list_dates$datum
  data <- select_tbl(tbl = data_list$tbl_pools, districtPerSchool = districtPerSchool, dates = date)
  #split per district
  district <- unique(data$district)
  dataPerDistrict <- split(data, data$district)
  
  #compute number of positive pools Per district
  Number_positive_Pools <- lapply(dataPerDistrict, function(tbl) length(unique(tbl[which(tbl$ergebnis == 1),"poolsId"])) )
  Number_positive_Pools <- do.call(rbind,Number_positive_Pools)
  Number_positive_Pools <- rbind('NRW'=sum(Number_positive_Pools[,1],na.rm = T),Number_positive_Pools)
  
  #compute number of pools Per district
  Number_Pools <- lapply(dataPerDistrict, function(tbl) length(unique((tbl$poolsId))) )
  Number_Pools <- do.call(rbind,Number_Pools)
  Number_Pools <- rbind('NRW'=sum(Number_Pools[,1],na.rm = T),Number_Pools)
  
  if(all.equal(rownames(Number_Pools),rownames(Number_positive_Pools))){
    # Percentage of positive Pools
    Percentage_pos_Pools <- round((Number_positive_Pools/Number_Pools)*100,2)
    Pools <- data.frame(rownames(Number_Pools),as.numeric(Number_Pools), as.numeric(Number_positive_Pools),
                        as.numeric(Percentage_pos_Pools))
    colnames(Pools) <-c('district','Number Pools','Number positive Pools',"Percentage positive Pools")
  }else{print("rownames of tables are not equal")}
  return(Pools)
}

select_tbl <- function(tbl, districtPerSchool, dates){
  data <- tbl[which(tbl$datum %in% dates), ]
  data <- merge(data, districtPerSchool, by='clientId')
  return(data)
}

subset_data_dates <- function(data_df, list_dates){
  List_df_date <-list()
  List_df_date$Uebersicht <- data_df[which(data_df$datum %in% list_dates),]
  List_df_date <- append(List_df_date,lapply(list_dates, function(date) subset(data_df, data_df$datum == date)))
  names(List_df_date)[-1] <- c(as.character(list_dates))
  return(List_df_date)
}
```
