## Supplementary Data 8 for "Effective high-throughput RT-qPCR screening for SARS-CoV-2 infections in children": poolResolution.R

```
###set global options###
Sys.setlocale("LC_TIME", "en_US.UTF-8")
###load required libraries###
library(writexl)
library(xlsx)
library(readxl)
###load data###
datum_heute <- as.Date('2021-11-26')
if (strftime(datum_heute, '%a') %in% c('Mon','Tue') ){KW <- as.numeric(strftime(datum_heute, '%V'))-1}else{KW <- as.numeric(strftime(datum_heute, '%V'))}
KW_names <- c(19:26,33:37)#2:KW#19:26,33:40,43:51,
KW<-37
df_list_cleaned <- readRDS(file=paste0("/cellnet/Ronja/LolliTest/Data/",datum_heute,"/",datum_heute,"_df_list_clean.rds"))
df_list_cleaned$tbl_pools <- subset(df_list_cleaned$tbl_pools, as.numeric(strftime(df_list_cleaned$tbl_pools$datum, '%V')) %in% KW_names)
df_list_cleaned$tbl_single <- subset(df_list_cleaned$tbl_single, as.numeric(strftime(df_list_cleaned$tbl_single$datum, '%V')) %in% KW_names)

schuldaten <- read_excel("/cellnet/Ronja/LolliTest/Data/Altersstruktur 20210630_SuS_Schulen mit Primarstufe.xlsx")
schuldaten <- as.data.frame(schuldaten)
###prepare data for analysis###
p <- df_list_cleaned$tbl_pools
s <- df_list_cleaned$tbl_single
cl <- df_list_cleaned$sys_clients
for(clname in colnames(cl)){
  if(is.character(cl[,clname])){
    Encoding(cl[,clname]) <- "latin1"
  }
}
asd <- df_list_cleaned$tbl_stammdaten
rownames(asd) <- asd$clientId

schuldaten_sub <- unique(schuldaten[,c("SCHULNR","Schulform")])
table(table(schuldaten_sub[,1]))#Confirm that there is only one type assigned to each school (i.e. elementary or special needs)
schulform <- sapply(cl$description,FUN=function(id){
  schuldaten_sub[which(schuldaten_sub[,"SCHULNR"]==id)[1],"Schulform"]
})
names(schulform) <- cl$clientId
schulform_red <- schulform
schulform_red[schulform_red%in%c("Grundschule","Freie Waldorfschule")] <- "Grundschule"
schulform_red[schulform_red%in%c("Förderschule G/H","Waldorfförderschule")] <- "Förderschule"
schulform_red[!schulform_red%in%c("Förderschule","Grundschule")] <- "Sonstige Schulform"

###In what time range do single PCRs arrive after a positive pool?###
schulenMitEinemPosPool <- names(which(table(p[which(p$ergebnis=="1"),"clientId"])==1))
p_einmal <- p[which(p$ergebnis=="1"&p$clientId%in%schulenMitEinemPosPool),]
tagNachPool <- lapply(schulenMitEinemPosPool,FUN=function(schule){
  poolDatum <- p_einmal[which(p_einmal$clientId==schule),"datum"]
  sDatum <- s[which(s$clientId==schule),"datum"]
  if(length(sDatum)==0){return(NULL)}
  dauerInTagen <- sDatum-poolDatum
  dauerInTagen <- dauerInTagen[dauerInTagen>=0]
  return(dauerInTagen)
})
wochenTagProSchule <- sapply(schulenMitEinemPosPool,FUN=function(schule){
  weekdays(p_einmal$datum[which(p_einmal$clientId==schule&p_einmal$ergebnis=="1")])
})
nachX <- sapply(0:10,FUN=function(i){
  sum(unlist(tagNachPool)==i)
})
nachX <- c(nachX,length(unlist(tagNachPool))-sum(nachX))
nachX <- cbind(c("am gleichen Tag",paste0(1:10, "Tag/e spaeter"),"mehr als 10 Tage spaeter"),nachX)
colnames(nachX) <- c("Zeitpunkt","Anzahl")
nachX <- cbind(nachX,Prozent=round(as.numeric(nachX[,2])/sum(as.numeric(nachX[,2]))*100,digits=2))
nachX <- cbind(nachX,Anzahl_kummulativ=cumsum(as.numeric(nachX[,2])))
nachX <- cbind(nachX,Prozent_kummulativ=round(as.numeric(nachX[,4])/sum(as.numeric(nachX[,2]))*100,digits=2))
###Collect information on positive pools###
#Which pool is positive?
p_pos <- p[which(p$ergebnis=="1"),]
#Were there additional positive pools in the same school?
posPoolDavorDanach <- t(sapply(1:nrow(p_pos),FUN=function(i){
  pID <- p_pos[i,"poolsId"]
  schule <- p_pos[i,"clientId"]
  poolDatum <- p_pos[i,"datum"]
  poolsAnDerSchule <- p_pos[p_pos$clientId==schule,]
  letzterPool <- tail(sort(poolsAnDerSchule[which(poolsAnDerSchule$poolsId!=pID&poolsAnDerSchule$datum<=poolDatum),"datum"]),n=1)
  if(length(letzterPool)==0){letzterPool <- NA}
  naechsterPool <- head(sort(poolsAnDerSchule[which(poolsAnDerSchule$poolsId!=pID&poolsAnDerSchule$datum>=poolDatum),"datum"]),n=1)
  if(length(naechsterPool)==0){naechsterPool <- NA}
  return(c(as.character(letzterPool),as.character(naechsterPool)))
}))
colnames(posPoolDavorDanach) <- c("letzterPosPool","naechsterPosPool")
posPoolDavorDanach <- data.frame(posPoolDavorDanach)
posPoolDavorDanach[,1] <- as.Date(posPoolDavorDanach[,1])
posPoolDavorDanach[,2] <- as.Date(posPoolDavorDanach[,2])
p_pos <- cbind(p_pos,posPoolDavorDanach)
##Which single PCR can be assigned to which pool?
grenze <- 5 #Number of days after the positive pool result in which the single PCR is expected.
poolProEinzel <- t(sapply(1:nrow(s),FUN=function(i){
  sDatum <- s[i,"datum"]
  schule <- s[i,"clientId"]
  poolsSeitX <- which(p_pos$clientId==schule&p_pos$datum%in%(sDatum-(0:grenze)))
  anzahlPoolsSeitX <- length(poolsSeitX)
  if(anzahlPoolsSeitX==0){
    return(c(NA,"kein positiver Pool der Schule")) # There was no positive pool at the respective school in the time frame of interest.
  }
  if(anzahlPoolsSeitX==1){
    return(c(poolsSeitX,"einziger positiver Pool"))#There was only one positive pool at the school in this time frame
  }
  einzelNamen <- s[i,"klasseLabor"]
  poolNamen <- p_pos[poolsSeitX,"klasseLabor"]
  if(sum(!is.na(poolNamen))>0&!is.na(einzelNamen)){
    if(einzelNamen%in%poolNamen){
      korrekterPool <- poolsSeitX[which(poolNamen==einzelNamen)]
      if(length(korrekterPool)>1){
        return(c(NA,"mehrere positive Pools ohne korrekte Namen (mehrfache Namen)")) #Several pools had the same name as the single pcr.
      }else{
        return(c(korrekterPool,"mehrere positive Pools mit korrekten Namen")) #There were multiple positive pools but the single pcrs could be matched through unique names.
      }
    }
  }
  return(c(NA,"mehrere positive Pools ohne korrekte Namen (nicht gefunden)")) #Names were not available or matched between pools and singles.
}))
poolProEinzel[!is.na(poolProEinzel[,1]),1] <- p_pos[as.numeric(poolProEinzel[!is.na(poolProEinzel[,1]),1]),"poolsId"]
colnames(poolProEinzel) <- c("poolId","Begruendung")
statusProSingle <- data.frame(singleId=s$singleId,poolProEinzel)

#How many single PCRs are available per pool, what are their results.
nEinzelProPool <- t(sapply(p_pos$poolsId,FUN=function(i){
  s_sub <- s[which(poolProEinzel[,1]==i),]
  nPos <- sum(s_sub[,"ergebnis"]=="1")
  nNeg <- sum(s_sub[,"ergebnis"]=="2")
  return(c(nPos+nNeg,nPos,nNeg))
}))
colnames(nEinzelProPool) <- c("gesamt","nPos","nNeg")
poolNachverfolgungStats <- data.frame(c("Pools ohne Einzelproben","Pools mit Einzelproben, aber ohne positive Einzelprobe","Pools mit mindestens einer positiven Einzelprobe"),
           c(sum(nEinzelProPool[,"gesamt"]==0),sum(nEinzelProPool[,"gesamt"]!=0&nEinzelProPool[,"nPos"]==0),sum(nEinzelProPool[,"nPos"]>0)))
colnames(poolNachverfolgungStats) <- c("Pool_Status","Anzahl")
#When was the first positive single, when was the last?
datenPosEinzel <- t(sapply(p_pos$poolsId,FUN=function(i){
  s_sub <- s[which(poolProEinzel[,1]==i),]
  erstePosEinzel <- min(s_sub$datumErgebnis[s_sub$ergebnis=="1"])-p[p$poolsId==i,"datum"]
  letztePosEinzel <- max(s_sub$datumErgebnis[s_sub$ergebnis=="1"])-p[p$poolsId==i,"datum"]
  out <- c(erstePosEinzel,letztePosEinzel)
  out[is.infinite(out)] <- NA
  return(out)
}))
tageBisAufloesungMin <- table(datenPosEinzel[,1])[as.character(0:5)]
print(head(tageBisAufloesungMin))
tageBisAufloesungMin <- c(tageBisAufloesungMin,sum(datenPosEinzel[,1]>5,na.rm=T))
print(head(tageBisAufloesungMin))
names(tageBisAufloesungMin) <- c(paste0(0:5," Tage"),"mehr als 5 Tage")
tageBisAufloesungMin <- data.frame(tageBisAufloesung=c(paste0(0:5," Tage"),"mehr als 5 Tage"),Anzahl=tageBisAufloesungMin)
rownames(tageBisAufloesungMin) <- NULL

#Determine class size per pool if possible.
klassenGroesseSpez <- sapply(1:nrow(p_pos),FUN=function(i){
  klassenNamen <- p_pos[i,"klasseLabor"]
  if(is.na(klassenNamen)){return(NA)}
  schule <- p_pos[i,"clientId"]
  schulID <- cl[schule,"description"]
  klassenDerSchule <- schuldaten[schuldaten$SCHULNR==schulID,]
  dieseKlasse <- klassenDerSchule[klassenDerSchule$LFDNR==klassenNamen,"SuS insgesamt"]
  gesamtSchueler <- sum(dieseKlasse)
  return(gesamtSchueler)
})
klassenGroesseSpez[which(klassenGroesseSpez==0)] <- NA
#Were the results transmitted after 6 am on the following day?
ergebnisNach6 <- (p_pos$datumErgebnis>p_pos$datum&p_pos$uhrzeitErgebnis>0.25)|(p_pos$datumErgebnis>(p_pos$datum+1))
ergebnisNach6 <- ifelse(ergebnisNach6,"spaet","frueh")
#On which weekday was the positive pool?
poolWochenTag <- weekdays(p_pos$datum)
#In which week was the pool?
kwProTag <- paste0("KW",as.numeric(strftime(p_pos$datum, format = "%V")))


#Combine information
p_pos_df <- data.frame(p_pos,
                       schulform=schulform_red[as.character(p_pos$clientId)],
                       durschnittlicheGroesse=asd[as.character(p_pos$clientId),"avgClassSize"],
                       spezKlassenGroesse=klassenGroesseSpez,
                       ergebnisNachSechs=ergebnisNach6,
                       poolWochenTag=poolWochenTag,
                       KW=kwProTag,
                       nEinzelProPool)
#Adjust class size after may.
p_pos_df <- data.frame(p_pos_df,angepassteGroesse=p_pos_df$durschnittlicheGroesse)
p_pos_df[which(p_pos_df$datum>=as.Date("2021-05-31")&p_pos_df$durschnittlicheGroesse>25),"angepassteGroesse"] <- p_pos_df[which(p_pos_df$datum>=as.Date("2021-05-31")&p_pos_df$durschnittlicheGroesse>25),"angepassteGroesse"]/2
###Analysis###
aufloesungProLabor <- t(sapply(unique(p_pos_df$laboratory),FUN=function(lab){
  ps <- p_pos_df[which(p_pos_df$laboratory==lab&p_pos_df$gesamt>0),]
  aufloesungProzent <- sum(ps$nPos>0)/nrow(ps)*100
  return(c(sum(ps$nPos>0),nrow(ps),round(aufloesungProzent,digits=2)))
}))
colnames(aufloesungProLabor) <- c("pools_mit_positiver_einzelprobe","positive_pools_mit_einzelproben","aufloesung_prozent")
#by school type
aufloesungProSchulform <- t(sapply(unique(p_pos_df$schulform),FUN=function(sf){
  ps <- p_pos_df[which(p_pos_df$schulform==sf&p_pos_df$gesamt>0),]
  aufloesungProzent <- sum(ps$nPos>0)/nrow(ps)*100
  return(c(sum(ps$nPos>0),nrow(ps),round(aufloesungProzent,digits=2)))
}))
colnames(aufloesungProSchulform) <- c("pools_mit_positiver_einzelprobe","positive_pools_mit_einzelproben","aufloesung_prozent")
#by time of transmission of the result
aufloesungNachUhrzeit <- t(sapply(as.vector(na.omit(unique(p_pos_df$ergebnisNachSechs))),FUN=function(tp){
  ps <- p_pos_df[which(p_pos_df$ergebnisNachSechs==tp&p_pos_df$gesamt>0),]
  aufloesungProzent <- sum(ps$nPos>0,na.rm=T)/nrow(ps)*100
  return(c(sum(ps$nPos>0,na.rm=T),nrow(ps),round(aufloesungProzent,digits=2)))
}))
colnames(aufloesungNachUhrzeit) <- c("pools_mit_positiver_einzelprobe","positive_pools_mit_einzelproben","aufloesung_prozent")
#by weekday
aufloesungNachTag <- t(sapply(sort(as.vector(na.omit(unique(p_pos_df$poolWochenTag))),decreasing=T),FUN=function(tp){
  ps <- p_pos_df[which(p_pos_df$poolWochenTag==tp&p_pos_df$gesamt>0),]
  aufloesungProzent <- sum(ps$nPos>0,na.rm=T)/nrow(ps)*100
  return(c(sum(ps$nPos>0),nrow(ps),round(aufloesungProzent,digits=2)))
}))
colnames(aufloesungNachTag) <- c("pools_mit_positiver_einzelprobe","positive_pools_mit_einzelproben","aufloesung_prozent")
aufloesungNachTag <- aufloesungNachTag[rowSums(aufloesungNachTag,na.rm=T)>0,]
if (all(c("Monday","Tuesday","Wednesday","Thursday","Friday") %in% rownames(aufloesungNachTag))){
  aufloesungNachTag <- aufloesungNachTag[c("Monday","Tuesday","Wednesday","Thursday","Friday"),]
}
#by week
aufloesungNachWoche <- t(sapply(sort(as.vector(na.omit(unique(p_pos_df$KW)))),FUN=function(tp){
  ps <- p_pos_df[p_pos_df$KW==tp&p_pos_df$gesamt>0,]
  aufloesungProzent <- sum(ps$nPos>0,na.rm=T)/nrow(ps)*100
  return(c(sum(ps$nPos>0),nrow(ps),round(aufloesungProzent,digits=2)))
}))
colnames(aufloesungNachWoche) <- c("pools_mit_positiver_einzelprobe","positive_pools_mit_einzelproben","aufloesung_prozent")

###Write xlsx###
xlList <- list(eingang_einzelproben=nachX,
               zuordnung_einzel_zu_pools=statusProSingle,
               tage_bis_aufloesung=tageBisAufloesungMin,
               nachverfolgung_pro_pool=poolNachverfolgungStats,
               gesammelte_pool_infos=p_pos_df,
               aufloesung_wochentag=aufloesungNachTag,
               aufloesung_KW=aufloesungNachWoche,
               aufloesung_labor=aufloesungProLabor,
               aufloesung_schulform=aufloesungProSchulform,
               aufloesung_zeit_ergebnis=aufloesungNachUhrzeit)
xlList <- lapply(xlList,FUN=function(l){
  if(!is.null(rownames(l))){
    l <- data.frame(Zeilennamen=rownames(l),l)
  }else{
    l <- as.data.frame(l)
  }
  return(l)
})

write_xlsx(x = xlList,path = paste0("/cellnet/Ronja/LolliTest/Reporting_output/",datum_heute,"/",datum_heute,"_poolAufloesung.xlsx"),col_names = T)

#Number of positive single PCRs per pool
kws <- na.omit(unique(p_pos_df$KW))
posSinglesProPoolUndKW <- sapply(kws,FUN=function(kw){
  ps <- p_pos_df[p_pos_df$nPos>0&p_pos_df$KW==kw,]
  mean(ps$nPos)
})
posSinglesProPoolUndKW
posSinglesProPoolUndKW2 <- sapply(kws,FUN=function(kw){
  ps <- p_pos_df[p_pos_df$KW==kw,]
  mean(ps$nPos)
})
posSinglesProPoolUndKW2
saveRDS(posSinglesProPoolUndKW,paste0("/cellnet/Ronja/LolliTest/Reporting_output/",datum_heute,"/",datum_heute,"posSinglesProPoolUndKW.rds"))
library(ggplot2)
df<-data.frame(y=names(posSinglesProPoolUndKW),x=posSinglesProPoolUndKW)
df$y <- as.factor(df$y)
df$y <- factor(df$y, levels=c(paste0('KW',KW_names)))
pdf(paste0("/cellnet/Ronja/LolliTest/Reporting_output/",datum_heute,"/",datum_heute,"_positive_einzel_pro_positivem_pool.pdf"),width = 10,height = 5)
plot<-ggplot(df) + geom_bar(aes(y=x,x=y),stat='identity') + xlab('Kalendarwoche')+ylab('Positive Einzel PCRs pro positivem Pool')
print(plot)
dev.off()
```
