## Supplementary Data 8 for "Effective high-throughput RT-qPCR screening for SARS-CoV-2 infections in children": SARS-CoV-2_infection_dynamics_in_schools.R

```
setwd("/data/public/rjohnen/LolliTest/Lolli-Test-NRW/")
Sys.setlocale("LC_TIME", "en_US.UTF-8")
source('scripts/compute_number_pools.R')
source('scripts/compute_number_children_adjusted.R')
library(readxl)
library(ggplot2)
library(ggplus)
library(reshape)

#####load data#####
# Pool and Single-PCR Results
df_list_clean <- readRDS(file='demo_data/df_list_clean_demo.rds')

#Use shorter variable names
p <- df_list_clean$tbl_pools
p$KW <-strftime( p$datum, "%V")
s <- df_list_clean$tbl_single
KWs <-  c(19:25,34:37)

#Inzidenz
inzidenz_nrw <- readRDS("demo_data/inzidenz_nrw.rds")
inzidenz_nrw_melt<- melt(inzidenz_nrw)
inzidenz_nrw_melt$KW <- sapply(as.character(inzidenz_nrw_melt$variable), function(x) strsplit(x, "KW")[[1]][[2]])
colnames(inzidenz_nrw_melt)[3] <- "incidence"

# SSI
SSIperSchool <- readRDS("demo_data/SSIperSchool_demo.rds")
# Poolauflösung
poolAufloesung <- readRDS("demo_data/poolAufloesung_demo.rds")
# Pool Infos
pool_infos <- readRDS("demo_data/pool_infos_demo.rds")
# Soll daten
Soll_daten <- readRDS("demo_data/Soll_daten_demo.rds")

#####Compute positiverate per district#####
#Load KreisProSchule
districtPerSchool<-readRDS('demo_data/districtPerSchool_demo.rds')

# add calendarweek and split per calendarweek
p_split <- split(p, p$KW)
Number_tested_pools <- list()
for(KW in 1:length(p_split) ){
  Number_tested_pools[[KW]] <- compute_number_pools(list("tbl_pools"=p_split[[names(p_split)[KW]]]), 
                                                    p_split[[names(p_split)[KW]]], 
                                                    districtPerSchool=districtPerSchool)
  Number_tested_pools[[KW]]$KW <- names(p_split)[KW]
}
names(Number_tested_pools)<-names(p_split)
Number_tested_pools<-do.call(rbind,Number_tested_pools)
colnames(Number_tested_pools)[4] <- "positiverate"

####Filter Pools####
# Filter pools based on the expected number of single PCRs performed.
# We use only the positive pools that have as many single PCRs as we would expect (+-20%).
pools_assignable <-subset(poolAufloesung, poolAufloesung$Begruendung == "einziger positiver Pool" |
                          poolAufloesung$Begruendung == "mehrere positive Pools mit korrekten Namen" )
pools_unassignable <-subset(poolAufloesung, poolAufloesung$Begruendung != "einziger positiver Pool" &
                                poolAufloesung$Begruendung != "mehrere positive Pools mit korrekten Namen" )

#filter for resolved pools (at least 1 positive single pcr)
resolved_pools <- pool_infos[pool_infos$nPos>0,"poolsId"]

pools_split<-split(pools_assignable,pools_assignable$poolId)
keep_pool <- rep(NA,length(pools_split))
for (i in 1:length(pools_split)){
  singles <- length(unique(pools_split[[i]]$singleId))
  school <-pool_infos[which(pool_infos$poolsId %in% pools_split[[i]]$poolId),'clientId']
  poolDate <- as.Date(pool_infos[which(pool_infos$poolsId==pools_split[[i]][1,2]),"datum"])
  
  if(poolDate >= "2021-08-16"){
    poolsize <- Soll_daten$NachFerien[[2]][which(Soll_daten$NachFerien[[2]]$clientId == school),'avgPoolGroesseAbJuni']
  }else if(poolDate < "2021-08-16" && poolDate >= "2021-05-31"){
    poolsize <- Soll_daten$VorFerien[[2]][which(Soll_daten$VorFerien[[2]]$clientId == school),'avgPoolGroesseAbJuni']
  }else if(poolDate < "2021-05-31"){
    poolsize <- Soll_daten$VorFerien[[2]][which(Soll_daten$VorFerien[[2]]$clientId == school),'avgPoolGroesseVorJuni']
  }else{print(i)}
  if(singles/poolsize >= 0.8 && singles/poolsize <= 1.2){
    keep_pool[i] <- T
  }else{
    keep_pool[i] <- F
  }
}
pools_keep <- names(pools_split)[keep_pool]
pools_rm <- names(pools_split)[!keep_pool]

#filter for resolved pools (at least 1 positive single pcr)
resolved_pools <- pool_infos[pool_infos$nPos>0,"poolsId"]
not_resolved_pools <- pool_infos[pool_infos$nPos==0,"poolsId"]

#combine
pools_keep_final <- intersect(resolved_pools, pools_keep) 
pools_rm_final <- unique(c(not_resolved_pools, pools_rm,unique(pools_unassignable$poolId))) 
pools_filter <- subset(pools_assignable, pools_assignable$poolId %in% pools_keep_final)
colnames(pools_filter)[2] <- "poolsId"

#remove singles ids where we don't have a corresponding pool
s <- s[s$ergebnis%in%c(1,2),]
s <- s[(s$singleId %in% pools_filter$singleId),]
#remove pools
p <- p[!(p$poolsId %in% pools_rm_final),]

####Calculate number of children tested per school#### 
p_split <- split(p, p$KW)
children_per_school<-list()
for(KW in 1:length(p_split)){
  children_per_school[[KW]] <- compute_number_children_adjusted(df=p_split[[KW]], list_dates=list(datum=unique(p_split[[KW]]$datum),KW=p_split[[KW]]$KW), Soll_daten=Soll_daten)
  children_per_school[[KW]] <- children_per_school[[KW]][[2]]
  children_per_school[[KW]]$KW <- unique(p_split[[KW]]$KW)
}
names(children_per_school) <- names(p_split)
children_per_school <- do.call(rbind,children_per_school)
colnames(children_per_school)[3]<-'ChildrenPerSchool'


####Match single and pool data####
p <- merge(p, pools_filter, by='poolsId', all.x = TRUE)

# add SSI, positiverate and incidence per Pool
p$district <- SSIperSchool$Kreisname[match(p$clientId,SSIperSchool$clientId)]
p$SSI <- as.numeric(SSIperSchool$Sozialindexstufe[match(p$clientId,SSIperSchool$clientId)])
p <- merge(p, inzidenz_nrw_melt[,c("district","KW","incidence")], by=c("district","KW"),all.x = T)
p <- merge(p, Number_tested_pools[,c("district","positiverate","KW")], by=c("district","KW"),all.x = T)

p$ergebnisEinzel <- s[match(p$singleId,s$singleId),"ergebnis"]
p <- p[p$ergebnisEinzel%in%c(1,2),]
p$ergebnisEinzel <- 1-(p$ergebnisEinzel-1) # 0 is negativ and 1 is positive

# for every school in each KW, sum up the positive children
p_agg = aggregate(p[,c("ergebnisEinzel")],
                  by = list(p$KW, p$clientId),
                  FUN = sum, na.rm = TRUE)
colnames(p_agg) <- c("KW","clientId","nPos")


####Combine in one table####
# one row is one school per calendarweek
SchoolData <- data.frame(clientId=p_agg$clientId,
                        SSI=SSIperSchool[match(p_agg$clientId,SSIperSchool$clientId),"Sozialindexstufe"],
                        district=SSIperSchool[match(p_agg$clientId,SSIperSchool$clientId),"Kreisname"],
                        KW=p_agg$KW,
                        nPos_Kind=p_agg$nPos)
SchoolData <- merge(SchoolData, inzidenz_nrw_melt[,c("district","KW","incidence")], by=c("district","KW"),all.x = T)
SchoolData <- merge(SchoolData, Number_tested_pools[,c("district","positiverate","KW")], by=c("district","KW"),all.x = T)
SchoolData <- merge(SchoolData, children_per_school, by=c("clientId","KW"),all.x = T)

SchoolData <- SchoolData[apply(!is.na(SchoolData),1,all),] # Remove special needs school and incomplete annotations
SchoolData$incidence <- as.numeric(SchoolData$incidence) # otherwise each number is a class
SchoolData$nPos_Kind <- as.numeric(SchoolData$nPos_Kind) 
SchoolData$nPos_Kind_perc <- (SchoolData$nPos_Kind/SchoolData$ChildrenPerSchool)
SchoolData$SSI <- as.numeric(SchoolData$SSI)

#### perform logistic regression####
Formula <- "nPos_Kind_perc ~ incidence:SSI + SSI + incidence + positiverate:SSI + positiverate"
glm_schule <- glm(Formula, SchoolData, family=quasibinomial(link = "logit"))

summary(glm_schule)

####Calculate expected value####
p<-p[!is.na(p$SSI),] # Remove special needs school and incomplete annotations 
# predict propability of a positive child based on the logistic model
p$pPerChild <- predict(glm_schule,p , type='response')

###Get expectations per pool### 
poolIds <- unique(p$poolsId)
statsPerPool <- t(sapply(poolIds,FUN=function(pID){
  nSingles <- sum(p$poolsId==pID)-1
  nPos <- sum(p$poolsId==pID&p$ergebnisEinzel==1)-1 # remove one positive child per pool because we want to get the expected value for an extra positive child
  nNeg <- (nSingles-nPos)
  pPerKid <- unique(p[p$poolsId==pID,'pPerChild'])
  p_0_1 <- dbinom(c(0:5),size = nSingles,prob = pPerKid)
  p_bigger1 <- 1-sum(p_0_1)
  return(c(as.numeric(pID),nSingles,nPos,nNeg,pPerKid,p_0_1,p_bigger1))
}))
colnames(statsPerPool) <- c("poolID","nChildren","nPos","nNeg","pPerKid","p0","p_1","p_2","p_3","p_4","p_5","p_bigger5")
statsPerPool <- data.frame(statsPerPool)

generalPoolInfos <- unique(p[,c("poolsId","KW","SSI","positiverate",'datum')])
identical(as.numeric(generalPoolInfos$poolsId),statsPerPool$poolID)
Final_df <-cbind(statsPerPool[,c('poolID','nChildren','nPos','nNeg','pPerKid','p0')],generalPoolInfos)
Final_df$p_bigger1 <- 1-Final_df$p0
Final_df$KW<-as.numeric(as.character(Final_df$KW))
# add variants 
Final_df$Variante <- c(rep(NA,nrow(Final_df)))
Final_df$Variante[Final_df$KW>=33] <- 'delta'
Final_df$Variante[Final_df$KW<33] <- 'alpha'

# for each variant compute the observed and expected value
Final_df_split <-split(Final_df,Final_df$Variante)
Observed<-sapply(Final_df_split, function(x) sum(x[,"nPos"]>0))
Expected<-sapply(Final_df_split, function(x) sum(x[,"p_bigger1"]))

#### Visualization ####
# prepare plot
df_plot<- data.frame(names(Observed),Observed,Expected,Observed/Expected)
colnames(df_plot) <- c("variant","observed","expected","observed/expected")

# Extended Figure 8a
ggplot(p[!duplicated(p),]) + geom_boxplot(aes(x=as.character(KW),y=pPerChild))+xlab('calendar week')+
  ylab('Probability to be positive per child')
# Figure 4e
ggplot( melt(df_plot[,-4]) ) + geom_bar(aes(x=variant,y=value,fill=variable),position="dodge",stat="identity")+theme_readable()+theme(legend.title = element_blank() )+ylab("Number of Pool-PCRs")
# Figure 4f
ggplot(df_plot) + geom_bar(aes(x=variant,y=observed/expected),stat="identity")+theme_readable()


#### Wilcoxon-test alpha vs delta ####
Final_df$Expected_Value <-(Final_df$pPerKid*Final_df$nChildren)
Final_df$enrichment <- Final_df$nPos-Final_df$Expected_Value
 
# Extended Figure 8c
ggplot(Final_df) + geom_violin(aes(x=Variante,y=enrichment))+ylab('Number of additional positiv children per pool observed -\n Number of additional positiv children per pool expected')

# subset only pools with positive enrichment (Observed more than expected)
Final_df_pos <- Final_df[which(Final_df$enrichment > 0),]
alpha<-subset(Final_df_pos, Final_df_pos$Variante =='alpha')
delta<-subset(Final_df_pos, Final_df_pos$Variante =='delta')
# perform wilcoxon rank test 
wilcox.test(alpha$enrichment,delta$enrichment)


#### Visualization per KW####
statsPerKW <- t(sapply(sort(unique(generalPoolInfos$KW)),FUN=function(KW){
  whichPools <- which(generalPoolInfos$KW==KW)
  poolsThisDay <- length(whichPools)
  observation <- c(sum(statsPerPool[whichPools,"nPos"]==0),sum(statsPerPool[whichPools,"nPos"]>0))
  expectation <- colSums(data.frame(statsPerPool[whichPools,c("p0")],1-statsPerPool[whichPools,c("p0")]))
  return(c(poolsThisDay,observation,expectation))
}))
colnames(statsPerKW) <- c("totalPools","observed0","observedBigger1",
                          "expected0","expectedBigger1")
statsPerKW <- data.frame(KW=as.character(sort(unique(generalPoolInfos$KW))),statsPerKW)
statsPerKW <- data.frame(statsPerKW,enrBiggerone=statsPerKW$observedBigger1/statsPerKW$expectedBigger1)

statsPerKW$KW<-as.numeric(as.character(statsPerKW$KW))
statsPerKW$Variante <- c(rep(NA,nrow(statsPerKW)))
statsPerKW$Variante[statsPerKW$KW>=33] <- 'delta'
statsPerKW$Variante[statsPerKW$KW<33] <- 'alpha'
statsPerKW$KW<-as.character(statsPerKW$KW)

# Extended Figure 8b
ggplot(statsPerKW)+geom_point(aes(x=expectedBigger1,y=observedBigger1,col=KW),size=5)+
  theme_readable()+geom_abline()+ylab('Observation - Pool-RT-qPCR with more than one positive child ')+xlab('Expectation - Pool-RT-qPCR with more than one positive child ')
```
